# Reversing Immunosenescence with Senolytics to Enhance Tumor Immunotherapy

**DOI:** 10.1101/2024.10.14.24315428

**Authors:** Niu Liu, Jiaying Wu, Enze Deng, Jianglong Zhong, Bin Wei, Tingting Cai, Xiaohui Duan, Sha Fu, David O. Osei-Hwedieh, Ou Sha, Yunsheng Chen, Xiaobin Lv, Yingying Zhu, Lizao Zhang, Hsinyu Lin, Qunxing Li, Peichia Lu, Jiahao Miao, Teppei Yamada, Lei Cai, Hongwei Du, Sylvan C. Baca, Qingpei Huang, Soldano Ferrone, Xinhui Wang, Fang Xu, Xiaoying Fan, Song Fan

## Abstract

Recent advancements in cancer immunotherapy have improved patient outcomes, yet responses to immunotherapy remain moderate. We conducted a Phase II clinical trial (NCT04718415) involving 51 cancer patients undergoing neoadjuvant chemoimmunotherapy and applied single-cell RNA and T/BCR sequencing on tumor and blood samples to elucidate the immune cell perturbations. Our findings associate poor response with reduced levels of CCR7^+^CD4 Naïve T cells and CD27^+^ Memory B cells, as well as higher expression of immunosenescence-related genes in T and B cell subsets. Using naturally aged and *Ercc1^+/-^* transgenic aging mouse models, we found that senolytics enhance the therapeutic efficacy of immunotherapy in multiple solid tumors by mitigating tumor immunosenescence. Notably, we launched a Phase II clinical trial, COIS-01 (NCT05724329), which pioneers the combination of senolytics with anti-PD-1 therapy. The clinical results demonstrate that this therapeutic strategy is associated with a favorable safety profile and therapeutic efficacy, significantly mitigating adverse effects and alleviating immunosenescence. These findings underscore the pivotal role of immunosenescence characteristics in influencing the effectiveness of immunotherapy and suggest a promising therapeutic efficacy along with a beneficial safety assessment for the combination of senolytics with anti-PD-1 therapy.

## Introduction

Despite recent advancements in blocking the PD-1/PD-L1 pathway, which have revolutionized the treatment of solid tumors, only a small fraction of patients benefits from immunotherapy, with fewer than 20% showing sustained responses ^1–3^. A major challenge in current immunotherapy and neoadjuvant strategies is our limited understanding of the highly dynamic and heterogeneous tumor immune microenvironment (TIME), which impacts treatment responses ^4–6^. The concept of cancer immunoediting emphasizes the immune system’s dual role in inhibiting tumor growth and shaping tumor immunogenicity. This process is described in three stages: elimination, equilibrium, and escape. During the elimination phase, the host’s innate and adaptive immune systems recognize and respond to tumor-specific antigens ^7,8^. T cells are the primary players in tumor immunity. Although tumor-reactive T cells are present in TIME, the phenotypic heterogeneity of T cells across different tumors and their divergent differentiation fates remain significant. Exhaustion still poses a major limitation to their anti-tumor potential^9,10^.

Immunosenescence, a well-established phenomenon occurring with age, results in the immune system losing its ability to effectively respond to pathogens and cancer cells ^11–15^. Recent studies have uncovered the complex role of immunosenescence in tumors, with factors such as cAMP, glucose competition, and oncogenic stress in the TIME inducing senescence in T cells, macrophages, natural killer (NK) cells, and dendritic cells, thereby impairing their immune cytotoxic function against tumors ^16^. However, it remains unclear whether individual differences in immunosenescence affect the efficacy of immunotherapy in cancer patients.

Previous studies have shown that blocking DNA damage signals can inhibit the senescence of tumor-infiltrating T cells, thereby enhancing the efficacy of immune checkpoint inhibitors (ICIs) against tumors ^17^. Recent research also indicate that chronic inflammation caused by an aging immune system can promote cancer development, regardless of the age of the cancerous or surrounding tissues ^18^. Therefore, we believe that the senescence status of immune cell subpopulations, which play a crucial role in tumor immunity, may be critical for treatment outcomes. We hypothesize that senolytics can be combined with ICIs to enhance their efficacy in treating solid tumors.

In this study, we first explored the variability in patient responses to neoadjuvant immunotherapy, starting with a Phase II clinical trial in head and neck squamous cell carcinoma (HNSCC, NCT04718415). Further analysis of single-cell RNA sequencing (scRNA-Seq) data from these patients allowed us to identify key immune cell subpopulations that influence immune responses and to reveal the negative impact of immunosenescence on immunotherapy efficacy. Additionally, animal study data highlighted the potential benefits of combining senolytics with ICIs for treating solid tumors, including HNSCC, bladder cancer, and breast cancer. We then initiated a pioneering Phase II clinical trial (COIS-01, NCT05724329) to evaluate the efficacy and safety of combining anti-aging drugs with ICIs in HNSCC patients. Our results indicate that the neoadjuvant combination of senolytics and ICIs significantly reduces toxic side effects compared to neoadjuvant chemotherapy. Furthermore, patients receiving the combination therapy exhibited a higher proportion of CCR7^+^ naïve T cells within their tumors, suggesting a mitigation of immunosenescence and an enhancement of the adaptive immune response.

## Results

### Phase II trial evaluating neoadjuvant chemotherapy combined with immunotherapy for oral cavity and oropharyngeal squamous cell carcinoma (OOC-001)

We enrolled 51 patients with stage II-IVA HNSCC for neoadjuvant chemoimmunotherapy (OOC-001, NCT04718415). One patient was excluded from the evaluation due to refusing to continue the treatment (Extended Data Fig. 1a). All patients underwent a minimum of two cycles of neoadjuvant treatment, with 24 (47.1%) receiving 2 cycles, 23 (45.1%) receiving 3 cycles, and 4 (7.8%) receiving 4 cycles of neoadjuvant treatment (Tables 1-2 and Supplementary Tables 1-2). Among the 48 evaluable patients, 23 patients achieved pathological complete response (pCR, 47.9%), 8 patients achieved major pathological response (MPR, 16.7%), 10 patients showed partial pathological response (pPR, 20.8%), and 7 patients displayed pathological no response (pNR, 14.6%) (Fig. 1a, Extended Data Fig. 1b). The comparison of radiologic and pathologic response indicators suggested that certain patients were pathologically assessed as pCR while radiologically assessed as PR (Partial response)/SD (Stable disease), highlighting significant inconsistency between pathological and radiological assessments (Fig. 1b). Compared to the baseline, 72.7% of primary tumors exhibited shrinkage, while only 46.7% of patients showed regression in lymph nodes, possibly indicating that primary tumors in HNSCC are more sensitive to neoadjuvant therapy (Fig. 1c and Extended Data Fig. 1c-e). Regarding treatment cycles, pathological assessment suggested that the efficacy of the 2-cycle treatment might be less than that of 3 or 4 cycles (Extended Data Fig. 1f). 48 patients (94%) experienced at least one treatment-related adverse effect (TRAE). Grade 3-4 TRAEs were observed in 26 patients (51%), and the most common TRAEs were alopecia (75%), asthenia or fatigue (53%), and nausea (49%) (Table 2).

**Fig. 1.**
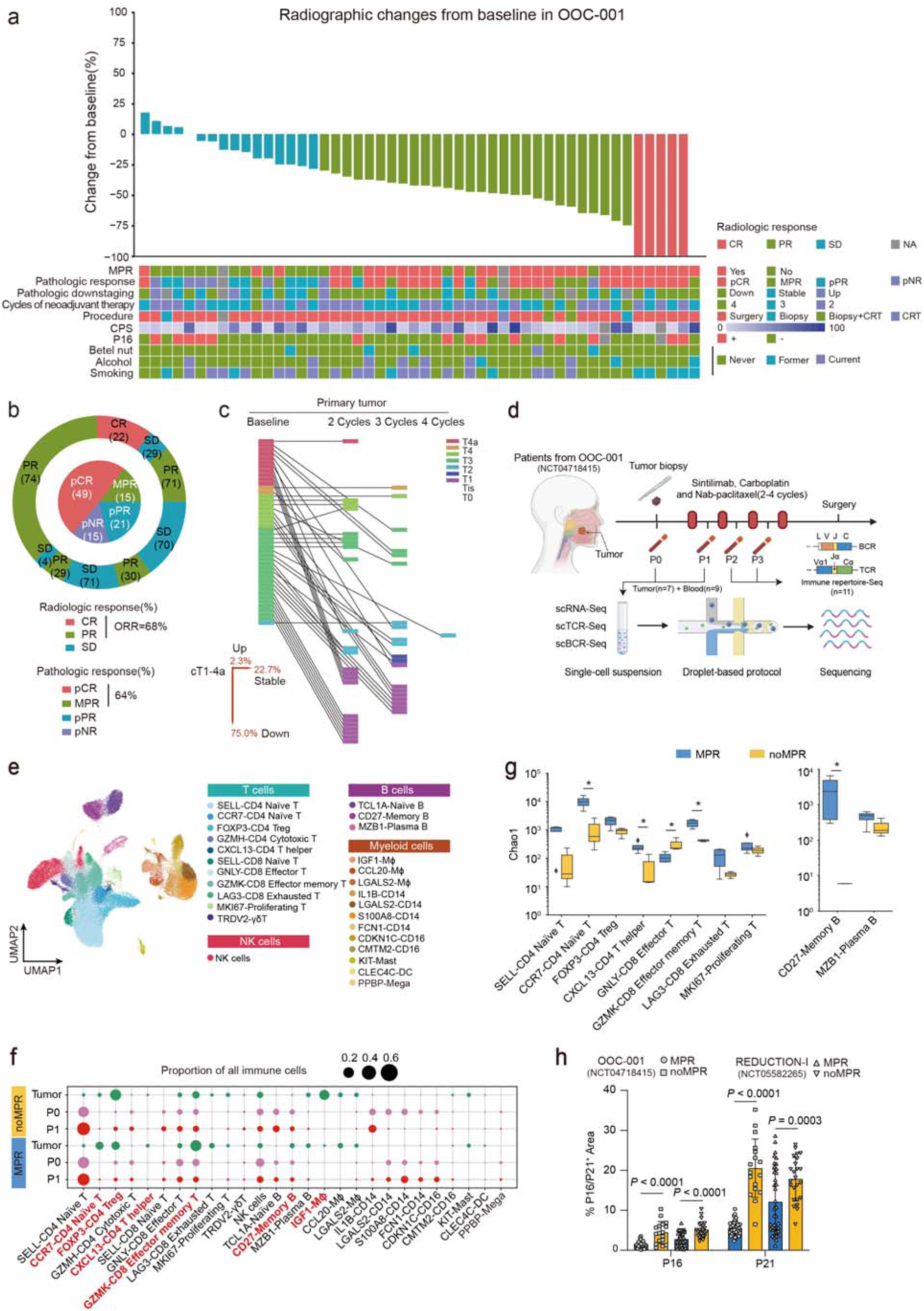
The results of the Phase II clinical trial (OOC-001) and scRNA-Seq data indicate that MPR patients demonstrate increased diversity in immune response-related cell subtypes and exhibit decreased expression of senescence markers. **a**, Waterfall plot depicting the treatment response of patients in the HNSCC neoadjuvant clinical trial OOC-001 (NCT04718415, n=50). **b**, Pie charts depicting radiological and pathological response rates in OOC-001. **c**, The changes and proportional statistics of TNM staging after treatment for primary tumors in different patients. **d**, Schematic overview of the experimental design and analytical workflow. **e**, UMAP plot of the immune cells that passed quality control. Immune cell clusters are annotated and marked by color code. **f**, Prevalence of each single-cell cluster (column) in all samples (n = 7) from different groupings of tissue (rows). Green dots represent tumors samples, pink dots represent peripheral blood samples of pre-treatment (P0), and red dots represent peripheral blood samples of post-treatment (P1). **g**, Estimation of size and diversity of TCR repertoires for T cell clusters and BCR repertoires for B cell clusters between MPR and noMPR groups in tumors. The number of TCR and BCR clonotypes estimated by Chao1 (Materials and Methods) illustrates the predicted theoretical clonotype diversity. Data are expressed as mean ± s.e.m. **h**, Quantifications of immunofluorescent staining for senescent markers P16 and P21 on paraffin sections from MPR and noMPR patients in two patient cohorts (Cohort1 n=50, Cohort2 n=61). *P < 0.05, **P < 0.01, ***P < 0.001.

**Table 1.** Demographic and clinical characteristics of OOC-001.

| Characteristics | Patients (n=50) |
| --- | --- |
| Age, median (range), years | 56 (26, 80) |
| **Sex, n (%)** |  |
| Male | 38 (76.0) |
| Female | 12 (24.0) |
| **Smoking, n (%)** |  |
| Never | 23 (46.0) |
| Former | 9 (18.0) |
| Current | 18 (36.0) |
| **Primary tumor site, n (%)** |  |
| Oral tongue | 23 (46.0) |
| Buccal mucosa | 4 (8.0) |
| Gingiva | 7 (14.0) |
| Floor of mouth | 7 (14.0) |
| Oropharynx | 7 (14.0) |
| Hard palate | 2 (4.0) |
| **ECOG scores, n (%)** |  |
| 0 | 38 (76.0) |
| 1 | 12 (24.0) |
| **Pretreatment clinical T stage, n (%)** |  |
| T1 | 1 (2.0) |
| T2 | 22 (44.0) |
| T3 | 10 (20.0) |
| T4 | 1 (2.0) |
| T4a | 16 (32.0) |
| **Pretreatment clinical N stage, n (%)** |  |
| N0 | 29 (58.0) |
| N1 | 5 (10.0) |
| N2a | 2 (4.0) |
| N2b | 5 (10.0) |
| N2c | 9 (18.0) |
| **AJCC overall clinical disease stage, n (%)** |  |
| II | 20 (40.0) |
| III | 8 (16.0) |
| IVA | 22 (44.0) |
| **Cycles of neoadjuvant therapy, n (%)** |  |
| 2 | 24 (48.0) |
| 3 | 22 (44.0) |
| 4 | 4 (8.0) |
| **HPV status, n (%)** |  |
| Positive | 14 (28.0) |
| Negative | 34 (68.0) |
| Not evaluable or missing | 2 (4.0) |
| **CPS, n (%)** |  |
| <10 | 15 (30.0) |
| ≥10 | 33 (66.0) |
| Not evaluable or missing | 2 (4.0) |

**Table 2.** Treatment-related adverse events of OOC-001 (*n*=51)

| TRAEs, n (%) | Any grade | Grade 1-2 | Grade 3-4 |
| --- | --- | --- | --- |
| Any treatment-related adverse events | 48 (94.1) | 48 (94.1) | 10 (19.6) |
| Alopecia | 38 (74.5) | 38 (74.5) | 3 (5.9) |
| Asthenia or fatigue | 27 (52.9) | 27 (52.9) | 0 |
| Nausea | 25 (49.0) | 25 (49.0) | 0 |
| Arthralgia | 13 (25.5) | 13 (25.5) | 0 |
| Skin disorders | 14 (27.5) | 14 (27.5) | 4 (7.8) |
| Diarrhoea | 12 (23.5) | 12 (23.5) | 0 |
| Gastroenteritis | 10 (19.6) | 9 (17.6) | 1 (1.9) |
| Myalgia | 8 (15.7) | 8 (15.7) | 0 |
| Vomiting | 8 (15.7) | 8 (15.7) | 0 |
| Decreased appetite or anorexia | 8 (15.7) | 8 (15.7) | 0 |
| Paraesthesia | 9 (17.6) | 8 (15.7) | 1 (1.9) |
| Pruritus | 6 (11.8) | 6 (11.8) | 0 |
| Anaemia | 6 (11.8) | 6 (11.8) | 0 |
| Hyperthyroidism | 7 (13.7) | 5 (9.8) | 2 (3.9) |
| Increased aminotransferases | 4 (7.8) | 4 (7.8) | 1 (1.9) |
| Leucopenia | 2 (3.9) | 1 (1.9) | 2 (3.9) |
| Pyrexia | 1 (1.9) | 1 (1.9) | 0 |

| **TRAEs, *n* (%)** | **Any grade** | **Grade 1-2** | **Grade 3-4** |
| --- | --- | --- | --- |
| Thrombopenia | 2 (3.9) | 1 (1.9) | 1 (1.9) |
| Hypothyroidism | 1 (1.9) | 1 (1.9) | 0 |
| Pulmonary infection | 1 (1.9) | 0 | 1 (1.9) |

The 24-month overall survival (OS) for the cohort was 93.9% (Extended Data Fig. 1g, h). Our findings show that combining sintilimab with carboplatin and nab-paclitaxel resulted in manageable side effects and achieved a high overall response rate (64.6%, MPR and pCR). Considering the trade-off between anti-tumor effectiveness and adverse effects, we observed that a 3-cycle treatment regimen offered superior benefits for patients with HNSCC.

### Immune landscape of HNSCC patients with neoadjuvant chemoimmunotherapy

ScRNA-Seq and scT/BCR-Seq was performed in seven treatment-naïve tumor samples and nine blood samples (including pre-and post-treatment) (Fig. 1d, Supplementary Table 1). Following quality filtering, we obtained scRNA-Seq profiles as well as TCR and BCR clonotypes from 143,056 cells, encompassing T cells, NK cells, B cells, myeloid cells, fibroblasts, endothelium, and tumor cells (Supplementary Figure 1a-f). A total of 27 cell clusters with distinct molecular features were observed by re-clustering each major immune cell type (Fig. 1e).

We analyzed the abundance of various immune subtypes and compared their proportions across the tumor, pre-treatment blood (P0) and post-treatment blood (P1) of MPR and noMPR patients. The proportion of CD4^+^ T cells showed the largest elevation among all immune cell types when comparing P1 to P0 (Supplementary Figure 1g, h). While compared to noMPR patients, MPR patients’ tumors exhibited higher proportions of CCR7^+^CD4 Naïve T cells, CXCL13^+^CD4 T cells, GZMK^+^CD8 Tem cells, and CD27^+^ Memory B cells. Conversely, the proportions of FOXP3^+^CD4 Treg cells and IGF1^+^ macrophages (IGF1^+^ Mφ) were lower (Fig. 1f and Supplementary Figure 2, 3a-f). The upregulated genes in noMPR patients of the main T/B cell subtypes were involved in aging-related pathways, indicating these pathways may contribute to variations in the treatment response status of HNSCC patients. (Supplementary Figure 3g, Supplementary Table 3). Using T/BCR diversity as markers of antigenic immune responses to neoadjuvant therapy, we investigated TCR and BCR clonality in MPR and noMPR groups through three different methods (Chao1, Shannon Index, and Richness). The results showed that MPR patients exhibited higher TCR and BCR clonal diversity in CCR7^+^CD4 Naïve T cells, CXCL13^+^CD4 T cells, GZMK^+^CD8 Tem cells, and CD27^+^ Memory B cells (Fig. 1g and Supplementary Figure 3h). Additionally, dominant clones present in the tumor and blood before treatment persisted as the dominant clones after treatment in MPR patients (Extended Data Fig. 2a-c). Further analysis of TCR/BCR clonotypes from P0, P1, and tumors revealed no significant differences in TCR/BCR clonotypes in blood before and after treatment, nor between MPR and noMPR patients. Interestingly, MPR patients exhibited a greater number of unique TCR/BCR clonotypes within tumors, suggesting a richer TCR/BCR clonal diversity (Extended Data Fig. 2d, e).

Immunosenescence refers to the decline in the immune system’s ability to effectively respond to pathogens and cancer cells ^11^. Recent studies have found that characteristics of immunosenescence include an imbalance in the proportion of naïve and memory T cells, a decrease in the number of TCR clones, and elevated levels of *p16^INK4a^* (also known as *CDKN2A*) and *p21^CIP1^*(*CDKN1A*) expression ^19,20^. We then examined the expressions of the senescent markers, P16 and P21, by immunostaining in tumor tissues from two clinical trials conducted at our center (OOC-001 and REDUCTION-I, NCT05582265). The results revealed that the tumor microenvironment of noMPR patients exhibited higher levels of senescent features (Fig. 1h and Extended Data Fig. 2f). In summary, our data suggests that non-responsive patients to neoadjuvant treatment demonstrate dysregulation in immune cell proportions and an increase in senescent characteristics.

### T cell and B cell subtypes exhibiting senescent features influence the response to immunotherapy

Recent studies have shown that in late life, the aging individual’s adaptive immune system exhibits dysfunction and increased autoimmunity, with T and B cell aging being a primary manifestation of immunosenescence ^21,22^. However, there is no gold standard marker for defining the senescence of T and B cells ^23^. To figure out this assumption, we first created an immunosenescence-associated gene set (IAGs) consists of 154 genes based on extensive scRNA-Seq datasets differentiating immune cells from young and elderly individuals (Extended Data Fig. 3a, Supplementary Table 4). The IAGs were significantly enriched in the immune cells of elderly individuals in three independent scRNA-Seq datasets (GSE157007 ^24^, GSE141595 ^25^ and PRJCA002856) (Fig. 2a and Extended Data Fig. 3b-d), indicating they could reflect human immunosenescence characteristics. Additionally, evaluating IAGs in scRNA-Seq datasets for HNSCC and triple-negative breast cancer (TNBC) ^26^ immunotherapies also revealed lower senescent scores in MPR patients compared to noMPR patients (Extended Data Fig. 3e). Subsequently, we used IAGs to score T cells and their subtypes, which revealed lower immunosenescence features in MPR patients with HNSCC (Fig. 2b and Extended Data Fig. 4a). Furthermore, we also identified senescent genes associated with T cells from the IAGs, which showed higher expression levels in multiple T cell subgroups of noMPR patients, with GO analysis revealed enrichment in pathways related to S100 protein binding (Fig. 2c and Extended Data Fig. 4b, c). Similar results were obtained from the TNBC dataset (Extended Data Fig. 4d-h).

**Fig. 2.**
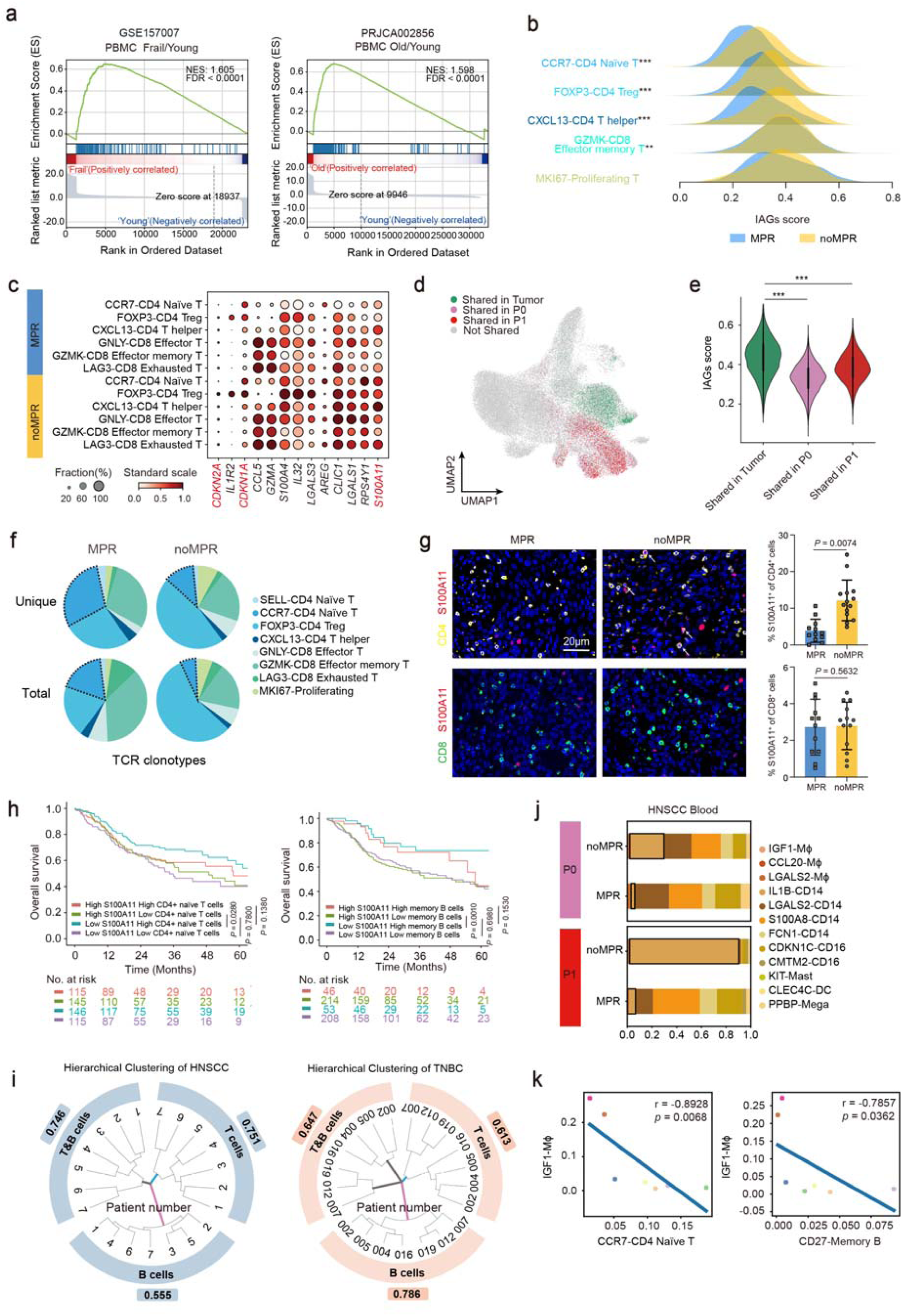
Patient responses to immunotherapy are associated with the expression of IAGs in both T and B cell subsets, as well as the interaction of IGF1^+^ Mφ with T and B cells. **a**, The IAGs significantly enriched in all immune cells in peripheral blood of frail elderly people (GSE157007) and old people (PRJCA002856). **b**, The Ridge plot of IAGs score analysis for major T cells clusters. **c**, Dot plot of intersection genes of HNSCC T cell DEGs and IAGs between MPR and noMPR groups. **d**, UMAP of T cells from 4 paired tumor and blood samples (12 samples total), highlighting shared clones between tumors (green) and P0 (pink, pre-treatment blood) and P1 (red, post-treatment blood). **e**, Shared clonal cells ratio between tumor (green) and P0 (pink, pre-treatment blood) and P1 (red, post-treatment blood) T cells from 4 paired tumor and blood samples. **f**, Paired TCR results Shows the overall number of TCR clones (upper row), and the status of single clones after normalization (lower row). **g**, Immunohistological staining quantifications of CD4 and S100A11 in HNSCC patients of two cohorts (n=12 per group). **h**, Low expressions of S100A11 and high proportion of CD4 Naïve T cells and memory B cells were significant associated with better OS. **i**, Hierarchical clustering tree shows different models distinguish pathological responses of patients following the combination therapy before surgery. T&B cells mean using the proportional distribution of T and B cells and representative genes as a model to predict the patient’s response. T cells alone or B cells alone means using the proportional distribution of T cells or B cells and representative genes as a model to predict patient response to treatment (the inner circle is the HNSCC patients’ number, and the outer circle is the cophenetic distance result to evaluate the effect). **j**, The cellular compositions of myeloid cell clusters in P0 and P1 between MPR and noMPR groups in HNSCC. **k**, Proportion of tumor-infiltrating IGF1^+^ Mφ was inversely correlated to the proportion of tumor infiltrating CCR7^+^CD4 Naïve T and CD27^-^ Memory B in HNSCC. Pearson correlation (two-sided).

Relative to noMPR patients, MPR patients exhibited lower Vα/β gene usage frequencies in CCR7^+^CD4 T cells and FOXP3^+^CD4 Treg cells in both tumors and blood, consistent with previous findings (Extended Data Fig. 5a). Pseudotime analysis of immune cells revealed that CD4^+^ T cell subsets in peripheral blood exhibit an early differentiation phenotype, while their tumor counterparts show a more differentiated phenotype. Notably, MPR patients demonstrated differentiation towards CCR7^+^CD4 Naïve T cells and CXCL13^+^CD4 Th cells, maintaining T cell vitality and the potential to differentiate into various functional subtypes. In contrast, CD4^+^ T cells in non-MPR patients tended to differentiate towards FOXP3^+^CD4 Tregs, leading to immunosenescence-related characteristics (Extended Data Fig. 5b-d).

Numerous T cells demonstrate shared clonotypes between tumors and blood. These clonal T cells in tumor showed elevated IAGs scores than those in blood, suggesting a higher level of senescent characteristics for the tumor-infiltrating T cells (Fig. 2d, e). Analysis of paired TCR data revealed a notable reduction of both unique and total clonotypes particularly in the CCR7^+^CD4 Naïve T cells in noMPR patients compared with MPR patients (Fig. 2f). Immunosenescence features are notably pronounced in B cells of noMPR patients, particularly in CD27^+^ Memory B cells (Extended Data Fig. 6a-e). Pairwise analysis of T and B cell receptors and ligands revealed strong interactions between FOXP3^+^CD4 Treg cells and CXCL13^+^CD4 T helper cells with B cells (Extended Data Fig. 6f). This indicates that in MPR patients, the reduced number of FOXP3^+^CD4 Tregs and increased number of CXCL13^+^CD4 T helpers may exert their effects through interactions with B cells.

Through immunofluorescence staining, we confirmed that CD4^+^ T cells and CD27^+^ memory B cells expressing the senescent marker S100A11 were more abundant in noMPR patients, while no difference was observed in the number of S100A11^+^CD8 T cells between noMPR and MPR patients, suggesting senescent status of CD4 T cells critically modulate patients’ response to neoadjuvant therapy (Fig. 2g and Extended Data Fig. 6g). Analysis of HNSCC Patients’ data from the TCGA database found that patients with high expression of IAGs such as *S100A11*, *AREG*, *CALD1*, *CDKN1A*, and *CASP4* together with low CD4 Naïve T cell or memory B cell abundance had significantly worse prognosis than those with low expression of senescent genes and high proportions of CD4 Naïve T cells or memory B cells (Fig. 2h and Supplementary Figure 4). T cells predict responses to immunotherapy more effectively in HNSCC patients, while in TNBC patients, B cells are more predictive (Fig. 2i). These analyses suggest that the IAGs we constructed were effective in distinguishing the immune cells of noMPR and MPR patients, and that lower expression levels of IAGs in CCR7^+^CD4 Naïve T and CD27^+^ Memory B cells are associated with a positive treatment response.

### IGF1^+^ Mφ induce senescent characteristics in T cells

Innate immune cells play an important role in early tumor cell recognition and subsequent initiation of inflammation and antitumor responses. To investigate whether non-lymphocyte immune cells are involved in the immunosenescence phenotype observed in our study subjects, we analyzed the gene expression features of myeloid cells. IGF1^+^ Mφ exhibited the highest differentiation score among all myeloid cell types according to pseudotime analysis (Extended Data Fig. 7a). The precursor cells of IGF1^+^ Mφ in the blood are IL1B-CD14 cells, which showed significant changes in the MPR group before and after treatment. Interestingly, the proportion of IGF1^+^ Mφ within tumors and IL1B-CD14 cells in blood were significantly higher in noMPR patients (Fig. 2j and Extended Data Fig. 7b-d). Additionally, IGF1^+^ Mφ negatively correlated with CCR7+CD4 Naïve T cells and CD27+ Memory B cells in tumors (Fig. 2k). We also examined the composition proportions of myeloid cell subtypes in TNBC patients and found that PR patients had a higher proportion of IGF1^+^ Mφ compared to SD patients, consistent with our HNSCC results (Extended Data Fig. 7e, f). By scoring the expression of six major senescence-associated secretory phenotype (SASP) factors in HNSCC and TNBC, we found that IL1B-CD14 monocytes and IGF1^+^ Mφ expressed higher levels of SASP factors compared to other myeloid cell subtypes (Extended Data Fig. 7g, h). Interaction analysis further revealed that IGF1^+^ Mφ had a strong potential for interaction with T cells (Supplementary Figure 5a-c). Enrichment analysis of pathways related to IGF1^+^ Mφ and IL1B-CD14 cells revealed involvement in senescent and inflammatory pathways, including ERK, MAPK and IL-1 pathways. Consequently, IGF1^+^ Mφ in the TIME may induce immunosenescence in T cells. Analysis of TCGA data for HNSCC patients demonstrated that those with high IGF1 signature had relatively poor OS and progression-free survival (PFS) (Extended Data Fig. 7i and Supplementary Figure 5d, e). In conclusion, our analysis of myeloid cell subtypes suggests that IGF1^+^ Mφ could be a crucial factor contributing to the limited response of tumor patients to immunotherapy. These cells may trigger an immunosenescent status in the TIME through their interactions with T cells.

### Senolytics combined αPD-1 alleviate TIME senescent burden in HNSCC

Currently, the role of the senescent phenotype in HNSCC and its interaction with tumor cells, immune cells, or other cell types remains poorly understood. The impact of senolytics treatments or their combination with other therapies in HNSCC is yet to be determined. Histochemical staining with β-galactosidase revealed a significantly more pronounced senescent phenotype in HNSCC tissue compared to the adjacent normal tissue (Fig. 3a). Then, we established a 4-NQO-induced (4-Nitroquinoline-N-oxide) HNSCC mouse model using 12-month-old C57BL/6 (C57) mice (Fig. 3b). The mice underwent the typical progression of normal epithelium-epithelial dysplasia-invasive cancer, with an average age of tumor occurrence around 20 months, reflecting the real progression of HNSCC in elderly patients more accurately (Extended Data Fig. 8a). Immunofluorescence staining on tumor sections confirmed significantly higher expression of the key senescent marker P16 in the tumor region compared to surrounding normal tissue (Fig. 3c).

**Fig. 3.**
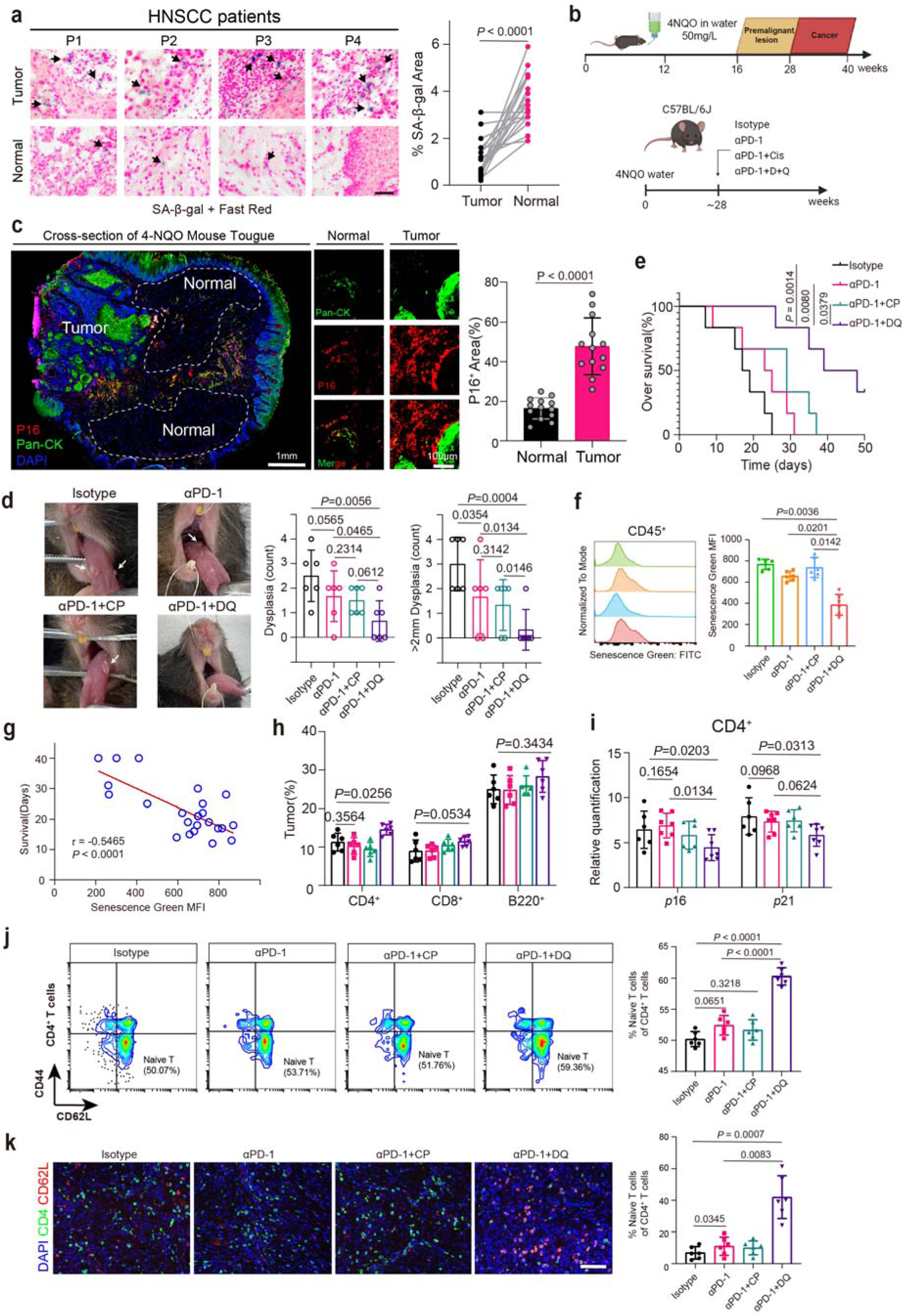
The combination of dasatinib and quercetin with αPD-1 enhances therapeutic efficacy in a chemically induced HNSCC mouse model by alleviating immunosenescence. **a,** Representative images of SA-β-gal and eosin staining for senescence markers in tumors and adjacent normal tissues from HNSCC patients, along with paired quantitative analysis of SA-β-gal positive areas. Scale bar, 50 μm. **b,** Flowchart for Inducing HNSCC in C57 mice by adding 4NQO to drinking water (Top). Schematic representation of different groups in 4NQO-induced mice, including Isotype intraperitoneal injection (Isotype), αPD-1 intraperitoneal injection (αPD-1), αPD-1+Cisplatin intraperitoneal injection (αPD-1+CP), αPD-1 intraperitoneal injection plus Dasatinib+Quercetin oral gavage (αPD-1+DQ). Isotype or αPD-1 was administered at 200 μg per mouse, cisplatin at 3 mg/kg, given on days 0, 7, and 14 of the experiment; Dasatinib (5 mg/kg) and Quercetin (50 mg/kg) were administered on days 0, 3, 7, 10 and 14 (Bottom). **c,** Immunohistological staining and staining area quantifications of SASP marker P16 and tumor marker Pan-CK in the 4NQO induced HNSCC mice. Scale bar, 10 μm. **d,** Representative images of tongue of mice after receiving Isotype, αPD-1, αPD-1+CP or αPD-1+DQ treatment. White arrows indicate tumor-like nodules (left); Dysplasia area quantifications of the mice treated with Isotype, αPD-1, αPD-1+CP or αPD-1+DQ (n= 6 per group) (right). **e,** Mouse over survival (Surviving mice/Total number of mice x 100%) after receiving Isotype, αPD-1, αPD-1+CP or αPD-1+DQ treatment (n= 6 per group). **f,** Flow cytometry analysis of the expression of the Senescence marker β-gal in immune cells within mouse tumors after receiving Isotype, αPD-1, αPD-1+CP or αPD-1+DQ treatment (n= 5 per group). **g,** Scatter plot showing the correlation between the expression level of β-galactosidase in immune cells and the survival time of mice. **h,** Flow cytometry analysis of CD4^+^, CD8^+^, and B cell proportions in 4-NQO mouse tumors after receiving Isotype, αPD-1, αPD-1+CP or αPD-1+DQ treatment (n= 6 per group). **i,** Expression of senescence markers (*p16^Ink4a^* and *p21^Cip1^* mRNA) in CD4^+^ TILs in mouse tumors after receiving Isotype, αPD-1, αPD-1+CP or αPD-1+DQ treatment (n= 6 per group). **j,** Flow cytometry analysis of CD4^+^ Naïve T cells proportions in mouse tumors after receiving Isotype, αPD-1, αPD-1+CP or αPD-1+DQ treatment (n= 6 per group). **k,** Representative images and quantifications of immunofluorescent staining for CD4^+^CD62L^+^ T cells after receiving Isotype, αPD-1, αPD-1+CP or αPD-1+DQ treatment (n= 6 per group). The statistical analysis was performed using t test (a), one-way ANOVA (c, d, f, h, i, j, k) and log-rank test (e). *P* values are shown, and error bars indicate the mean ± SEM.

To further investigate the effect of senolytics on HNSCC, we randomly divided the 4-NQO-induced tumor mice into four groups, administering Isotype, αPD-1 (anti PD-1), αPD-1+CP (anti PD-1+Cisplatin), or αPD-1+DQ (anti PD-1+Dasatinib+Quercetin) treatments (Fig. 3b). The αPD-1+DQ group exhibited fewer lesions compared to other groups. Interestingly, there was no significant difference between the αPD-1 group and the αPD-1+CP group (Fig. 3d and Extended Data Fig. 8b) while αPD-1+DQ group had a longer lifespan compared to other groups (Fig. 3e and Extended Data Fig. 8c). Moreover, flow cytometry revealed that αPD-1+DQ treatment effectively decreased β-galactosidase expression in CD45^+^ immune cells within tumors (Fig. 3f). Correlation analysis further demonstrated a negative relationship between mice survival time and β-galactosidase expression in immune cells, confirming the anti-tumor effect of αPD-1+DQ by reversing immunosenescence in HNSCC mice (Fig. 3g).

Subsequently, we evaluated changes in CD4^+^, CD8^+^, and B220^+^ cells in the spleen, blood, bone marrow, and tumors across different treatment groups using flow cytometry. The αPD-1+DQ group showed an increased proportion of CD4^+^ cells within tumors, while CD8^+^ and B220^+^ cell levels exhibited no significant differences compared to other groups (Fig. 3h). Similar results were observed in the spleen (Extended Data Fig. 8d, e). However, B220^+^ cell counts were relatively elevated in peripheral blood samples from the αPD-1+DQ group (Extended Data Fig. 8f-g). Additionally, the expression levels of *p16* and *p21* were significantly reduced in tumor-infiltrating CD4^+^ cells from the αPD-1+DQ group (Fig. 3i), whereas no significant differences were noted in CD8^+^ cells (Extended Data Fig. 8h). Furthermore, an increased proportion of CD62L^+^CD44^-^ naïve CD4^+^ cells was observed in the αPD-1+DQ group (Fig. 3j). Immunofluorescence staining consistently confirmed a significant increase in CD4^+^CD62L^+^ cells in the αPD-1+DQ group relative to the other groups (Fig. 3k and Supplementary Figure 6). Together, these findings suggest that immunosenescence in HNSCC surpasses that in surrounding normal tissue, and the combination of αPD-1+DQ may enhance the therapeutic efficacy of αPD-1 by mitigating the senescent phenotype of tumor immune cells, particularly in CD4^+^ cells.

### Synergistic reduction of SASP and increase in CD4^+^ Naïve T cells by senolytics and αPD-1 in solid tumors

To determine the effectiveness of αPD-1+DQ in various transplant tumor models, including HNSCC, we established two mouse oral squamous cell carcinoma cell lines, 4N-MS1 and 4N-MS2, from the 4-NQO model (Fig. 4a, Extended Data Fig. 9a, b). To simulate the systemic senescence in normal elderly mice, we used *Ercc1^+/-^* mice, which exhibit rapid senescent characteristics such as hair loss and graying like 20-month-old wild-type mice by the age of 10 months (Fig. 4b). Further measurements of spleen weight and 8-hydroxy-2-deoxyguanosine (8-OHdG) content in the spleen showed that *Ercc1^+/-^* 10m mice had similar 8-OHdG levels and spleen weight as WT 20m mice. Immunofluorescence staining revealed that 10-month-old *Ercc1^+/-^* mice expressed more γH2AX in the spleen, heart, liver, and lungs compared to wild-type mice (Fig. 4c-e). In summary, *Ercc1^+/-^*10m mice exhibited senescent characteristics like naturally aging wild-type mice.

**Fig. 4.**
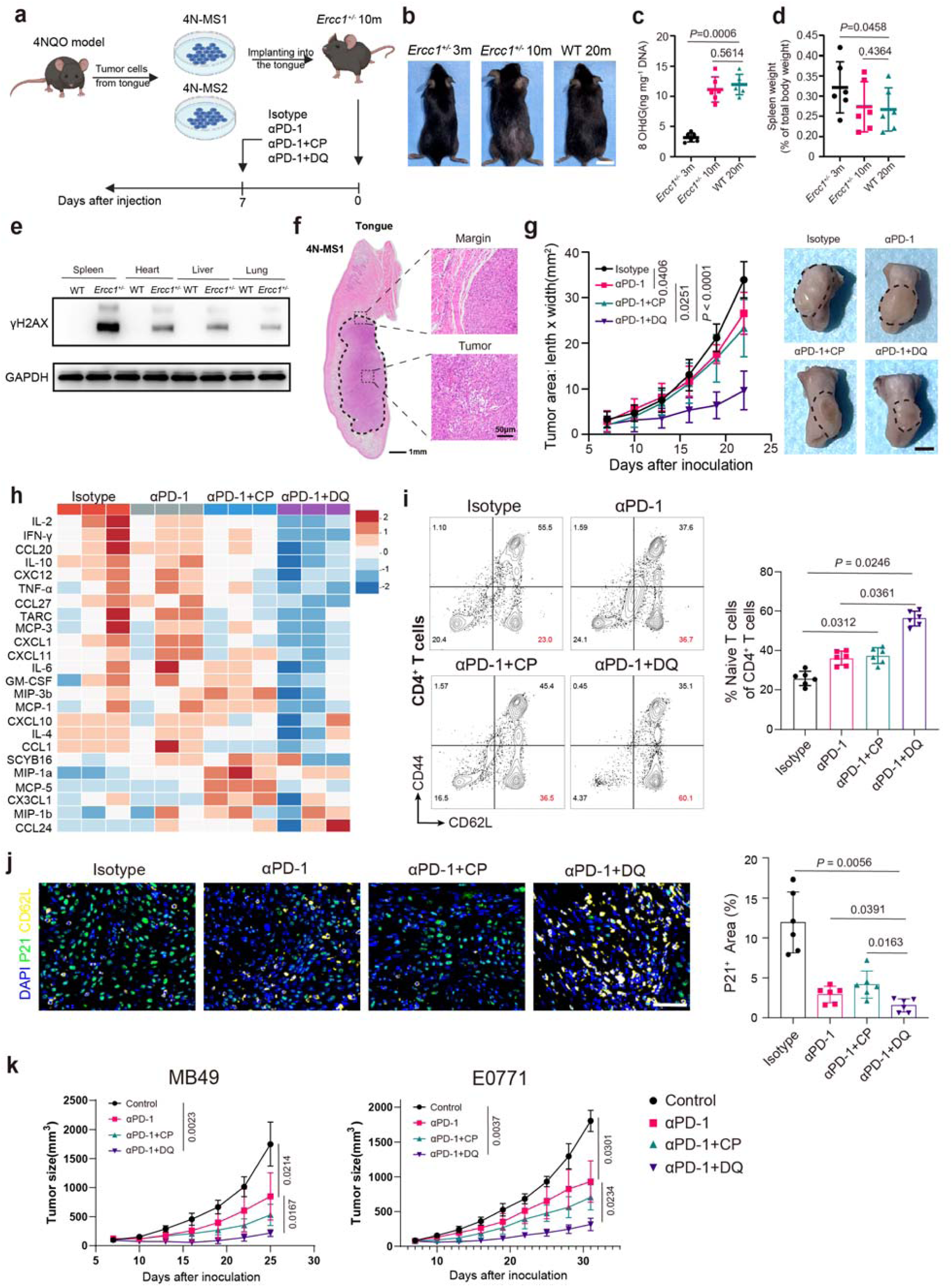
Senolytics combined αPD-1 improve the efficacy of solid tumors in *Ercc1^+/-^* transgenic aging models by reducing SASP and increasing tumor-infiltrating CD4^+^ naïve T cells. **a,** Schematic diagram of orthotopic transplantation of tumor cells derived from the 4-NQO model into the tongues of *Ercc1^+/-^* transgenic mice. **b,** Representative photos of 3-month-old *Ercc1^+/-^* mice (*Ercc1^+/-^* 3m), 9-month-old *Ercc1^+/-^* mice (*Ercc1^+/-^* 9m), and 20-month-old wild type (WT 20m) C57 mice. **c,** Levels of 8-OHdG in spleens from *Ercc1^+/-^* 3m, *Ercc1^+/-^* 9m and WT 20m mice (n=6 per group). **d,** Splenic weights normalized to body weight of *Ercc1^+/-^* 3m, *Ercc1^+/-^* 9m and WT 20m mice (n=6 per group). **e,** Immunoblot detection of γH2AX in spleen, heart, liver and lung from 10-month-old WT or 10-month-old *Ercc1^+/-^* (*Ercc1^+/-^*) mice. **f,** Representative images of H&E staining for 4N-MS1 tongue orthotopic transplant tumors. **g,** C57 mice were implanted with 5×105 4N-MS1 cells into the tongue. Administration of different drug treatments around the 7th day after 4N-MS1 implantation. Tumor growth curve showing the tumor size after receiving Isotype, αPD-1, αPD-1+CP or αPD-1+DQ treatment (n= 6 per group). **h,** Multiplex bead-based protein analysis of lysate of mouse tongue tumor after receiving Isotype, αPD-1, αPD-1+CP or αPD-1+DQ treatment (n= 3 per group). **i**, Flow cytometry analysis of CD4^+^ Naïve T cells proportions in mouse tumors after receiving Isotype, αPD-1, αPD-1+CP or αPD-1+DQ treatment (n= 6 per group). **j,** Representative images and quantitative analysis of P21 and CD62L staining in 4N-MS1 tongue orthotopic transplant tumors. (n= 6 per group). **k,** Tumor growth curves for MB49 and E0771 transplant models after receiving Isotype, αPD-1, αPD-1+CP or αPD-1+DQ treatment (n= 6 per group). The statistical analysis was performed using one-way ANOVA (c, d, h, j) and log-rank test (g, k). *P* values are shown, and error bars indicate the mean ± SEM.

We implanted 4N-MS1cells into the tongues of 10-month-old *Ercc1^+/-^*mice and, on the 5th day, treated the mice with Isotype, αPD-1, αPD-1+CP, or αPD-1+DQ (Fig. 4f). Tumor growth curves showed that mice in the αPD-1+DQ group had better treatment outcomes than other groups (Fig. 4g). Additionally, we employed the Multiplex bead-based protein analysis method to measure the expression of various SASP factors in tumor tissue lysates. The results revealed that αPD-1+DQ treatment significantly reduced the expression of various SASP factors within the tumor, including IL-2, TNF-α, IFN-γ, IL-10, CCL20, MCP-3, and others (Fig. 4h and Supplementary Table 5). Furthermore, flow cytometric analysis showed an increase in CD62L^+^CD44^-^CD4^+^ Naïve cells in the αPD-1+DQ group compared to the Isotype group (Fig. 4i). Consistently, immunofluorescence staining confirmed a significant decrease of P21^+^ cells in the αPD-1+DQ group compared to the other three groups (Fig. 4j). Moreover, we implanted MB49 (bladder cancer) and E0771 (breast cancer) tumors subcutaneously in 10-month-old *Ercc1^+/-^* mice and observed therapeutic outcomes like those seen with the 4N-MS1 transplant model. (Fig. 4k and Extended Data Fig. 9c-f). In summary, these data suggest that αPD-1+DQ treatment can suppress tumor growth in multiple solid tumors while enhancing immune microenvironment vitality through reduced SASP and increased CD4^+^ Naïve cells.

### Senolytics reduce senescent characteristics and enhance activation of tumor-infiltrating CD4^+^ Naïve T cells

To investigate the unique functionality of significantly increased CD4^+^CD62L^+^ Naïve T cells in tumors after combined αPD-1+DQ therapy in anti-tumor immunity, we isolated CD62L^+^CD44^-^CD4^+^ cells from tumors of mice treated with Isotype, αPD-1, or αPD-1+DQ, and performed RNA-Seq and ATAC-Seq (Fig. 5a). The results of RNA-Seq showed that, compared to Isotype or αPD-1 treatment, the αPD-1+DQ group exhibited significantly higher expression of genes related to T-cell migration (*Ccr7*) or antigen response (*Lck*, *Cfd*) in Naïve T cells (Fig. 5b, Extended Data Fig. 10a and Supplementary Table 6). GO analysis revealed enrichment in pathways related to T-cell maintenance, differentiation, and antigen presentation for both αPD- 1+DQ and Isotype groups, indicating that the combination of senolytics with αPD-1 may enhance immune response levels by promoting proliferation and differentiation of Naïve T cells (Fig. 5c and Extended Data Fig. 10b). Additionally, ATAC-Seq analysis demonstrated that, relative to Isotype or αPD-1 treatment, the αPD-1+DQ group exhibited significantly higher chromatin accessibility of the *Ccr7* gene, crucial for adaptive immune responses. Conversely, genes associated with immunosenescence and chronic inflammation, such as *S100a11*, showed lower chromatin accessibility (Fig. 5d, Extended Data Fig. 10c-g and Supplementary Table 7). Furthermore, joint GO analysis of RNA-Seq and ATAC-Seq results from the αPD-1+DQ and Isotype groups yielded similar findings (Fig. 5e). qPCR measurements confirmed that αPD-1+DQ treatment upregulated genes associated with T cell activation, such as *Ccr7*, *Lck*, and *Cfd*, and downregulated genes linked to immunosenescence, including *p16*, *p21*, and *S100a11*, in tumor-infiltrating CD4^+^ Naïve T cells. (Fig. 5f). Additionally, flow cytometry and immunofluorescence staining also demonstrated a significant increase in CCR7 and LCK- positive CD4^+^ Naïve T cells following αPD-1+DQ treatment (Fig. 5g, h). In conclusion, our results indicate that αPD-1+DQ treatment effectively reduces the senescent phenotype while markedly enhancing the quantity and differentiation potential of tumor-infiltrating CD4^+^ Naïve T cells.

**Fig. 5.**
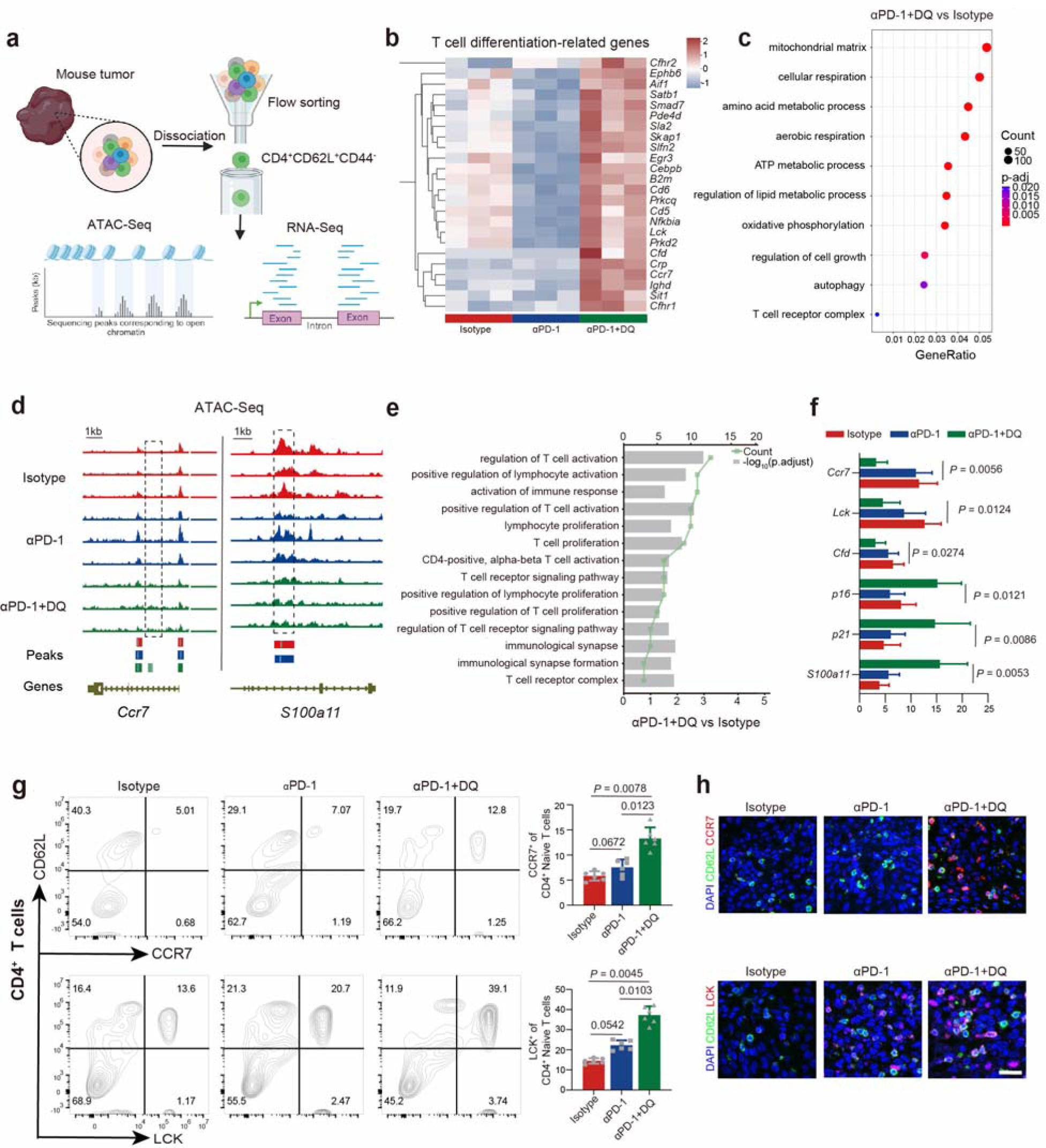
ATAC-Seq and RNA-Seq analyses of CD4^+^ naïve T cells provide evidence for the potential of αPD-1+DQ in the regulation of immunosenescence. **a,** Pattern diagram of cell sorting, as well as combined sequencing with ATAC-Seq and RNA-Seq. **b,** Heatmap illustrating differences in the expression levels of T cell di-related genes among different groups. **c,** DEGs (Log_2_Foldchages > 0.5) of CD4 naïve T cells in tumors between αPD-1 and DQ and isotype groups enriched GO terms. **d,** ATAC-seq tracks showing the representative genes chromatin accessibility in the *Ccr7* and *S100a11* loci for CD4 naïve T cells in mouse tumors from isotype, αPD-1 and DQ, and αPD-1 groups. **e,** RNA-seq and ATAC-seq detected GO terms that were simultaneously enriched in the αPD-1 and DQ and isotype groups. **f,** Expression of *Ccr7*, *Lck*, *Cfd*, *S100a11*, *p16^Ink4a^*, *p21^Cip1^*in CD4^+^ naïve T cells in mouse tumors after receiving Isotype, αPD-1 or αPD-1+DQ treatment (n= 6 per group). **g,** Flow cytometry analysis of CCR7 and LCK in CD4^+^ Naïve T cells proportions in mouse tumors after receiving Isotype, αPD-1 or αPD- 1+DQ treatment (n= 6 per group). **h,** Representative images of CCR7 and LCK staining in 4N- MS1 tongue orthotopic transplant tumors. The statistical analysis was performed using one-way ANOVA (**f**, **g**). *P* values are shown, and error bars indicate the mean ± sem.

### Combination of anti-PD-1 with dasatinib and quercetin demonstrates safety and efficacy in patients with HNSCC

Previous studies have indicated that neoadjuvant chemotherapy reduces recurrence in patients with locally advanced oral cancer but does not improve survival rates ^27,28^. Recent research indicates that chemotherapy can exacerbate the adverse effects associated with immunotherapy. The most prevalent adverse effects of neoadjuvant immunochemotherapy include alopecia (100.0%), nausea/vomiting (60.4%), and fatigue (50.0%) ^29^. Therefore, it is essential to investigate novel therapeutic agents that can enhance the efficacy of immunotherapy while minimizing systemic side effects, preserving the function of vital organs, and ultimately improving the quality of life for patients.

Based on our findings from single-cell multi-omics analysis and animal models, we hypothesized that combining ICIs with dasatinib and quercetin would be both effective and safe for patients with HNSCC, significantly reducing TRAEs compared to standard neoadjuvant chemotherapy. To evaluate this hypothesis, we conducted a prospective, open-label, single-center, Phase II clinical trial (COIS-01), assessing the neoadjuvant efficacy of tislelizumab combined with dasatinib and quercetin in resectable HNSCC patients. The primary endpoint of the study was the MPR rate (Fig. 6a). Between February 5, 2023, and April 5, 2024, a total of 24 patients were enrolled according to the study protocol. Efficacy evaluations revealed that 4 patients (16.7%) achieved a pCR, while another 4 (16.7%) achieved a MPR, resulting in an overall treatment response rate of 33.3% (pCR + MPR) (Fig. 6b, Table 3 and Supplementary Table 8). Representative imaging, pathological assessments, and clinical photographs of patients achieving pCR are provided (Fig. 6c).

**Fig. 6.**
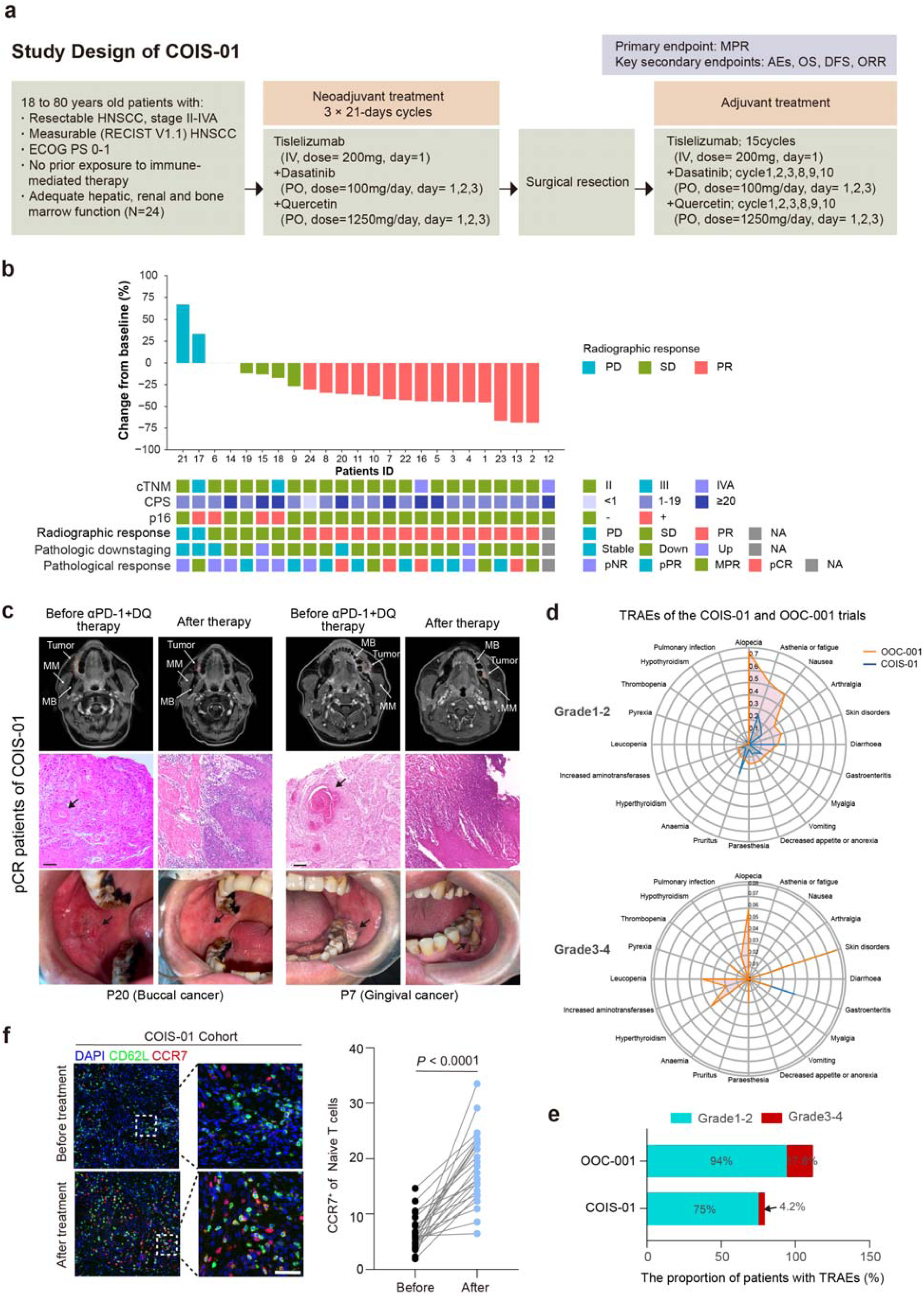
The Phase II clinical trial COIS-01 validated the safety and efficacy of combining senolytic drugs with anti-PD-1 therapy. **a,** Schematic representation of the clinical trial (COIS- 01) investigating the combination of ICIs with senolytics as neoadjuvant therapy for HNSCC. **b,** The waterfall chart displays the clinical characteristics of 24 patients in the COIS-01 trial. **c,** Representative imaging, HE pathology, and intraoral photographs of two pCR patients from COIS-01. MM represents the masseter muscle, MB represents the mandible, and the tumor area is indicated by the circled region. Scale bar, 50 μm. **d,** The radar chart shows the proportions of various types of TRAEs in two clinical trials. **e,** Comparison of TRAEs between the COIS-01 and OOC-001 clinical trials. **f,** Representative images and quantifications of immunofluorescent staining for CCR7^+^ Naïve T cells before and after treatment (n= 24 per group). The statistical analysis was performed using t test (**f**). *P* values are shown, and error bars indicate the mean ± sem.

**Table 3.**
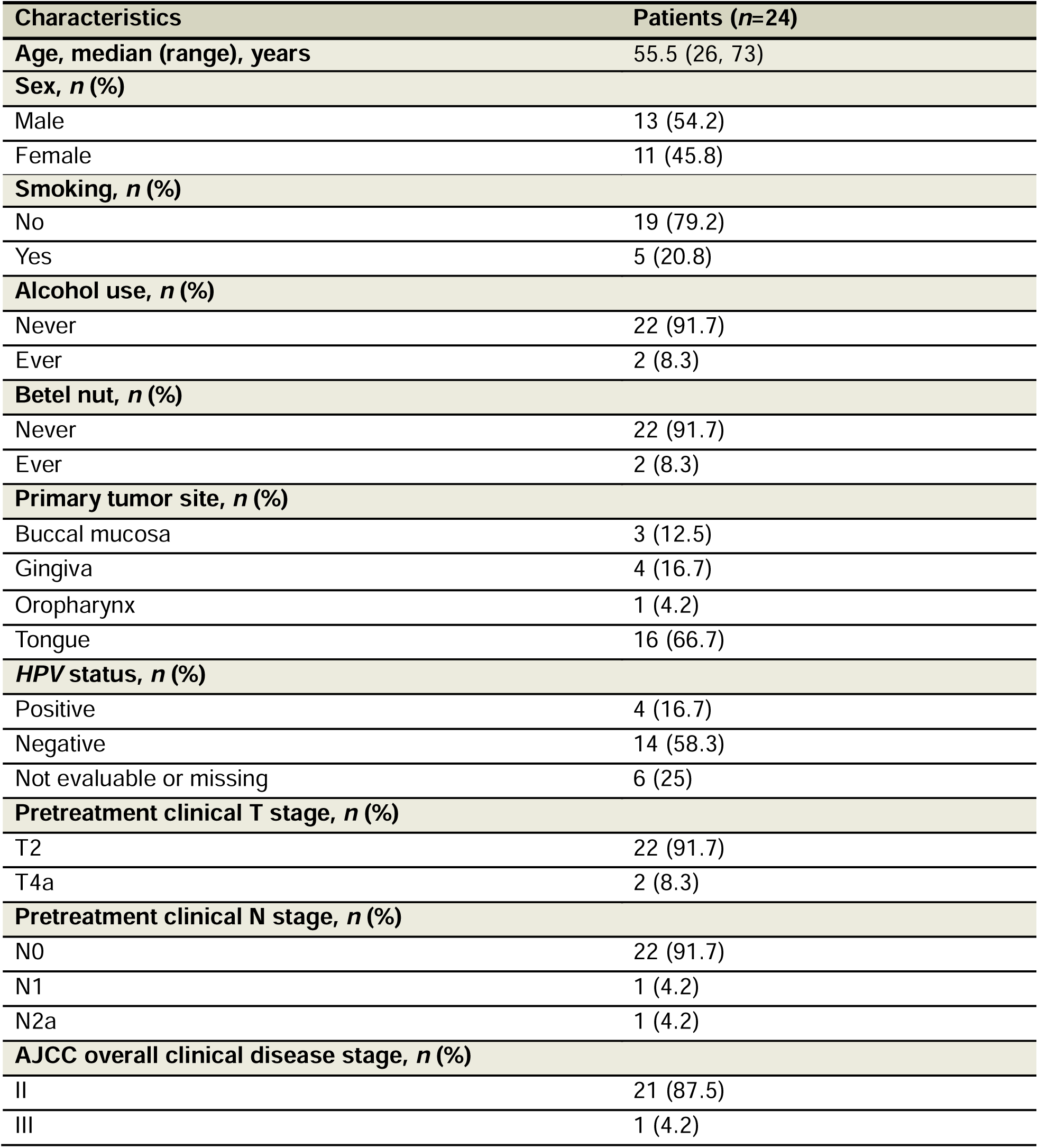
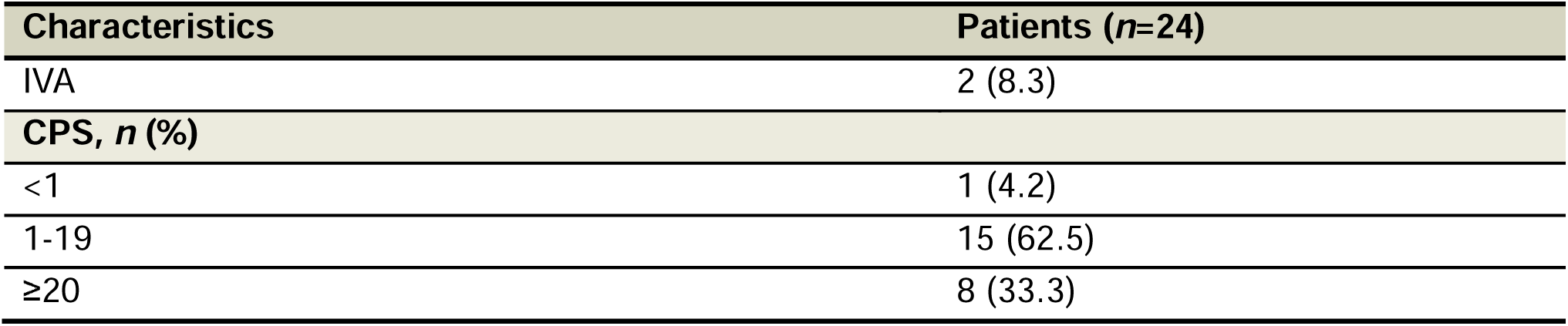
Demographic and clinical characteristics of COIS-01.

Safety analyses of the 24 patients demonstrated that only 1 patient experienced grade 3-4 TRAEs (Fig. 6d). In contrast, the OOC-001 trial, which evaluated neoadjuvant chemotherapy, showed the most common TRAEs were alopecia (75%), asthenia or fatigue (53%), and nausea (49%) (Fig. 6e, Table 2). In the COIS-01 trial, the corresponding incidences were notably lower, with 0% for alopecia, 25% for asthenia or fatigue, and 16.7% for nausea (Table 4 and Supplementary Table 9). Although the survival period in COIS-01 has not yet allowed for statistical analysis of overall survival, no recurrences or deaths have been observed to date. Furthermore, immunohistochemical analysis of patient paraffin sections before and after αPD-1 + DQ treatment revealed an upregulation of CCR7 expression in naïve T cells post-treatment (Fig. 6f). These early findings from the COIS-01 trial suggest that the combination of tislelizumab with dasatinib and quercetin demonstrates promising antitumor efficacy, favorable tolerability, and a potential to mitigate immunosenescence-related features.

**Table 4.**
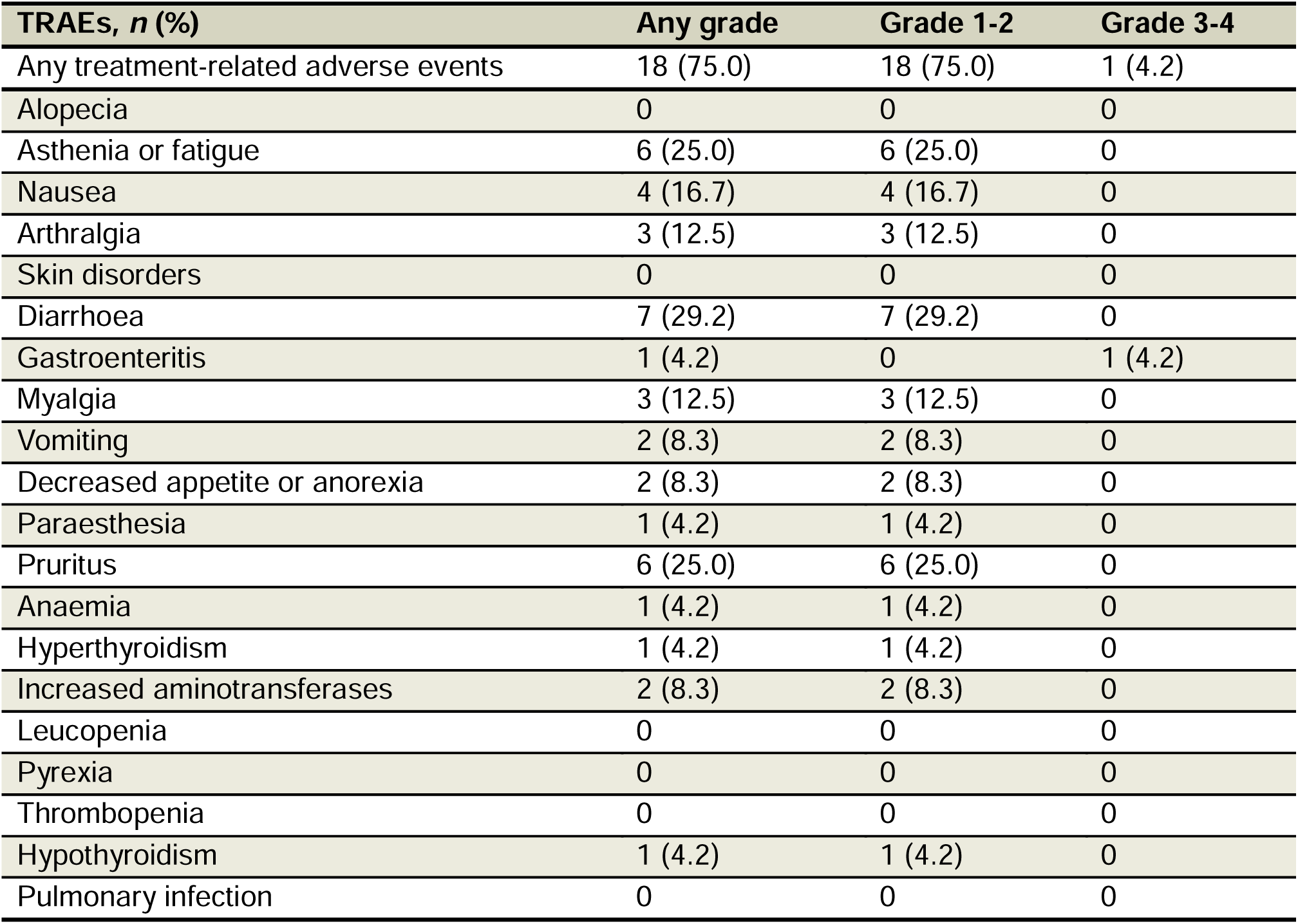
Treatment-related adverse events of COIS-01 (*n*=24)

| TRAEs, n (%) | Any grade | Grade 1-2 | Grade 3-4 |
| --- | --- | --- | --- |
| Any treatment-related adverse events | 18 (75.0) | 18 (75.0) | 1 (4.2) |
| Alopecia | 0 | 0 | 0 |
| Asthenia or fatigue | 6 (25.0) | 6 (25.0) | 0 |
| Nausea | 4 (16.7) | 4 (16.7) | 0 |
| Arthralgia | 3 (12.5) | 3 (12.5) | 0 |
| Skin disorders | 0 | 0 | 0 |
| Diarrhoea | 7 (29.2) | 7 (29.2) | 0 |
| Gastroenteritis | 1 (4.2) | 0 | 1 (4.2) |
| Myalgia | 3 (12.5) | 3 (12.5) | 0 |
| Vomiting | 2 (8.3) | 2 (8.3) | 0 |
| Decreased appetite or anorexia | 2 (8.3) | 2 (8.3) | 0 |
| Paraesthesia | 1 (4.2) | 1 (4.2) | 0 |
| Pruritus | 6 (25.0) | 6 (25.0) | 0 |
| Anaemia | 1 (4.2) | 1 (4.2) | 0 |
| Hyperthyroidism | 1 (4.2) | 1 (4.2) | 0 |
| Increased aminotransferases | 2 (8.3) | 2 (8.3) | 0 |
| Leucopenia | 0 | 0 | 0 |
| Pyrexia | 0 | 0 | 0 |
| Thrombopenia | 0 | 0 | 0 |
| Hypothyroidism | 1 (4.2) | 1 (4.2) | 0 |
| Pulmonary infection | 0 | 0 | 0 |

## Discussion

The central conclusion of this study is that the senescence of TIME is a critical factor affecting the efficacy of tumor immunotherapy. The first evidence supporting this conclusion comes from the multidimensional single-cell data analysis of immune cells from both blood and tumors of patients in clinical trials. We observed that noMPR patients exhibited more pronounced immunosenescence features, including a decreased proportion of naïve T cells and memory B cells, an increased proportion of immunosuppressive cell subtypes, and a reduction in the TCR/BCR repertoire. Additionally, we identified IAGs in immune cells from noMPR patients, which showed higher expression levels across various T and B cell subsets. When tumor-bearing mice received a combination of ICI and senolytics, we observed an increase in CD4^+^ naïve T cells and a reduction in senescence markers in immune cells within the tumors. This led to reduced tumor burden and extended survival in the mice. A clinical trial based on our findings further confirmed the potential advantage of combining ICI with senolytics for treating solid tumors.

Immune cells are the cornerstone of tumor immunotherapy ^30,31^. T cell-centered therapies have become powerful tools against cancer. ICI is the most widely used immunotherapy method for solid malignancies, with an increasing number of monoclonal antibodies targeting different inhibitory receptors entering clinical practice ^32^. Antibodies against PD-1, CTLA-4, and LAG-3 have been FDA-approved for several types of solid tumors. Although ICI can induce objective responses even in the challenging context of metastatic disease, only a minority of patients (estimated at <30%) achieve sustained and/or complete clinical responses ^33–35^. The TIME is rich in tumor-reactive T cells, but exhaustion remains a major factor limiting their antitumor capabilities. Studies on chronic antigen stimulation models in mice ^36–38^ and the characteristics of tumor-infiltrating lymphocytes (TILs) in human “hot” tumors ^39–41^ emphasize that antitumor T cells can progressively acquire a dysfunctional state, allowing tumor immune evasion ^42,43^. These findings suggest that the TIME is not isolated but interconnected with the systemic immune system. The dynamic processes of TILs dysfunction and continuous recruitment of immune cells to the tumor provide a new perspective for exploring ways to improve the TIME during tumor progression.

Notably, our study shows no significant differences in the proportions of most immune cell types in the peripheral blood of responsive and non-responsive patients before and after treatment. This indicates that predicting immunotherapy responses based on peripheral blood tests may be challenging. Although cellular senescence drives various age-related complications through SASP, identifying senescent immune cells in vivo remains difficult ^44–46^. Recent studies have identified a set of genes (SenMayo) enriched in bone biopsies from elderly individuals, capable of identifying senescent hematopoietic or stromal cells at the single-cell level in human and mouse bone marrow/bone scRNA-Seq data ^46^. Researchers also established AgeAnno, a human aging single-cell annotation knowledge base, offering dynamic functional annotations of 1,678,610 cells from 28 healthy tissue samples ^47^. Currently, there are no specific markers or gene sets dedicated to reflecting the degree of immunosenescence. We have constructed an immunosenescence-related gene set comprising 154 genes (IAGs), which has effectively distinguished between young and elderly individuals in external datasets. This provides a powerful tool for future research on the senescent status of the immune system. It is important to note that the immune system consists of diverse cell types, each with distinct functions. For example, the gene expression profiles of senescent states in T cells, dendritic cells, and macrophages may differ significantly. Understanding these differences is crucial for developing targeted therapies for senescent phenotypes.

Senolytics therapies for various cancers are currently under investigation. Studies have shown that senolytics ABT-263 can exert antitumor effects by selectively clearing senescent cells in obese patients’ tumor tissues ^48^. Additionally, the plant compound Rutin, when combined with chemotherapy, can intervene in tumor development ^49^. These studies suggest that senolytics may offer unique advantages in cancer treatment, with fewer side effects and promoting overall systemic health. However, the combination of senolytics and ICI has not been explored in TIME. Previous research discussed the potential benefits of DQ therapy, a novel senolytics regimen that disrupts survival pathways of senescent cells and selectively targets them ^50–52^. Recent research on systemic administration of DQ for treating intervertebral disc degeneration has provided new insights, highlighting the tissue-specific effects of DQ and the importance of treatment timing ^53^. Additionally, a clinical trial on DQ treatment for idiopathic pulmonary fibrosis (IPF) demonstrated significant improvements in patients’ physical function ^54,55^. Our COIS-01 trial has demonstrated the efficacy and safety of combining senolytics with ICIs in HNSCC patients. Previous studies have shown that ICIs monotherapy in HNSCC has limited effectiveness, with reported MPR rates of 7-13.9% ^56–58^. In contrast, the COIS-01 trial achieved an MPR rate of 33.3%. While immunochemotherapy can achieve higher MPR rates, it is associated with significant toxicity, with grade ≥ 3 adverse events occurring in up to 35% of patients ^59^, substantially higher than in the COIS-01 trial, where the incidence of such events was considerably lower (4.2%). Grade ≥3 toxicities can severely impact patient quality of life and pose challenges for subsequent adjuvant immunotherapy. Discontinuation of adjuvant immunotherapy due to toxicity may compromise the overall therapeutic efficacy, potentially affecting survival outcomes ^60^.

Given this balance between efficacy and risk, the COIS-01 trial offers a novel therapeutic strategy that maximizes the antitumor efficacy of ICIs while minimizing adverse events. Preclinical studies in multiple mouse models have demonstrated the superior potential for long-term survival with the combination of αPD-1 and senolytics (dasatinib and quercetin) compared to monoimmunotherapy or immunochemotherapy. Consistently, the combination of senolytics and anti-PD-1 therapy has been shown to dramatically upregulate CCR7^+^ naïve T cells in COIS-01 patients, a critical feature in the modulation of immunosenescence ^61^. This upregulation enhances the homing ^62^, immune surveillance ^63^, and circulating maintenance ^64^ of these specific T cells, potentially predicting improved survival outcomes in clinical settings. However, rigorous randomized clinical trials are needed to further validate these findings. Additionally, further investigation is required to determine the optimal type and dosage of senolytics in combination with ICIs. In summary, this study provides valuable insights into the variability of TIME across individuals and highlights the potential to enhance antitumor efficacy by targeting immunosenescence.

## Acknowledgments

We thank all patients and their families for participation during sample collection. We are grateful to the support of the Clinical Research Design Division and Clinical Research Center at Sun Yat-sen Memorial Hospital, Sun Yat-sen University. The co-author Dr. Soldano Ferrone sadly passed away on January 10, 2023, at the age of 82, after an 8-week-long battle with COVID-19. He was the supervisor of Dr. Song Fan during Dr. Fan’s post-doctoral work in his lab at Massachusetts General Hospital (MGH). Due to his constantly invaluable guidance and suggestions, which greatly helped improve and organize this study, all contributing authors agree to keep him as a co-author of this manuscript. We thank all the members of the Fan lab for valuable discussions and help with experimental techniques and analysis of the manuscript. This work was supported by the Joint Funds of the National Natural Science Foundation of China (U21A20381), the General Funds of the National Natural Science Foundation of China (82373452), the Guangdong Natural Science Funds for Distinguished Young Scholar (2022B1515020061), the Guangdong Basic and Applied Basic Research Foundation (2021A1515220138), the Guangzhou Basic Research Program Jointly Funded by Municipal Schools (Institutes) (202201020367), the Fundamental Research Funds for the Central Universities, Sun Yat-sen University (16ykpy10), the Fundamental Research Funds for the Central Universities, Sun Yat-sen University (19ykzd20), the General Funds of the National Natural Science Foundation of China (32071451), the Guangdong Provincial Pearl River Talents Program (2021QN02Y747), the Shenzhen Science and Technology Program (RCYX20210706092100003), and by the Shenzhen Medical Research Funds grant (A2303005).

## Author contributions

X.W., S.F., X.F., F.X., N.L., J.W. and E.D. designed the study and wrote the manuscript, S.F., X.F., F.X. and X.W. supervised the study. Single-cell RNA and T/BCR sequencing, RNA-Seq and ATAC-Seq involves library preparation and data analysis by N.L., J.W., E.D., T.C, Q.L. and J.Z., B.W., J.W., E.D., X.D., S.F., T.C and Y.Z. collected patient samples and analyzed clinical data. S.F., N.L., J.W. and E.D. performed experiments and interpreted the data. All authors critically revised the paper.

## Methods

### Patients

The inclusion criteria and exclusion criteria for OOC-001 and COIS-01 are the same.

Eligibility criteria included: age 18-80 years; newly diagnosed histologically or cytologically confirmed HNSCC; stage II-IVA AJCC 8th edition; disease was determined resectable by the treating head and neck surgeon; Eastern Cooperative Oncology Group (ECOG) performance status ≤ 1; and adequate organ function.

Key exclusion criteria included: prior therapy with immunotherapy or or have had invasive malignancies and related treatments in the past 5 years; autoimmune diseases requiring recent systemic steroid use, active infections, or a history of allergic reactions to similar compounds; pregnancy, psychiatric disorders, HIV, active Hepatitis B or C.

### Trial design and treatments

#### OOC-001 (NCT04718415)

This study is a prospective, single-arm, phase II clinical study. This study plans to enroll 51 patients with resectable oral or oropharyngeal squamous cell carcinoma, and preoperatively use sintilimab, carboplatin, and albumin-bound paclitaxel. Tumor tissue and paracancerous tissue of patients will be collected to observe the changes of imaging and pathology before and after treatment. At the same time, clinical information of patients, such as pathological grade, stage, treatment, prognosis, serology, imaging, etc., will be collected to evaluate the safety and feasibility of sintilimab combined with carboplatin and albumin-bound paclitaxel for neoadjuvant treatment of resectable oral and oropharyngeal squamous cell carcinoma.

Patients received treatment with sintilimab, nab-paclitaxel, and carboplatin for up to 2 to 4 cycles: Sintilimab (IV), dose: 200 mg, day: 1, cycle length: 21 days; Carboplatin (IV), dose: 300 mg/m², day: 1, cycle length: 21 days; Nab-paclitaxel (IV), dose: 260 mg/m², day: 1, cycle length: 21 days. Surgical therapy will be performed at the discretion of the treating surgeon according to standard of care.

#### COIS-01 (NCT05724329)

This study is a prospective, open-label, single-center, phase II clinical study evaluating the safety and efficacy of the combination of tislelizumab with dasatinib and quercetin in the new adjuvant treatment of resectable head and neck squamous cell carcinoma. This research aims to expand the indications for new adjuvant immunotherapy in solid tumors and provide a new strategy for significantly improving the prognosis of head and neck squamous cell carcinoma patients. This study plans to enroll 24 participants with resectable head and neck squamous cell carcinoma. We will collect tumor tissues, adjacent tissues, fat, epidermis, whole blood samples, saliva, feces, and urine from the patients. The study aims to observe imaging and pathological changes before and after treatment while also collecting clinical information, such as pathological grading, staging, treatment details, prognosis, and serological and imaging data. Patients will receive neoadjuvant treatment with tislelizumab, dasatinib, and quercetin for up to 3 cycles (cycle length: 21 days): Tislelizumab (IV), dose: 200 mg, day: 1; Dasatinib (PO), dose: 100 mg/day, days: 1, 2, 3; Quercetin (PO), dose: 1250 mg/day, days: 1, 2, 3. Surgical therapy will be performed at the discretion of the treating surgeon according to standard of care. Following surgical resection, participants will receive adjuvant therapy: Tislelizumab (200 mg, day: 1) for cycles 1 to 15; Dasatinib (100 mg/day, days: 1, 2, 3) for cycles 1, 2, 3, 8, 9, and 10; Quercetin (1250 mg/day, days: 1, 2, 3) for cycles 1, 2, 3, 8, 9, and 10.

### Ethics statement

This study includes two independent clinical trials for HNSCC: OOC-001 (NCT04718415) and COIS-01 (NCT05724329). Eligible HNSCC patients received prior approval from the Institutional Review Board (IRB) at Sun Yat-sen Memorial Hospital, Sun Yat-sen University, and were required to provide written informed consent to participate in the study. All patients were diagnosed with HNSCC by expert histopathologists, and all invasive procedures and systemic treatments were ordered by the patients’ attending physicians, individually determined based on the response after the administration of pharmacological drugs. All images of research participants in this article have obtained consent for publication.

### End points

#### OOC-001

The primary endpoint were Safety, defined as the severity of treatment-related adverse events will be graded according to NCI CTCAE (version 5.0) during the study and follow-up; the major pathological response rate, defined as the proportion of participants with viable tumor cells ≤10% in the resected specimens. Secondary endpoints were Disease-Free Survival, defined as the time from treatment until the date of the first relapse (local/regional recurrence or distant metastasis) or death (from any cause) whichever comes firsts; Overall Survival, defined as the time from day 1 of study treatment until death from any cause; Objective Response Rate (ORR), defined as assessed through imaging according to the Response Evaluation Criteria in Solid Tumors (RECIST 1.1); Surgery Delay Rate, defined as participants who cannot undergo surgery within 8 weeks after the last dose will be classified as having surgery delay.

#### COIS-01

The primary endpoint was the major pathological response rate, defined as the proportion of participants with viable tumor cells ≤10% in the resected specimens. Secondary endpoints were Safety, defined as the severity of treatment-related adverse events will be graded according to NCI CTCAE (version 5.0) during the study and follow-up; Overall Survival, defined as time from enrollment to the date of death from any cause; Disease-Free Survival, defined as the time from treatment until the date of the first relapse (local/regional recurrence or distant metastasis) or death (from any cause) whichever comes firsts; Objective Response Rate (ORR), defined as assessed through imaging according to the Response Evaluation Criteria in Solid Tumors (RECIST 1.1); Surgery Delay Rate, defined as participants who cannot undergo surgery within 8 weeks after the last dose will be classified as having surgery delay.

### Statistical analysis of clinical trials

#### OOC-001

All enrolled cases that have received at least one dose of the investigational drug and have safety records post-treatment will be included in the Safety Analysis Set. Effectiveness analysis will be conducted for all enrolled cases that received neoadjuvant therapy in conjunction with surgery, biopsy, or chemoradiotherapy.

This is a single-arm, pilot study with a fixed sample size. No strict hypothesis testing will be conducted, and the study plans to enroll 51 participants. The MPR rate and 95% CI were calculated using the Clopper-Pearson method. The OS, and DFS were plotted using the Kaplan-Meier method. Other clinical outcomes, demographic characteristics, and safety were summarized descriptively. We performed all statistical tests using GraphPad Prism 9.0.0.

#### COIS-01

Effectiveness analysis will be conducted for all enrolled cases that received at least one dose of the drug according to the Intent-to-Treat (ITT) principle. All enrolled cases that have received at least one dose of the investigational drug and have safety records post-treatment will be included in the Safety Analysis Set.

Based on a two-sided type 1 error of 0.05 and a power of 90%, we estimated that a total sample size of 24 patients would be required to show an improvement of 23% in the major pathological response rate after DQ (ie, from 7% in the previous study ^58^ to 30% in the current study). Assuming a 5% dropout rate, the final sample size was estimated to comprise 24 patients.

The MPR rate and 95% CI were calculated using the Clopper-Pearson method. The OS, and DFS were plotted using the Kaplan-Meier method. Other clinical outcomes, demographic characteristics, and safety were summarized descriptively. We performed all statistical tests using GraphPad Prism 9.0.0.

### Human specimens

The eight patients used for the scRNA-Seq study were part of the OOC-001 clinical trial and received the described treatment regimen. Specifically, all patients had their primary tumor tissue and peripheral blood collected through biopsy at the time of enrollment, and the peripheral blood was collected after sintilimab in combination with carboplatin and nab-paclitaxel treatment before surgery. Among the 7 patients (SH1-SH7) who underwent surgery, 3 patients are MPR, and 4 patients are noMPR. Paired biopsy samples of primary tumor tissue and peripheral blood were collected at baseline before treatment, and peripheral blood samples were collected 3 weeks after the first dosing cycle. Except for SH1, peripheral blood samples were collected from the other patients before and after the first medication cycle for matching. For patient SH1, blood samples were collected only before the first medication cycle. Peripheral blood samples were collected after the second medication cycle for SH1, SH2, SH3, and SH4, and peripheral blood samples were collected for SH3, SH4, and SH7 after the third medication cycle. Paraffin sections from two ongoing clinical study patient cohorts were used for immunofluorescence staining of senescent markers P16 and P21. This included 33 MPR patients and 17 noMPR patients from OOC-001, as well as 37 MPR patients and 24 noMPR patients from REDUCTION-I. Another set from COIS-01 collected paraffin sections from 7 patients for immunofluorescence staining before and after treatment.

### Sample collection and processing

Peripheral blood mononuclear cells (PBMCs) were isolated according to the manufacturer’s protocol. Briefly, before the start of treatment, 5 mL of fresh peripheral blood was collected in EDTA anticoagulated tubes and subsequently extracted with Lymphoprep™ (Stem Cells) solution. After centrifugation, carefully transfer lymphocytes to a new 50 mL tube and wash with PBS. Next, incubate the lymphocytes for 10 minutes to lyse the blood cells. Finally, cells were resuspended in sorting buffer containing PBS plus 2% FBS.

For tumor biopsy samples, cut tissue with a diameter of about 5 mm at the junction of the tumor and normal tissue as a representative sample. All 7 tumor biopsy samples were collected as a discovery cohort through routine clinical practice and then stored in MACS Tissue Storage Solution (Miltenyi). Fresh biopsy samples of oral HNSCC were minced and isolated using a human tumor isolation kit (Miltenyi), followed by enzymatic digestion on a rotor at 37°C for 45– 60 min. The detached cells were then passed through 70 μm and 40 μm cell filters (Biosharp) and centrifuged at 300 × g for 10 min. Pelleted cells were suspended in red blood cell lysis buffer (Thermo-Fisher) and incubated on ice for 2 min, then washed and resuspended in phosphate buffer supplemented with 2% fetal bovine serum (FBS, ExCell) PBS twice in saline (PBS, Gibco). The viability of all samples was confirmed to be >90% using the trypan blue (Thermo-Fisher) exclusion method.

### Single-cell RNA-seq library and TCR-seq library preparation and sequencing

According to the manufacturer’s protocol, scRNA-seq libraries were constructed. In summary, the density of cells was determined after washing once with PBS containing 0.04% bovine serum albumin (BSA, Invitrogen). Next, 2×10^5^ cells were loaded on a 10x Genomics GemCode Single cell instrument (10x Genomics) that generates single-cell Gel Bead-In-EMulsion (GEMs). Libraries were generated and sequenced from the cDNAs with the Chromium Next GEM Automated Single Cell 3’ cDNA Kitv3.1 (10x Genomics). Upon dissolution of the Gel Bead in a GEM, primers were released and mixed with cell lysate and Master Mix. Barcoded, full length cDNAs were then reverse-transcribed from polyadenylated mRNA. Full-length, barcoded cDNAs were then amplified by quantitative real-time PCR (qRT PCR) for library construction. Finally, single-cell RNA libraries were sequenced by an Illumina HiSeq X Ten sequencer with 150 bp paired-end reads.

After reverse transcription and cell barcoding, the cDNA was de-emulsified and purified using silane magnetic beads, followed by PCR amplification. The amplified cDNA was then used for 5’ gene expression library construction and TCR V(D)J targeted enrichment amplification using the Illumina® bridge. Libraries prepared according to the manufacturer’s user guide were then purified and analyzed for quality assessment. Single-cell RNA and TCR V(D)J libraries were sequenced by an Illumina HiSeq X Ten sequencer using 150 bp paired-end reads.

### Bulk TCR-seq library preparation and sequencing

Whole Blood Samples Blood was collected in EDTA tubes and sent to isolated PBMCs, and DNA was extracted for TCRβ analysis by immunoSEQ ^65^. Briefly, extracted genomic DNA was amplified using a bias-controlled multiplex PCR system, followed by high-throughput sequencing. Raw data processing and analysis were performed by immunoSEQ. Subsequently, we used the International ImMunoGeneTics Information System (IMGT; http://imgt.cines.fr) database for comparison. After the alignment, the relative abundance of each TCRβ CDR3 sequence was clarified and calculated. Furthermore, we use batch correction to eliminate batch effects across different datasets. Subsequently, multiple TCR data statistics were performed.

### Bulk ATAC and RNA isolation and sequencing

Isolation, library construction and sequencing of bulk ATAC and RNA were performed at Gene Denovo Biotechnology Co. (Guangzhou, China). For ATAC sequencing, CD4 naïve cells from C57 mouse tongue tumors were extracted. Tn5 transposase was added to the nuclear suspension. After the reaction was completed, the DNA fragment was purified; the amplified product was then used for PCR amplification. The fragments were purified using AMPure XP magnetic beads (Beckman Coulter, Brea, CA, USA) to construct a sequencing library. After the library construction was completed, Agilent 2100 (Agilent, Santa Clara, CA) was used to detect the quality of the library. Libraries that pass the quality inspection will be used for on-machine sequencing (Novaseq 6000) to obtain sequence information of the open chromatin region fragments to be tested.

Extraction of CD4 naïve cells from C57 mouse tongue tumors. Total RNA was extracted using Trizol reagent kit (Invitrogen) according to the manufacturer’s protocol. RNA quality was assessed on an Agilent 2100 Bioanalyzer (Agilent Technologies). After total RNA was extracted, eukaryotic mRNA was enriched by Oligo(dT) beads. Then the enriched mRNA was reversely transcribed into cDNA by using NEBNext Ultra RNA Library Prep Kit for Illumina (NEB #7530, New England Biolabs). The resulting cDNA library was sequenced using Illumina Novaseq6000 by Gene Denovo Biotechnology Co. (Guangzhou, China).

### scRNA-Seq data processing

The Cell Ranger toolkit (version 3.1.0) provided by 10x Genomics was applied to aggregate raw data, filter low-quality reads, align reads to the human reference genome (GRCh38), assign cell barcodes and generate a UMI matrix. The toolkit Scanpy (version 1.9.3) ^66^ was used to analyze single-cell RNA sequencing (scRNA-Seq) data. Specifically, the original UMI matrix was processed to filter out genes expressed in less than 10 cells and cells with less than 200 genes. We further quantified the number of genes and UMI counts per cell and maintained thresholds for high-quality cells of 1000-50,000 UMIs, 400-6,000 genes and less than 10% mitochondrial gene counts, ensuring that most heterogeneous cell types were Incorporate into downstream analyses. Scrublet (version 0.2.3) ^67^ was then applied to each sequencing library to remove potential doublets, using default parameters. The normalized expression matrix was calculated from raw UMI counts normalized to total counts per cell (library size) and then scaled by 1e4 and log-transformed.

### Dimensionality reduction and unsupervised clustering of scRNA-seq data

Dimensionality reduction and unsupervised clustering were performed according to standard workflows in Scanpy ^66^. Briefly, use highly variable genes (HVG) based on the highly_variable_genes default parameters. Unwanted sources of variation, including total counts, and mitochondrial gene counts were further regressed from the normalized expression matrix. Primary component analysis (PCA) was performed on the variable gene matrix to reduce noise and the top 50 components were used for downstream analysis. To correct for batch effects across different patients, we applied BBKNN to generate a batch-balanced k-nearest neighbor (KNN) map, which identifies the top neighbors of each cell in each batch individually, rather than the entire pool of cells ^67^. The Leiden algorithm is then applied to such nearest neighbor graphs to detect communities and find cell clusters ^68^. It is worth noting that the same principal components are also used for nonlinear dimensionality reduction to generate uniform distributions for visualization in Uniform Manifold Approximation and Projection (UMAP). After the first round of unsupervised clustering, we annotated each cell cluster according to canonical immune cell markers and identified the main immune cell types, including T cells, NK cells, B cells, myeloid cells, fibroblasts, endothelial cells and tumor cells. Use the rank_genes_groups function with parameter method Wilcoxon to detect marker gene.

### TCR sequences assembly

Applying the TCR sequence assembly Cell Ranger toolkit (version 3.1.0) provided by 10x Genomics will perform FASTQ sequence quality filtering, sequence alignment, V(D)J assembly and TCRαβ pairing. Only TCRs containing paired TRAV-CDR3-TRAJ and TRBV-CDR3-TRBJ-TRBC chains were considered valid and retained for downstream analysis. Each cell is assigned a pair of α and β chains with the highest UMI count. Cells with identical TCR pairs are defined as clonal and are thought to originate from a common ancestor.

### Calculation of Diversity Index

Characterize the immune repertoire by examining diversity and clonality. Immune repertoire analysis involves a variety of diversity indices, such as Chao1 index, Clone Diversity Index, Richness index and Shannon index ^24^.

### Bulk RNA-sequencing and ATAC-sequencing data processing

For bulk RNA-Seq data, reads were filtered using FASTQ (0.23.3) default parameters, and then clean reads were aligned to the mouse genome GRCm39 and gencode. vM30 annotations were using STAR (2.7.10b) and using RSEM (1 .3.1) rsem-calculate-expression is in quantitative analysis of gene expression. Difference analysis was done using DESeq2 (1.42.0).

For bulk ATAC data, use FASTQ (0.23.3) for quality control, then use bwa (0.7.17) to align the reads to the reference genome (GRCm39), and peak using Genrich (0.6.1, https://github.com/jsh58/Genrich). Peak is annotated to gencode. vM30 through annotatePeaks.pl of HOMER (4.11). The difference peak is obtained by BEDTools (v2.31.1) subtract. (https://github.com/jsh58/ATAC-seq2)

### Generation of Immunosenescence gene set

Our own senescence-related gene set was generated by combining genes reported to be enriched in senescent immune cells in previous studies and experimentally validated in at least human or mouse cells. We screened 1745 studies, but after removing studies reporting duplicates, case reports, non-human genes, and non-high-throughput sequencing, a list of 6 studies was developed (Senmayo ^46^, GSE157007 ^24^, HRA000624 ^69^, HRA000395 ^70^, HRA000615 ^71^ and HSA000203 ^72^ and 1 database (https://genomics.senescence.info/cells/)). We screen immune cells and compare samples from young and old people to obtain differential genes that are highly expressed in the elderly, remove duplicate genes, and screen genes that appear in at least two data sets. Based on the enrichment of relevant gene pathways and machines Learning method, 154 genes constituting the immunosenescence gene set were identified (Supplementary Table 4).

### GO enrichment analysis and Gene set enrichment analysis (GSEA)

Using the R package ClusterProfiler (version 4.8.3), gene function annotation and GO were performed on genes that were down-regulated and up-regulated in T cells and B cells in the MPR and noMPR groups, as well as differential genes between macrophages and monocytes and other cells. Analysis, a program that supports statistical analysis and visualization of functional profiles of genes and gene clusters.

GSEA was performed by the GSEA software (http://software.broadinstitute.org/gsea/index.jsp)^73^. Gene sets used in this article were c2.cp.kegg.v6.2.symbols.gmt downloaded from the Molecular Signatures Database (MSigDB, http://software.broadinstitute.org/gsea/msigdb/index.jsp).

### Gene Set Score Analysis

The scoregenes function in Scanpy is used to calculate the module score of a gene expression program in a single cell. First, all analyzed genes were binned according to their average expression, and control genes were randomly selected from each bin. Then, the average expression value of the gene set at the single-cell level minus the aggregate expression of the control gene set was calculated. The gene set was obtained from the immunosenescence gene set constructed by ourselves. The genes in each gene set are listed in Supplementary Table 4.

### Cell-cell interaction

We used the CellChat package (1.6.1), which allows analysis of scRNA-Seq data. We studied the interaction between immune cells (i.e. T cells and B cells) and myeloid cells (i.e. macrophages and monocytes), one of which contributes ligands or receptors during the interaction, using CellChat Signaling pathway networks are analyzed and visualized.

### Trajectory inference of immune cell subsets across tissues

The status of immune cell subpopulations in the periphery and in tumors is dynamic, and they may differentiate into different cell states and exert different biological functions in different patients. We use the pseudo-temporal inference algorithm Monocle 2 for trajectory analysis to reconstruct the cell differentiation trajectory of immune cells across tissues, reveal the progression of cells and reconstruct the trajectory of cells progressing through biological processes under study.

### Survival analysis

Survival analysis uses TCGA and HNSCC data to evaluate the prognostic performance of different immune cell populations. Gene expression data and clinical information of HNSCC patients were downloaded from cBioPortal (https://www.cbioportal.org/study/clinicalData). Use Cibersort and Xcell to extract T cells and B cells subtypes and group them based on the expression levels of characteristic genes. Specifically, HNSCC patients were divided into four groups based on the median proportion of immune cell subsets and the median mean expression of a given signature gene. In the analysis of myeloid cells, a subset of cells within a single cell is characterized using a set of signature genes, defined based on a combination of typical marker genes for major immune cell types and cluster-specific marker genes. Specifically, for different macrophage clusters, we used the typical IGF1^+^ Mφ marker genes “*CD276*”, “*APOE*”, “*TREM2*”, “*HTRA1*”, “*PTGR1*”, “*RBP1*”, “*IGF1*”, “*CDKN1A*”, “*CD81*” define its signature. HNSCC patients were divided into two groups based on the median of the mean expression of the given signature. Survival analysis was performed using the COX proportional hazards model implemented in the R package “survival”.

### Hierarchical Clustering

In the end, cell subpopulations and characteristic genes were used to construct a neoadjuvant treatment model for patients before surgery, and a hierarchical clustering method was used to create groups. Cophenetic distance was used to evaluate the objects in the group so that they were like each other and different from objects in other groups. Verify different cell subpopulations and gene signatures to evaluate the efficacy of neoadjuvant therapy in patients before surgery.

### TNBC single cell RNA sequencing data processing

TNBC single-cell RNA sequencing data comes from GSE169246. Patients who received combined neoadjuvant chemotherapy and immune therapy before surgery were screened, and the patients were divided into PR (response) and SD (non-response) groups based on the corresponding imaging conditions. The rest of the analysis process as before, similar cell subpopulations were evaluated after dimensionality reduction clustering, and senescent immune cells were evaluated on the cell subpopulations. Hierarchical clustering was used to verify the effect of different cell subpopulations and gene characteristics on patients receiving neoadjuvant therapy before surgery to evaluate.

### Animal experiments

Twelve-month-old female C57BL/6J mice (SPF grade), weighing 18-20 g, along with *Ercc1* knockout mice, were purchased from GemPharmatech Co., Ltd. The housing conditions were maintained at a temperature of 18-22°C and a humidity of 50-60%. Water was changed 2-3 times per week, and the mice were kept under a 12-hour light/dark cycle. The animal experiments in this study were conducted with the approval of the Animal Ethics Committee of Sun Yat-sen University and the Institutional Animal Care and Use Committee at the Shenzhen Institutes of Advanced Technology, Chinese Academy of Sciences (CAS). All experimental procedures adhered to the Regulations for the Administration of Affairs Concerning Experimental Animals issued by the People’s Republic of China. The animal experiments also complied with laboratory practice guidelines and standard operating procedures.

### 4-NQO-induced mouse HNSCC model

4NQO (Sigma, USA) was prepared at a concentration of 500 mg·L^−1^ in 1,2-propanediol and stored in a light-protected container at 4 °C. For administration, the 4NQO solution was diluted with distilled water to a final concentration of 50 mg·L^−1^ and provided in light-protected bottles as drinking water for female C57 mice (12 months old) for 16 weeks. After 16 weeks, the mice were switched to distilled water until the end of the experiment. Starting from the initiation of 4NQO administration, one mouse was randomly sacrificed each week to collect tongue tissue for paraffin embedding. The tissues were routinely embedded, and serial sections were made until no tissue remained. Sections with intact tissue samples were selected for further analysis. Hematoxylin-eosin (HE) staining was performed according to standard protocols. Pathological diagnosis was conducted following the WHO 2005 diagnostic criteria.

### Establishment of cell lines and tissue culture

Mice fed with 4NQO for 28-32 weeks (all exhibiting visible tumors) were euthanized by cervical dislocation under anesthesia. The mice were disinfected by soaking in 75% ethanol for 1 minute, followed by two rounds of povidone-iodine disinfection of the oral cavity. Tumor tissues were excised from the oral cavity using micro scissors and a scalpel. The tissues were then minced with ophthalmic scissors in a small volume of PBS and digested with enzymes (in 2.5% trypsin-DMEM) for 30 minutes in a shaking water bath at 37°C. After digestion, the cell suspension was filtered through a 70 µm strainer. The isolated tumor cells were plated at a density of 2 million cells per well in 6-well tissue culture plates and continuously cultured to select for cancer cells.

### SA-β-gal staining

Tumor tissues excised during surgery were washed three times with saline and then fixed in 4% PFA for 12 hours. The tissues were embedded in cryo-embedding medium (OCT) and rapidly frozen in liquid nitrogen. Serial sections of 5 μm thickness were cut using a cryostat. β-Galactosidase staining was performed according to the manufacturer’s protocol (C0602, Beyotime), followed by nuclear staining with eosin. After staining, images were captured under a 20x optical microscope.

### RNA isolation and qRT–PCR

Add 1 mL of TRIzol to a sterile culture tube. Place the frozen tissue into the tube and homogenize on ice using a homogenizer set to 25/30, grinding the tissue twice for 10 seconds each time. Total RNA was isolated using the TRIzol method according to the manufacturer’s instructions (Thermo Fisher). Subsequently, cDNA synthesis was performed using the SuperScript IV VILO Master Mix (Thermo Fisher) following the manufacturer’s protocol, with reactions done in duplicate for each sample. Data were analyzed using the ΔΔCt method, and expression levels were normalized to *Gapdh*.

### Quantitation of 8-OHdG DNA lesions

Tissues from mice were analysed for 8-OHdG levels using the ELISA kit (Abcam) according to manufacturer’s specifications.

### Immunoblotting

After rapidly freezing the spleen, liver, heart, and lung tissues from mice, homogenize the tissues using a tissue homogenizer. Lyse the cells with RIPA buffer (Thermo-Fisher) and collect the lysate into centrifuge tubes. Mix the lysate on a shaker for 4-15 minutes, then centrifuge at 14,000g for 15 minutes at 4°C. Discard the pellet and measure the protein concentration in the supernatant using the Bradford method. Load equal amounts of protein and molecular weight markers into the wells of an SDS-PAGE gel. Load 20-30 μg of total protein from the cell lysate or tissue homogenate per well. Transfer the proteins from the gel to a membrane and block the membrane with blocking buffer at room temperature for 1 hour. Incubate the membrane overnight at 4°C with anti-γH2AX (1:2000) and anti-GAPDH (1:5000) antibodies in blocking buffer. Wash the membrane three times with TBST, each wash lasting 5 minutes. Incubate the membrane with a secondary antibody conjugate in blocking buffer at room temperature for 1 hour, then wash and develop using ECL (Thermo-Fisher).

### TIL isolation and flow cytometry

After euthanizing the mice by cervical dislocation, tumor tissues were excised and placed on ice for 10 minutes. The tissues were washed twice with 1× PBS, minced, and digested with a solution containing collagenase II (1 mg/mL), collagenase IV (1 mg/mL), and DNase I (100 U/mL) for 30 minutes. The digested tumor tissues were filtered through a 70 µm strainer and washed once with 1× PBS to stop the digestion. The cell suspension was washed again with 1× PBS. A 10 µL aliquot of the cell suspension was diluted 10-fold with 1× PBS for cell counting. The single-cell suspension was adjusted to a cell density of 1×10^7 cells/mL, and 100 µL of the cell suspension was taken per tube for flow cytometry staining. For detection, the samples were divided into two groups. First, the cells were incubated with anti-mouse CD45 antibody at 4°C in the dark for 10 minutes. Group 1 was then stained with anti-mouse CD3, CD4, CD8, B220 antibodies, and corresponding isotype control antibodies. Group 2 was stained with anti-mouse CD45, CD4, CD8, CD44, CD62L antibodies, and corresponding isotype control antibodies. Intracellular staining for CCR7 and LCK was performed using the eBioscience transcription factor buffer set from Invitrogen. After incubating at 4°C in the dark for 30 minutes, the cells were washed once with 1× PBS and analyzed by flow cytometry. The immunophenotyping of the cells was defined as follows: CD4+ T cells (CD45+ CD3+ CD4+), CD8+ T cells (CD45+ CD3+ CD8+), B cells (CD45+ B220+), and CD4+ Naive T cells (CD4+ CD62L+ CD44-). Flow cytometry data were analyzed using FlowJo-V10 software. The number of infiltrating cells per unit volume of the tumor was calculated as the number of each cell subset divided by the tumor volume.

### Immunofluorescence and image quantification

The method for multiplex immunofluorescence staining of paraffin sections was referenced from our previously published literature^74^. Briefly, tissues were harvested, fixed, and paraffin embedded. Slides were stained for P16 (Abcam, ab185620, 1:500), P21 (Abcam, ab109520, 1:1000), CD4 (Abcam, ab133616, 1:1000), CD8a (Cell signaling Technology, 70306S, 1:500), CCR7 (Abcam, ab253187, 1:1000), PAN-CK (Cell signaling Technology, 4545S, 1:2000), LCK (Cell signaling Technology, 2984S), CD62L (Invitrogen, PA5-95721, 1:500) and S100A11 (Abcam, ab180593, 1:1000) antibodies. Quantification of immune infiltration was done using QuPath, an open-source software for digital pathology image analysis. For the quantification, at least three regions of interest (ROI) were selected for each condition and the percentage of positive cells was calculated.

## Data availability

All data associated with this study are present in the paper or the Supplementary Materials. The raw sequence data including single-cell sequencing, TCR/BCR sequencing of human, as well as RNA and ATAC sequencing of CD4 naïve T cells datasets generated and analyzed during the current study have been deposited in Genome Sequence Archive (Genomics, Proteomics & Bioinformatics 2021) with accession code **PRJCA027702 and PRJCA028161**.

## Code availability

No unique code was used in the study. All relevant references are included in Methods.

