## Supplementary Information for "Reversing Immunosenescence with Senolytics to Enhance Tumor Immunotherapy"

#### Supplementary Information Guide

| Supplementary Information | Title |
|---|---|
| Extended Data Figure 1 | **The trial design and outcomes of OOC-001.** |
| Extended Data Figure 2 | **Dynamic characterization of TCR landscape.** |
| Extended Data Figure 3 | **Development and validation of IAGs.** |
| Extended Data Figure 4 | **Anti-tumor reactivity and immunosenescence features of T cell subtypes.** |
| Extended Data Figure 5 | **CD4 T cell differentiation pathway.** |
| Extended Data Figure 6 | **Immunosenescence features of B cells.** |
| Extended Data Figure 7 | **Dynamics of myeloid cell subtypes after neoadjuvant chemoimmunotherapy.** |
| Extended Data Figure 8 | **The effect of senolytics combined with αPD-1 on TIME of 4-NQO mice.** |
| Extended Data Figure 9 | **αPD-1+DQ enhanced the anti-tumor effect in breast and bladder cancer xenograft models.** |
| Extended Data Figure 10 | **Transcriptomic and epigenetic dynamics of CD4 naive T cells following the αPD-1+DQ therapy.** |
| Supplementary Figure 1 | **Single-cell profiling of all cells in HNSCC.** |
| Supplementary Figure 2 | **Differential gene expression analysis for all immune cells.** |
| Supplementary Figure 3 | **Immune cells proportion between MPR and noMPR patients.** |
| Supplementary Figure 4 | **Survival curve between the expression levels of senescent genes and multiple T and B cells in TCGA samples.** |
| Supplementary Figure 5 | **Interaction between myeloid cells and T/B cells.** |

| Supplementary Information | Title |
| --- | --- |
| Supplementary Figure 6 | **Senolytics combined with αPD-1 reduced the expression of the aging marker P16 in multiple organs.** |
| Supplementary Protocol | **The protocol of OOC-001** |
| Supplementary Protocol | **The protocol of COIS-01** |
| Supplementary Table 1-1 | **Demographic and clinical characteristics of OOC-001, related to Figure 1.** |
| Supplementary Table 1-2 | **Demographic and clinical characteristics of 7 HNSCC patients, related to Figure 1.** |
| Supplementary Table 2 | **Treatment-related adverse events of OOC-001** |
| Supplementary Table 3-1 | **DE genes of $CXCL13^{+}$CD4 T helper in tumors between MPR and noMPR.** |
| Supplementary Table 3-2 | **DE genes of $CCR7^{+}$CD4 naïve T cells in tumors between MPR and noMPR.** |
| Supplementary Table 3-3 | **DE genes of $CD27^{+}$Memory B cells in tumors between MPR and noMPR.** |
| Supplementary Table 4 | **Immunosenescence-related genes (IAGs).** |
| Supplementary Table 5 | **Luminex assay.** |
| Supplementary Table 6-1 | **DE genes of CD4 naïve T cells between anti-PD-1+DQ and Control groups, related to Figure 5.** |
| Supplementary Table 6-2 | **DE genes of CD4 naïve T cells between anti-PD-1 and Control groups.** |
| Supplementary Table 6-3 | **DE genes of CD4 naïve T cells between anti-PD-1 and anti-PD-1+DQ groups.** |
| Supplementary Table 7-1 | **Signature peaks and chromatin dynamics of CD4 naïve T cells, related to Figure 5.** |
| Supplementary Table 7-2 | **Signature peaks and chromatin dynamics of T cell clusters, related to Figure 5.** |
| Supplementary Table 8 | **Demographic and clinical characteristics of COIS-01** |
| Supplementary Table 9 | **Treatment-related adverse events of COIS-01** |

### Extended Data Figures 1-10:

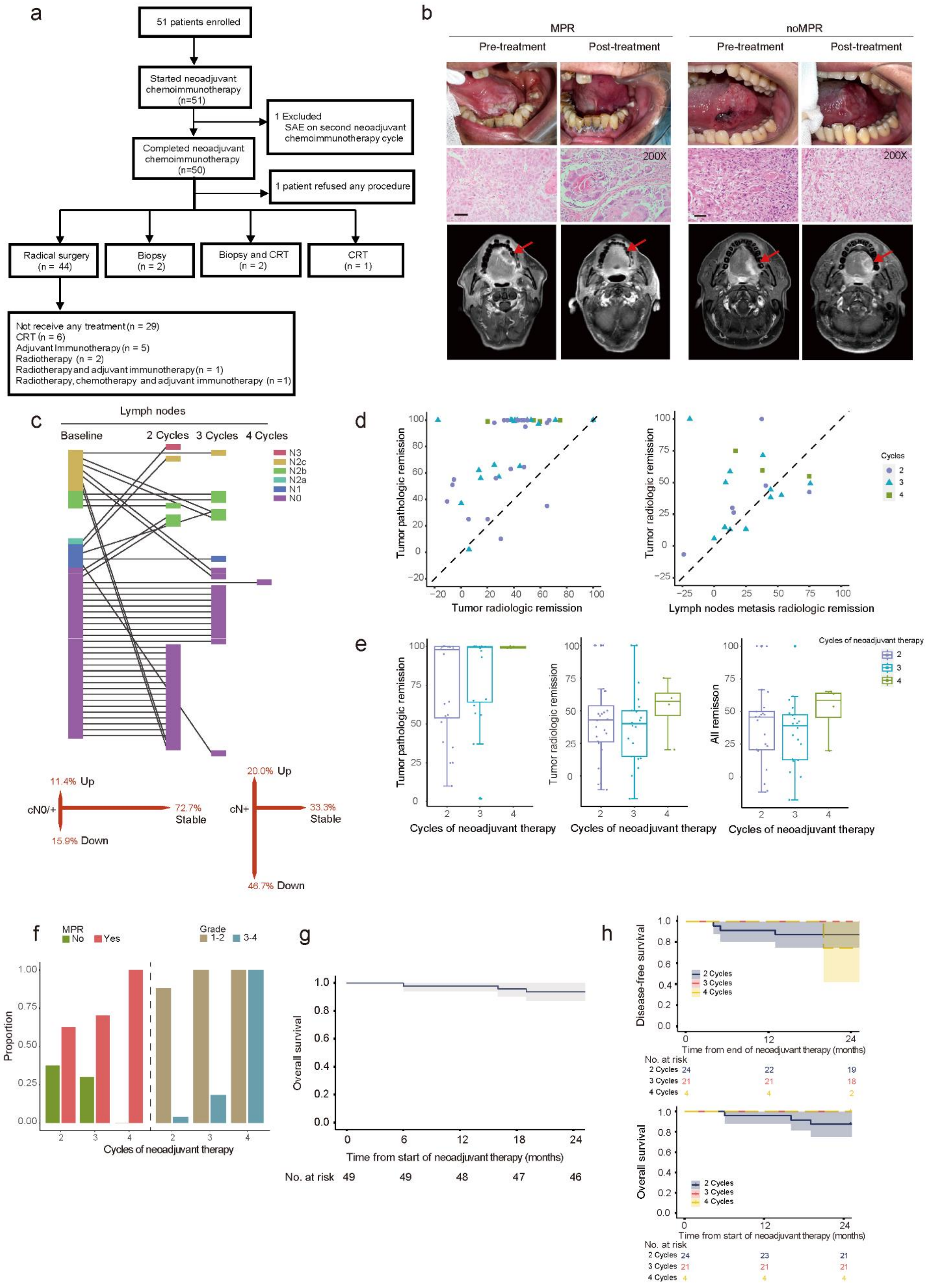

**Extended Data Fig. 1 The trial design and outcomes of OOC-001.**

**a**, Trial flow diagram. **b**, Representative pre- and post-treatment intraoral photographs, H&E staining, and MRI images for MPR and noMPR patients. Scale bar = 50 μm. **c**, The changes and proportional statistics of TNM staging after treatment for lymph nodes in different patients. **d**, Scatter plot depicts the pathological response and radiographic response of the primary tumor after neoadjuvant treatment in the same patient (left), and the radiographic response of the primary tumor and lymph node metastasis in the same patient after neoadjuvant treatment (right). **e**, Pathological response of the primary tumor in patients treated with different cycles during neoadjuvant therapy (left), radiographic response of the primary tumor in patients treated with different cycles during neoadjuvant therapy (middle), and all remission include primary tumor and lymph node metastasis in patients treated with different cycles during neoadjuvant therapy (right). **f**, The rates of MPR for patients after 2 cycles, 3 cycles, and 4 cycles of treatment, as well as the rates of Grade 1-2 adverse events and Grade 3-4 adverse events. **g**, Survival curves for patients in this clinical trial at the 24-month follow-up. **h**, Disease-free survival (DFS) and OS curves for patients in this clinical trial at the 24-month follow-up.

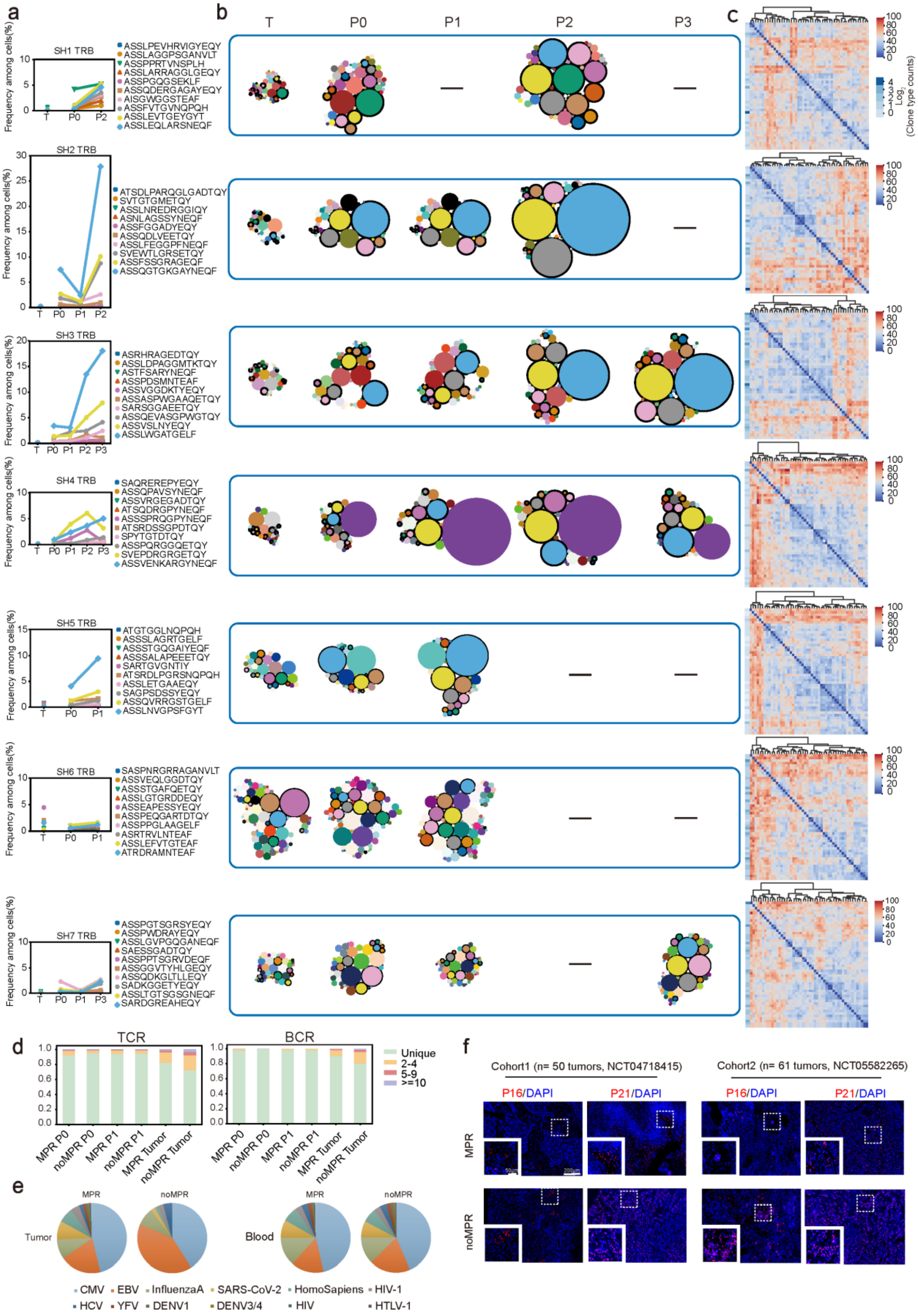
a
b
c
T
P0
P1
P2
P3
SH1 TRB
ASSLPEVHRVIGYEQY
ASSLAGGPSGANVLT
ASSPPRTVNSPLH
ASSLARRAGGLGEQY
ASSPGQGSEKLF
ASSQDERGAGAYEQY
AISGWGGSTEAF
ASSFVTGVNQPQH
ASSLEVTGEYGYT
ASSLEQLARSNEQF
SH2 TRB
ATSDLPARQGLGADTQY
SVTGTGMETQY
ASSLNREDRGGIQY
ASNLAGSSYNEQF
ASSFGGADYEQY
ASSQDLVEETQY
ASSLFEGGPFNEQF
SVEWTLGRSETQY
ASSFSSGRAGEQF
ASSQGTGKGAYNEQF
SH3 TRB
ASRHRAGEDTQY
ASSLDPAGGMTKTQY
ASTFSARYNEQF
ASSPDSMNTEAF
ASSVGGDKTYEQY
ASSASPWGAAQETQY
SARSGGAEETQY
ASSQEVASGPWGTQY
ASSVSLNYEQY
ASSLWGATGELF
SH4 TRB
SAQREREPYEQY
ASSQPAVSYNEQF
ASSVRGEGADTQY
ATSQDRGPYNEQF
ASSSPRQGPYNEQF
ATSRDSSGPDTQY
SPYTGTDTQY
ASSPQRGGQETQY
SVEPDRGRGETQY
ASSVENKARGYNEQF
SH5 TRB
ATGTGGLNQPQH
ASSSLAGRTGELF
ASSSTGQGAIYEQF
ASSSALAPEEETQY
SARTGVGNTIY
ATSRDLPGRSNQPQH
ASSLETGAAEQY
SAGPSDSSYEQY
ASSQVRRGSTGELF
ASSLNVGPSFGYT
SH6 TRB
SASPNRGRRAGANVLT
ASSVEQLGGDTQY
ASSSTGAFQETQY
ASSLGTGRDDEQY
ASSEAPESSYEQY
ASSPEQGARTDTQY
ASSPPGLAAGELF
ASRTRVLNTEAF
ASSLEFVTGTEAF
ATRDRAMNTEAF
SH7 TRB
ASSPGTSGRSYEQY
ASSPWDRAYEQY
ASSLGVPGQGANEQF
SAESSGADTQY
ASSPPTSGRVDEQF
ASSGGVTYHLGEQY
ASSQDKGLTLLEQY
SADKGGETYEQY
ASSLTGTSGSGNEQF
SARDGREAHEQY
Frequency among cells(%)
Log2 (Clone type counts)
d
TCR
BCR
Unique
2-4
5-9
>=10
MPR P0
noMPR P0
MPR P1
noMPR P1
MPR Tumor
noMPR Tumor
e
MPR
noMPR
Tumor
Blood
CMV
EBV
InfluenzaA
SARS-CoV-2
HomoSapiens
HIV-1
HCV
YFV
DENV1
DENV3/4
HIV
HTLV-1
f
Cohort1 (n= 50 tumors, NCT04718415)
Cohort2 (n= 61 tumors, NCT05582265)
P16/DAPI
P21/DAPI
MPR
noMPR

**Extended Data Fig. 2 Dynamic characterization of TCR landscape.**

**a**, TCR-seq was performed on T cells from tumors (pre-treatment biopsy tumors), P0 (pre-treatment blood), and P1 (post-treatment blood) obtained from patients (from top to bottom, patients SH1, SH2, SH3, SH4, SH5, SH6, SH7). The top 10 clonotype frequencies detected in T cells that differed most in peripheral blood before and after treatment are shown in serial peripheral blood (left) and tumor tissue (right). **b**, From left to right represents changes in high abundance clonotypes in tumors and blood before and after patient treatment, and patients from top to bottom are SH1, SH2, SH3, SH4, SH5, SH6, SH7. Each circle represents a clonotype. The diameter of the circle indicates the concentration of the clonotype. Persistent clonotypes are marked as bright circles, with a single color representing one clonotype. For patients with MPR, the predominant high-abundance clonotype present in tumors and blood before treatment remained the predominant high-abundance clonotype after treatment. **c**, Pairwise similarity (Grantham distance) between the top 50 most common intratumoral TCR CDR3b clones. Evaluate the similarity between the most common lengths of CDR3b amino acid sequences in each patient (from top to bottom, patients SH1, SH2, SH3, SH4, SH5, SH6, SH7). Red indicates the most expanded clone. **d**, Bar plot of TCR and BCR clonotypes across all T cells and B cells in MPR or noMPR groups from P0, P1 and tumors. One clonotype consists strictly of one paired α-/β-chain V(D)J TCR. **e**, Pie chart of the antigen specificity of detected TCRs when considering α-, β-, and paired strands. CMV, cytomegalovirus; EBV, Epstein-Barr virus; InfluenzaA, influenza A virus; SARS-CoV-2, severe acute respiratory syndrome coronavirus 2; HS, Homo sapiens antigen; HIV-1, human immunodeficiency virus 1; HCV, hepatitis C virus; YFV, yellow fever virus; DEV, dengue virus. **f**, Representative immunofluorescence images of senescent markers P16 and P21 on paraffin sections from MPR and noMPR patients in two patient cohorts (Cohort1 n=50, Cohort2 n=61).

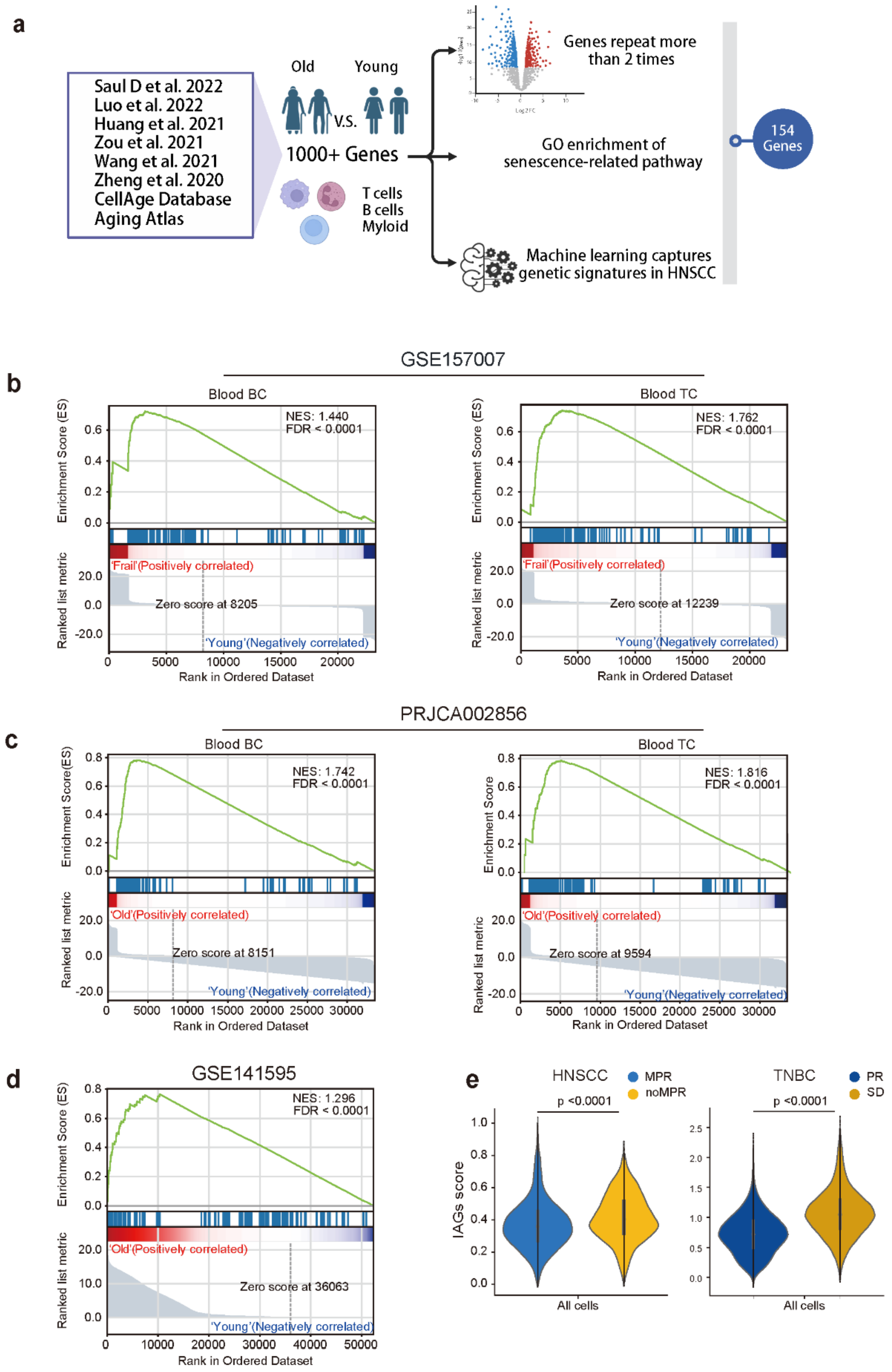
a
Saul D et al. 2022
Luo et al. 2022
Huang et al. 2021
Zou et al. 2021
Wang et al. 2021
Zheng et al. 2020
CellAge Database
Aging Atlas
Old
Young
V.S.
1000+ Genes
T cells
B cells
Myloid
Genes repeat more than 2 times
GO enrichment of senescence-related pathway
Machine learning captures genetic signatures in HNSCC
154 Genes
b
GSE157007
Blood BC
NES: 1.440
FDR < 0.0001
'Frail'(Positively correlated)
Zero score at 8205
'Young'(Negatively correlated)
Blood TC
NES: 1.762
FDR < 0.0001
'Frail'(Positively correlated)
Zero score at 12239
'Young'(Negatively correlated)
Enrichment Score (ES)
Ranked list metric
Rank in Ordered Dataset
c
PRJCA002856
Blood BC
NES: 1.742
FDR < 0.0001
'Old'(Positively correlated)
Zero score at 8151
'Young'(Negatively correlated)
Blood TC
NES: 1.816
FDR < 0.0001
'Old'(Positively correlated)
Zero score at 9594
'Young'(Negatively correlated)
Enrichment Score(ES)
Enrichment Score
Ranked list metric
Rank in Ordered Dataset
d
GSE141595
NES: 1.296
FDR < 0.0001
'Old'(Positively correlated)
Zero score at 36063
'Young'(Negatively correlated)
Enrichment Score (ES)
Ranked list metric
Rank in Ordered Dataset
e
HNSCC
MPR
noMPR
p <0.0001
IAGs score
All cells
TNBC
PR
SD
p <0.0001
All cells

**Extended Data Fig. 3 Development and validation of IAGs.**

**a,** Human immune cell samples from multiple data sets were used for single-cell RNA sequencing analysis to construct the IAGs. **b,** The IAGs significantly enriched in B cells (middle) and T cells (right) in peripheral blood of frail elderly people (GSE157007). **c,** The IAGs is significantly enriched in B cells (middle) and T cells (right) in the peripheral blood of elderly women (PRJCA002856). **d,** The IAGs is significantly enriched in all immune cells in bone marrow of older women (GSE141595). **e,** Violin plot of expression of the different all cells IAGs score in HNSCC tumors between MPR and noMPR groups (left), and in TNBC tumors between PR and SD groups (right). Black lines with different lengths indicate which two groups were compared. Wilcoxon signed-rank test.

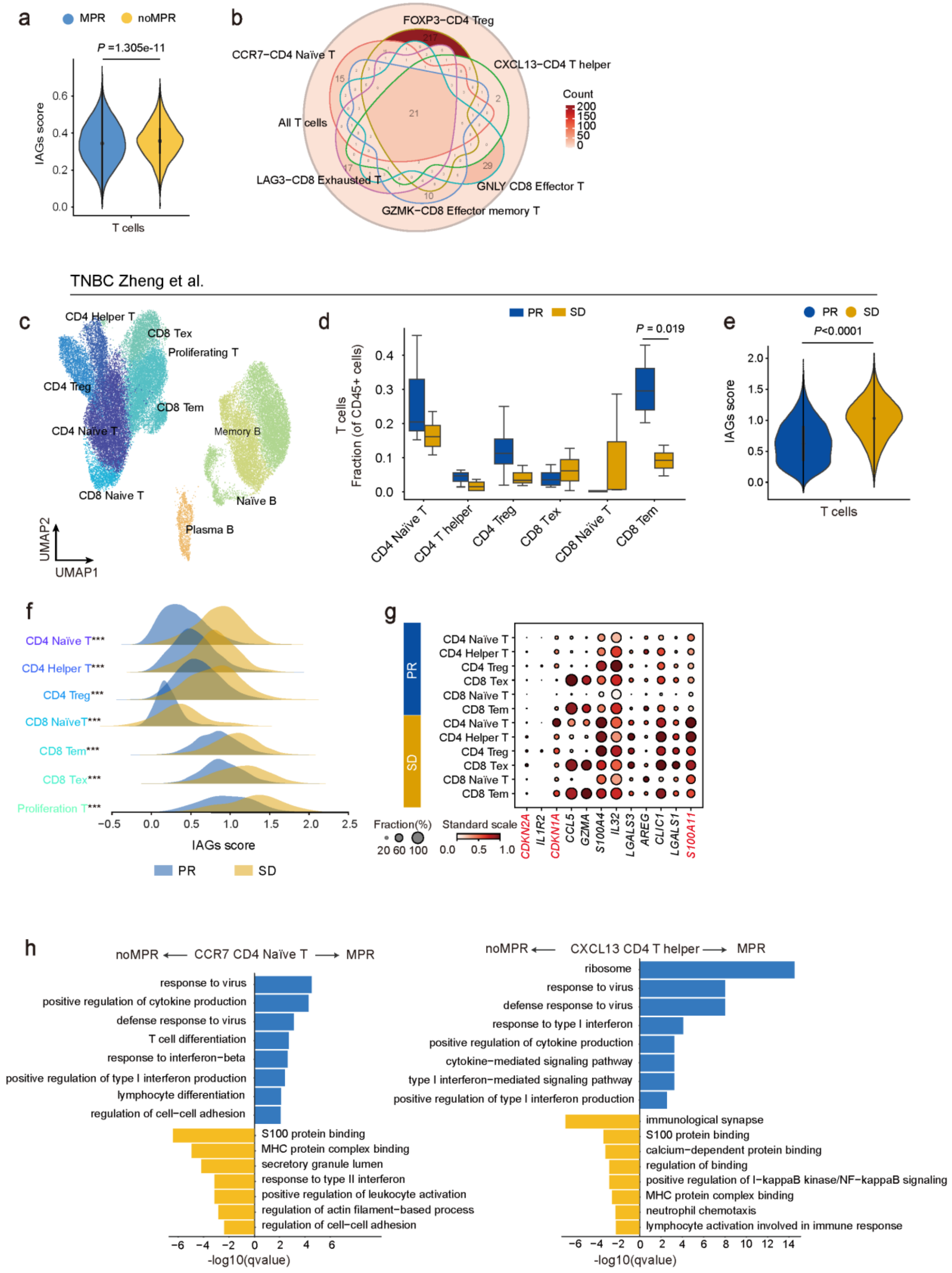

a
MPR
noMPR
P =1.305e-11
IAGs score
T cells
b
FOXP3−CD4 Treg
CCR7−CD4 Naïve T
CXCL13−CD4 T helper
All T cells
LAG3−CD8 Exhausted T
GNLY CD8 Effector T
GZMK−CD8 Effector memory T
Count
TNBC Zheng et al.
c
CD4 Helper T
CD8 Tex
Proliferating T
CD4 Treg
CD8 Tem
CD4 Naïve T
Memory B
CD8 Naïve T
Naïve B
Plasma B
UMAP2
UMAP1
d
PR
SD
P = 0.019
T cells
Fraction (of CD45+ cells)
CD4 Naïve T
CD4 T helper
CD4 Treg
CD8 Tex
CD8 Naïve T
CD8 Tem
e
PR
SD
P<0.0001
IAGs score
T cells
f
CD4 Naïve T***
CD4 Helper T***
CD4 Treg***
CD8 NaïveT***
CD8 Tem***
CD8 Tex***
Proliferation T***
IAGs score
PR
SD
g
PR
SD
CD4 Naïve T
CD4 Helper T
CD4 Treg
CD8 Tex
CD8 Naïve T
CD8 Tem
Fraction(%)
Standard scale
CDKN2A
IL1R2
CDKN1A
CCL5
GZMA
S100A4
IL32
LGALS3
AREG
CLIC1
LGALS1
S100A11
h
noMPR ⟵ CCR7 CD4 Naïve T ⟶ MPR
response to virus
positive regulation of cytokine production
defense response to virus
T cell differentiation
response to interferon−beta
positive regulation of type I interferon production
lymphocyte differentiation
regulation of cell−cell adhesion
S100 protein binding
MHC protein complex binding
secretory granule lumen
response to type II interferon
positive regulation of leukocyte activation
regulation of actin filament−based process
regulation of cell−cell adhesion
-log10(qvalue)
noMPR ⟵ CXCL13 CD4 T helper ⟶ MPR
ribosome
response to virus
defense response to virus
response to type I interferon
positive regulation of cytokine production
cytokine−mediated signaling pathway
type I interferon−mediated signaling pathway
positive regulation of type I interferon production
immunological synapse
S100 protein binding
calcium−dependent protein binding
regulation of binding
positive regulation of I−kappaB kinase/NF−kappaB signaling
MHC protein complex binding
neutrophil chemotaxis
lymphocyte activation involved in immune response
-log10(qvalue)

**Extended Data Fig. 4 Anti-tumor reactivity and immunosenescence features of T cell subtypes.**

**a**, Violin plot of expression of all T cells IAGs score in HNSCC tumors between MPR and noMPR groups. Black lines with different lengths indicate which two groups were compared. Wilcoxon signed-rank test. **b**, Gene Venn diagram of differential genes of T cell subtypes in tumors between MPR and noMPR groups. **c**, UMAP shows the distribution of T cell and B cell subtypes in TNBC. **d**, Box plot of the percentage of the T cell clusters in TNBC tumors between PR and SD groups. The boxed plot on the left of each T cell cluster is the PR group, and on the right is SD group. Box middle lines, median; box limits, upper and lower quartiles. Black lines with different lengths indicate which two groups were compared. Wilcoxon signed-rank test. **e**, Violin plot of expression of all T cells IAGs score in TNBC tumors between PR and SD groups. Black lines with different lengths indicate which two groups were compared. Wilcoxon signed-rank test. **f**, The Ridge plot of IAGs score analysis for T cells clusters in TNBC. **g**, Dot plot of intersection genes of TNBC T cell DEGs and IAGs between PR and SD groups. **h**, DEGs (Log2Foldchages > 0.5) of CCR7$^{+}$ CD4 naïve T cells and CXCL13 CD4 T helper cells between MPR and noMPR enriched GO terms in tumors. Log-rank test (two-sided). *P < 0.05, **P < 0.01, ***P < 0.001.

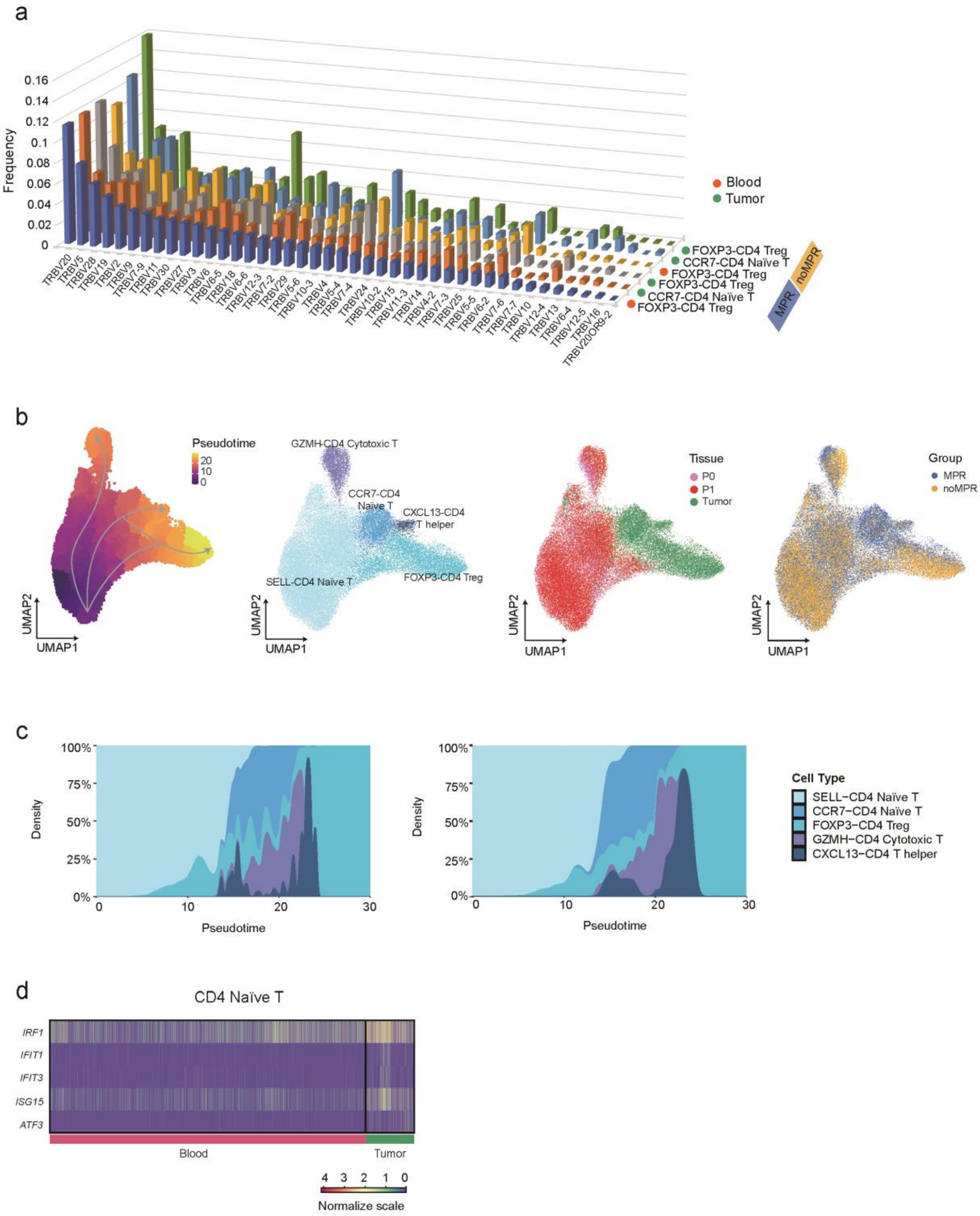

a
Frequency
0.16
0.14
0.12
0.1
0.08
0.06
0.04
0.02
0
TRBV20
TRBV5
TRBV28
TRBV19
TRBV2
TRBV9
TRBV7-9
TRBV11
TRBV30
TRBV27
TRBV3
TRBV6
TRBV6-5
TRBV18
TRBV6-6
TRBV12-3
TRBV7-2
TRBV29
TRBV5-6
TRBV10-3
TRBV4
TRBV5-4
TRBV7-4
TRBV24
TRBV10-2
TRBV15
TRBV11-3
TRBV14
TRBV4-2
TRBV7-3
TRBV25
TRBV5-5
TRBV6-2
TRBV7-6
TRBV7-7
TRBV10
TRBV12-4
TRBV13
TRBV6-4
TRBV12-5
TRBV16
TRBV20OR9-2
Blood
Tumor
FOXP3-CD4 Treg
CCR7-CD4 Naïve T
FOXP3-CD4 Treg
FOXP3-CD4 Treg
CCR7-CD4 Naïve T
FOXP3-CD4 Treg
noMPR
MPR
b
Pseudotime
20
10
0
UMAP2
UMAP1
GZMH-CD4 Cytotoxic T
CCR7-CD4 Naïve T
CXCL13-CD4 T helper
SELL-CD4 Naïve T
FOXP3-CD4 Treg
Tissue
P0
P1
Tumor
Group
MPR
noMPR
c
100%
75%
50%
25%
0%
Density
0
10
20
30
Pseudotime
Cell Type
SELL−CD4 Naïve T
CCR7−CD4 Naïve T
FOXP3−CD4 Treg
GZMH−CD4 Cytotoxic T
CXCL13−CD4 T helper
d
CD4 Naïve T
IRF1
IFIT1
IFIT3
ISG15
ATF3
Blood
Tumor
4 3 2 1 0
Normalize scale

**Extended Data Fig. 5 CD4 T cell differentiation pathway.**

**a**, Frequency of Vα/β gene usage among $CCR7^+$ CD4 T cells and $FOXP3^+$ CD4 Treg cells in tumors and $CCR7^+$ CD4 T cells in blood (V genes ordered by decreasing frequency in $CCR7^+$ CD4 T cells in blood). **b**, UMAP embedded CD4 T cell trajectory with mapping of pseudotime (top left), cell subtype distribution (top right), single-cell transcriptional profiles of each tissue (bottom left), and different groups (bottom right) sampled in the study. **c**, Pseudotime distribution diagram of the proportion of CD4 T cells in tumors, the MPR group on the left, the noMPR group on the right. **d**, Differential genes of CD4 Naïve T cells in tumors and peripheral blood.

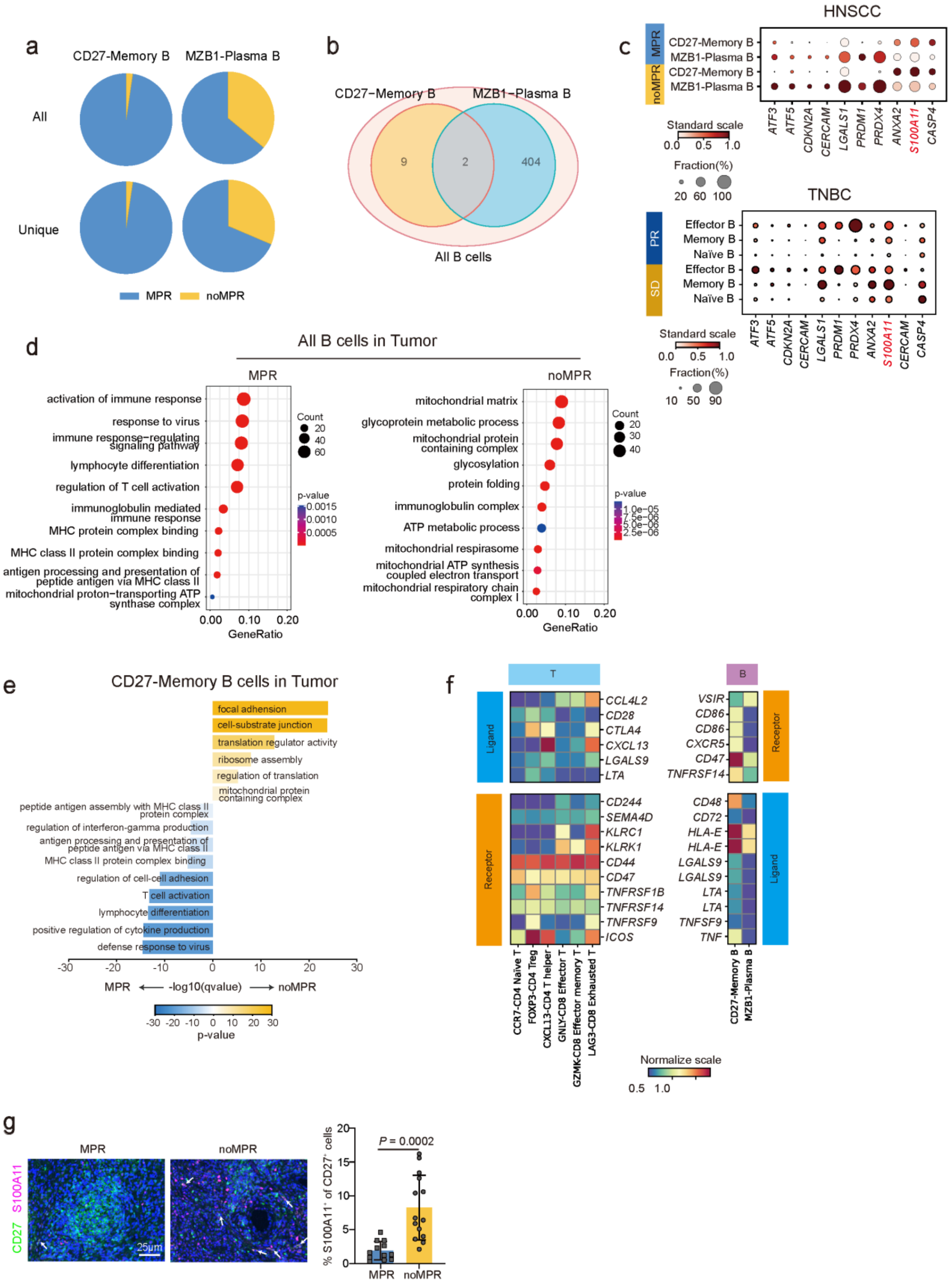

a
CD27-Memory B
MZB1-Plasma B
All
Unique
MPR
noMPR
b
CD27−Memory B
MZB1−Plasma B
9
2
404
All B cells
c
HNSCC
MPR
noMPR
CD27-Memory B
MZB1-Plasma B
CD27-Memory B
MZB1-Plasma B
ATF3
ATF5
CDKN2A
CERCAM
LGALS1
PRDM1
PRDX4
ANXA2
S100A11
CASP4
Standard scale
0.0 0.5 1.0
Fraction(%)
20 60 100
TNBC
PR
SD
Effector B
Memory B
Naïve B
Effector B
Memory B
Naïve B
ATF3
ATF5
CDKN2A
CERCAM
LGALS1
PRDM1
PRDX4
ANXA2
S100A11
CERCAM
CASP4
Standard scale
0.0 0.5 1.0
Fraction(%)
10 50 90
d
All B cells in Tumor
MPR
activation of immune response
response to virus
immune response−regulating signaling pathway
lymphocyte differentiation
regulation of T cell activation
immunoglobulin mediated immune response
MHC protein complex binding
MHC class II protein complex binding
antigen processing and presentation of peptide antigen via MHC class II
mitochondrial proton−transporting ATP synthase complex
Count
20
40
60
p-value
0.0015
0.0010
0.0005
0.00 0.10 0.20
GeneRatio
noMPR
mitochondrial matrix
glycoprotein metabolic process
mitochondrial protein containing complex
glycosylation
protein folding
immunoglobulin complex
ATP metabolic process
mitochondrial respirasome
mitochondrial ATP synthesis coupled electron transport
mitochondrial respiratory chain complex I
Count
20
30
40
p-value
1.0e−05
7.5e−06
5.0e−06
2.5e−06
0.00 0.10 0.20
GeneRatio
e
CD27-Memory B cells in Tumor
focal adhesion
cell-substrate junction
translation regulator activity
ribosome assembly
regulation of translation
mitochondrial protein containing complex
peptide antigen assembly with MHC class II protein complex
regulation of interferon-gamma production
antigen processing and presentation of peptide antigen via MHC class II
MHC class II protein complex binding
regulation of cell-cell adhesion
T cell activation
lymphocyte differentiation
positive regulation of cytokine production
defense response to virus
-30 -20 -10 0 10 20 30
MPR ⟵ -log10(qvalue) ⟶ noMPR
-30 -20 -10 0 10 20 30
p-value
f
T
B
Ligand
Receptor
CCL4L2
CD28
CTLA4
CXCL13
LGALS9
LTA
CD244
SEMA4D
KLRC1
KLRK1
CD44
CD47
TNFRSF1B
TNFRSF14
TNFRSF9
ICOS
VSIR
CD86
CD86
CXCR5
CD47
TNFRSF14
CD48
CD72
HLA-E
HLA-E
LGALS9
LGALS9
LTA
LTA
TNFSF9
TNF
CCR7-CD4 Naïve T
FOXP3-CD4 Treg
CXCL13-CD4 T helper
GNLY-CD8 Effector T
GZMK-CD8 Effector memory T
LAG3-CD8 Exhausted T
CD27-Memory B
MZB1-Plasma B
Normalize scale
0.5 1.0
g
MPR
noMPR
CD27 S100A11
25μm
P = 0.0002
% S100A11⁺ of CD27⁺ cells
0
5
10
15
20
MPR
noMPR

**Extended Data Fig. 6 Immunosenescence features of B cells.**

**a**, Paired BCR results Shows the overall number of BCR clones (upper row), and the status of single clones after normalization (lower row). **b**, Gene Venn diagram of differential genes of B cell subtypes in tumors between MPR and noMPR groups. **c**, Dot plot of intersection genes of HNSCC B cell DEGs and IAGs between MPR and noMPR groups (upper), and between PR and SD groups in TNBC. **d**, The dot plot showing DEGs (Log2Foldchages > 0.5) of B cells between MPR and noMPR groups enriched GO terms in tumors. **e**, GO terms enriched in $CD27^{-}$Memory B cells in tumors between MPR and noMPR groups. adjusted p value < 0.05. **f**, Heatmap showing the expression of ligand-receptor pairs highly expressed in T cell subtypes and B cell subtypes in tumor. **g**, Immunohistological staining quantifications of CD27 and S100A11 in HNSCC patients of two cohorts (MPR n=12, noMPR n=14).

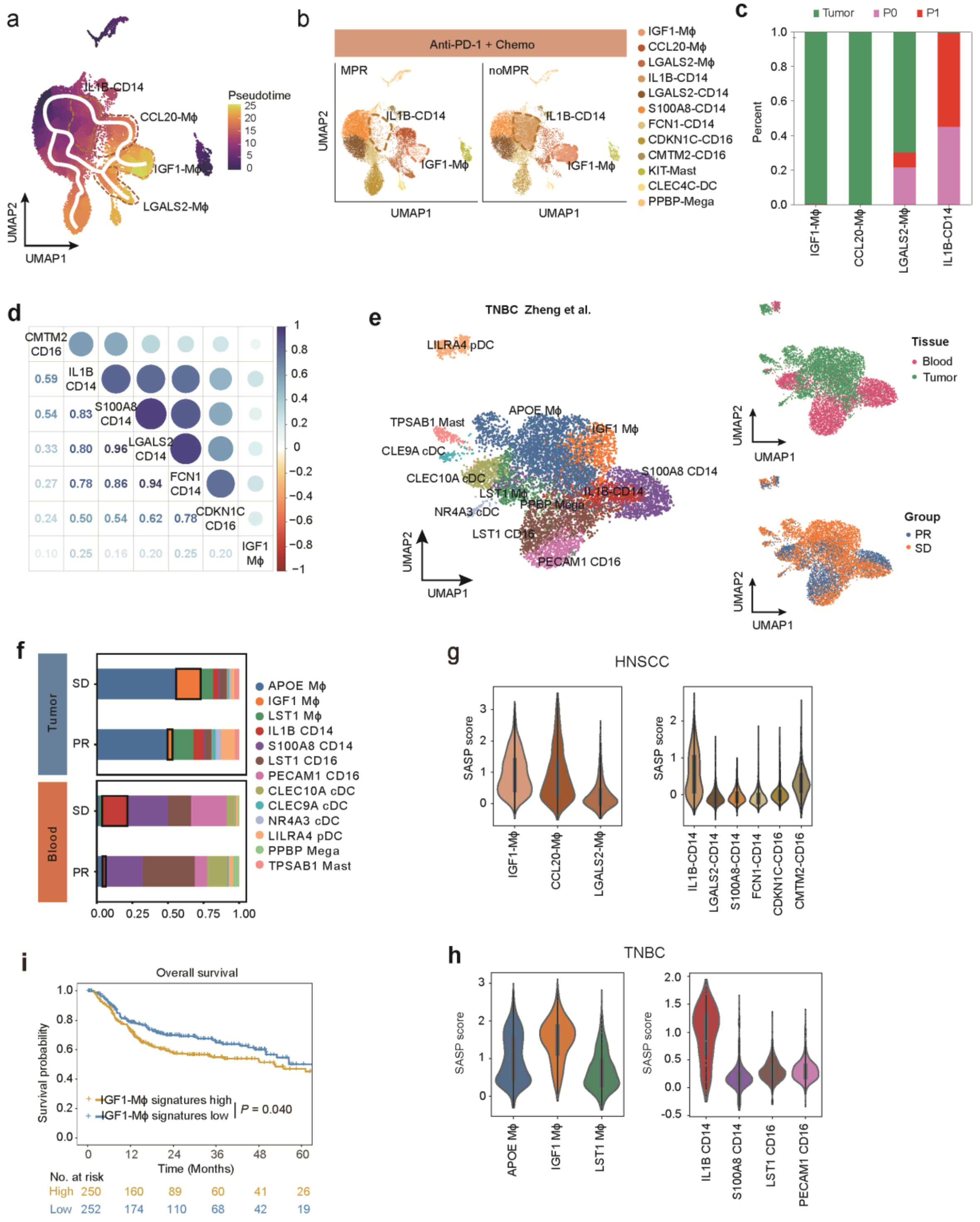

a
Pseudotime
IL1B-CD14
CCL20-Mϕ
IGF1-Mϕ
LGALS2-Mϕ
UMAP1
UMAP2
b
Anti-PD-1 + Chemo
MPR
noMPR
IGF1-Mϕ
CCL20-Mϕ
LGALS2-Mϕ
IL1B-CD14
LGALS2-CD14
S100A8-CD14
FCN1-CD14
CDKN1C-CD16
CMTM2-CD16
KIT-Mast
CLEC4C-DC
PPBP-Mega
c
Tumor
P0
P1
Percent
d
e
TNBC Zheng et al.
LILRA4 pDC
TPSAB1 Mast
APOE Mϕ
IGF1 Mϕ
CLE9A cDC
CLEC10A cDC
S100A8 CD14
LST1 Mϕ
PPBP Mega
NR4A3 cDC
LST1 CD16
PECAM1 CD16
Tissue
Blood
Tumor
Group
PR
SD
f
Tumor
Blood
LST1 Mϕ
IL1B CD14
CLEC9A cDC
LILRA4 pDC
TPSAB1 Mast
g
HNSCC
SASP score
h
TNBC
i
Overall survival
Survival probability
IGF1-Mϕ signatures high
IGF1-Mϕ signatures low
P = 0.040
Time (Months)
No. at risk
High 250 160 89 60 41 26
Low 252 174 110 68 42 19

**Extended Data Fig. 7 Dynamics of myeloid cell subtypes after neoadjuvant chemoimmunotherapy.**

**a**, UMAP embedding myeloid cell trajectory with mapping of pseudotime. **b**, UMAP plots showing the distributions myeloid cells between MPR and noMPR groups. **c**, The proportion of myeloid cells in different tissues. **d**, Gene similarity between IGF1$^{+}$ Mφ and myeloid cell clusters from peripheral blood. **e**, UMAP showing the distribution of myeloid cell clusters (left), embedding the single-cell transcriptional profiles of myeloid cell from each tissue sampled (top right), and from different groups (bottom right) in TNBC. **f**, The cellular compositions of myeloid cell clusters in tumors and peripheral blood between PR and SD groups in TNBC. **g**, Violin plot of six key SASP expression of the IAGs score in myeloid cell clusters in tumors (left) and peripheral blood (right). **h**, Violin plot of six key SASP expression of the IAGs score in myeloid cell clusters in tumors (left) and peripheral blood (right) from TNBC. **i**, Low expressions of IGF1$^{+}$ Mφ signature gene were significant associated with better OS and DFS. Log-rank test (two-sided).

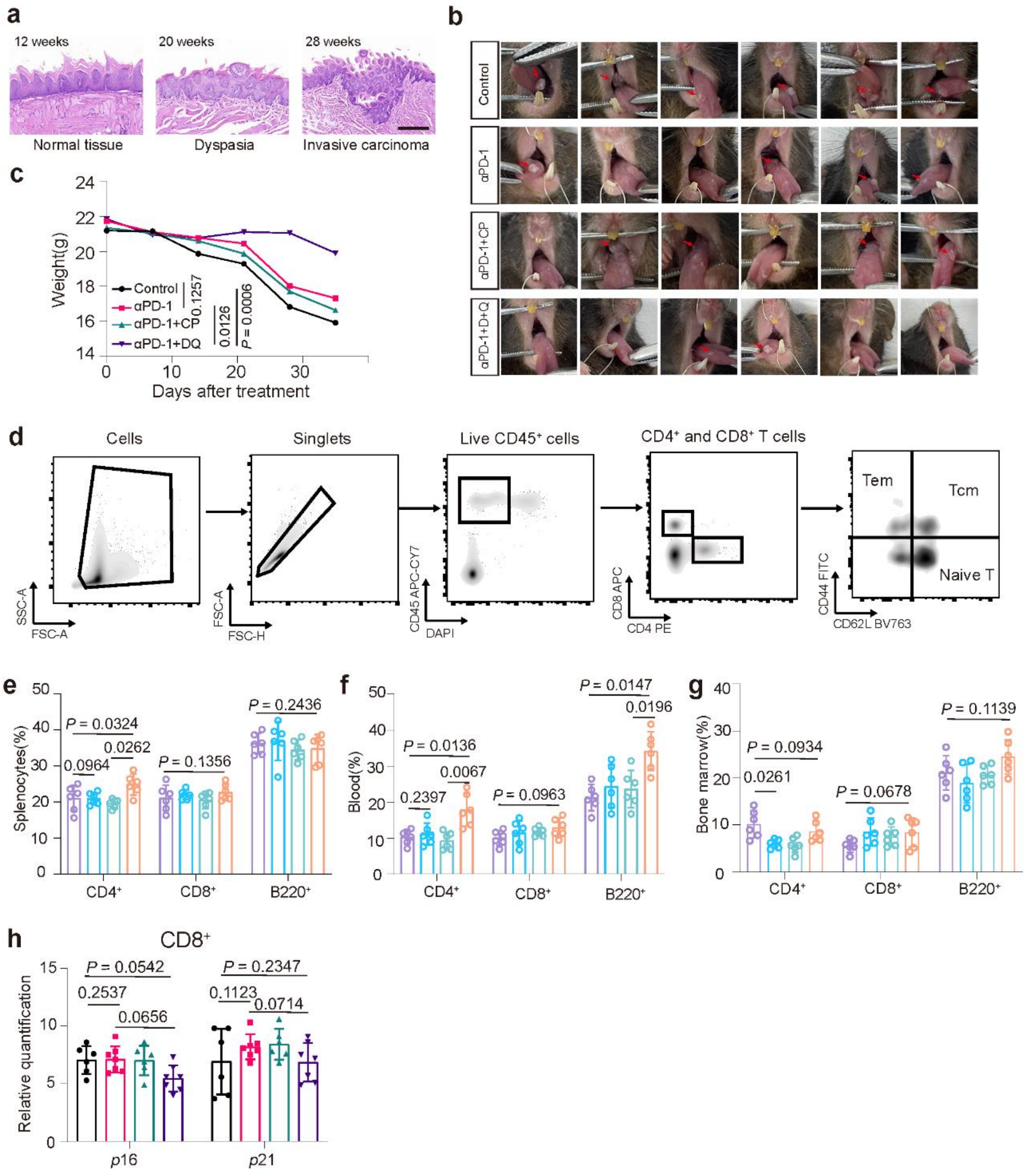

a
12 weeks
20 weeks
28 weeks
Normal tissue
Dyspasia
Invasive carcinoma
b
Control
αPD-1
αPD-1+CP
αPD-1+D+Q
c
Weight(g)
Days after treatment
Control
αPD-1
αPD-1+CP
αPD-1+DQ
0.1257
0.0126
P = 0.0006
d
Cells
Singlets
Live CD45⁺ cells
CD4⁺ and CD8⁺ T cells
Tem
Tcm
Naive T
SSC-A
FSC-A
FSC-H
CD45 APC-CY7
DAPI
CD8 APC
CD4 PE
CD44 FITC
CD62L BV763
e
Splenocytes(%)
P = 0.0324
0.0964
0.0262
P = 0.1356
P = 0.2436
CD4⁺
CD8⁺
B220⁺
f
Blood(%)
P = 0.0136
0.2397
0.0067
P = 0.0963
P = 0.0147
0.0196
CD4⁺
CD8⁺
B220⁺
g
Bone marrow(%)
P = 0.0934
0.0261
P = 0.0678
P = 0.1139
CD4⁺
CD8⁺
B220⁺
h
CD8⁺
Relative quantification
P = 0.0542
0.2537
0.0656
P = 0.2347
0.1123
0.0714
p16
p21

**Extended Data Fig. 8 The effect of senolytics combined with αPD-1 on TIME of 4-NQO mice.** **a,** Representative images of tongue sections from mice, fed with 4-nitroquinoline-1-oxide (4-NQO)-containing drinking water for 12, 20, and 28 weeks, stained with hematoxylin and eosin (HE). Scale bar, 50μm. **b,** Images of tongue of mice after receiving Isotype, αPD-1, αPD-1+CP or αPD-1+DQ treatment (n= 6 each group). Red arrows indicate tumor-like nodules. **c,** Line graph illustrating the change in mouse body weight over time after receiving Isotype, αPD-1, αPD-1+CP or αPD-1+DQ treatment (n= 6 per group). **d,** Flow cytometry gating strategy for assessing the proportions of $CD4^+$, $CD8^+$, as well as T cell subtypes. **e-g,** Flow cytometry immunophenotyping of splenic cells, bone marrow Cells and blood showing the frequencies of $CD4^+$, $CD8^+$ and $B220^+$ cells in mice after receiving Isotype, αPD-1, αPD-1+CP or αPD-1+DQ treatment (n= 6 each group). **h,** Senescence markers *p16* and *p21* expression in $CD8^+$ TILs from the mice treated with Isotype, αPD-1, αPD-1+CP or αPD-1+DQ (n=6 per group).

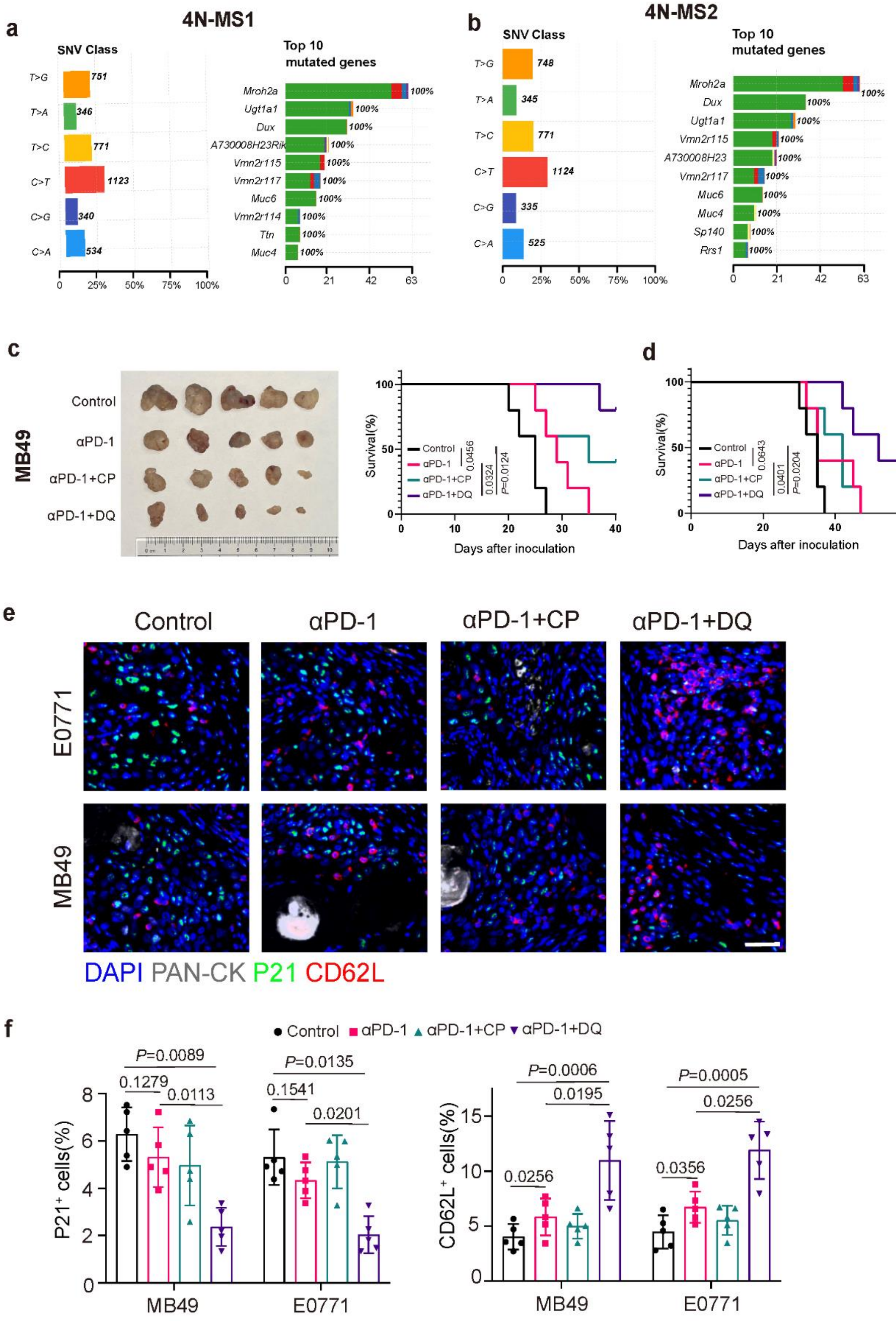
a
4N-MS1
SNV Class
T>G 751
T>A 346
T>C 771
C>T 1123
C>G 340
C>A 534
Top 10 mutated genes
Mroh2a 100%
Ugt1a1 100%
Dux 100%
A730008H23Rik 100%
Vmn2r115 100%
Vmn2r117 100%
Muc6 100%
Vmn2r114 100%
Ttn 100%
Muc4 100%
b
4N-MS2
SNV Class
T>G 748
T>A 345
T>C 771
C>T 1124
C>G 335
C>A 525
Top 10 mutated genes
Mroh2a 100%
Dux 100%
Ugt1a1 100%
Vmn2r115 100%
A730008H23 100%
Vmn2r117 100%
Muc6 100%
Muc4 100%
Sp140 100%
Rrs1 100%
c
MB49
Control
αPD-1
αPD-1+CP
αPD-1+DQ
Survival(%)
Days after inoculation
0.0456
0.0324
P=0.0124
d
Survival(%)
Days after inoculation
0.0643
0.0401
P=0.0204
e
Control
αPD-1
αPD-1+CP
αPD-1+DQ
E0771
MB49
DAPI PAN-CK P21 CD62L
f
Control αPD-1 αPD-1+CP αPD-1+DQ
P21+ cells(%)
P=0.0089
0.1279
0.0113
P=0.0135
0.1541
0.0201
MB49
E0771
CD62L+ cells(%)
P=0.0006
0.0195
0.0256
P=0.0005
0.0256
0.0356
MB49
E0771

**Extended Data Fig. 9 αPD-1+DQ enhanced the anti-tumor effect in breast and bladder cancer xenograft models.**

**a-b,** The whole-exome sequencing (WES) of 4N-MS1 cells reveals the classification of Single Nucleotide Variants (SNV Class) and the top 10 genes with the highest mutation frequencies. **c,** Tumor images and survival curves for the MB49 transplant model after receiving Isotype, αPD-1, αPD-1+CP or αPD-1+DQ treatment (n=6 per group). **d,** The survival curves for the E0771 transplant model after receiving Isotype, αPD-1, αPD-1+CP or αPD-1+DQ treatment (n=6 per group). **e-f,** Representative immunofluorescence images and statistical analysis of the senescent marker P21 and the Naïve cell marker CD62L in MB49 and E0771 tumor xenografts after receiving Isotype, αPD-1, αPD-1+CP or αPD-1+DQ treatment (n=5 per group).

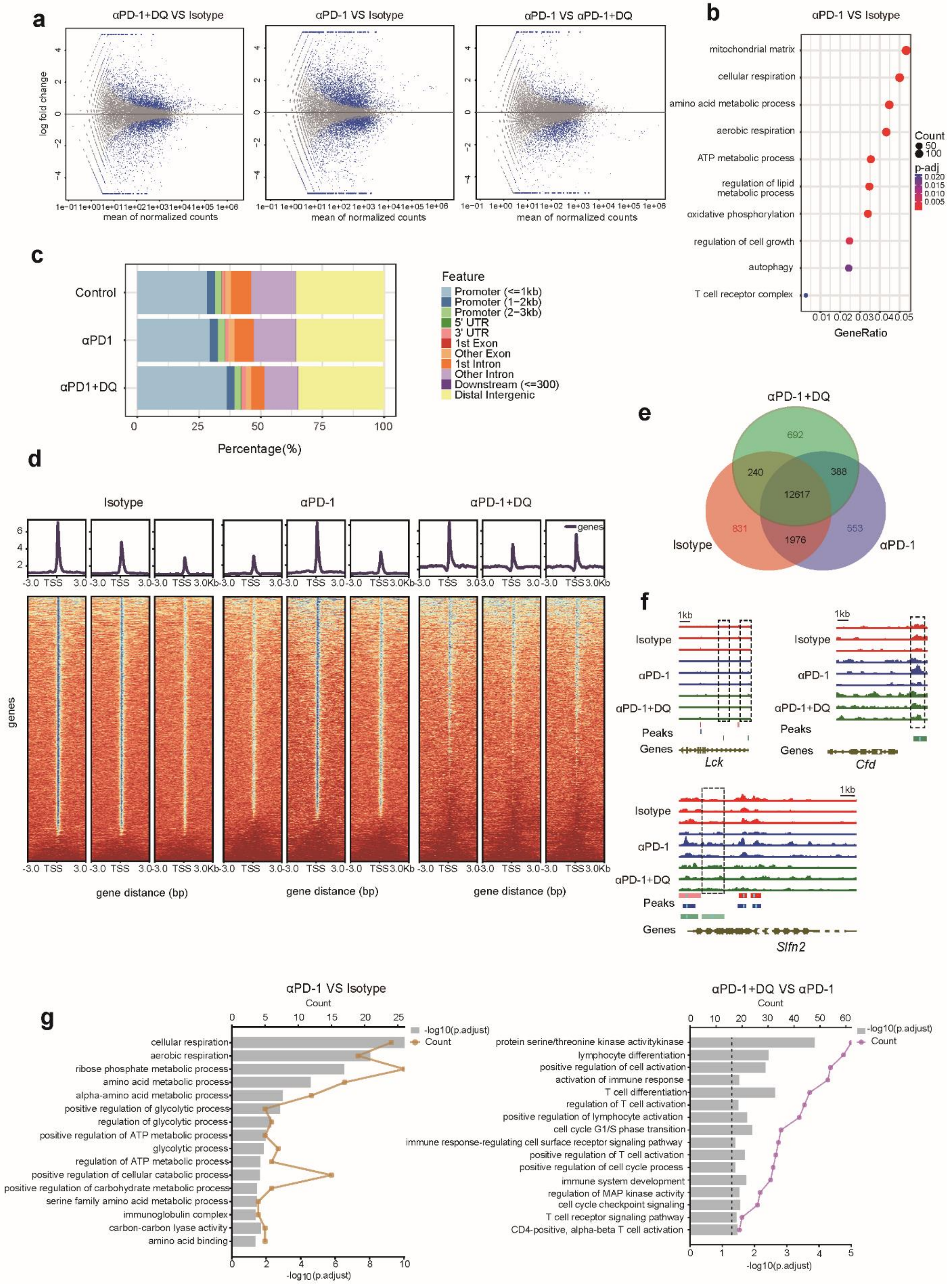

a
αPD-1+DQ VS Isotype
αPD-1 VS Isotype
αPD-1 VS αPD-1+DQ
log fold change
mean of normalized counts
b
αPD-1 VS Isotype
mitochondrial matrix
cellular respiration
amino acid metabolic process
aerobic respiration
ATP metabolic process
regulation of lipid metabolic process
oxidative phosphorylation
regulation of cell growth
autophagy
T cell receptor complex
Count
p-adj
GeneRatio
c
Control
αPD1
αPD1+DQ
Feature
Promoter (<=1kb)
Promoter (1–2kb)
Promoter (2–3kb)
5' UTR
3' UTR
1st Exon
Other Exon
1st Intron
Other Intron
Downstream (<=300)
Distal Intergenic
Percentage(%)
d
Isotype
αPD-1
αPD-1+DQ
genes
gene distance (bp)
e
αPD-1+DQ
692
240
388
12617
831
1976
553
Isotype
αPD-1
f
Isotype
αPD-1
αPD-1+DQ
Peaks
Genes
Lck
Cfd
Slfn2
g
αPD-1 VS Isotype
Count
-log10(p.adjust)
cellular respiration
aerobic respiration
ribose phosphate metabolic process
amino acid metabolic process
alpha-amino acid metabolic process
positive regulation of glycolytic process
regulation of glycolytic process
positive regulation of ATP metabolic process
glycolytic process
regulation of ATP metabolic process
positive regulation of cellular catabolic process
positive regulation of carbohydrate metabolic process
serine family amino acid metabolic process
immunoglobulin complex
carbon-carbon lyase activity
amino acid binding
αPD-1+DQ VS αPD-1
protein serine/threonine kinase activitykinase
lymphocyte differentiation
positive regulation of cell activation
activation of immune response
T cell differentiation
regulation of T cell activation
positive regulation of lymphocyte activation
cell cycle G1/S phase transition
immune response-regulating cell surface receptor signaling pathway
positive regulation of T cell activation
positive regulation of cell cycle process
immune system development
regulation of MAP kinase activity
cell cycle checkpoint signaling
T cell receptor signaling pathway
CD4-positive, alpha-beta T cell activation

**Extended Data Fig. 10 Transcriptomic and epigenetic dynamics of CD4 naive T cells following the αPD-1+DQ therapy.**

**a,** Volcano plot of differentially expressed genes between αPD-1+DQ and Isotype groups (left), αPD-1 and Isotype groups (middle), αPD-1+DQ and αPD-1 groups (right). **b,** DEGs (Log2Foldchanges > 0.5) of CD4 naïve T cells in tumors between αPD-1 and Isotype groups enriched GO terms. **c,** Proportions of the ATAC-seq peak regions representing various genome annotations identified in Isotype, αPD-1+DQ and αPD-1 groups. **d,** The heatmap of different annotated genes in Isotype, αPD-1 and αPD-1+DQ groups from ATAC-seq. **e,** Venn diagram of peaks of CD4 naïve T cells in Isotype, αPD-1 and αPD-1+DQ groups. **f,** ATAC-seq tracks showing the representative genes chromatin accessibility in the *Lck*, *Cfd*, and *Slfn2* loci for CD4 naïve T cells in Isotype, αPD-1+DQ and anti-PD-1 groups. **g,** RNA-seq and ATAC-seq detected GO terms that were simultaneously enriched in the αPD-1 group and Isotype group (left) and GO terms that were enriched in open chromatin regions unique to the αPD-1+DQ group and αPD-1group (right).

### Supplementary Figures 1-6:

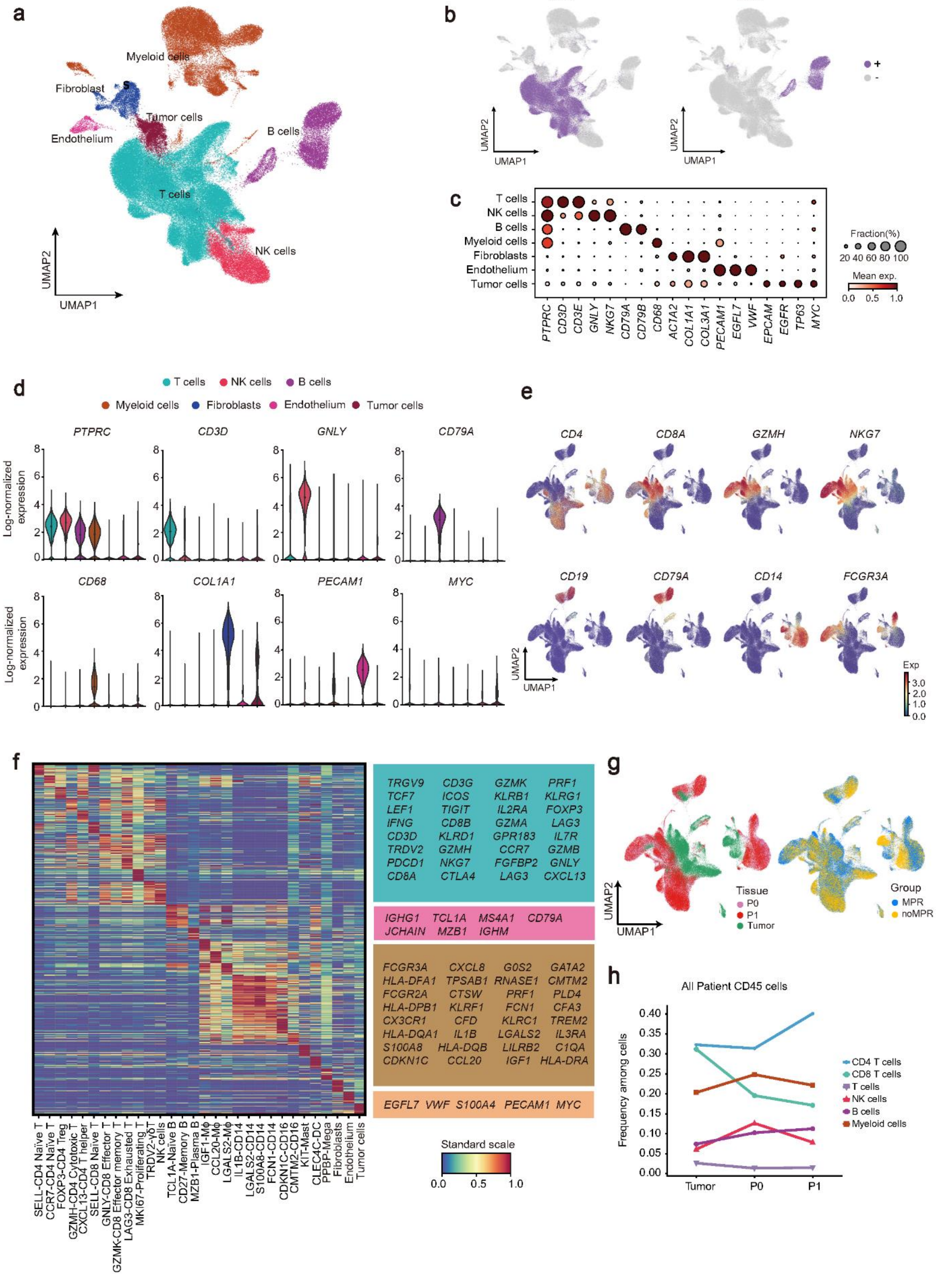

**Supplementary Figure 1. Single-cell profiling of all cells in HNSCC.**

**a**, UMAP plot of all cells that passed quality control. all cell types, defined by 7 unique clusters, are annotated and marked by color code. **b**, Productive TCR (left) and BCR (right) detection projected onto UMAP. **c**, Dot plot of marker genes in distinct the cell types. **d**, Normalized expression of differentially expressed genes between all cell type. Violin plot top and bottom lines indicate range of normalized expression; width indicates number of cells at the indicated expression level. **e**, Normalized expression of selected markers defining the immune cell clusters. For all UMAP plots, red indicates high log-normalized expression; blue indicates low. **f**, The expression patterns of signature genes in distinct tumor-infiltrating all cell clusters. **g**, UMAP embedding of single-cell transcriptional profiles from each tissue sampled in the study (left), from different groups (right). **h**, Mean frequencies of immune cell populations in tumor, P0, and P1.

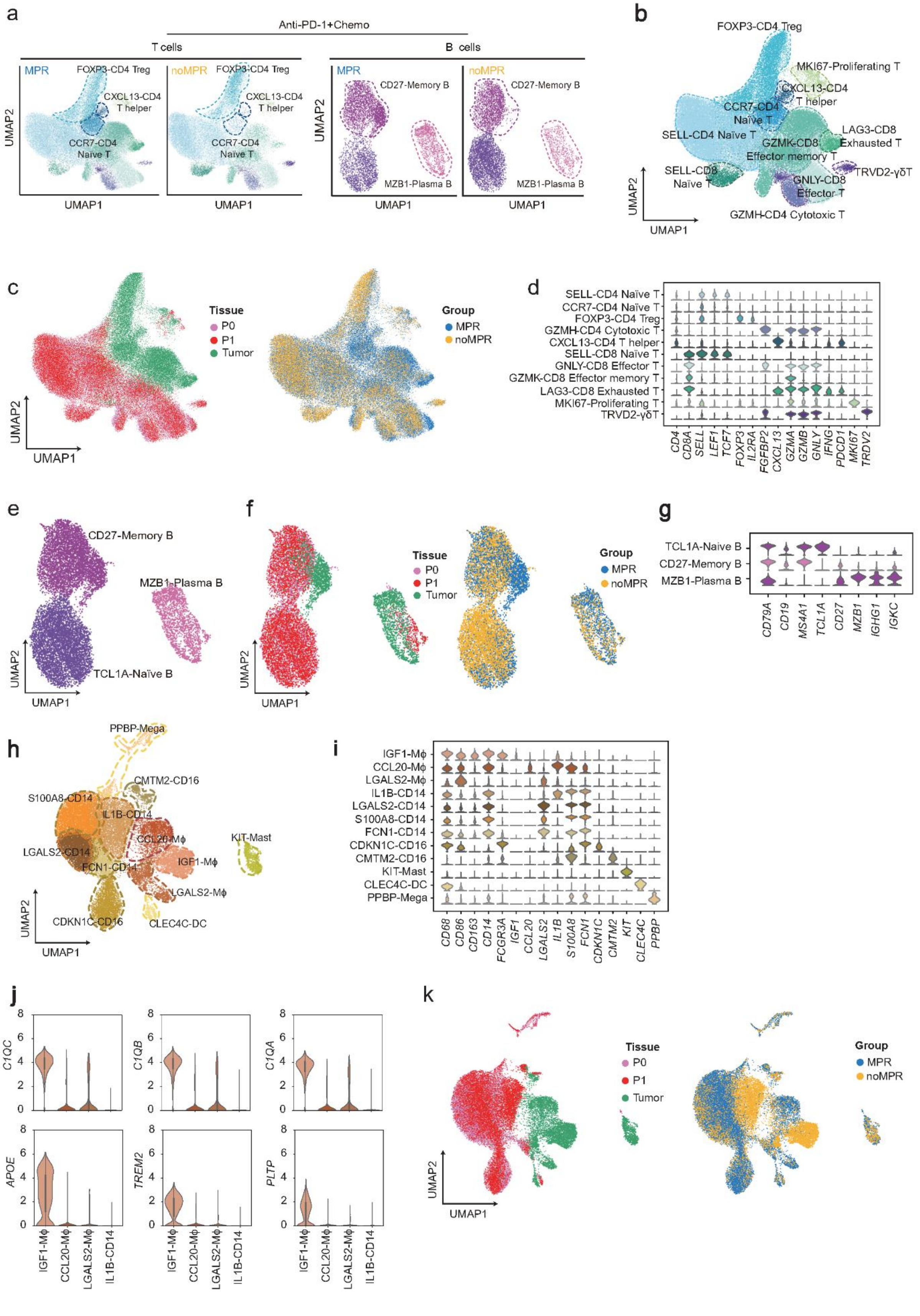
a
Anti-PD-1+Chemo
T cells
B cells
MPR
noMPR
FOXP3-CD4 Treg
CXCL13-CD4 T helper
CCR7-CD4 Naïve T
CD27-Memory B
MZB1-Plasma B
UMAP1
UMAP2
b
FOXP3-CD4 Treg
MKI67-Proliferating T
CXCL13-CD4 T helper
CCR7-CD4 Naïve T
SELL-CD4 Naïve T
LAG3-CD8 Exhausted T
GZMK-CD8 Effector memory T
SELL-CD8 Naïve T
TRVD2-γδT
GNLY-CD8 Effector T
GZMH-CD4 Cytotoxic T
c
Tissue
P0
P1
Tumor
Group
MPR
noMPR
d
SELL-CD4 Naïve T
CCR7-CD4 Naïve T
FOXP3-CD4 Treg
GZMH-CD4 Cytotoxic T
CXCL13-CD4 T helper
SELL-CD8 Naïve T
GNLY-CD8 Effector T
GZMK-CD8 Effector memory T
LAG3-CD8 Exhausted T
MKI67-Proliferating T
TRVD2-γδT
CD4 CD8A SELL LEF1 TCF7 FOXP3 IL2RA FGFBP2 CXCL13 GZMA GZMB GNLY IFNG PDCD1 MKI67 TRDV2
e
CD27-Memory B
MZB1-Plasma B
TCL1A-Naïve B
f
g
TCL1A-Naive B
CD27-Memory B
MZB1-Plasma B
CD79A CD19 MS4A1 TCL1A CD27 MZB1 IGHG1 IGKC
h
PPBP-Mega
CMTM2-CD16
S100A8-CD14
IL1B-CD14
CCL20-Mϕ
KIT-Mast
LGALS2-CD14
FCN1-CD14
IGF1-Mϕ
LGALS2-Mϕ
CDKN1C-CD16
CLEC4C-DC
i
IGF1-Mϕ
CCL20-Mϕ
LGALS2-Mϕ
IL1B-CD14
LGALS2-CD14
S100A8-CD14
FCN1-CD14
CDKN1C-CD16
CMTM2-CD16
KIT-Mast
CLEC4C-DC
PPBP-Mega
CD68 CD86 CD163 CD14 FCGR3A IGF1 CCL20 LGALS2 IL1B S100A8 FCN1 CDKN1C CMTM2 KIT CLEC4C PPBP
j
C1QC
C1QB
C1QA
APOE
TREM2
PLTP
0 2 4 6 8
IGF1-Mϕ
CCL20-Mϕ
LGALS2-Mϕ
IL1B-CD14
k
Tissue
P0
P1
Tumor
Group
MPR
noMPR
UMAP1
UMAP2

**Supplementary Figure 2. Differential gene expression analysis for all immune cells.**

**a**, UMAP plots showing the distributions T cells and B cells between MPR and noMPR groups. **b**, UMAP shows the distribution of T cell clusters. **c**, UMAP embeds single-cell transcriptional profiles of T cells from each tissue sampled in the study (left), and from different groups (right). **d**, Stacked violin plot of normalized expression between T cell clusters markers. **e**, UMAP shows the distribution of B cell clusters. **f**, UMAP embeds single-cell transcriptional profiles of B cells from each tissue sampled in the study (left), and from different groups (right). **g**, Stacked violin plot of normalized expression between B cell clusters markers. **h**, UMAP showing the distribution of myeloid cell clusters. **i**, Stacked violin plot of normalized expression between myeloid cell clusters markers. **j**, Violin plot of normalized expression genes of IGF1 Mφ, CCL20 Mφ, LGALS2 Mφ and IL1B Monocyte. The top and bottom lines of the violin plot represent the range of normalized expression; the width represents the number of cells at the specified expression level. **k**, UMAP displays myeloid cells from each tissue sampled in the study (left), and from different groups (right).

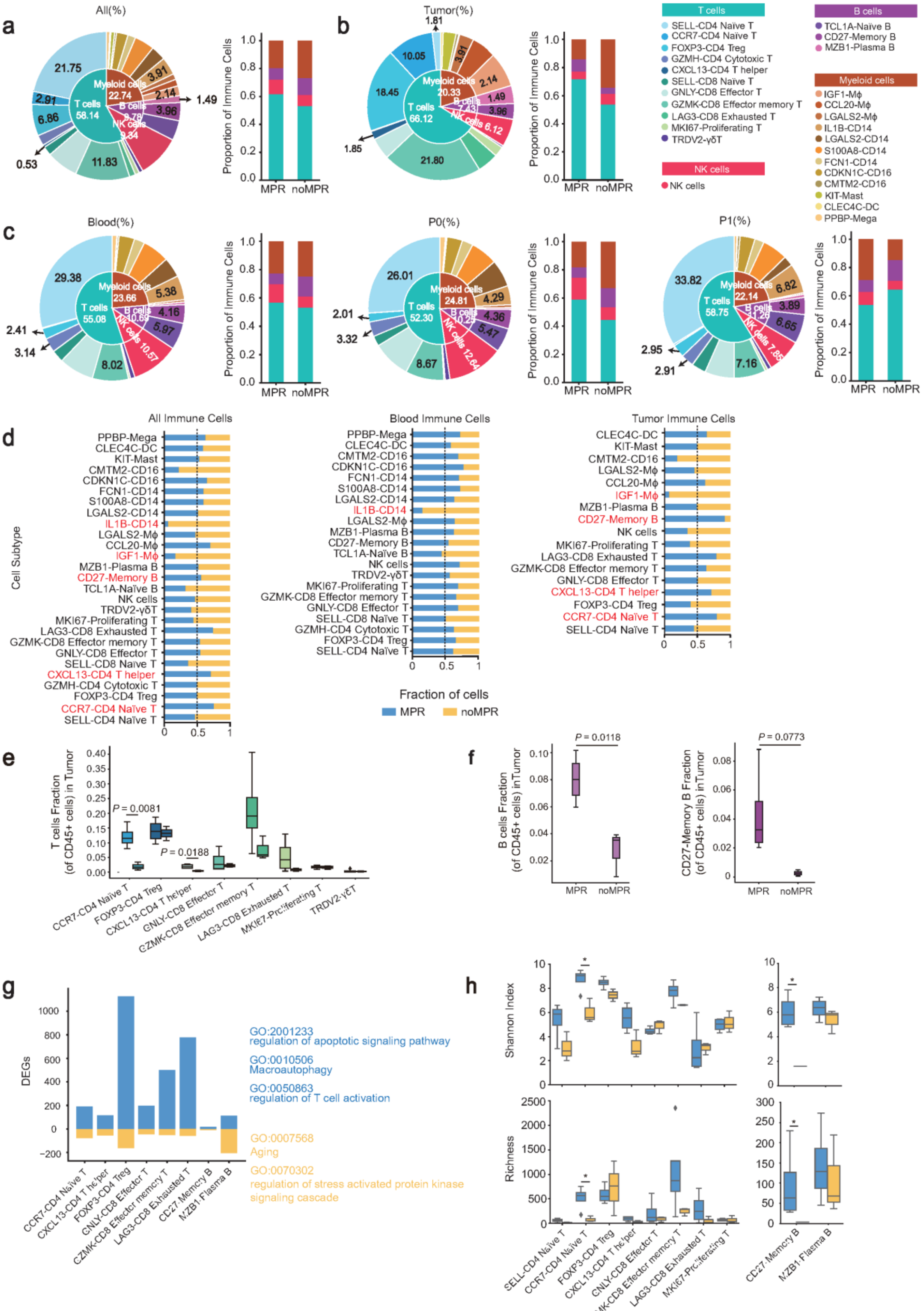

a
All(%)
21.75
2.91
6.86
0.53
11.83
T cells 58.14
Myeloid cells 22.74
B cells 9.78
NK cells 9.34
3.91
2.14
3.96
1.49
Proportion of Immune Cells
MPR
noMPR
b
Tumor(%)
1.81
10.05
18.45
1.85
21.80
T cells 66.12
Myeloid cells 20.33
B cells 7.43
NK cells 6.12
3.91
2.14
1.49
3.96
T cells
SELL-CD4 Naïve T
CCR7-CD4 Naïve T
FOXP3-CD4 Treg
GZMH-CD4 Cytotoxic T
CXCL13-CD4 T helper
SELL-CD8 Naïve T
GNLY-CD8 Effector T
GZMK-CD8 Effector memory T
LAG3-CD8 Exhausted T
MKI67-Proliferating T
TRDV2-γδT
NK cells
NK cells
B cells
TCL1A-Naïve B
CD27-Memory B
MZB1-Plasma B
Myeloid cells
IGF1-Mϕ
CCL20-Mϕ
LGALS2-Mϕ
IL1B-CD14
LGALS2-CD14
S100A8-CD14
FCN1-CD14
CDKN1C-CD16
CMTM2-CD16
KIT-Mast
CLEC4C-DC
PPBP-Mega
c
Blood(%)
29.38
2.41
3.14
8.02
T cells 55.08
Myeloid cells 23.66
B cells 10.69
NK cells 10.57
5.38
4.16
5.97
P0(%)
26.01
2.01
3.32
8.67
T cells 52.30
Myeloid cells 24.81
B cells 10.25
NK cells 12.64
4.29
4.36
5.47
P1(%)
33.82
2.95
2.91
7.16
T cells 58.75
Myeloid cells 22.14
B cells 11.26
NK cells 7.85
6.82
3.89
6.65
d
All Immune Cells
Blood Immune Cells
Tumor Immune Cells
Cell Subtype
Fraction of cells
MPR
noMPR
e
T cells Fraction (of CD45+ cells) in Tumor
P = 0.0081
P = 0.0188
CCR7-CD4 Naïve T
FOXP3-CD4 Treg
CXCL13-CD4 T helper
GNLY-CD8 Effector T
GZMK-CD8 Effector memory T
LAG3-CD8 Exhausted T
MKI67-Proliferating T
TRDV2-γδT
f
B cells Fraction (of CD45+ cells) in Tumor
P = 0.0118
CD27-Memory B Fraction (of CD45+ cells) in Tumor
P = 0.0773
MPR
noMPR
g
DEGs
CCR7-CD4 Naïve T
CXCL13-CD4 T helper
FOXP3-CD4 Treg
GNLY-CD8 Effector T
GZMK-CD8 Effector memory T
LAG3-CD8 Exhausted T
CD27-Memory B
MZB1-Plasma B
GO:2001233
regulation of apoptotic signaling pathway
GO:0010506
Macroautophagy
GO:0050863
regulation of T cell activation
GO:0007568
Aging
GO:0070302
regulation of stress activated protein kinase signaling cascade
h
Shannon Index
Richness
SELL-CD4 Naïve T
CCR7-CD4 Naïve T
FOXP3-CD4 Treg
CXCL13-CD4 T helper
GNLY-CD8 Effector T
GZMK-CD8 Effector memory T
LAG3-CD8 Exhausted T
MKI67-Proliferating T
CD27-Memory B
MZB1-Plasma B

**Supplementary Figure 3. Immune cells proportion between MPR and noMPR patients.**

**a**, The pie charts show the proportion of the immune cells' clusters from tumors and peripheral blood. The bar plot shows the proportion of the immune cells' clusters from tumors and peripheral blood. **b**, The pie charts show the proportion of major immune cell types and corresponding subtypes from tumors. The bar plot shows the proportion of the immune cells' clusters from tumor. **c**, The pie charts show the proportion of major immune cell clusters and corresponding subtypes from peripheral blood, P0 and P1. The bar plot shows the proportion of the immune cells clusters from peripheral blood, P0 and P1. **d**, Comparing the proportion distribution of different immune cell clusters between MPR and noMPR group in tumors and peripheral blood (left), only in peripheral blood (middle) and only in tumors (right). **e**, Box plot of the percentage of the T cell clusters in HNSCC tumors between MPR and noMPR groups. The boxed plot on the left of each T cell cluster is the MPR group, and on the right is noMPR group. Box middle lines, median; box limits, upper and lower quartiles. Black lines with different lengths indicate which two groups were compared. Wilcoxon signed-rank test. **f**, Box plot of the percentage of the overall B cells and $CD27^{+}$Memory B cells in HNSCC tumors between MPR and noMPR groups. Box middle lines, median; box limits, upper and lower quartiles. Black lines with different lengths indicate which two groups were compared. Wilcoxon signed-rank test. **g**, Distribution of genes upregulated in all T cell and B cell clusters in MPR and noMPR groups respectively. On the right is the GO terms enriched for each group of up-regulated genes. **h**, Estimation of size and diversity of TCR repertoires for T cell clusters and BCR repertoires for B cell clusters between MPR and noMPR groups in tumors. The number of TCR and BCR clonotypes estimated by Shannon index (Methods) is a metric used to quantify the diversity of detected TCR clonotypes and BCR clonotypes - a higher index means greater diversity (upper). Richness (Materials and Methods) also measures the diversity of the TCR repertoire and the BCR repertoire - higher scores indicate greater diversity (bottom). Data are expressed as mean ± s.e.m. $^{*}P < 0.05$, $^{**}P < 0.01$, $^{***}P < 0.001$.

a

T cell Kaplan-Meier plotter

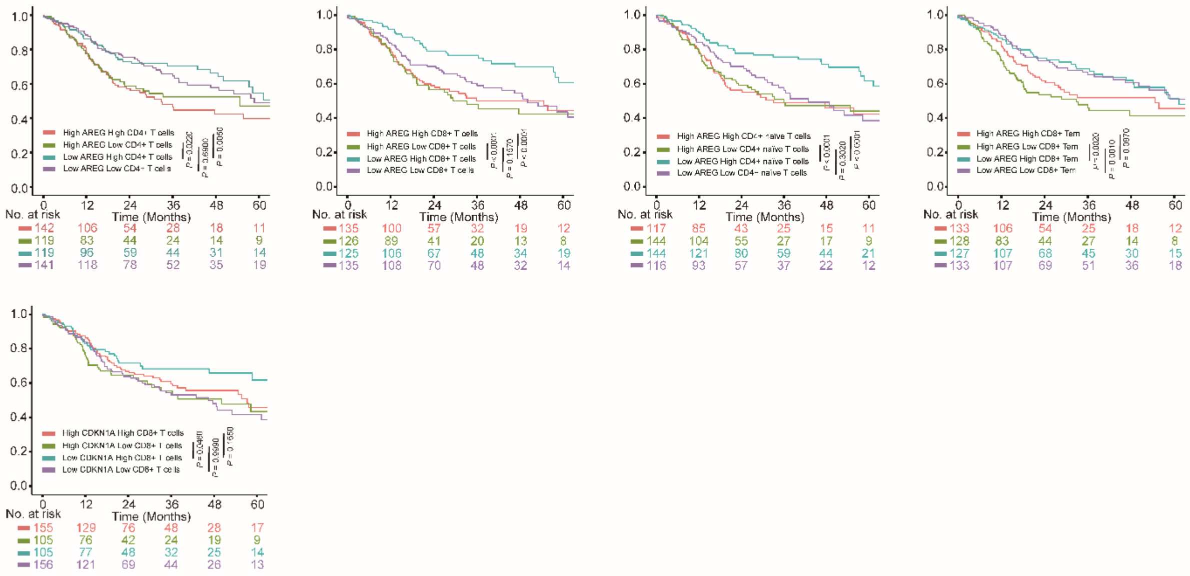

b

B cell Kaplan-Meier plotter

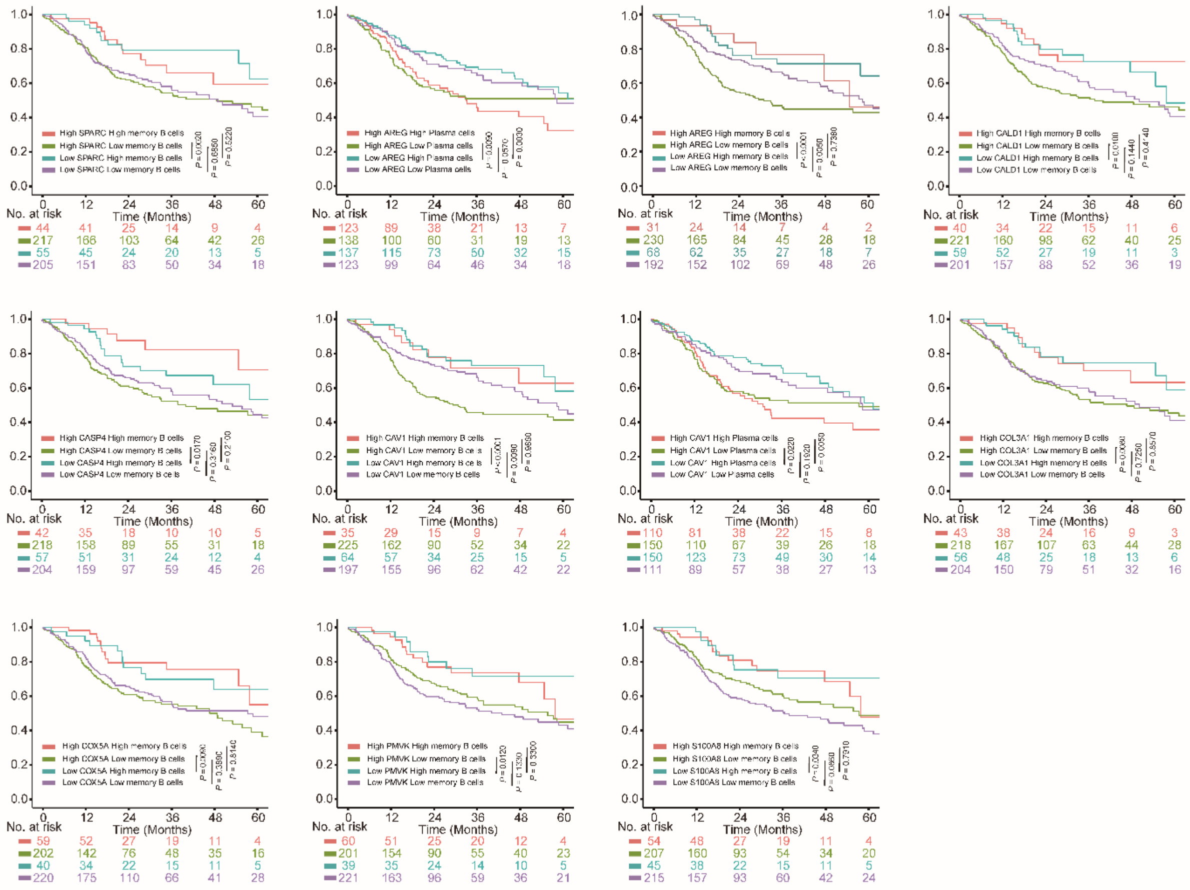

**Supplementary Figure 4. Survival curve between the expression levels of senescent genes and multiple T and B cells in TCGA samples.**

**a**, The survival curve depicts the relationship between the expression levels of *AREG* and *CDKN1A* and the survival period in multiple T-cell subpopulations with high and low abundance in HNSCC TCGA samples. **b**, The survival curve depicts the relationship between the expression levels of *SPARC*, *AREG*, *CALD1*, *CASP4*, *CAV1*, *COL3A1*, *COX5A*, *PMVK* and *S100A8* and the survival period in multiple B-cell subpopulations with high and low abundance in HNSCC TCGA samples.

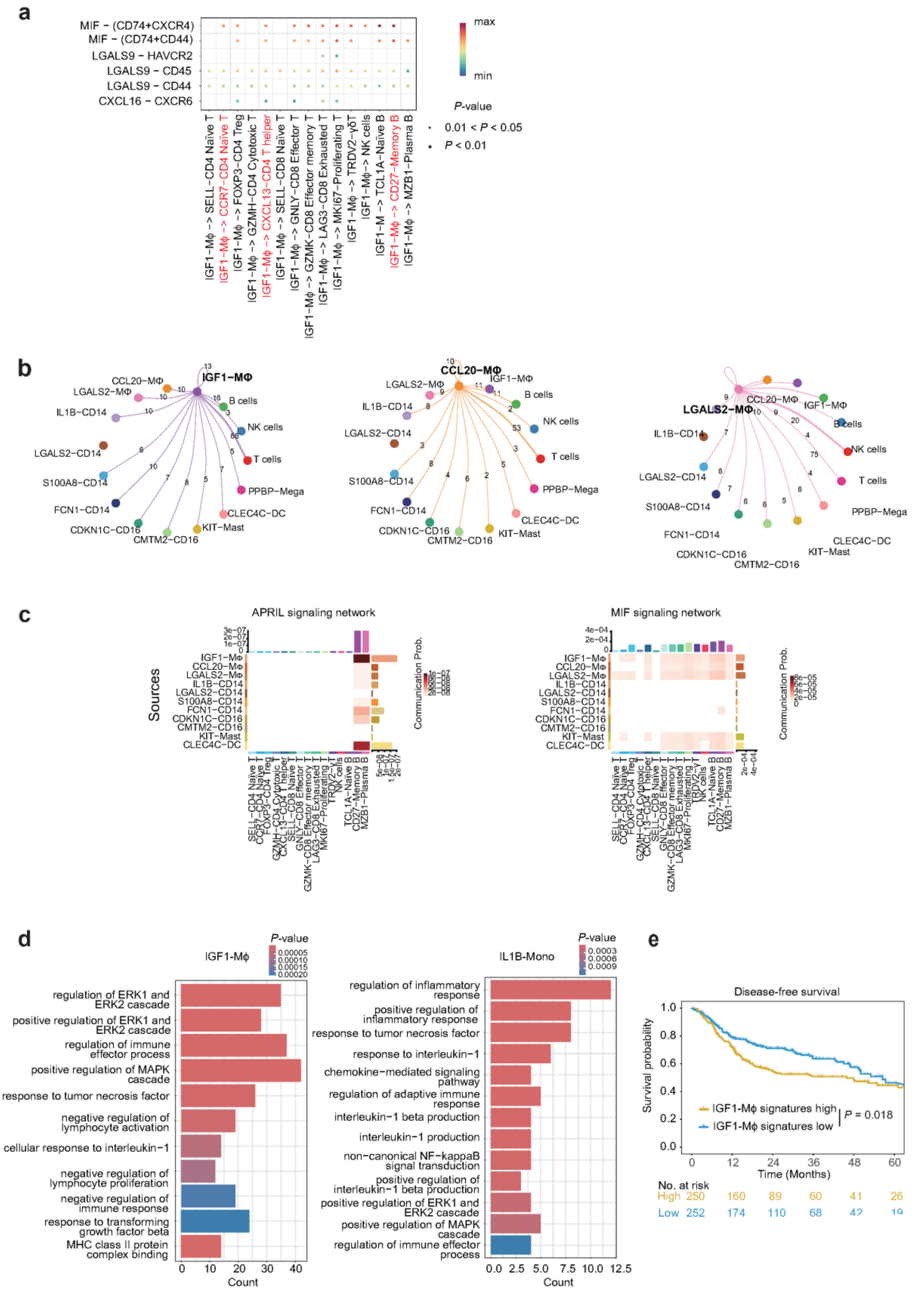

a
MIF − (CD74+CXCR4)
MIF − (CD74+CD44)
LGALS9 − HAVCR2
LGALS9 − CD45
LGALS9 − CD44
CXCL16 − CXCR6
max
min
P-value
0.01 < P < 0.05
P < 0.01
b
IGF1−MΦ
CCL20−MΦ
LGALS2−MΦ
B cells
NK cells
T cells
PPBP−Mega
CLEC4C−DC
KIT−Mast
CMTM2−CD16
CDKN1C−CD16
FCN1−CD14
S100A8−CD14
LGALS2−CD14
IL1B−CD14
c
APRIL signaling network
MIF signaling network
Sources
Communication Prob.
d
IGF1-Mϕ
IL1B-Mono
P-value
regulation of ERK1 and ERK2 cascade
positive regulation of ERK1 and ERK2 cascade
regulation of immune effector process
positive regulation of MAPK cascade
response to tumor necrosis factor
negative regulation of lymphocyte activation
cellular response to interleukin−1
negative regulation of lymphocyte proliferation
negative regulation of immune response
response to transforming growth factor beta
MHC class II protein complex binding
regulation of inflammatory response
positive regulation of inflammatory response
response to tumor necrosis factor
response to interleukin−1
chemokine−mediated signaling pathway
regulation of adaptive immune response
interleukin−1 beta production
interleukin−1 production
non−canonical NF−kappaB signal transduction
positive regulation of interleukin−1 beta production
positive regulation of ERK1 and ERK2 cascade
positive regulation of MAPK cascade
regulation of immune effector process
Count
e
Disease-free survival
Survival probability
Time (Months)
IGF1-Mϕ signatures high
IGF1-Mϕ signatures low
P = 0.018
No. at risk
High 250 160 89 60 41 26
Low 252 174 110 68 42 19

**Supplementary Figure 5. Interaction between myeloid cells and T/B cells.**

**a**, Dot plot showing the expression of ligand-receptor pairs highly expressed in IGF1 Mφ and T cell and B cell clusters in tumors. **b**, Strength of *IGF1*, *CCL20* and *LGALS2* interaction between macrophages, T cells and B cells and other myeloid cells in tumors and blood. **c**, Heat map of myeloid cells interaction pathways with T cells and B cells in tumors and blood. **d**, GO terms enriched in IGF1 Mφ and IL1B Monocyte, including ERK, MAPK and other pathways that regulate senescence and inflammation. Adjusted p value < 0.05. **e**, Low expressions of IGF1 Mφ signature gene were significant associated with better disease-free survival. Log-rank test (two-sided).

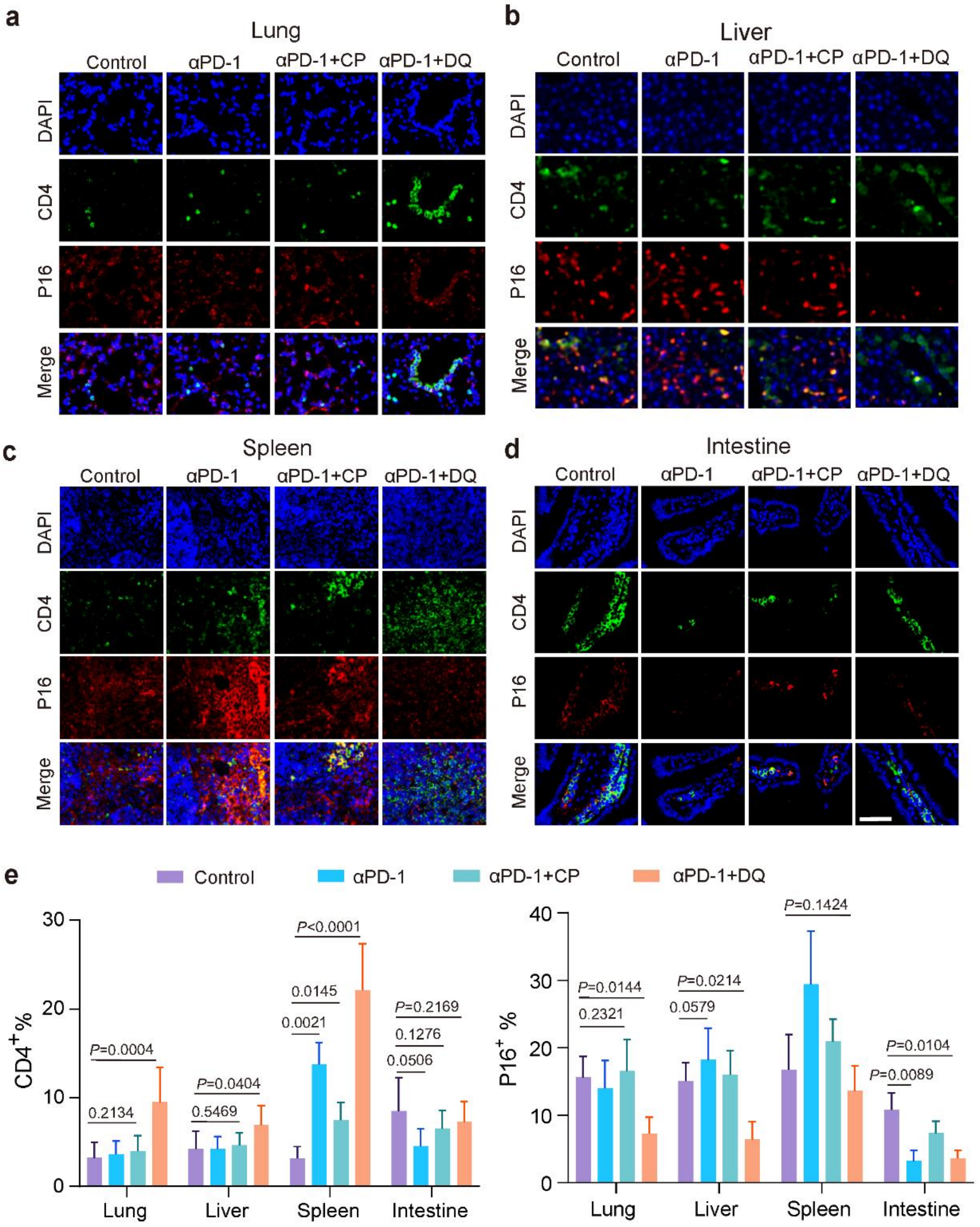
a
Lung
Control
αPD-1
αPD-1+CP
αPD-1+DQ
DAPI
CD4
P16
Merge
b
Liver
Control
αPD-1
αPD-1+CP
αPD-1+DQ
DAPI
CD4
P16
Merge
c
Spleen
Control
αPD-1
αPD-1+CP
αPD-1+DQ
DAPI
CD4
P16
Merge
d
Intestine
Control
αPD-1
αPD-1+CP
αPD-1+DQ
DAPI
CD4
P16
Merge
e
Control
αPD-1
αPD-1+CP
αPD-1+DQ
CD4+%
P=0.0004
0.2134
P=0.0404
0.5469
P<0.0001
0.0145
0.0021
P=0.2169
0.1276
0.0506
0
10
20
30
Lung
Liver
Spleen
Intestine
P16+ %
P=0.0144
0.2321
P=0.0214
0.0579
P=0.1424
P=0.0104
P=0.0089
0
10
20
30
40
Lung
Liver
Spleen
Intestine

**Supplementary Figure 6. Senolytics combined with αPD-1 reduced the expression of the aging marker P16 in multiple organs.**

**a-d,** Representative images of CD4 (green) and P16 (red) staining in lung (**a**), liver (**b**), spleen (**c**), and intestinal (**d**) sections from 4-NQO induced HNSCC mice after receiving Isotype, αPD-1, αPD-1+CP or αPD-1+DQ treatment (n= 6 mice per group for Isotype and αPD-1 group, n= 7 mice per group for αPD-1+CP and αPD-1+DQ group). Scale bar, 10μm. **e,** Quantitative immunostaining of CD4 and P16. Statistical analysis was performed using one-way ANOVA with Tukey's multiple-comparison test (**e**).

### CLINICAL STUDY PROTOCOL

| | |
|---|---|
| **Protocol Title:** | A Prospective, Single-arm, Phase II Clinical Study of Sintilimab (Anti-PD-1 Antibody) Combined with Carboplatin and Nab-paclitaxel in Patients with Resectable Oral or Oropharyngeal Squamous Cell Carcinoma |
| **Protocol Identifier:** | OOC-001 |
| **Investigation Agents:** | Sintilimab |
| **Indication:** | Resectable cT1-T4aN0-3M0 Oral or Oropharyngeal Squamous Cell Carcinoma |
| **Sponsor:** | Sun Yat-Sen Memorial Hospital, Sun Yat-Sen University<br>No. 107 Yanjiangxi Road, Yuexiu District, Guangzhou |
| **Version No.:** | Version 2.2/February 1, 2022 |

Note: This is an English translation of the protocol. The trial is conducted following the original version of the protocol wrote in Chinese.

| Confidentiality Statement |
|---|
| The information contained in this document (especially unpublished data) is the proprietary property of sponsor. The confidential information in this document is provided to you (as an investigator, potential investigator, or consultant) for review by you, your staff, and relevant Ethics Committee. This document may not be disclosed to any other individuals without the prior written approval of sponsor unless for the purpose of providing necessary information to obtain informed consents from candidate patients. |

### SYNOPSIS

This study plans to enroll 51 patients with resectable oral or oropharyngeal squamous cell carcinoma, and preoperatively use Sintilimab, carboplatin, and albumin-bound paclitaxel. Tumor tissue and paracancerous tissue of patients will be collected to observe the changes of imaging and pathology before and after treatment. At the same time, clinical information of patients, such as pathological grade, stage, treatment, prognosis, serology, imaging, etc., will be collected to evaluate the safety and feasibility of Sintilimab combined with carboplatin and albumin-bound paclitaxel for neoadjuvant treatment of resectable oral and oropharyngeal squamous cell carcinoma.

### LIST OF ABBREVIATIONS AND TERMS

| **Abbreviations** | **Definition** |
|---|---|
| CI | Confidence Interval |
| CRs | Complete Responses |
| CTLA-4 | Cytotoxic T-lymphocyte-associated Protein 4 |
| DFS | Disease-free Survival |
| ECOG | Eastern Cooperative Oncology Group |
| OSCC | oral squamous cel1 carcinoma |
| OSPCC | oropharyngeal squamous cel1 carcinoma |
| HR | Hazard Ratio |
| ICIs | Immune Checkpoint Inhibitors |
| irAEs | Immune-related Adverse Events |
| MPR | Major Pathologic Response |
| ORR | Objective Response Rate |
| OS | Overall Survival |
| pCR | Pathological Complete Response |
| PD-1 | Programmed Death Receptor 1 |
| PD-L1 | Programmed Cell Death 1 Ligand 1 |
| PD-L2 | Programmed Cell Death Ligand-2 |
| TRAEs | Treatment-related Adverse Events |
| PTX | Paclitaxel |

### 1. Introduction

#### 1.1 Background on Oral or Oropharyngeal Squamous Cell Carcinoma

Since the twentieth century, with the aging of the population and population growth, cancer incidence and mortality rates have increased rapidly worldwide, and malignant tumors have become the most threatening diseases to human health and life. According to the latest global cancer statistics GLOWBOCAN2018 report, the standardized incidence rate of cancer in China is 201.7 per 100,000 people, while the world average is 197.9 per 100,000 people, and China's standardized incidence rate of cancer ranks 68th globally, and the report also shows that the number of new-onset patients of oral squamous cell carcinoma (OSCC) is as high as 350,000 per year, and the number of deaths exceeds 350,000 per year. Reports also show that oral squamous cel1 carcinoma (OSCC) has a global incidence of 350,000 patients and more than 170,000 deaths per year, while oropharyngeal squamous cel1 carcinoma (OPSCC) has a global incidence of 90,000 patients and more than 50,000 deaths per year [1]. In China, patients with oral cavity squamous carcinoma and oropharyngeal squamous carcinoma are facing a severe survival situation. According to the data of China Cancer Statistics Bureau in 2015, 48,100 new cases of oral cavity squamous carcinoma and oropharyngeal squamous carcinoma were found in China, which accounted for 1% of the systemic malignant tumors, and the number of deaths was as high as 22,100, with a trend of increasing year by year [2]. Risk factors for oral cavity squamous carcinoma and oropharyngeal squamous carcinoma include smoking, alcohol consumption, betel nut chewing, bacterial and viral infections, and malnutrition. In the past decades, the main treatments for oral cavity and oropharyngeal squamous carcinoma include surgery, radiotherapy and chemotherapy, however, the five-year survival rate of the patients still remains around 50%, and even worse for patients with intermediate and advanced stages, which is only around 27%. Therefore, how to improve the clinical treatment effect of oral cavity and oropharyngeal squamous carcinoma, improve the prognosis of patients, and enhance the quality of patients' survival has been a difficult problem that needs to be solved urgently.

#### 1.2. Current Neoadjuvant Therapy for Oral or Oropharyngeal Squamous Cell Carcinoma

##### 1.2.1. Platinum-based chemotherapy regimen for Oral or Oropharyngeal Squamous Cell Carcinoma

Chemotherapy is the main method of non-surgical treatment of head and neck tumors. Paclitaxel (PTX) is a taxane diterpenoid compound with a very unique anti-tumor mechanism. Drug studies have shown that its anti-tumor mechanism is mainly to change the normal cell cycle. PTX can directly act on the cell microtubule network through passive diffusion through the cell membrane, increase the number and polymerization rate of microtubule dimers, inhibit the normal formation of spindles, and block the cell cycle at the G2M cycle, thereby causing abnormal mitosis of tumor cells. By inducing the cascade reaction of Raf-1, the expression of p53 in some cells is increased, causing tumor cell cycle to be blocked and die in the G1 phase. In addition,

PTX induces phosphorylation of Bcl-2 protein, which can inhibit its binding with Bax protein to form heterodimers, increase the level of free Bax, and create favorable conditions for cell apoptosis. Albumin bound paclitaxel (nab-PTX) is a nanoparticle that combines PTX with human albumin to form particles with an average diameter of 130 μm based on ordinary PTX. It uses the biological mechanism of tumor nutrient uptake to make the anticancer drug accumulate at the tumor site. Compared with ordinary PTX, nab-PTX increases the PTX concentration in the tumor stroma and does not require anti-allergic pretreatment before treatment. It has the characteristics of high efficiency and low toxicity [20, 21]. Carboplatin is the main platinum drug used to treat oral cancer and oropharyngeal squamous cell carcinoma. It has a specific inhibitory effect on the cell cycle. When the drug enters the cell, it can irreversibly bind to DA to hinder the replication process of DA, thereby inhibiting cell proliferation and achieving an anti-cancer effect.

Taxol combined with carboplatin is the first-line chemotherapy for recurrent or metastatic oral cancer and oropharyngeal squamous cell carcinoma. Several studies have shown that induction chemotherapy with carboplatin combined with paclitaxel can provide complete remission rates and partial remission rates of 8% to 33% and 50% to 85% in oral cancer and oropharyngeal squamous cell carcinoma, respectively [22-30]. In addition, PFS and OS are always between 60-80%. In many such prospective studies, patients tolerated induction chemotherapy with carboplatin combined with paclitaxel and subsequent concurrent chemoradiotherapy with minimal toxicity.

##### 1.2.2. Anti-PD-1/Anti-PD-L1 Therapy for Oral or Oropharyngeal Squamous Cell Carcinoma

In recent years, tumor immune checkpoint therapy has gradually become a hot topic in research and development, and has continuously made great breakthroughs. Unlike cytotoxic drugs or monoclonal antibodies or small molecule tyrosine kinase inhibitors targeting tumor driver genes, tumor immune checkpoint therapy does not act directly on tumor cells, but blocks the inhibitory signals of T cell proliferation and activation, relieves the immune escape mechanism of tumor cells, restores T cell activity, and improves the effective recognition and killing of tumor cells by T cells [3]. At present, tumor immune checkpoint targets that have shown significant clinical efficacy include programmed death receptor and programmed death receptor 1/ligand 1 (PD-1/PD-L1) and cytotoxic T lymphocyte anti-4 (CTLA-4). Among them, immune checkpoint inhibitors targeting PD1/PD-L1 have better clinical application prospects due to their better safety and wider indications [4]. Programmed death receptor-1 (PD-1, CD279) is a type 1 transmembrane protein with a molecular weight of 55kD and is a member of the CD28 family of cellular co-stimulatory molecules, which also includes CD28, CTLA-4, ICOS, and BTLA.

PD-1 contains an intracellular membrane-proximal immunoreceptor tyrosine-based inhibitory motif (TM) and a membrane-distal immunoreceptor tyrosine-based switch motif (TSM). Two specific ligands for PD-1 have been identified: PD-L1 (B7-H1/CD274) and PD-L2 (B7DC/CD273). Both PD-L1 and PD-L2 have been shown to downregulate T cell activation when bound to PD-1 in both mouse and human systems. PD-1 transmits negative signals by recruiting SHP-2 to phosphorylated tyrosine residues in the TSM cytoplasmic region. Immune checkpoint inhibitors (ICIS) are a class of broadly effective immunotherapies that block inhibitory immune checkpoint pathways to reactivate immune responses to cancer. PD-L1, which is usually expressed by tumor cells, connects to PD-1 protein, which can be expressed by T cells, resulting in the inhibition of T cell immune response, which is one of the mechanisms of tumor immune escape [5-7]. Neoadjuvant immune checkpoint therapy has achieved significant clinical efficacy in melanoma, non-small cell lung cancer, bladder cancer and glioblastoma [8-14]. In 2016, PD-1 blockers nivolumab and pembrolizumab were approved by FAD for patients with recurrent/metastatic head and neck squamous cell carcinoma, including oral and oropharyngeal squamous cell carcinoma, with a response rate of approximately 20%, and overall survival benefit compared with chemotherapy [15, 16].

In recent years, immune checkpoint inhibitors such as anti-PD-1 monoclonal antibodies have been rapidly developed in advanced head and neck squamous cell carcinoma. In the checkmate-141 study, compared with standard single-agent systemic treatment, nivolumab significantly prolonged the overall survival of patients, and the one-year OS rate increased from 16.6 to 36% (HR, 0.70) [17]. In the keynote-040 study, it was also proved that pembrolizumab could improve the median OS of patients from 6.9 months to 8.4 months (R, 0.80) compared with standard treatment. At the same time, more significant benefits were observed in the population with PD-L1 positive and CPS ⩾ 1 [18]. In a phase II clinical trial in 2020, it was demonstrated that patients who received nivolumab before surgery had a 50% volume response, a 53% pathological downgrade, and a 50% pathological response. The median follow-up was 14.2 months, the 1-year progression-free survival rate was 85%, and the overall survival rate was 89%, demonstrating the feasibility and safety of preoperative use of the PD-1 blocker nivolumab in patients with resectable oral squamous cell carcinoma and oropharyngeal cancer [19].

For locally advanced oral and oropharyngeal squamous cell carcinoma, surgical resection of the primary tumor and lymph node drainage, followed by risk-adapted adjuvant radiotherapy, with or without platinum-based chemotherapy, or initially confirmed concurrent chemoradiotherapy, remains the main treatment, but its efficacy is limited. Locally advanced squamous cell carcinoma of the oral cavity and oropharynx is usually treated with surgical resection, followed by adjuvant therapy based on pathological analysis. Despite advances in surgery and radiotherapy for patients with oral and oropharyngeal squamous cell carcinoma, their prognosis remains relatively poor. Most patients have locally advanced disease and are at high risk of recurrence, so it is promising to explore the clinical application of immunotherapy in the first-line and locally advanced oral and oropharyngeal squamous cell carcinoma. At present, a better understanding of emerging immunotherapies, including appropriate patient selection, treatment sequencing, response monitoring, adverse event management, and biomarker testing, can gradually improve patient outcomes and quality of life.

#### 1.3. Background Information on Sintilimab

Sintilimab (R&D code: IBI308) is a recombinant fully human IgG type PD-1 monoclonal antibody. It is a Class 1 new drug independently developed by Innovent Biologics (Suzhou) Co., Ltd. (hereinafter referred to as "Innovent"), which can specifically bind to the PD-1 molecules on the surface of T lymphocytes, thereby blocking the PD 1PD-L1 pathway that causes tumor immune tolerance, reactivating the anti-tumor activity of T lymphocytes and achieving the purpose of treating tumors. Sintilimab targets the same target as nivolumab and pembrolizumab, but has different amino acid sequences. The effect of sintilimab in blocking the PD-1 pathway has been verified in multiple preclinical in vitro tests. The results of various preclinical pharmacodynamics, animal pharmacokinetics and toxicology studies have shown that sintilimab has the characteristics of clear target, reliable cell line source, good drug stability, etc., and has shown good activity in various completed preclinical studies. In September 2016, the Phase Ia dose escalation trial of sintilimab (study code CIBI308A101-1a) was launched. In Phase I, subjects with advanced solid tumors who had failed standard treatment were enrolled. The dose increase decision followed the classic "3+3" design, and four dose levels (1mg/kg, 3mg/kg, 200mg and 10mgkg) were evaluated. After the 1mg/kg dose group was completed, the subjects were randomly assigned 1:1 to undergo independent evaluation of the 3g/kg and 200mg dose groups. The DLT observation period is 28 days after the first dose of each dose group. Only subjects who have completed the DLT observation period can enter the subsequent treatment of sintilimab once every two weeks (1m/kg, 3mg/kg and 10mg/kg) or once every three weeks (200mg) until disease progression, intolerable toxicity, withdrawal of informed consent, or other reasons for stopping study treatment (whichever occurs first).

In a pharmacodynamic study of patients with advanced solid tumors, a single dose of 1 mg/kg (N=3) of Sintilimab can quickly (24 hours) saturate (mean $\geqslant$ 95%) the PD-1 receptor on the surface of CD3+T cells in the peripheral blood of solid tumor subjects, and can maintain the occupancy level during the study period (28 days) when the concentration continues to decrease and during continuous multi-dose treatment. The PD-1 occupancy results of the 3 mg/kg (N=3), 200 mg (N=3), and 10 mg/kg (N=3) dose groups are similar to those of 1 mg/kg, indicating that the PD-1 receptor occupancy level is not dose- and concentration-dependent within the dose range of 1-10 mg/kg.

Based on the previous kinetic and pharmacodynamic results, as well as acceptable safety events, and taking into account potential individual differences, the dose of Sintilimab used in subsequent studies is 200 mg Q3W. Currently, Sintilimab has conducted multiple phase II clinical studies in different tumor types. Based on the results of a multicenter, single-arm phase II clinical study (ORIENT-1) in relapsed or refractory classical Hodgkin's lymphoma, Sintilimab was officially approved for marketing by the National Medical Products Administration (NMPA) on December 24, 2018 for the treatment of relapsed or refractory classical Hodgkin's lymphoma after at least two lines of systemic chemotherapy.

As of October 16, 2018, a total of 540 tumor patients in 5 studies were treated with Sintilimab. The overall safety characteristics are similar to those of anti-PD-1 monoclonal antibodies approved abroad. The incidence of adverse reactions of all grades in 540 patients treated with Sintilimab was 86.1%, and the adverse reactions with an incidence of $\geqslant$ 10% included fever, anemia, increased aspartate aminotransferase, increased alanine aminotransferase, fatigue, and decreased white blood cell count. The incidence of grade 3 and above adverse reactions was 30.6%, and the incidence of $\geqslant$1% included lung infection, anemia, increased lipase, decreased platelet count, pneumonia, decreased neutrophil count, hyponatremia, increased γ-glutamyl transferase, infectious pneumonia, upper gastrointestinal bleeding, and lymphocyte count.

#### 1.4. Study Rationale

Based on current domestic and foreign literature, immunotherapy combined with chemotherapy has achieved great success in many cancer types such as lung cancer and nasopharyngeal carcinoma, and other tumor types are also exploring their efficacy through clinical trials. For example, a multicenter, randomized, bilingual study published in the New England Journal of Medicine evaluated the efficacy of PD-L1 inhibitors Atezolizumab and Nab-paclitaxel versus placebo plus Nab-paclitaxel in the treatment of locally advanced or metastatic triple-negative breast cancer. The overall OS was encouraging in the PD-L1 positive population.

In 2017, an international open clinical trial OPTIMA-(NCT03107182) was launched to evaluate the tumor response rate in patients with oral cancer and oropharyngeal squamous cell carcinoma using a combination of nivolumab (PD-1 inhibitor), carboplatin and albumin-bound paclitaxel. At the same time, in another phase II open clinical trial LCC1621 (NCT03174275), Durvalumab (Durvalumab. ) combined with carboplatin and albumin-bound paclitaxel was used preoperatively for untreated stage -V head and neck cancer patients (including oral squamous cell carcinoma and oropharyngeal squamous cell carcinoma). The primary outcome indicator was to evaluate the pathological complete remission rate after induction chemotherapy. The trial is still going smoothly.

Based on the above research background, we believe that PD-1 (Sintilimab) combined with carboplatin and albumin-bound paclitaxel has a good application prospect for oral squamous cell carcinoma and oropharyngeal squamous cell carcinoma. To this end, we designed this prospective, single-arm, phase || clinical study. The aim is to explore the efficacy and safety of PD-1 (Sintilimab) combined with carboplatin and albumin-bound paclitaxel in patients with resectable oral squamous cell carcinoma or oropharyngeal squamous cell carcinoma before surgery, to provide a new approach to further improve the prognosis of patients with oral squamous cell carcinoma and oropharyngeal squamous cell carcinoma, and to provide valuable information for prospective clinical trials of anti-PD-1 and other immunotherapies combined with chemotherapy in the perioperative and advanced disease settings of patients with oral and oropharyngeal squamous cell carcinoma.

### 2. Study Objectives

#### 2.1. Main Research Indicators:

●Safety: During the study and follow-up period, the severity of adverse events related to neoadjuvant therapy will be graded and evaluated according to NCICTCAE (version 5.0);

Study-related adverse events (90 days after the first administration of Sintilimab or 30 days after surgery, whichever occurs later);

Effectiveness:

●Major pathological response (MPR) rate, defined as the proportion of subjects with ≤10% viable tumor cells in the resected specimen among all subjects.

#### 2.2. Secondary objectives:

●Disease-free survival (DFS): The time from tumor disappearance to confirmed tumor recurrence or death (whichever occurs first);

● Overall survival (OS), defined as the time from enrollment to the death of the subject due to any cause;

● Radiological response: Imaging assessment was performed using the Response Evaluation Criteria in Solid Tumors 1.1 (RECIST1.1);

- Surgery postponement rate: If surgery cannot be performed within 8 weeks after the last dose of medication, the subject will be included in the category of surgical postponement.

### 3. Study Design

This study is a prospective, single-arm, multicenter, phase II clinical study to evaluate the use of Sintilimab combined with carboplatin and albumin-bound paclitaxel for neoadjuvant treatment of resectable oral or oropharyngeal squamous cell carcinoma. The main target population is patients with surgically resectable oral or oropharyngeal squamous cell carcinoma. The treatment plan is to give Sintilimab to subjects who meet the inclusion criteria before surgery: 200mg, intravenous infusion on the first day of each cycle, one cycle every 3 weeks (Q3W), a total of 24 cycles: At the same time, the administration cycle of carboplatin and albumin-bound paclitaxel is: one cycle every 3 weeks (Q3W), a total of 24 cycles. Carboplatin is administered on the first day of each cycle, 300mg/m$^{2,}$ intravenous drip, drip time ≥1h, albumin-bound paclitaxel 260 mg/m$^2$, administered on the first day of each cycle, intravenous drip 30min. Surgery is scheduled within 22-56 days after neoadjuvant treatment.

The study used RECIST1.1 for imaging assessment, and enhanced CT examinations were performed at baseline and before surgery (before the second dose and within 1 week before surgery), and CT or enhanced CT examinations were performed 30 days after surgery and every 3 months after surgery until 2 years after surgery, disease recurrence or death, or the end of this study. Comprehensive imaging examinations are required at baseline to exclude distant metastasis. During the study, subjects will receive safety follow-up (90 days after the first dose of Sintilimab combined with chemotherapy or 30 days after surgery, whichever occurs later), and postoperative recurrence-free survival follow-up will be performed according to the above cycle.

### 4. Research plan and technical route

#### 4.1. Sample size calculation and statistical analysis methods:

This study is a single-arm, exploratory, fixed sample size study. No strict hypothesis testing is performed. This study plans to enroll 51 cases. The measurement data will be statistically described using mean ± standard deviation or median (minimum, maximum). The counting data will be statistically described using frequency (percentage). All statistical analyses were completed using SAS9.2 (or higher).

#### 4.2. Eligible subjects for this study must meet all of the following criteria:

1. Sign written informed consent before implementing any test-related procedures;

2. 18 years old ≤ age ≤ 80 years old;
3. Cytological or histological diagnosis of oral or oropharyngeal squamous cell carcinoma: The researcher assessed it as surgically resectable oral or oropharyngeal squamous cell carcinoma without distant metastasis. (Based on the NCCN Clinical Practice Guidelines for Head and Neck Cancers (2021. V1)): II-IV;
4. According to the Response Evaluation Criteria in Solid Tumors (RECIST1.1 version), there is at least one radiographically measurable lesion: First-line patients: have not received any systemic anti-tumor treatment for advanced metastatic disease. Patients who have received platinum-containing adjuvant/neoadjuvant chemotherapy or radical chemoradiotherapy for advanced disease are allowed to join this study if the interval between disease progression or recurrence and the end of the last chemotherapy drug treatment is at least 6 months;
5. ECOG score 0-1 points;
6. Expected survival time>3 months;
7. Sufficient organ function, subjects must meet the following laboratory indicators:
1) In the absence of granulocyte colony-stimulating factor in the past 14 days, the absolute neutrophil count (ANC) ≥ 1. 5x$10^9$L;
2) In the absence of blood transfusion in the past 14 days, platelets ≥ 100×$10^9$L;
3) In the absence of blood transfusion or erythropoietin in the past 14 days, hemoglobin>9g/dL;
4) Total bilirubin ≤1. 5×Upper limit of normal (ULN);
5) Aspartate aminotransferase (AST) and alanine aminotransferase (ALT) ≤25×ULN (patients with liver metastasis) Patients with ALT or AST ≤ 5×ULN are allowed);
6) Serum creatinine ≤ 1.5 × ULN and creatinine clearance (calculated by Cockcroft-. Gault formula) ≥ 60 ml/min;
7) Good coagulation function, defined as international normalized ratio (NR) or coagulation transfer time (PT) ≤ 1.5 times ULN;
8) Normal thyroid function, defined as thyroid stimulating hormone (TSH) within the normal range. If the baseline TSH exceeds the normal range, subjects with total T3 (or FT3) and FT4 within the normal range can also be enrolled;
9) Myocardial enzyme spectrum within the normal range (if the researcher comprehensively judges that the simple laboratory abnormality is not clinically significant, it is also allowed to be enrolled);
8. For female subjects of childbearing age, a urine or serum pregnancy test should be performed within 3 days before the first dose of study drug (Day 1 of Cycle 1) and the result should be negative. If the urine pregnancy test result cannot be confirmed as negative, a blood pregnancy test is required. Non-childbearing women are defined as those who have been postmenopausal for at least 1 year, or have undergone surgical sterilization or hysterectomy:

9. If there is a risk of pregnancy, all subjects (both male and female) must use contraceptive measures with an annual failure rate of less than 1% throughout the treatment period until 120 days after the last dose of study drug (or 180 days after the last dose of chemotherapy drug).

##### 4.3. Subjects who meet the following criteria are not eligible for inclusion in this study:

1. Patients diagnosed with other malignant diseases other than head and neck squamous cell carcinoma within 5 years before the first dose (excluding radically cured basal cell carcinoma of the skin, squamous cell carcinoma of the skin, and or radically resected in situ temples);
2. Patients currently participating in interventional clinical research treatment, or receiving other research drugs or using research devices within 4 weeks before the first dose;
3. Patients who have previously received the following therapies: anti-PD-1, anti-PD-L1 or anti-PD-L2 drugs or drugs targeting another micro- or synergistic inhibitory T cell receptor (e.g., CTLA-4, OX-40, CD137);
4. Patients who have received systemic treatment with traditional Chinese medicine or immunomodulatory drugs (including thymosin, interferon, interleukin, except for local use to control pleural effusion) with indications for oral or oropharyngeal squamous cell carcinoma within 2 weeks before the first dose;
5. Patients who have had systemic treatment required (e.g., use of disease-modifying drugs, glucocorticoids or immunosuppressants) within 2 years before the first dose. Replacement therapy (e.g., thyroxine, insulin, or physiologic glucocorticoids for adrenal or pituitary insufficiency) is not considered systemic therapy:
6. Patients who are receiving systemic glucocorticoid therapy (excluding topical glucocorticoids via nasal spray, inhalation or other routes) or any other form of immunosuppressive therapy within 7 days before the first dose of the study;

Note: Physiological doses of glucocorticoids (≤10mg/day of prednisone or equivalent drugs) are allowed;

7. Patients with clinically uncontrollable pleural effusion/peritoneal effusion (patients who do not need to drain the effusion or who do not have a significant increase in the effusion after stopping drainage for 3 days can be enrolled);
8. Patients with known allogeneic organ transplantation (except corneal transplantation) or allogeneic hematopoietic stem cell transplantation;
9. Patients with known allergies to the active ingredients or excipients of the study drugs Sintilimab, carboplatin, and albumin-bound paclitaxel;
10. Patients who have not fully recovered from toxicity and/or complications caused by any intervention before starting treatment (i.e., ≤1 grade or reaching baseline, excluding fatigue or alopecia):

11. Patients with known history of human immunodeficiency virus (HIV) infection (i.e., positive HIV1/2 antibodies):
12. Untreated active hepatitis B (defined as HBsAg positive and HBV-DNA copy number detected greater than the upper limit of normal value of the laboratory of the research center);
Note: Hepatitis B subjects who meet the following criteria can also be enrolled:
1) HBV viral load <1000 copies/ml (200U/ml) before the first dose, the subject should receive anti-HBV treatment during the entire study treatment period to avoid viral reactivation
2) For subjects with anti-HBc (+), HBsAg (-), anti-HBs (-) and HBV viral load (-), no preventive anti-HBV treatment is required, but close monitoring of viral reactivation is required
13. Active HCV infected subjects (HCV antibody positive and HCV-RNA level above the detection limit);
14. Live vaccine received within 30 days before the first dose (1st cycle, day 1): day
Note: Injectable inactivated virus vaccine for seasonal influenza within 30 days before the first dose is allowed: but live attenuated influenza vaccine for intravenous use is not allowed;
15. Pregnant or lactating women;
16. Any serious or uncontrolled systemic disease, such as:
1) Abnormalities in rhythm, conduction or morphology on resting electrocardiogram with serious and difficult-to-control symptoms, such as complete left bundle branch block, heart block of more than Ⅱ degree, ventricular arrhythmia or atrial fibrillation;
2) Unstable angina, congestive heart failure, chronic heart failure of New York Heart Association (NYHA) grade ≥2;
3) Any arterial thrombosis, embolism or ischemia within 6 months before treatment, such as myocardial infarction, unstable angina, cerebrovascular accident or transient ischemic attack;
4) Poor blood pressure control (systolic blood pressure>140 mmHg, diastolic blood pressure>90 mmHg);
5) History of non-infectious pneumonia requiring glucocorticoid treatment within 1 year before first administration, or current clinically active interstitial lung disease;
6) Active pulmonary tuberculosis;
7) Active or uncontrolled infection requiring systemic treatment;
8) Clinically active diverticulitis, abdominal abscess, gastrointestinal obstruction;
9) Liver disease such as cirrhosis, decompensated liver disease, acute or chronic active hepatitis;
10) Poorly controlled diabetes (fasting blood glucose (FBG)>10 mmol/L);

11) Patients with urine protein ⩾ + and confirmed 24-hour urine protein quantification >1.0g;
12) Patients with mental disorders and unable to cooperate with treatment;
17. Patients with medical history or disease evidence that may interfere with the test results and prevent the subject from participating in the study, abnormal treatment or laboratory test values, or other situations that the researcher believes are not suitable for inclusion in the group. The researcher team believes that there are other potential risks and are not suitable for participation in this study.

#### 4.4. Efficacy evaluation:

The efficacy indicators of this study are evaluated by the researcher in accordance with RECIST1.1;

##### 4.4.1. Main research indicators:

● Safety:

During the study and follow-up period, the severity of adverse events related to neoadjuvant therapy will be graded and evaluated according to NCI CTCAE (version 5.0): Study-related adverse events (90 days after the first dose of Sintilimab or 30 days after surgery, whichever occurs later):

● Effectiveness: Major pathological response (MPR) rate, defined as the proportion of subjects with ⩽10% viable tumor cells in the resected specimen to the total subjects:

##### 4.4.2. Secondary objectives:

● Disease-free survival (DFS): The time from tumor disappearance to confirmed tumor recurrence or death (whichever occurs first):

● Overall survival (OS), defined as the time from enrollment to the death of the subject due to any cause:

● Radiological response: Imaging assessment was performed using the Response Evaluation Criteria in Solid Tumors 1.1 (RECIST1.1):

● Surgery postponement rate: If surgery cannot be performed within 8 weeks after the last dose of medication, the subject will be included in the category of surgery postponement.

#### 4.5. Criteria for Subjects to Stop Treatment and Exit the Study

##### 4.5.1. Stop Study Treatment

Subjects may stop treatment at any time for any reason, or the investigator may decide whether to stop treatment when any adverse event occurs. In addition, the investigator may stop treatment of the subject if the subject is not suitable for treatment, violates the study protocol, or for management and other safety reasons.

Subjects must stop treatment for any of the following reasons, but may continue to be monitored in the study:

The subject or the subject's legal representative requests that treatment be stopped.

An adverse event occurs that requires treatment to be stopped as specified in the protocol.

Another malignant tumor occurs that requires active treatment.

A concurrent disease occurs that prevents further treatment.

The investigator decides to withdraw the subject from the study.

The subject's serum pregnancy test result is positive.

The subject has poor compliance.

The investigator believes that based on the subject's disease condition or personal circumstances, if the study drug is continued, it will put the subject at unnecessary risk.

Complete the treatment prescribed by the protocol.

##### 4.5.2. Withdrawal from the study

If the subject or the subject's legal representative withdraws the informed consent to participate in the study, the subject must withdraw from the study. If the subject withdraws from the study, he or she will no longer receive treatment or planned visits. Subjects can receive survival follow-up after withdrawal from the study with the subject's consent. If the subject is lost to follow-up, he or she must withdraw from the study.

#### 4.6. Statistical analysis plan

##### 4.6.1. Sample size:

This study is a single-arm, exploratory, fixed sample size study. No strict hypothesis testing is performed. This study plans to enroll 51 cases.

##### 4.6.2. Definition of analysis set:

Full analysis set: According to the intention-to-treat (ITT) principle, all cases enrolled and using the drug at least once are analyzed for efficacy. For case data where the entire treatment process cannot be observed, the last observation data is carried forward to the final result of the trial (LOCF).

Per-protocol set: All cases that meet the trial prescription, have good compliance, do not take prohibited drugs during the trial, and complete the content specified in the case report form. No imputation is performed for missing data.

Statistical analysis of the efficacy of the drug is performed on both AS and PPS.

Safety analysis set: All enrolled cases who have used the trial drug at least once and have safety records after using the drug belong to the safety analysis set. This data set is used for safety analysis.

**4.6.3. Statistical software:**

SAS9.2 (or higher version) statistical software was used for statistical analysis.

**4.6.4. Missing data:**

This trial did not make special treatment for missing data of efficacy indicators, and no missing values were estimated in the safety evaluation.

**4.6.5. Descriptive statistics:**

In this study, unless otherwise specified, data will be summarized using descriptive statistics according to the following general principles.

The median (minimum value, maximum value) will be used for statistical description of measurement data. The frequency (percentage) will be used for statistical description of count data. In addition, the Kaplan Meier method will be used to estimate the PFS survival rate and draw the survival curve.

**4.6.6. Analysis of baseline data:**

The mean, standard deviation, median, maximum value, minimum value, gender, C0G score and other quantitative data are calculated, and the frequency and percentage of qualitative data such as age, height and weight are listed.

**4.6.7. Analysis of influencing factors:**

None

**4.6.8. Sensitivity analysis:**

None

**4.6.9. General methods of statistical analysis:**

This study will conduct statistical analysis on the drug administration in the trial to calculate the drug exposure during the entire trial. Baseline data will be analyzed according to the full analysis set, all efficacy indicators will be analyzed according to the full analysis set and the protocol-compliant set, and the analysis groups will be based on the randomized medication groups: safety analysis will use the safety analysis set, and the analysis groups will be based on the actual medication groups. If a patient receives incorrect medication in a certain cycle, but receives correct medication in other treatment cycles, then the patient will be grouped according to the correct treatment for safety analysis.

The measurement data of each visit will be statistically described using the mean ±

standard deviation or median (minimum, maximum).The count data of each visit in the trial group will be statistically described using frequency (constituent ratio).

1) Major pathological response(MPR) rate: Major pathological response rate, defined

as the proportion of subjects with ≤10% viable tumor cells in the resected specimen to the total number of subjects, 1PR = number of subjects who achieved PR/total number of subjects 100%, and its 95% CI is calculated using binomial distribution.
2) pathological complete response(pCR) rate: pathological complete response rate, defined as the proportion of subjects with no residual viable tumor cells under the microscope and negative lymph nodes to the total number of subjects, pCR = number of subjects achieving pCR / total number of subjects * 100, using binomial distribution to calculate its 95%CI.
3) ORR: objective response rate, defined as the proportion of subjects with complete response (CR) and partial response (PR) to the total number of subjects, ORR = number of subjects with CR + PR / total number of subjects * 100%, using binomial distribution to calculate its 95%CI, the calculation of objective response rate is based on the optimal efficacy evaluation of the tumor confirmed twice during the study.
4) DFS: disease-free survival, the time from the first study drug treatment to the death or recurrence of the subject, the 1-year and 2-year DRS rates are estimated by Kaplan-Meier;
5) OS: the time from the first use of the study drug to the death of the subject, at the end of the study, if the subject is still alive, the known "last survival date of the subject" is used as the loss date. The comparison of OS between groups was performed by log-log test, and the COX proportional hazard model was used to estimate the inter-group R and its 95%CI. The Kaplan Meier method was used to estimate the median OS and its 95%CI, and the survival curve was drawn.
6) Analysis of safety indicators. The safety analysis used the SS population. The main safety indicators included: study-related adverse events (90 days after the first administration of Sintilimab or 30 days after surgery, whichever occurred later): study drug-related adverse events of grade 3 or above, surgical complications: defined as intraoperative and perioperative complications of grade 3 or above or severe complications, and no delayed surgery rate (no surgery for more than 56 days from the first medication date is considered a delayed surgery).

##### 4.7. Informed consent

In this study, no subject shall be enrolled before obtaining a signed written informed consent from the subject. All updated versions of the informed consent and written information will be provided to the subject during the subject's participation. The informed consent should be kept for reference as an important document of the clinical study.

##### 4.8. Research-related ethics

This protocol and the written informed consent and materials directly related to the subject must be submitted to the Ethics Committee, and the study can only be formally carried out after obtaining written approval from the Ethics Committee. The researcher must submit an annual research report to the Ethics Committee at least once a year (if applicable). When the study is terminated and/or completed, the researcher must notify the Ethics Committee in writing. The researcher must promptly report to the Ethics Committee all changes that occur in the research work (such as revisions to the protocol and/or informed consent number), and these changes shall not be implemented without the approval of the Ethics Committee, unless the changes are made to eliminate obvious and direct risks to the subject. In such cases, the Ethics Committee will be notified.

##### 4.9. Confidentiality measures

The information of patients participating in this project will be kept confidential. The results of this project may be published in medical journals, but we will keep the patient's information confidential in accordance with the requirements of the law. Unless required by relevant laws, the patient's personal information will not be disclosed. When necessary, government management departments and hospital ethics committees and their relevant personnel may review the patient's information in accordance with regulations.

### 5. Safety evaluation

##### 5.1 Definition of adverse events

Adverse events (AE) are defined as any unfavorable and unexpected medical events that occur to clinical research subjects since the signing of the informed consent form. Regardless of whether they are causally related to the research drug, they are all judged as adverse events. AE includes but is not limited to the following situations: Exacerbation of the original (before entering the clinical trial) medical condition/disease (including symptoms, signs, and laboratory abnormalities):

Any new adverse medical condition (including symptoms, signs, and newly diagnosed diseases)

Abnormal laboratory results with clinical significance.

##### 5.2 Definition of serious adverse events

Serious adverse events (SAE) are adverse events that meet at least one of the following criteria: Leading to death, except for death caused by progression of the disease of the research indication.

Life-threatening (the definition of "life-threatening" means that when this AE occurs, the subject is at risk of death, and does not include AEs that may cause death if the event worsens).

Requires hospitalization or prolonged hospitalization, excluding the following situations:

√ Rehabilitation institution

√ Nursing home Routine emergency room admission

√ Same-day surgery (such as outpatient/same-day ambulatory surgery)

√ Hospitalization or prolonged hospitalization that is not related to the worsening of

AE is not an SAE in itself. For example: hospitalization for existing diseases, no new adverse events, and no worsening of existing diseases (such as: abnormal laboratory results that persisted before the clinical trial to the present): hospitalization for other reasons (such as: annual routine physical examination): hospitalization stipulated in the trial protocol during the clinical trial (such as: operation according to the requirements of the trial protocol): elective hospitalization not related to the worsening of adverse events (such as elective surgery): scheduled treatment or surgical operation should be recorded in the entire trial protocol and/or the subject's personal baseline data: hospitalization only due to the use of blood products.
Resulting in permanent or severe disability.
Leading to congenital anomalies and birth defects.
Other important medical events: defined as events that harm the subject or require medical intervention to prevent any of the above situations.

##### 5.3 Assessment of adverse events

The investigator will assess all adverse events in accordance with the CI Common Terminology for Adverse Events (CTCAE) Version 5.0. Any adverse event that changes the CTCAE grade will be recorded in the Adverse Event Case Report Form Worksheet. All adverse events, regardless of the CTCAE grade, must be assessed for serious adverse events.
The details of the adverse event assessment are shown in the table below
Table 1 Assessment details of adverse events

| | | |
|---|---|---|
| V5.0 CTCAE classification | Level 1 | Mild; no symptoms or mild symptoms; only clinical or diagnostic manifestations; no intervention required |
| | Level 2 | Moderate; minimal, partial, or noninvasive intervention required; limitations on age-appropriate daily activities |
| | Level 3 | Severe or significant medical event requiring medication but not immediately life-threatening; hospitalization or prolonged hospitalization; disability; limited ability to care for oneself (ADL) |

| | | |
|---|---|---|
| | Level 4 | Life-threatening consequences; urgent intervention required |
| | Level 5 | Deaths related to AEs |
| Severity | Serious adverse events are any of the following adverse events occurring at any dose or during any use of the study drug: | |
| | Causes death; | |
| | Life-threatening; or in the opinion of the investigator, the occurrence of the event puts the subject at risk of immediate death (Note: this does not include adverse events that may lead to death if they occur in a more serious form); | |
| | Causes permanent or severe disability/incapacity (severely interferes with the ability to carry out a normal life); | |
| | Causing or prolonging an existing hospitalization; (Hospitalization is defined as a single hospitalization, regardless of duration, even if the hospitalization is a precautionary measure for continued observation. Note: Hospitalization for a pre-existing condition that was not exacerbated during the study [including hospitalization for elective surgery], does not constitute a serious adverse event. Pre-existing conditions are clinical conditions that were diagnosed prior to the start of study drug and are documented in the patient's medical history) | |
| | Congenital anomalies/birth defects; (offspring of subjects who used the product, regardless of when they were diagnosed); | |
| | Other important medical events; although not resulting in death, not life-threatening, or not requiring hospitalization, which, based on appropriate medical judgment, could endanger the subject and may require medical or surgical intervention to prevent one of the outcomes listed previously (marked † above) may also be considered a serious adverse event. | |
| Duration | Record the start and end dates of the adverse event. If | |

| | | |
|---|---|---|
| | less than 1 day, indicate the appropriate length of time and units. | |
| Take action | Did the study drug adverse event lead to discontinuation of the study drug? | |
| Relationship to study drug | Did the study drug cause the adverse event? A medically qualified investigator is required to provide the results of a causal assessment between the study drug and the adverse event. The investigator will sign/date (with initials) the original document or worksheet to support the causal relationship assessment on the AE form to ensure that a medically qualified causal relationship assessment was performed. This signed document must be retained for the required regulatory time frame. The following criteria are intended to serve as a reference guide to assist investigators in evaluating the relationship between the investigational drug and the adverse event based on the available information.<br>The following elements are used to evaluate the relationship between the study drug and the AE; The greater the correlation between the items and their corresponding elements (in terms of quantity and/or intensity), the greater the possibility that the study drug caused the adverse event; | |
| | Exposure | Is there evidence that subjects were actually exposed to the investigational drug, such as: credible medical history, acceptable compliance assessments (drug counts, diaries, etc.), expected pharmacological effects, drug/metabolite measurements in specimens collected in vivo? |
| | Schedule | Is there a reasonable temporal sequence between the adverse event and the trial drug treatment? Is the timing of |

| | | |
|---|---|---|
| | | the adverse event consistent with a drug-induced adverse event? |
| | Possible Causes | Whether the adverse event cannot be explained by other etiologies, such as underlying disease, other drugs/vaccines, or other host or environmental factors |
| | De-challenge test | Was the study drug discontinued or the dose/exposure/frequency reduced? If yes, did the AE resolve or improve?<br>If yes, the dechallenge test was positive. If no, the dechallenge test was negative.<br>Note: This criterion does not apply if: (1) the adverse event resulted in death or permanent disability; (2) the AE resolved/improved despite continued use of the study drug; (3) the trial was a single-dose drug trial; (4) the study drug was used only once. |
| | Rechallenge test | Has the subject been repeatedly exposed to the study drug in this trial? If so, has the AE recurred or worsened?<br>If so, the rechallenge test is positive. If not, the rechallenge test is negative. Note: This criterion does not apply if: (1) the initial AE resulted |

| | | |
|---|---|---|
| | | in death or permanent disability, or (2) the trial is a single-dose trial, or (3) the study drug is used only once.<br>Note: If a rechallenge test is planned for an adverse event that is serious and possibly caused by the study drug, or if re-exposure to the study drug may pose a serious potential risk to the subject/patient, a rechallenge test is not recommended unless it is considered that continued use of the drug may benefit the patient and there is no available alternative treatment, and it may be performed with the approval of the principal investigator in advance. |
| | Consistency with trial treatment characteristics | Are the clinical/pathological manifestations of the adverse event consistent with previous treatment information on the study drug or pharmacology and toxicology trials of this drug class? |
| The medically qualified investigator will report the results of the relationship assessment on the case report form/worksheet based on his/her best clinical judgment, including consideration of the above-mentioned relationships. | | |

| | |
|---|---|
| Documenting cause and effect | The following table can be used to make a causal link assessment (not all criteria need to be met): |
| related | There is evidence of exposure to study |

| | |
|---|---|
| | drug. The temporal sequence of the AE to the administration of study drug is reasonable. The AE is more likely to be explained by study drug than by other causes. |
| Unrelated | The subject did not receive the study drug or the administration of the study drug was not in a reasonable time relationship to the occurrence of the AE or there was another reason that was more likely to explain the adverse event than the study drug (also applies to subjects who have an overdose but do not experience an associated adverse event) |

### 6. Data Collection and Management

The clinical test reports of patients in this project strictly adopt the national legal measurement units. The information of patient test reports and follow-up records must be complete, including date, record items, record results and normal value range, etc., and relevant personnel should sign. The clinical data and follow-up data of cases are recorded and collected by trained and professional data collection and management personnel, and handed over to the team leader for verification and confirmation before recording in the database.

### 7. Quality Management Plan

This project is composed of a well-structured and professional head and neck tumor clinical researchers and data recorders and data analysts to ensure the implementation of the research plan: The specific projects involved in this project are equipped with specific team leaders and technical personnel. The general project leader and team leader keep in close contact with the leaders of each team. The team leader will feedback information to the general leader at any time. The general leader will check the corresponding information and solve possible problems in time.

### 8. Preliminary assessment of project risks and benefits and risk control plan

#### 8.1. Risks and treatment related to Sintilimab

Patients treated with Sintilimab may experience immune-related adverse reactions, including severe and fatal cases. Immune-related adverse reactions may occur during and after discontinuation of Sintilimab treatment and may involve any tissue or organ. The incidence of all levels of adverse reactions of this product alone was 86.1%, and adverse reactions with an incidence of ≥10% included: fever, anemia, increased aspartate aminotransferase, increased alanine aminotransferase, fatigue, and decreased white blood cell count. The incidence of grade 3 and above adverse reactions was 30.6%, and the incidence of ≥1% included: lung infection, anemia, increased lipase, decreased platelet count, pneumonia, decreased neutrophil count, hyponatremia, Y. Increased glutamyl transferase, infectious pneumonia, upper gastrointestinal bleeding, lymphocyte count.

For suspected immune-related adverse reactions, adequate evaluation should be performed to exclude other causes. Most immune-related adverse reactions are reversible and can be managed by interrupting sintilimab, corticosteroid treatment and/or supportive care. Overall, most grade 3-4 and certain specific grade 2 immune-related adverse reactions require dosing suspension. For grade 4 and certain specific grade 3 immune-related adverse reactions, permanent discontinuation is required.

For grade 3-4 and certain specific grade 2 immune-related adverse reactions, 1-2 mg/kg day prednisone equivalent and other treatments are given until improvement to ≤ grade 1. Corticosteroids need to be gradually reduced for at least one month until they are discontinued. Rapid reduction may cause adverse reactions to worsen or recur. If adverse reactions continue to worsen or do not improve after corticosteroid treatment, non-corticosteroid immunosuppressant therapy should be added.

Any recurrent grade 3 immune-related adverse reactions after monoclonal antibody administration, grade 2-3 immune-related adverse reactions that do not improve to grade 0 within 12 weeks after the last dose (excluding endocrine diseases), and corticosteroids that fail to decrease to ≤10 mg of prednisone equivalent within 12 weeks after the last dose should be permanently discontinued.

| Risks associated with immunotherapy | Grading | Treatment adjustment plan |
|---|---|---|
| pneumonia | Grade 2 | Withhold medication until improvement to grade 0-1 |
| | Grade 3-4 or recurrent grade 2 | Permanently stop the drug |
| Colitis/diarrhea | Grade 2-3 | Withhold medication until improvement to grade 0-1 |
| | Grade 4 | Permanently stop the drug |
| Thyroid disease | Grade 1-2: No symptoms | Give thyroxine |

| | | |
|---|---|---|
| | or mild symptoms | replacement therapy. |
| | Grade3-4: Severe symptoms | Please consult the endocrinology department and give hormone or β-blocker treatment if necessary. |
| hepatitis | Grade 2: aspartate aminotransferase (AST) or alanine aminotransferase (ALT) 3-5 times the upper limit of normal (ULN) or total bilirubin 1.5-3 times ULN | Withhold medication until improvement to grade 0-1 |
| | Grade 3-4, AST or ALT > 5 times ULN, or total bilirubin > 3 times ULN | Permanently stop the drug |
| Nephritis | Grade 2-3 elevated serum creatinine | Withhold medication until improvement to grade 0-1 |
| | Grade 4 increased serum creatinine | Permanently stop the drug |
| Endocrine diseases | Symptomatic grade 2-3 hypothyroidism, grade 2-3 hyperthyroidism, grade 2-3 hypophysitis, grade 2 adrenal insufficiency, grade 3 hyperglycemia or type 1 diabetes mellitus | Withhold medication until improvement to grade 0-1 |
| | Grade 4 hypothyroidism, Grade 4 hyperthyroidism, Grade 4 hypophysitis, Grade 3-4 adrenal insufficiency, Grade 4 hyperglycemia or type 1 diabetes mellitus | Permanently stop the drug |
| Skin Adverse Reactions | Grade 3 rash | Withhold medication until improvement to grade 0-1 |
| | Grade 4 rash, Stevens-Johnson syndrome (SJS) or toxic epidermal necrotic | Permanently stop the drug |

| | | |
|---|---|---|
| | syndrome (TEN) | |
| Thrombocytopenia | Grade 3 | Withhold medication until improvement to grade 0-1 |
| | Grade 4 | Permanently stop the drug |

##### 8.2. Risks and countermeasures of the relevant operation process in the study

Blood collection: Blood collection may cause some discomfort, bleeding or bruising at the puncture site. Small blood stains or swelling may form at the site. In rare cases, fainting or local infection may occur.
MRI scan: For most people, MRI examination is risk-free. However, if the subject has certain objects or devices implanted in the body, such as pacemakers, insulin pumps, ear implants, joint replacements, permanent dentures, piercings or shrapnel, MRI scans may be very dangerous. You may feel claustrophobic during the test because you have to stay still in a small space.
Infusion reaction: If the subject has an infusion-related reaction (IRR), the infusion should be interrupted immediately (for subjects with non-serious IRRs, the infusion rate can be slowed down according to the decision of the investigator) and the subject should be monitored until all symptoms are resolved. Antipyretic analgesics can be used for treatment. If mild to moderate IRR occurs after the first infusion, preventive diphenhydramine or other antihistamines will be used before the next infusion according to local diagnostic and treatment standards. If severe IRR occurs, dexamethasone, diphenhydramine or other antihistamines must be used. If severe infusion-related reactions occur again after preventive steroid use, the study treatment will be permanently terminated.
The researchers will conduct this study in accordance with the protocol, the ethical principles of the Declaration of Helsinki, China's GCP and relevant regulatory requirements.
The research team of this project group has published many SCI articles and regularly participates in cutting-edge academic conferences. These factors provide strong technical guarantee and support for the development of this project.

### CLINICAL STUDY PROTOCOL

| | |
|---|---|
| **Protocol Title:** | A Prospective, Open-label, Single-center, Phase II Single-arm Clinical Study of Tislelizumab Combined with Dasatinib and Quercetin as Neoadjuvant Therapy for Patients with Resectable Head and Neck Squamous Cell Carcinoma. |
| **Protocol Identifier:** | COIS-01 |
| **Investigation Agents:** | Tislelizumab, Dasatinib, Quercetin |
| **Indication:** | Resectable Head and Neck Squamous Cell Carcinoma |
| **Sponsor:** | Sun Yat-Sen Memorial Hospital, Sun Yat-Sen University<br>No. 33 Yingfeng Road, Haizhu District, Guangzhou |
| **Version No.:** | Version 1.1/March 22, 2024 |

Note: This is an English translation of the protocol. The trial is conducted following the original version of the protocol wrote in Chinese.

| **Confidentiality Statement** |
|---|
| The information contained in this document (especially unpublished data) is the proprietary property of sponsor. The confidential information in this document is provided to you (as an investigator, potential investigator, or consultant) for review by you, your staff, and relevant Ethics Committee. This document may not be disclosed to any other individuals without the prior written approval of sponsor unless for the purpose of providing necessary information to obtain informed consents from candidate patients. |

### SYNOPSIS

The new adjuvant treatment model using monotherapy with immune agents has shown promising efficacy in clinical trials; however, some patients do not benefit. Studies indicate that combining immunotherapy with tyrosine kinase inhibitors has demonstrated good efficacy and acceptable toxicity in solid tumors such as head and neck squamous cell carcinoma, endometrial cancer, clear cell renal cell carcinoma, advanced EGFR-mutant NSCLC, and prostate cancer. Quercetin, a flavonoid compound, has been shown to positively modulate the immune system by promoting immune cell proliferation and enhancing overall immune function. Additionally, preclinical studies in gliomas have demonstrated that combining immunotherapy with dasatinib and quercetin is safe and can significantly improve prognosis. Therefore, to avoid the adverse effects associated with chemotherapy and achieve a chemo-free integrated treatment model while enhancing the efficacy of immunotherapy to ultimately improve survival outcomes for head and neck squamous cell carcinoma patients, we propose to conduct a prospective, open-label, single-center, phase II clinical study evaluating the safety and efficacy of the combination of Tislelizumab with Dasatinib and Quercetin in the new adjuvant treatment of resectable head and neck squamous cell carcinoma. This research aims to expand the indications for new adjuvant immunotherapy in solid tumors and provide a new strategy for significantly improving the prognosis of head and neck squamous cell carcinoma patients.

This study plans to enroll 24 participants with resectable head and neck squamous cell carcinoma. We will collect tumor tissues, adjacent tissues, fat, epidermis, whole blood samples, saliva, feces, and urine from the patients. The study aims to observe imaging and pathological changes before and after treatment while also collecting clinical information, such as pathological grading, staging, treatment details, prognosis, and serological and imaging data. The primary endpoint of this study is the major pathologic response rate.

### LIST OF ABBREVIATIONS AND TERMS

| Abbreviations | Definition |
|---|---|
| ADCC | Antibody-dependent Cellular Cytotoxicity |
| ADCP | Antibody-dependent Cellular Phagocytosis |
| ASCO | American Society of Clinical Oncology |
| C1q | Complement 1q |
| CDC | Complement-dependent Cytotoxicity |
| CI | Confidence Interval |
| CPS | Combined Positive Score |
| CRs | Complete Responses |
| CTLA-4 | Cytotoxic T-lymphocyte-associated Protein 4 |
| DCR | Disease Control Rate |
| DFS | Disease-free Survival |
| DOR | Duration of Response |
| ECOG | Eastern Cooperative Oncology Group |
| ESMO | European Society for Medical Oncology |
| FcγR | Gamma Fragment Crystallizable Region Receptors |
| GLP | Good Laboratory Practice |
| HNSCC | Head and Neck Squamous Cell Carcinoma |
| HR | Hazard Ratio |
| ICIs | Immune Checkpoint Inhibitors |
| IPF | Idiopathic Pulmonary Fibrosis |
| irAEs | Immune-related Adverse Events |
| IRC | Independent Review Committee |
| ITIM | Immune Receptor Tyrosine Inhibitory Motif |
| ITSM | Immune Receptor Tyrosine Switch Motif |
| MPR | Major Pathologic Response |
| MSI | Microsatellite Instability |
| MSS | Microsatellite Stable |
| MTD | Maximum Tolerated Dose |
| nPCR | Near pathological Complete Response |
| NOAEL | No-observed-adverse-effect-level |
| ORR | Objective Response Rate |
| OS | Overall Survival |
| pCR | Pathological Complete Response |
| PD-1 | Programmed Death Receptor 1 |
| PD-L1 | Programmed Cell Death 1 Ligand 1 |

| Abbreviations | Definition |
|---|---|
| PD-L2 | Programmed Cell Death Ligand-2 |
| PF | Cisplatin and 5-Fluorouracil |
| PFS | Progression-free Survival |
| pMMR | Proficient Mismatch Repair |
| PRs | Partial Responses |
| RFS | Relapse-free Survival |
| RP2D | Recommended Phase 2 Dose |
| SFKs | Src Family Kinases |
| TKIs | Tyrosine Kinase Inhibitors |
| TPF | Docetaxel, Cisplatin, and 5-Fluorouracil |
| TRAEs | Treatment-related Adverse Events |
| Vd | Volume of Distribution |

### 1 INTRODUCTION

#### 1.1 Current Status and Challenges of HNSCC Treatment

HNSCC primarily refers to squamous cell carcinoma occurring on the mucosal surfaces of the head and neck anatomical areas, including the oral cavity, nasal cavity, pharynx, and larynx [1]. According to the latest global cancer statistics from GLOBOCAN 2018, there were approximately 890,000 new cases of head and neck squamous cell carcinoma worldwide, with a death toll of 450,000. It is projected that new cases will increase at a rate of 30% annually, exceeding 1 million cases by 2030 [2]. In China, the situation is similarly concerning, with over 130,000 new cases and nearly 70,000 deaths each year [3]. The main risk factors for head and neck squamous cell carcinoma include smoking, alcohol consumption, betel quid chewing, bacterial and viral infections, and malnutrition. Despite the comprehensive treatment approach focusing on surgery supplemented by radiation and chemotherapy, the treatment outcomes remain unsatisfactory, with an overall five-year survival rate of only 50-60%, and just 26% for patients with advanced stages [3]. Therefore, improving the clinical treatment outcomes for oral and oropharyngeal squamous cell carcinoma, enhancing patient prognosis, and increasing quality of life remain urgent challenges we need to address.

For early-stage head and neck squamous cell carcinoma patients, surgery or radiation therapy alone is sufficient, and these patients have a good prognosis. For locally advanced patients, treatment may involve concurrent chemoradiotherapy or surgical resection combined with postoperative adjuvant therapy, depending on the possibility of surgical excision. Postoperative adjuvant therapy is tailored based on the presence of adverse prognostic factors, involving either postoperative radiation therapy or chemoradiotherapy [4]. Chemotherapy is the main non-surgical treatment for oral and oropharyngeal squamous cell carcinoma. Research has explored the value of induction chemotherapy combined with standard treatment regimens in locally advanced head and neck tumors. The first Phase III clinical trial reported on the PF regimen as an induction chemotherapy scheme, with results indicating that the use of induction chemotherapy on top of standard treatment does not reduce local recurrence rates or distant metastasis rates, nor does it improve tumor survival [5]. Due to the findings from the TAX323 and TAX324 Phase III clinical trials, which showed that the addition of docetaxel to the PF regimen can improve overall survival in patients with unresectable locally advanced head and neck tumors, it was suggested that the TPF regimen may offer benefits in the induction treatment of locally advanced head and neck tumors. Subsequently, further research explored the value of TPF induction chemotherapy combined with standard treatment regimens in resectable locally advanced head and neck tumors. The study was divided into two groups: one receiving TPF induction chemotherapy combined with standard treatment and the other receiving standard treatment alone. The results indicated that the objective response rate in the induction chemotherapy group was 80.6%; however, it did not improve DFS (HR, 0.974; 95% CI, 0.654 to 1.45; P = 0.897) or OS (HR, 0.977; 95% CI, 0.634 to 1.507; P = 0.918) [6]. The meta-analysis based on the two studies mentioned also shows that the use of induction chemotherapy prior to standard treatment does not reduce the local recurrence rate or improve survival time [5]. Paclitaxel combined with cisplatin is a first-line chemotherapy regimen for recurrent or metastatic HNSCC. Multiple studies indicate that induction chemotherapy with cisplatin and paclitaxel can provide imaging complete response rates of 8% to 33% and partial response rates of 50% to 85%. Approximately 50% of patients experience grade 3-4 chemotherapy-related adverse reactions during treatment, which can lead to surgical delays and decreased quality of life for patients[5–13]. Although induction chemotherapy can improve the ORR in patients, it does not enhance the long-term survival benefits in HNSCC [5,14]. Neoadjuvant chemoradiotherapy or targeted therapy for oral squamous cell carcinoma and oropharyngeal squamous cell carcinoma has been associated with an increased risk of toxicity, with grade 3-4 adverse events occurring in up to 50% of cases. There is an urgent need for a more effective and safer systemic treatment option for these cancers [15,16].

#### 1.2 Preliminary Exploration of Immunotherapy in HNSCC

In recent years, tumor immune checkpoint therapy has gradually become a hotspot for research and development, continuously achieving breakthroughs. Unlike cytotoxic drugs or monoclonal antibodies and small molecule tyrosine kinase inhibitors that target tumor-driving genes, tumor immune checkpoint therapy does not directly act on tumor cells. Instead, it works by blocking the inhibitory signals that suppress T cell proliferation and activation, thereby releasing the immune evasion mechanisms of tumor cells, restoring T cell activity, and enhancing the effective recognition and killing of tumor cells by T cells [17]. Currently, the tumor immune checkpoint targets that have shown significant clinical efficacy include PD-1/PD-L1 and CTLA-4. Among these, immune checkpoint inhibitors targeting PD-1/PD-L1 have better clinical application prospects due to their favorable safety profile and broader indications [18]. PD-1 (CD279) is a 55 kD type I transmembrane protein and a member of the CD28 co-stimulatory molecule family. PD-1 contains an intracellular ITIM and a membrane-distal ITSM. Two specific ligands for PD-1 have been identified: PD-L1 (B7-H1/CD274) and PD-L2 (B7-DC/CD273). Both PD-L1 and PD-L2 have been shown to downregulate T cell activation upon binding to PD-1 in both mouse and human systems. PD-1 transmits negative signals by recruiting SHP-2 to phosphorylated tyrosine residues in the ITSM cytoplasmic region. ICIs are a broadly effective class of immunotherapies that block inhibitory immune checkpoint pathways to reactivate immune responses against cancer. PD-L1, typically expressed by tumor cells, binds to the PD-1 protein expressed on T cells, leading to the inhibition of T cell immune responses and contributing to the mechanisms of tumor immune evasion [19–21]. In recent years, immune checkpoint inhibitors such as anti-PD-1 monoclonal antibodies have rapidly developed for advanced HNSCC. In the CheckMate 141 study, compared to standard treatment, Nivolumab (a PD-1 inhibitor) significantly extended OS, increasing the one-year OS rate from 16.6% to 36% (HR, 0.70) [22]. In the KEYNOTE-040 study, Pembrolizumab (a PD-1 inhibitor) also demonstrated improved median OS compared to standard treatment, increasing from 6.9 months to 8.4 months (HR, 0.80). Additionally, more significant benefits were observed in the population with PD-L1 positivity and CPS≥1 [23]. In 2016, PD-1 inhibitors Nivolumab and Pembrolizumab were approved by the FDA for patients with recurrent/metastatic HNSCC, showing overall survival benefits compared to chemotherapy [22,24]. The 2020 version of the CSCO Guidelines for Head and Neck Cancer reinforced the role of immunotherapy in the second-line treatment of recurrent or metastatic head and neck squamous cell carcinoma (excluding nasopharyngeal carcinoma), elevating the recommendation level for Nivolumab from grade II to grade I, establishing it as the standard second-line treatment for recurrent or metastatic head and neck squamous cell carcinoma after first-line failure in China.

#### 1.3 Neoadjuvant Immunotherapy

In recent years, neoadjuvant immunotherapy with checkpoint inhibitors has shown significant clinical efficacy in melanoma, non-small cell lung cancer, bladder cancer, and glioblastoma [25–30]. A multicenter phase II clinical study aimed to evaluate the efficacy and safety of cemiplimab as neoadjuvant therapy for patients with resectable stage II-IV cutaneous squamous cell carcinoma. A total of 79 patients were enrolled, among which 40 cases (51%) achieved complete pathological response, and 10 patients (13%) experienced significant pathological response. Imaging objective response was observed in 54 patients (68%). Adverse events of grade 3 or higher occurred in 14 patients (18%) during the study period [31]. ICIs, such as PD-1 inhibitors, have rapidly developed in HNSCC. In a phase II clinical trial conducted in 2020 involving patients with stage II-IVA oral squamous cell carcinoma, neoadjuvant use of nivolumab (a PD-1 inhibitor) resulted in a 50% volumetric response, 53% pathological downgrade, and 50% pathological response among patients. With a median follow-up of 14.2 months, the one-year progression-free survival rate was 85%, and the overall survival rate was 89%, demonstrating the feasibility and safety of preoperative nivolumab in patients with resectable oral squamous cell carcinoma [32]. In 2020, data from the IMCISION study was presented at the ESMO annual meeting. This study, an Ib/II phase trial, evaluated the use of nivolumab alone (Ib phase, 6 patients) and in combination with ipilimumab (II phase, 26 patients) for neoadjuvant therapy in resectable HNSCC. Among the 29 evaluable patients, 31% (9 patients) achieved nPCR with nivolumab ± ipilimumab. At a follow-up of 14 months, the RFS rate for patients with pCR reached 100%. The safety profile was manageable, with an overall incidence of grade 3-4 irAEs at 38%. The safety, feasibility, and effectiveness of nivolumab monotherapy and its combination with ipilimumab for neoadjuvant treatment of head and neck squamous cell carcinoma have been preliminarily validated. Currently ongoing trials, IMSTAR-HN and KEYNOTE-689, are exploring the value of nivolumab ± ipilimumab and pembrolizumab in perioperative settings combined with existing postoperative adjuvant treatment modalities for tumor control, with promising results anticipated. Data from a phase II clinical study presented at the 2021 ASCO annual meeting showed good tolerability for two cycles of the PD-1 inhibitor pembrolizumab as neoadjuvant therapy for locally advanced head and neck squamous cell carcinoma, with only one patient experiencing a grade 3 adverse event and no grade 4 events. The pathological response rate (pTR-2) was 44%, confirming that two cycles of monotherapy with a PD-1 inhibitor have good safety and certain efficacy, although new treatment strategies are needed to further enhance effectiveness [33].

#### 1.4 Immunotherapy Combined with Other Treatments.

The abnormal activation of tyrosine kinases caused by mutations, translocations, or amplifications is associated with the development, progression, invasion, and metastasis of malignant tumors. Additionally, wild-type tyrosine kinases can serve as critical nodes in the activation of cancer pathways. Therefore, tyrosine kinases have become a major target in drug discovery [34,35]. TKIs inhibit the corresponding kinases, preventing the phosphorylation of tyrosine residues on their substrates and thereby blocking the activation of downstream signaling pathways [36]. In the past 20 years, various robust and well-tolerated TKIs have been developed, targeting single or multiple pathways, contributing to the realization of precision oncology based on individual patient genetic alterations. TKIs have significantly improved patient survival rates and quality of life, transforming the treatment paradigms for various solid tumors. Additionally, TKIs can induce the reconstruction of the immune microenvironment and activate tumor immune response processes [35]. Current research indicates that immunotherapy combined with tyrosine kinase inhibitors shows promising efficacy and acceptable toxicity in cancers such as endometrial carcinoma and clear cell renal cell carcinoma. The KEYNOTE-146 study is a Phase 1b/2, multicenter, open-label, single-arm trial designed to evaluate the efficacy and safety of pembrolizumab combined with lenvatinib in patients with advanced endometrial carcinoma who have previously received no more than two lines of chemotherapy and have measurable lesions, reporting a 24-week ORR of 38.0%. For patients who have previously received this treatment, regardless of MSI status, the median DOR was 21.2 months, the median PFS was 7.4 months, and the median OS was 16.7 months. The results of this study indicate that the combination therapy for recurrent endometrial carcinoma is not only significantly effective in MSI-H patients but also shows better efficacy in MSS patients compared to pembrolizumab monotherapy [37]. KEYNOTE-775 is a multicenter, open-label, randomized controlled phase III clinical study comparing the efficacy and safety of pembrolizumab combined with lenvatinib to physician's choice of treatment (doxorubicin or paclitaxel) in patients with advanced/recurrent endometrial cancer who had previously received at least one line of platinum-based therapy. Compared to the control group, pembrolizumab combined with lenvatinib resulted in a median PFS increase of 2.8 months and a 40% reduction in the risk of recurrence or death for patients with pMMR. The median OS increased by 5.4 months, with a 32% reduction in mortality risk, and the ORR improved by 15.2%. For the overall patient population, the median PFS was extended by 3.4 months, with a 44% reduction in recurrence or death risk, a median OS increase of 6.9 months, a 38% reduction in mortality risk, and an ORR improvement of 17.2%. The study indicates that in patients with advanced/recurrent endometrial cancer who have previously received at least one line of platinum-based therapy, the combination of Pembrolizumab and Lenvatinib can improve the prognosis for all patients, including those with pMMR. The TATTON study evaluates the combination of Osimertinib (an EGFR TKI) and Durvalumab for advanced EGFR-mutant non-small cell lung cancer, achieving an ORR of 50%-80%, with 15% (5/34) experiencing grade 3-4 interstitial lung disease [38]. The KEYNOTE-426 trial is an open-label phase 3 study comparing the efficacy and safety of Pembrolizumab combined with Atezolizumab versus Sunitinib in previously untreated patients with advanced renal cell carcinoma. After a median follow-up of 12.8 months, the 12-month OS rates were 89.9% for the Pembrolizumab-Atezolizumab group and 78.3% for the Sunitinib group. The median PFS was 15.1 months for the Pembrolizumab-Atezolizumab group and 11.1 months for the Sunitinib group. The ORR was 59.3% for the Pembrolizumab-Atezolizumab group and 35.7% for the Sunitinib group. Additionally, 75.8% of patients in the Pembrolizumab-Atezolizumab group and 70.6% in the Sunitinib group experienced grade 3 or higher adverse events of any cause [34]. In a phase Ib study of neoadjuvant therapy for locally advanced oral squamous cell carcinoma, 20 patients with resectable locally advanced oral squamous cell carcinoma received three cycles of Camrelizumab (an anti-PD-1 monoclonal antibody) combined with Apatinib before surgery. The results showed that the neoadjuvant therapy was well tolerated, with a MPR rate of 40%, achieving the primary endpoint and confirming that the combination of Camrelizumab and Apatinib is safe and effective for resectable oral squamous cell carcinoma [36].

Dasatinib, as a member of tyrosine kinase inhibitors, is a potent oral ATP-competitive inhibitor that can inhibit various tyrosine kinases, including BCR-ABL, c-KIT, platelet-derived growth factor receptor, and SFKs. Quercetin, a flavonoid compound, is widely found in various plants, fruits, and vegetables in glycoside form and has been added to functional foods as a dietary supplement. Quercetin not only exhibits anti-inflammatory, anti-tumor, anti-platelet aggregation, endothelial cell protection, and antioxidant effects, but also plays a positive immunomodulatory role in the immune system, promoting immune cell proliferation and enhancing the body's immune function [39]. The combination of Dasatinib and Quercetin is a widely used therapeutic regimen. In a multicenter, open-label, phase I clinical study evaluating the efficacy and feasibility of this combination in patients with IPF (n = 14), the primary endpoints were retention and completion rates assessed clinically. The results indicated a retention rate of 100%, with no patients discontinuing treatment; only one patient experienced a serious adverse event [40]. Currently, the D+Q treatment model has been widely studied in both preclinical and clinical research [41–49]. In gliomas, preclinical studies have shown that the combination of immunotherapy with Dasatinib and Quercetin is well-tolerated and significantly improves prognosis [50].

#### 1.5 Background Information on Tislelizumab

##### 1.5.1 Pharmacology

Tislelizumab (also known as BGB-A317) is a humanized IgG4 variant monoclonal antibody targeting PD-1. It was approved for marketing in China on December 27, 2019, with the indication for classical Hodgkin lymphoma, and is currently in clinical development for various other human malignancies.

Tislelizumab binds to the extracellular domain of human PD-1 with high specificity and high affinity (dissociation constant [KD] = 0.15 nM), exerting its pharmacological effects. It competitively blocks the binding of PD-L1 and PD-L2, thereby inhibiting PD-1 mediated negative signaling in T cells. In in vitro cell assays, Tislelizumab has been shown to enhance the functional activity of human T cells and pre-activated primary peripheral mononuclear cells in a sustained and dose-dependent manner. Additionally, Tislelizumab has demonstrated anti-tumor activity in several xenograft models, where peripheral blood mononuclear cells were co-injected with human cancer cells (A431 [epithelial carcinoma]) or tumor fragments (BCCO-028 [colon cancer]) into immunocompromised mice.

In addition, Tislelizumab is an IgG4 variant antibody to FcγR such as FcγRI and FcγRIIIA. It has very low binding affinity to C1q, a subunit of complement 1. In vitro assays with Tislelizumab suggest either very low or no ADCC, ADCP, or CDC effects in humans [51,52].

##### 1.5.2 Toxicology

The toxicity and safety profile of Tislelizumab was characterized in single-dose toxicology studies in mice and monkeys and in a 13-week, repeat-dose toxicology study in cynomolgus monkeys. The tissue cross-reactivity was evaluated in the normal frozen tissues from both humans and monkeys. The cytokine release assays were also evaluated using fresh human whole blood cells. The pivotal toxicology studies were conducted following GLP regulations. The single-dose regimens spanned from the intended human dose to 10-fold higher than the maximum of the intended human dose, and the repeated-dose regimens spanned from the intended human dose to 3-fold higher than the maximum of the intended human dose. Cynomolgus monkey was the only relevant species based on target sequence homology and binding activity. Overall, no apparent toxicity was noted in mice and monkey toxicity studies. No tissue cross-reactivity was found in either human or monkey tissues, nor was any effect on cytokine release observed in human whole-blood assay. The toxicokinetic profile was well characterized, with dose proportional increases in systemic exposure without apparent accumulation or sex difference. Immunogenicity was observed without apparent immunotoxicity or effect on the systemic exposure. The NOAEL of Tislelizumab in the 13-week monkey toxicity study was considered to be 30 mg/kg. The safety profile of Tislelizumab is considered adequate to support the current study COIS-01.

Please refer to the Tislelizumab Prescribing Information for more detailed information on the toxicology of Tislelizumab.

##### 1.5.3 Clinical Pharmacology

In Phase 1 studies BGB-A317_Study_001 and BGB-A317-102, interim PK analysis (cut-off date 28 August 2017) was conducted by noncompartmental methods, using serum concentrations from patients who received doses of 0.5, 2.0, 5.0, 10 mg/kg Q2W and 2.0mg/kg, 5.0 mg/kg, 200 mg Q3W (Phase 1a Parts 1, 2, and 3, and Phase 1b in BGBA317_Study_001) and patients who received doses of 200 mg Q3W in

Phase 1 of Study BGB-A317-102 (n = 19). The maximum observed plasma concentration (Cmax) and the area under the concentration-time curve (AUC) increased in a nearly dose-proportional manner from 0.5 mg/kg to 10 mg/kg, both after single-dose administration and at steady state. Preliminary PK data from 27 patients who were administered 1 dose of 200 mg Q3W(Phase 1a, Part 3 and Study BGB-A317-102) showed Tislelizumab concentrations between the range of concentrations observed for patients who were administered 2 mg/kg and 5 mg/kg doses. Preliminary population PK analysis using a 2-compartment model with first-order elimination showed a systemic plasma clearance (CL) of Tislelizumab of 0.173 L/d, Vd in the central and peripheral compartments of 2.89 L and 1.76 L, respectively, and half-life (t1/2) of approximately 19 days. Race, gender, and body weight were not significant covariates on the CL of Tislelizumab, which supports fixed-dosing across different ethnic groups.

##### 1.5.4 Prior Clinical Experience With Tislelizumab

As of 20 May 2019, there are 22 ongoing studies with Tislelizumab worldwide, including monotherapy and combination studies in solid tumors and hematological malignancies and 1705 patients have received Tislelizumab administration. Of the ongoing monotherapy studies in solid tumors, available data of urothelial carcinoma from BGB-A317_Study_001 and BGB-A317-102 are summarized in Section 1.3.4.1and Section 1.3.4.2(with a data cutoff date of 1 December 2018). Data from the Phase 2 registrational clinical study BGBA317-204 with Tislelizumab in inoperable locally advanced or metastatic urothelial carcinoma are summarized in Section 1.3.4.3 (with a data cut-off date of 28 February 2019).

Please refer to the Tislelizumab Prescribing Information for more detailed information on efficacy and safety of Tislelizumab.

###### 1.5.4.1 BGB-A317_Study_001 (Data Cut-off Date: 28 August 2017)

Study BGB-A317_Study_001 is a 2-stage study consisting of a Phase 1a dose-escalation and dose-expansion component with 3 parts to establish the MTD, if any, a RP2D for the Phase 1b, and a flat dose (fixed dose) followed by a Phase 1b component to investigate efficacy in selected tumor types in indication expansion arms and to further evaluate safety and tolerability of Tislelizumab.

As of 28 August 2017, in Phase 1a, 116 patients have received Tislelizumab at dose regimens including 0.5 mg/kg, 2 mg/kg, 5 mg/kg, or 10 mg/kg Q2W; 2 mg/kg or 5 mg/kg Q3W; and 200 mg Q3W. In Phase 1b, 323 patients across 9 indication-expansion cohorts had received Tislelizumab.

Overall, for the 439 patients in the study, the median age was 60.0 years, 53.8% of the population was male, and 65.6% of patients were white. The median number of prior anticancer therapy regimens was 2 (range: 0 to 12). The median treatment exposure duration was 2.50 months (range: 0 to 23.0 months) and the median study follow-up duration was 5.56 months (range: 0.0 to 26.9 months). As of 28 August 2017, there were 210 patients (47.8%) remaining on study in BGB-A317_Study_001.

**Preliminary Safety**

Of the 439 total patients in the Safety Population for study BGB-A317_Study_001, 240 patients (54.7%) experienced at least 1 treatment-emergent adverse event (TEAE) assessed as related to Tislelizumab by investigator and 34 patients (7.7%) experienced at least 1 ≥ Grade 3 Tislelizumab-related TEAE. The most commonly reported treatment related TEAEs for patients treated with Tislelizumab monotherapy in study BGBA317_Study_001 were fatigue (12.8%), rash (7.7%), nausea (6.8%), diarrhea (6.6%), and hypothyroidism (4.8%). The ≥ Grade 3 TEAEs occurring in ≥ 2 patients were pneumonitis (6 patients, 1.4%); colitis and alanine aminotransferase (ALT) increased (4 patients each, 0.9%); fatigue, type 1 diabetes mellitus, and aspartate aminotransferase (AST) increased (3 patients each, 0.7%); and diarrhoea, gamma-glutamyltransferase (GGT) increased, and diabetic ketoacidosis (2 patients each, 0.5%). All other AEs occurred in single patients. Lastly, 18 patients (4.1%) experienced infusion-related reactions; all were mild/moderate in severity.

**Preliminary Efficacy**

- For patients in Phase 1a (n = 116, evaluable), there were 20 patients with a confirmed response and 42 patients with a best overall response (BOR) of stable disease.

- For patients in Phase 1b (n = 286 evaluable), a total of 26 patients had a confirmed response. Additionally, there were 101 patients with a BOR of stable disease.

###### 1.5.4.2 Study BGB-A317-204 (Data Cut-off Date: 28 February 2019)

Study BGB-A317-204 (CTR20170071) is a Phase 2 clinical study to evaluate the efficacy, safety, and tolerability of Tislelizumab (200 mg Q3W) in patients with PD-L1+ urothelial bladder cancer who have received ≥ 1 lines of prior platinum-containing chemotherapy in China and South Korea. The study is designed as a single-arm study which enrolls secondline patients with PD-L1+ advanced urothelial bladder cancer. Patients will receive Tislelizumab monotherapy until documented disease progression or intolerance to Tislelizumab. Primary endpoint is ORR assessed by IRC per RECIST v1.1, and secondary endpoints are DOR, PFS, DCR, and OS. The study is planned to enroll 113 patients, including 108 patients from China.

**Preliminary Efficacy**

As of the DCO date of 28 February 2019, the Efficacy Evaluable Analysis Set included 104 patients with measurable disease per IRC according to RECIST v1.1 at baseline, which was the primary analysis set for the efficacy analyses. The median duration of Tislelizumab treatment of all these patients was 15.3 weeks with a median follow-up time of 7.6 months, and 30 patients remained on Tislelizumab treatment. Of the 104 patients evaluated, 24 patients achieved ORs (ORR = 23.1%, 95% CI: 15.4 to 32.4) as confirmed by the IRC, including 8 CRs and 16 PRs as assessed by the IRC; 19 of 24 patients (79%) had ongoing response; subgroup analysis suggested that the response rate was independent of baseline factors; additionally, the response rate reported was similar to the pooled response rate in Phase 1 study (PD-L1+: + 24%, PD-L1-/unknown: 21%). The median PFS was 2.1 months and OS was 9.8 months.

**Preliminary Safety**

- Safety analysis showed an overall TRAEs incidence of 92.9%, most of which were ⩽ Grade 2 in severity.
- TRAEs that occurred in > 15% of patients included anemia (26.5%), decreased appetite (18.6%) , and pyrexia (16.8%); anemia (7%) was the only Grade 3 to 4 TRAEs.
- Immune-related adverse events were reported in 64% of patients, and the most common irAEs included: immune-related skin adverse reactions (n = 38; 34%), immune-related hepatitis (n = 27; 24%), immune-related thyroid disorder (n = 15; 13%), and immune-related nephritis/renal dysfunction (n = 13; 12%).

- There were no ≥ Grade 3 immune related adverse events with incidence > 5%.

##### 1.5.4.3 Pooled Analyses for Immune-Related Adverse Events

Immune-related adverse events (irAEs) were reported in 23.1% of patients who received Tislelizumab monotherapy in Study BGB-A317_Study_001 and 16% of patients in Study BGB-A317-102; irAEs that were reported in at least 2% of patients in the overall population are detailed in Table 1 below.

**Table 1: Immune-Related TEAEs of Tislelizumab as adjudicated by Investigator**

| | BGB-A317_Study_001 Study [a]<br>N = 451<br>n (%) | BGB-A317-102 Study [b]<br>N = 300<br>n (%) |
|---|---|---|
| Immune-related abnormal thyroid function | 39 (8.6) | 25 (8.3) |
| Skin reactions | 39 (8.6) | 9 (3.0) |
| Immune-related pneumonitis | 12 (2.7) | 7 (2.3) |
| Hepatitis | 10 (2.2) | 5 (1.7) |
| Colitis | 9 (2.0) | 0 |

[a] As of 27 October 2018. [b] As of December 1, 2018.

Of these irAEs, immune-related skin adverse reactions were reported more frequently in Study BGB-A317_Study_001 in the overall population as compared with Study BGBA317-102. Most of these events were Grade 1 or 2 in severity with Grade 3 events occurring less frequently.

In study BGB-A317_Study_001, the potential irTEAEs assessed as ≥ Grade 3 were: Grade 3 pneumonitis and ALT increased in 6 patients (1.3%) each, diarrhea and colitis in 3 patients (0.7%) each, GGT increased and hyperglycemia in 2 patients (0.4%) each. All other potential irTEAEs assessed as ≥ Grade 3 were events in single patients, including a Grade 4 event of diabetic ketoacidosis in a patient with adenoid cystic carcinoma, and 2 patients with Grade 5 events of acute hepatitis in a patient with HCC and pneumonitis in a patient with NSCLC.

In Study BGB-A317-102, the only Grade 4 event reported was a single case of an adverse skin reaction. There were no deaths due to immune-related adverse events in

Study BGBA317-102.

In summary, the immunotherapy monotherapy neoadjuvant treatment model has shown good efficacy in clinical trials, but some patients still do not benefit. Research indicates that in solid tumors such as endometrial cancer, clear cell renal cell carcinoma, advanced EGFR mutation NSCLC, and prostate cancer, the combination of immunotherapy with tyrosine kinase inhibitors demonstrates good efficacy and acceptable toxicity. Additionally, preclinical studies in gliomas have shown that the combination of immunotherapy with Dasatinib and Quercetin is well-tolerated and can significantly improve prognosis. Therefore, to avoid the adverse effects of chemotherapy and achieve a chemo-free comprehensive treatment model while enhancing the efficacy of immunotherapy to ultimately improve the survival outcomes of patients with head and neck squamous cell carcinoma, we propose to conduct a prospective, open-label, single-center, phase II clinical study of Tislelizumab combined with Dasatinib and Quercetin as neoadjuvant therapy for resectable head and neck squamous cell carcinoma patients. The aim is to evaluate its safety and efficacy. This study will also expand the indications for neoadjuvant immunotherapy in solid tumors and provide new strategies to improve the prognosis of patients with HNSCC.

### 2 STUDY OBJECTIVES AND ENDPOINTS

#### 2.1 STUDY OBJECTIVES

##### 2.1.1 Primary Objective

Aimed at evaluating the efficacy of Tislelizumab combined with Dasatinib and Quercetin as neoadjuvant therapy in patients with resectable HNSCC, providing new avenues to further improve the prognosis of these patients.

##### 2.1.2 Secondary Objectives

• Safety: Adverse events related to treatment during the study and follow-up period will be assessed by the investigator according to NCI CTCAE (version 5.0).

• Overall Survival (OS) assessed by the investigator at 5 years.

• Disease-Free Survival (DFS) assessed by the investigator at 2 years.

• Objective Response Rate (ORR) assessed by the investigator according to RECIST version 1.1.

• Surgery Delay Rate: Participants who cannot undergo surgery within 8 weeks after the last dose will be classified as having surgery delay.

##### 2.1.3 Exploratory Objective

To evaluate biomarkers in tumor tissue and blood associated with the efficacy, resistance, and prognosis of this neoadjuvant treatment regimen, including but not limited to the expression of PD-1, tumor mutational burden, and immune-related gene expression profiles.

#### 2.2 Study Endpoints

##### 2.2.1 Primary Endpoint

Major Pathological Response (MPR) rate : the proportion of participants with viable tumor cells ≤ 10% in the resected specimen among the total participants.

##### 2.2.2 Secondary Endpoints

• Safety: The severity of treatment-related adverse events will be graded according to NCI CTCAE (version 5.0) during the study and follow-up.

• Overall Survival (OS): Defined as the time from enrollment to the date of death from any cause (5 years).

• Disease-free survival (DFS)： DFS is defined as the time from treatment until the date of the first relapse (local/regional recurrence or distant metastasis) or death (from any cause) whichever comes firsts and regardless of whether the patient withdraws from treatment or receives another anti-cancer therapy prior to disease relapse.

• Objective Response Rate (ORR): Assessed through imaging according to the Response Evaluation Criteria in Solid Tumors (RECIST 1.1).

• Surgery Delay Rate: Participants who cannot undergo surgery within 8 weeks after the last dose will be classified as having surgery delay.

##### 2.2.3 Exploratory Endpoint

To evaluate biomarkers in tumor tissue and blood associated with the efficacy, resistance, and prognosis of this neoadjuvant treatment regimen, including but not limited to the expression of PD-1, tumor mutational burden, and immune-related gene expression profiles.

### 3 STUDY DESIGN

This study is a prospective, open-label, single-center, phase II clinical trial evaluating the safety and efficacy of Tislelizumab in combination with Dasatinib and Quercetin as neoadjuvant therapy for resectable head and neck squamous cell carcinoma. The study aims to expand the indications for neoadjuvant immunotherapy in solid tumors and provide new strategies to improve the prognosis of patients with head and neck squamous cell carcinoma.

The treatment regimen is as follows: Tislelizumab 200 mg, administered intravenously on day 1; Dasatinib, orally at 100 mg/day on days 1, 2, and 3; Quercetin (phytosome Thorne Research Sophora japonica concentrate [leaf]/phosphatidylcholine complex from Sunflower), orally at 1250 mg/day on days 1, 2, and 3. The treatment will be given every 3 weeks for a total of 3 cycles. Surgery is scheduled to take place between days 22 and 56 after the completion of neoadjuvant therapy. Postoperative treatment will include chemoradiotherapy as per guidelines.

Neoadjuvant treatment will be administered according to the Q3W schedule until one of the following occurs (whichever comes first): 1) completion of 3 cycles of treatment; 2) occurrence of unacceptable toxicity; 3) documentation of progressive disease (PD) according to the RECIST v1.1 criteria; 4) clinical assessment by the clinician indicates that the patient can no longer benefit from neoadjuvant therapy; 5) withdrawal of consent, loss to follow-up, or death. Subsequent treatment will be determined by the investigator based on the patient's treatment status.

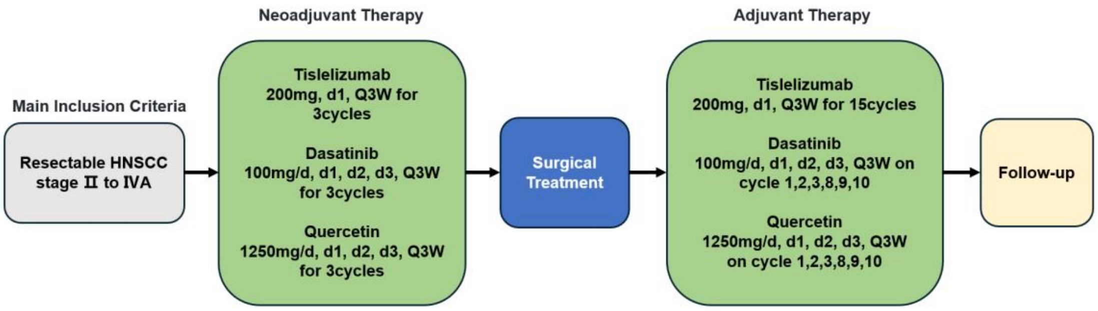

**Figure 1: Study Design**

**Table 2: Schedule of Assessments**

| | Screening | Treatment Period | | | Maintenance Treatment Period[p] | Medication Safety Follow-up[q] | Survival Follow-up[r] |
|---|---|---|---|---|---|---|---|
| | | Cycle 1 | Cycle 2 | Cycle 3 | | | |
| Days (window) | -28~-1 | 1 (+3) | 22 (±3) | 43 (±3) | every three weeks (±3) | 30 days after the last dose (±7) | every 90 days (±7) |
| General Research Process | | | | | | | |
| Informed Consent[a] | X | | | | | | |
| Inclusion/Exclusion Criteria | X | | | | | | |
| Demographic Data/History of Treatment[b] | X | | | | | | |
| Previous and Concomitant Medications | X | X | X | X | X | X | |
| Physical Examination/Vital signs[c] | X | X | X | X | X | X | |
| ECOG Assessments | X | X | X | X | X | X | |
| Electrocardiogram[d] | X | | X | X | X | X | |
| Laboratory Evaluation | | | | | | | |
| Hematology[e] | X | | X | X | X | X | |
| Serum Chemistry[e] | X | | X | X | X | X | |
| Urinalysis[e] | X | | X | X | X | X | |
| Coagulation[f] | X | | X | X | X | X | |
| Pregnancy Test[g] | X | | | | | | |
| Thyroid Function[h] | X | | X | X | X | X | |
| Cardiac Enzyme Profile[h] | X | | X | X | X | | |

| | Screening | Treatment Period | | | Maintenance Treatment Period[p] | Medication Safety Follow-up[q] | Survival Follow-up[r] |
|---|---|---|---|---|---|---|---|
| | | Cycle 1 | Cycle 2 | Cycle 3 | | | |
| Days (window) | -28~-1 | 1 (+3) | 22 (±3) | 43 (±3) | every three weeks (±3) | 30 days after the last dose (±7) | every 90 days (±7) |
| Viral Antibody Testing (HIV, HBV and HCV)[i] | X | | | | | | |
| Test for HCV-RNA (If HCV antibody is positive)[j] | X | | X | X | X | X | |
| Test for HBV-DNA (If HBsAg and/or HBcAb is positive)[k] | X | | X | X | X | X | |
| Safety Monitoring and Survival. | | | | | | | |
| Adverse Events[l] | X | X | X | X | X | X | X |
| Subsequent Antitumor Treatment | | | | | | | X |
| Survival Status | | X | X | X | X | X | X |
| Efficacy Assessment | | | | | | | |
| Tumor Imaging Assessment[m] | X | | | X | X | X | |
| Biomarker Exploration | | | | | | | |
| Tumor Tissue[n] | X | | | | | | |
| Whole Blood[o] | | | | | | | |

a. Written informed consent is required before performing any study-specific tests or procedures. Results of standard-of-care tests or examinations performed before obtaining informed consent and ≤ 28 days before enrollment may be used for the purposes of screening rather than repeating the standard-of-care tests unless otherwise indicated.

b. Including year of birth (or age), gender and ethnicity/race; history of treatments for the initial diagnosis, including prior drug therapy(ies), locoregional treatment(s), and surgical treatment(s). Relevant information on radiological studies performed prior to study entry may be collected for review by the investigator.

c. Vital signs collected on study include temperature, pulse rate, and blood pressure (systolic and diastolic) while the patient has been resting for 10 minutes. The patient's vital signs are required to be recorded within 1 hour before, during, and 1 hour after the first 2 Tislelizumab infusions. For subsequent infusions, vital signs will be collected within 60 minutes before infusion and if clinically indicated, during and 30 minutes after the infusion. Investigators should solicit patients regarding changes in vision, visual disturbance, or ocular inflammation at each scheduled study visit during Tislelizumab treatment. For any change in vision, referral to an appropriate specialist will be made for further management guidance.

d. Patients should rest for at least 10 minutes prior to each ECG collection.

e. The complete blood count (CBC) includes: red blood cell count (RBC), hemoglobin (HGB), hematocrit (HCT), white blood cell count (WBC), platelet count (PLT), and white blood cell differential [lymphocytes, neutrophils, monocytes, eosinophils, basophils]. Serum biochemistry includes: liver function [total bilirubin (TBIL), direct bilirubin (DBIL), alanine aminotransferase (ALT), aspartate aminotransferase (AST), gamma-glutamyl transferase (γ-GT), alkaline phosphatase (ALP), albumin (ALB), total protein (TP), lactate dehydrogenase (LDH), creatine kinase (CK)], renal function [urea (UREA), creatinine (Cr)], electrolytes (Na, K, Cl, Mg, Ca, P), amylase, and blood glucose (GLU). Routine urinalysis includes: pH, urine white blood cells (UWBC), urine protein (UPRO), urine red blood cells (URBC), urine glucose (UGLU), and specific gravity. Participants with urine protein ≥ 2+ during the screening phase must undergo 24-hour urine protein quantification. Assessments will be conducted within 7 days prior to the first administration of the investigational drug during the screening phase and at the start of Cycle 2 before each administration of the investigational drug and during safety follow-up. Tests will be performed at each study center.

f. Coagulation function tests include: prothrombin time (PT) and international normalized ratio (INR). These tests will be conducted within 7 days prior to the first dose of the investigational treatment during the screening phase, before each administration of the investigational drug at the start of Cycle 2, and during safety follow-up. Assessments will be performed at each study center.

g. Women of childbearing potential will undergo urine or serum pregnancy testing within 3 days prior to the first dose during the screening phase. If the urine pregnancy test results are inconclusive for negative, a serum pregnancy test will be conducted, with serum results taking precedence. Assessments will be performed at each study center.

h. Thyroid function tests and cardiac enzyme profiles will be conducted within 28 days prior to the first dose, before each administration of the investigational drug at the start of Cycle 2, and during safety follow-up. Assessments will be performed at each study center.

i. Testing for HIV and HCV antibodies, as well as the hepatitis B panel (HBsAg, HBsAb, HBcAb, HBeAg, HBeAb), must be completed within 28 days prior to the first dose. Assessments will be conducted at each study center during the screening phase.

j. HCV antibody testing will be conducted during the screening visit. For participants who test positive for HCV antibodies, HCV-RNA levels will be measured every 3 weeks (21±7 days) starting from the first dose, or sooner if clinically indicated.

k. The hepatitis B panel will be tested during the screening visit. For participants who test positive for HBsAg and/or HBcAb, HBV-DNA levels will be measured every 3 weeks (21±7 days) starting from the first dose, or sooner if clinically indicated.

l. The assessment of adverse events (AEs) and laboratory safety evaluations will be conducted according to CTCAE v5.0. Definitions, documentation, causality assessment, severity classification, reporting timelines, and management of AEs and serious adverse events (SAEs) will follow the descriptions in Section 5 of the protocol.

m. Tumor assessment includes RECIST 1.1 evaluation. Imaging assessments for tumors typically involve contrast-enhanced CT or MRI, with the examination areas including the chest and abdomen. At baseline, pelvic CT or MRI is also required, along with cranial MRI and whole-body bone scans. If cervical lymph node enlargement is present, contrast-enhanced CT of the neck should be performed. PET/CT may be used as a screening method for baseline assessment, but if abnormalities are found, corresponding CT/MRI should be conducted to facilitate follow-up evaluations. For subjects without metastatic lesions at baseline, routine imaging of the head, pelvis, and whole-body bones is not necessary unless clinically indicated thereafter. The imaging techniques used for a subject during the study should remain consistent. Baseline assessments will be conducted within 28 days prior to enrollment. From the first administration of the study drug, tumor imaging evaluations will be performed every 6 weeks (±7 days) for the first 48 weeks, and then every 12 weeks (±7 days) thereafter. If the investigator determines radiographic progression based on RECIST 1.1 during the first assessment, but the subject's disease is clinically stable with no evidence of rapid radiographic progression, and the investigator believes the subject may continue to benefit from the study drug, the subject may continue the current treatment. Imaging should be re-evaluated at least 4 weeks later to confirm progression (based on RECIST 1.1). If progression (PD) is confirmed upon re-evaluation, the subject should discontinue study treatment. If progression is not confirmed, the subject may continue treatment, with imaging evaluations performed at the protocol-specified time points until radiographic progression is confirmed. If a subject discontinues study treatment for reasons other than objective disease progression, imaging evaluations should be performed at the time of treatment discontinuation. Imaging assessments should continue at the protocol-specified time points until any of the following events occur: initiation of a new anti-tumor therapy, objective disease progression, loss to follow-up, death, or withdrawal of informed consent (ICF) by the subject, whichever occurs first. If a subject exhibits clinical disease instability during the study, an unscheduled imaging assessment may be conducted at any time. If clinical disease instability is observed after the first assessment of radiographic progression, a 4-6 week confirmation is not required, and the subject should discontinue study treatment immediately.

n. Subjects are required to provide at least 5-15 archived or fresh tumor tissue samples that meet testing criteria during the screening period. If clinically feasible, a biopsy is requested at the time of radiographic disease progression.

o. Subjects are required to provide 10 ml of whole blood samples for tumor biomarker testing at the following time points: prior to the first administration of the study drug, before each imaging assessment during the treatment period (before starting the next treatment), and upon confirmation of disease progression. Testing will be conducted at the central laboratory.

p. The patient underwent adjuvant therapy post-surgery with the following regimen: Tislelizumab 200 mg, IV, on day 1 every 3 weeks, for 15 cycles. Dasatinib, PO, 100 mg/day, on days 1, 2, 3 every 3 weeks, on cycles 1, 2, 3, 8, 9, 10. Quercetin, PO, 1250 mg/day, on days 1, 2, 3 every 3 weeks, on cycle 1, 2, 3, 8, 9, 10.

q. The safety follow-up will be conducted 30±7 days after the last administration of the drug or before the start of new antitumor treatment, whichever occurs first. All adverse events (AEs) that occurred prior to the safety follow-up should be recorded until they resolve to grade 0-1 or baseline levels, or until the investigator deems that further follow-up is unnecessary for justifiable reasons (such as irreversible conditions or improvement), whichever occurs first. Serious adverse events (SAEs) occurring within 90 days after the last administration of the drug or before the subject starts new anticancer treatment (whichever occurs first) will be followed up and recorded.

r. Survival follow-up: Safety assessments will be conducted every 90 days (± 7 days) after the safety follow-up and may include phone interviews.

### 4 STUDY APPROACH

#### 4.1 Sample Size Calculation and Statistical Analysis Methods

This study is a prospective, open-label, single-center, Phase II clinical trial investigating the neoadjuvant treatment of resectable head and neck squamous cell carcinoma patients using Tislelizumab in combination with Dasatinib and Quercetin, with the primary target population being patients with operable head and neck squamous cell carcinoma.

This study is a single-arm trial, with the primary endpoint being the MPR rate. Previous research has indicated that the MPR rate for neoadjuvant treatment with PD-1 inhibitors (such as Pembrolizumab) in operable head and neck squamous cell carcinoma is 7%[53]. Assuming that this study aims to increase the MPR rate to 30% after neoadjuvant treatment, a one-sided alpha of 0.025, a power of 0.9, and an annual dropout rate of 5% lead to a calculated sample size of 24 cases.

#### 4.2 Inclusion Criteria

• Before implementing any trial-related procedures, written informed consent must be obtained.

• Age must be between 18 years and under 80 years.

• Cytological or histological diagnosis of head and neck squamous cell carcinoma. The investigator assesses the head and neck squamous cell carcinoma as resectable, with no distant metastasis. According to the 8th edition of the AJCC staging, it is classified as stage II-IVA.

• According to the Response Evaluation Criteria in Solid Tumors (RECIST 1.1), there must be at least one measurable lesion on imaging; for first-line patients: they have not received any systemic antitumor therapy for advanced/metastatic disease. For patients who have previously undergone platinum-based adjuvant/neoadjuvant chemotherapy or definitive chemoradiotherapy for advanced disease, if disease progression or recurrence occurs at least 6 months after the last chemotherapy treatment, enrollment in this study is permitted.

• ECOG score of 0-1

• Expected survival time > 3 months

• Sufficient organ function: subjects must meet the following laboratory criteria: Absolute neutrophil count (ANC) ⩾ 1.5 x 10^9/L without the use of granulocyte colony-stimulating factor in the past 14 days; Platelet count ⩾ 100 x 10^9/L without blood transfusion in the past 14 days; Hemoglobin > 9 g/dL without blood transfusion or use of erythropoietin in the past 14 days;Total bilirubin ⩽ 1.5 x upper limit of normal (ULN); Aspartate aminotransferase (AST) and alanine aminotransferase (ALT) ⩽ 2.5 x ULN (for patients with liver metastases, ALT or AST ⩽ 5 x ULN is allowed); Serum creatinine ⩽ 1.5 x ULN and creatinine clearance (calculated using the Cockcroft-Gault formula) ⩾ 60 mL/min; Adequate coagulation function, defined as international normalized ratio (INR) or prothrombin time (PT) ⩽ 1.5 x ULN; Normal thyroid function, defined as thyroid-stimulating hormone (TSH) within the normal range; If baseline TSH is outside the normal range, subjects can be included if total T3 (or FT3) and FT4 are within the normal range; Normal range for myocardial enzyme levels (subjects may also be included if the investigator judges that the abnormal laboratory results have no clinical significance); For women of childbearing potential, a negative urine or serum pregnancy test must be obtained within 3 days prior to the first dose of study medication (Cycle 1, Day 1); If the urine pregnancy test cannot be confirmed as negative, a blood pregnancy test is required. Women not of childbearing potential are defined as those who have been menopausal for at least one year or have undergone surgical sterilization or hysterectomy; If there is a risk of pregnancy, all subjects (regardless of gender) must use contraception with a failure rate of less than 1% throughout the treatment period and for 120 days after the last dose of study medication (or 180 days after the last chemotherapy).

#### 4.3 Exclusion Criteria

• Diagnosis of malignancies other than head and neck squamous cell carcinoma within 5 years prior to the first administration (excluding completely resected basal cell carcinoma, squamous cell carcinoma of the skin, and/or completely resected carcinoma in situ).

• Currently participating in an interventional clinical trial treatment, or received other investigational drugs or used investigational devices within 4 weeks prior to the first administration.

• Previously received the following therapies: anti-PD-1, anti-PD-L1, or anti-PD-L2 drugs, or medications targeting another stimulatory or co-inhibitory T cell receptor (e.g., CTLA-4, OX-40, CD137).

• Received systemic therapy with traditional Chinese medicine or immunomodulatory drugs with indications for anti-head and neck squamous cell carcinoma within 2 weeks prior to the first administration (including thymosin, interferon, interleukin, excluding local use for the control of pleural effusion)

• Active autoimmune diseases requiring systemic treatment (e.g., use of disease-modifying drugs, glucocorticoids, or immunosuppressants) occurring within 2 years prior to the first administration. Alternative therapies (e.g., thyroid hormone, insulin, or physiological glucocorticoids for adrenal or pituitary insufficiency) are not considered systemic treatment

• Receiving systemic glucocorticoid treatment (excluding nasal, inhaled, or other forms of topical glucocorticoids) or any other form of immunosuppressive therapy within 7 days prior to the first administration of the study drug.

• Presence of clinically uncontrollable pleural effusion/ascites (patients who do not require drainage or have had drainage stopped for 3 days with no significant increase in effusion can be enrolled)

• Known allogeneic organ transplantation (excluding corneal transplantation) or allogeneic hematopoietic stem cell transplantation.

• Known allergy to the active ingredients or excipients of the study drug.

• Not fully recovered from any toxicities and/or complications arising from any interventions before starting treatment (i.e., ≤ grade 1 or returned to baseline, excluding fatigue or hair loss)

• Known history of human immunodeficiency virus (HIV) infection (i.e., positive for HIV 1/2 antibodies).

• Untreated active hepatitis B (defined as HBsAg positive with HBV-DNA copy number exceeding the upper limit of normal values at the study center).

• Participants with active HCV infection (HCV antibody positive and HCV-RNA levels above the detection limit).

• Received live vaccines within 30 days before the first administration (Cycle 1, Day 1).

• Pregnant or breastfeeding women.

• Presence of any serious or uncontrolled systemic diseases, such as significant and severely symptomatic abnormalities in rhythm, conduction, or morphology on resting electrocardiogram, including complete left bundle branch block, second-degree or higher heart block, ventricular arrhythmias, or atrial fibrillation; unstable angina; congestive heart failure with New York Heart Association (NYHA) classification ≥ 2; any arterial thromboembolism or ischemic event within the 6 months prior to enrollment, such as myocardial infarction, unstable angina, cerebrovascular accident, or transient ischemic attack; poorly controlled blood pressure (systolic blood pressure > 140 mmHg, diastolic blood pressure > 90 mmHg); history of non-infectious pneumonia requiring corticosteroid treatment within 1 year prior to the first administration, or currently active clinical interstitial lung disease; active tuberculosis; presence of active or uncontrolled infections requiring systemic treatment; clinically active diverticulitis, abdominal abscess, or gastrointestinal obstruction; liver diseases such as cirrhosis, decompensated liver disease, or acute or chronic active hepatitis; poorly controlled diabetes (fasting blood glucose (FBG) > 10 mmol/L); urinalysis indicating urine protein ≥ ++, confirmed by 24-hour urine protein quantification > 1.0 g; patients with mental disorders who are unable to cooperate with treatment.

• History or evidence of diseases that may interfere with trial results, hinder the subject's full participation in the study, abnormal treatment or laboratory test values, or other conditions deemed unsuitable for enrollment by the investigator due to potential risks.

#### 4.4 Criteria for Discontinuing Treatment/Withdrawing from the Study

##### 4.4.1 Stop Study Treatment

Participants may discontinue treatment at any time for any reason, or the investigator may decide to stop treatment in the event of any adverse events. Additionally, if a participant is deemed unsuitable for treatment, violates the study protocol, or for management and/or other safety reasons, the investigator may discontinue the participant's treatment.

For any of the following reasons, participants must discontinue treatment but may continue to receive monitoring in the study:

- The participant or their legal representative requests to discontinue treatment.
- Adverse events occur that require discontinuation of treatment as specified in the protocol.
- Another malignant tumor requiring active treatment occurs.
- Complications occur that hinder further treatment.
- The investigator decides to withdraw the participant from the study.
- The participant's serum pregnancy test result is positive.
- Poor participant compliance.
- The investigator believes that continuing to administer the study drug would place the participant at unnecessary risk based on their medical condition or personal circumstances.
- Completion of treatment as specified in the protocol.

##### 4.4.2 Withdrawal from the Study

If the participant or their legal representative withdraws their informed consent to participate in the study, the participant must be withdrawn from the study. If the participant withdraws from the study, they will no longer receive treatment or attend scheduled visits. With the participant's consent, post-withdrawal, they may receive survival follow-up. If the participant is lost to follow-up, they must be withdrawn from the study.

#### 4.5 Statistical Analysis

##### 4.5.1 Analysis Set

- **Full Analysis Set:** Effectiveness analysis will be conducted for all enrolled cases that received at least one dose of the drug according to the Intent-to-Treat (ITT) principle. For cases with incomplete treatment data, the last observation carried forward (LOCF) method will be used to carry forward the last observed data to the final study results.

- **Per-protocol Set:** All cases that meet the study protocol criteria, have good compliance, have not taken prohibited medications during the trial, and have completed the required content of the case report form. No imputation will be performed for missing data. The efficacy of the drug will be statistically analyzed for both the Full Analysis Set (FAS) and the Per Protocol Set (PPS).

- **Safety Analysis Set:** All enrolled cases that have received at least one dose of the investigational drug and have safety records post-treatment will be included in the Safety Analysis Set. This dataset will be used for safety analysis.

##### 4.5.2 Statistical Software

Statistical analysis will be performed using SAS 9.2 (or a higher version) statistical software.

##### 4.5.3 Missing Data

This trial does not apply special handling for missing data related to efficacy endpoints, and no estimation will be performed for missing values in safety assessments.

##### 4.5.4 Descriptive Statistics

In this study, unless otherwise specified, data will be summarized using descriptive statistics based on the following general principles. Continuous data will be described using median (minimum, maximum), while categorical data will be summarized using frequency (percentage).

##### 4.5.5 Analysis of Baseline Data

Calculate the mean, standard deviation, median, maximum, and minimum for quantitative data such as age, height, and weight. For qualitative data such as gender and ECOG scores, list the frequency and percentage.

##### 4.5.6 General Statistical Analysis Methods

This study will conduct statistical analysis of the dosing regimen to calculate drug exposure throughout the trial. Baseline data will be analyzed using the Full Analysis Set (FAS), with all efficacy endpoints assessed using both the FAS and the Per Protocol Set (PPS), categorized by randomization group. Safety analysis will use the Safety Analysis Set (SAS), categorized by actual treatment group. If a patient receives incorrect treatment in one cycle but correct treatment in others, they will be analyzed according to the correct treatment group for safety. Continuous data from each visit will be described using mean ± standard deviation or median (minimum, maximum), while categorical data for each visit in the trial group will be summarized using frequency (proportion).

• The major pathological response rate (MPR) is defined as the proportion of participants with viable tumor cells ≤10% in the resected specimens. MPR is calculated as the number of participants achieving MPR divided by the total number of participants, multiplied by 100%. A binomial distribution will be used to calculate its 95% confidence interval (CI).

• Overall survival (OS) is defined as the time from the first use of the study drug until the participant's death. If a participant is still alive at the end of the study, the last known survival date will be used as the censoring date. Comparisons between OS groups will be conducted using the log-rank test, and a Cox proportional hazards model will estimate the hazard ratio (HR) and its 95% confidence interval (CI) between groups. The median OS and its 95% CI will be estimated using the Kaplan-Meier method, along with the creation of survival curves.

• Disease-free survival (DFS) is defined as the time from treatment until the date of the first relapse (local/regional recurrence or distant metastasis) or death (from any cause) whichever comes firsts and regardless of whether the patient withdraws from treatment or receives another anti-cancer therapy prior to disease relapse. The DFS rate will be estimated using the Kaplan-Meier method.

• Objective response rate (ORR) is defined as the proportion of participants who achieve complete response (CR) or partial response (PR) out of the total participants.

ORR is calculated as (number of participants with CR + number of participants with PR) divided by the total number of participants, multiplied by 100%. A binomial distribution will be used to calculate its 95% confidence interval (CI). The calculation of the objective response rate is based on the best efficacy assessment of the tumors confirmed during the study.

• The safety analysis was conducted using the SS population, with the primary safety endpoints including study-related adverse events (occurring within 90 days after the first administration of tiragolumab or within 30 days post-surgery, whichever is later), grade 3 or higher study drug-related adverse events, and the occurrence of surgical complications. Surgical complications are defined as grade 3 or severe complications occurring intraoperatively or in the perioperative period.

##### 4.6 Informed Consent

Clinical researchers must fulfill the obligation to provide comprehensive information to participants, explaining that participation in the clinical trial is entirely voluntary and that they have the right to withdraw at any time without discrimination or retaliation, without affecting their medical treatment or rights. Participants should be informed that their involvement and personal data will remain confidential. They must also be informed about the nature of the trial, its purpose, potential benefits, and possible risks and inconveniences. Additionally, participants should be made aware of other available treatment options and their rights and obligations according to the Declaration of Helsinki. Sufficient time must be given for participants to consider their willingness to participate and to sign the informed consent form.

##### 4.7 Research-related Ethics

This protocol, along with the written informed consent form and any materials directly related to the participants, must be submitted to the ethics committee for written approval before officially commencing the research. Researchers are required to submit annual reports to the ethics committee at least once a year, if applicable. Upon the cessation or completion of the study, researchers must notify the ethics committee in writing. Any changes in the research process (such as modifications to the protocol and/or informed consent) must be promptly reported to the ethics committee, and such changes cannot be implemented without prior approval, except for modifications made to eliminate apparent and direct risks to participants. In such cases, the ethics committee will be notified.

#### 4.8 Confidentiality Measures

The information of patients participating in this project will be kept confidential. While the findings from this research may be published in medical journals, we will protect patient information as required by law, and personal information will not be disclosed unless mandated by relevant legal requirements. If necessary, regulatory authorities and the hospital's ethics committee, along with their personnel, may access patient data in accordance with regulations.

#### 4.9 Follow-Up

Safety follow-up will be conducted 30±7 days after the last administration of medication or before the start of new antitumor treatment, whichever comes first. All adverse events (AEs) occurring before the safety follow-up visit must be recorded until they resolve to grade 0-1 or baseline levels, or until the investigator determines that further follow-up is unnecessary for valid reasons (whichever comes first). Serious adverse events (SAEs) occurring within 90 days after the last medication or before the initiation of new antitumor therapy will also be followed up and recorded. Survival follow-up will occur every 90 days (±7 days) after the safety visit, with phone visits being acceptable.

### 5 SAFETY MONITORING AND REPORTING

#### 5.1 Adverse Events

An adverse event (AE) in clinical research is defined as any unfavorable and unintended medical occurrence that begins after a participant signs the informed consent form, regardless of whether it is related to the study drug. AEs include, but are not limited to, the following situations:

• Worsening of pre-existing medical conditions/diseases (including exacerbation of symptoms, signs, or laboratory abnormalities) prior to entering the clinical trial.

• Any new adverse medical conditions (including symptoms, signs, or newly diagnosed diseases).

• Abnormal clinically significant laboratory results.

#### 5.2 Serious Adverse Events

A serious adverse event (SAE) is defined as an adverse event that meets at least one of the following criteria:

• Results in death, excluding death due to disease progression related to the study indication.

• Is life-threatening (defined as when the AE poses a risk of death to the participant at the time of occurrence, not including AEs that may cause death only if they worsen).

• Requires hospitalization or prolongation of existing hospitalization, excluding the following: Rehabilitation facilities, Nursing homes, Routine emergency room admissions, Same-day surgeries (e.g., outpatient/non-inpatient surgeries), Hospitalizations or extensions of stay not related to the worsening of an AE (e.g., hospitalization for pre-existing conditions without new AEs).

• Results in permanent or significant disability/incapacity.

• Results in congenital anomalies/birth defects.

• Other important medical events: defined as events that pose a risk to the participant or require medical intervention to prevent any of the above outcomes.

#### 5.3 Assessment of Adverse Events

The investigators will assess all adverse events according to the NCI Common Terminology Criteria for Adverse Events (CTCAE) version 5.0. Any adverse events that result in a change in CTCAE grade will be recorded in the adverse event case report form/workbook. All adverse events, regardless of CTCAE grade, must be evaluated for whether they constitute a serious adverse event.

Detailed criteria for adverse event assessment are provided in the figure below:

**Common Terminology Criteria for Adverse Events (CTCAE) v5.0**
**Publish Date: November 27, 2017**

**Introduction**
The NCI Common Terminology Criteria for Adverse Events is a descriptive terminology which can be utilized for Adverse Event (AE) reporting. A grading (severity) scale is provided for each AE term.

**SOC**
System Organ Class (SOC), the highest level of the MedDRA[1] hierarchy, is identified by anatomical or physiological system, etiology, or purpose (e.g., SOC Investigations for laboratory test results). CTCAE terms are grouped by MedDRA Primary SOCs. Within each SOC, AEs are listed and accompanied by descriptions of severity (Grade).

**CTCAE Terms**
An Adverse Event (AE) is any unfavorable and unintended sign (including an abnormal laboratory finding), symptom, or disease temporally associated with the use of a medical treatment or procedure that may or may not be considered related to the medical treatment or procedure. An AE is a term that is a unique representation of a specific event used for medical documentation and scientific analyses. Each CTCAE v4.0 term is a MedDRA LLT (Lowest Level Term).

**Grades**
Grade refers to the severity of the AE. The CTCAE displays Grades 1 through 5 with unique clinical descriptions of severity for each AE based on this general guideline:

**Grade 1** Mild; asymptomatic or mild symptoms; clinical or diagnostic observations only; intervention not indicated.
**Grade 2** Moderate; minimal, local or noninvasive intervention indicated; limiting age-appropriate instrumental ADL*.
**Grade 3** Severe or medically significant but not immediately life-threatening; hospitalization or prolongation of hospitalization indicated; disabling; limiting self care ADL**.
**Grade 4** Life-threatening consequences; urgent intervention indicated.
**Grade 5** Death related to AE.

A Semi-colon indicates 'or' within the description of the grade.

A single dash (-) indicates a Grade is not available. Not all Grades are appropriate for all AEs. Therefore, some AEs are listed with fewer than five options for Grade selection.

**Grade 5**
Grade 5 (Death) is not appropriate for some AEs and therefore is not an option.

**Definitions**
A brief Definition is provided to clarify the meaning of each AE term. A single dash (-) indicates a Definition is not available.

**Navigational Notes**
A Navigational Note is used to assist the reporter in choosing a correct AE. It may list other AEs that should be considered in addition to or in place of the AE in question. A single dash (-) indicates a Navigational Note has not been defined for the AE term.

**Activities of Daily Living (ADL)**
*Instrumental ADL refer to preparing meals, shopping for groceries or clothes, using the telephone, managing money, etc.
**Self care ADL refer to bathing, dressing and undressing, feeding self, using the toilet, taking medications, and not bedridden.

[1] CTCAE v5.0 incorporates certain elements of the MedDRA terminology. For further details on MedDRA refer to the MedDRA MSSO Web site (https://www.meddra.org/).

**Figure 2: CTCAE Version 5.0**

### 6 Data Management

#### 6.1 Electronic Case Report Form

This study utilizes an electronic case report form (eCRF). For each patient who enters screening (after signing the informed consent), the eCRF must be completed and signed by the principal investigator or a delegated research staff member. The investigator must ensure the accuracy and completeness of all data.

#### 6.2 Data Query and Response

While data entry is performed in the EDC system, the system can automatically generate queries regarding issues such as dates, eligibility criteria, exclusion criteria, dropout, and missing values. Data managers can also manually issue queries on data points within the EDC system. Investigators can respond to queries online directly, or monitors can download a data query form, which the investigator will answer in writing and sign before being entered into the system by research assistants. The offline query forms will be properly stored at each research center. Investigators should respond to system-generated queries as promptly as possible, and if necessary, additional queries may be issued after the responses are provided.

#### 6.3 Data Review and Database Lock

After all data queries in the system have been resolved, 'clean' data will be exported and submitted to the statistical personnel, along with the finalized statistical analysis plan. Once the investigator submits their electronic signature and the finalized statistical plan on the EDC, the data will be locked, and statistical analysis will be conducted according to the statistical analysis plan.

### 7 STUDY ASSESSMENT

Throughout the study, patient safety and tolerability will be closely monitored. All assessments for each patient will be conducted and documented in their medical records. Administration of the study drug will only occur after reviewing clinical evaluations and local laboratory results (which must be obtained prior to each administration) to ensure compliance with the dosing criteria outlined in the study protocol.

#### 7.1 Screening

Screening assessments will be conducted within 28 days prior to enrollment, and patients who agree to participate will sign the Informed Consent Form (ICF) before undergoing any screening procedures. The screening period begins on the first day the screening procedures are performed. Screening assessments may be repeated as necessary during this period; the investigator will evaluate the patient's eligibility based on the most recent screening assessment results. Standard tests or examination results obtained ≤28 days prior to obtaining informed consent may be used for screening assessments, and these standard tests do not need to be repeated unless otherwise specified.

##### 7.1.1 Demographic Data and Medical History

Demographic Data will include date of birth (or age), gender, and self-reported ethnicity/race. It will encompass various clinically relevant medical histories, including past illnesses, surgeries, or cancer history; reproductive status; and history of alcohol and tobacco use. The cancer history will include assessments of previous treatments, including treatment start and end dates, best response, and reasons for treatment cessation. Imaging results collected prior to study enrollment will also be gathered.

##### 7.1.2 Women of Childbearing Potential and Contraception

Fertility is defined as the physiological capability to conceive. For guidance on contraception and definitions of "women of childbearing potential" and "women not of childbearing potential," please refer to Appendix 2.

##### 7.1.3 Informed Consent and Screening Records

Prior to executing any study-specific procedures, written informed consent must be obtained from participants voluntarily agreeing to participate in the study. Informed consent forms (ICF) for all enrolled and screened but not enrolled patients will be retained at the research center. All screening assessments must be completed and reviewed before confirming that a patient meets all eligibility criteria for enrollment. The investigator will maintain a screening log documenting detailed information for all screened patients, noting those who qualify for enrollment or recording reasons for screening failures.

#### 7.2 Enrollment

##### 7.2.1 Eligibility Confirmation

The investigator will assess the eligibility of each patient. All screening procedures and relevant medical history must be obtained before determining eligibility. All inclusion criteria must be met, and no exclusion criteria may be violated. No waivers are allowed for any eligibility criteria. After screening, the investigator must confirm the patient's eligibility for enrollment.

##### 7.2.2 Patient ID

After obtaining the informed consent form, the staff at the research center will assign a unique patient ID to the potential study participant.

#### 7.3 Safety Assessment

##### 7.3.1 Vital Signs

Vital signs include measuring temperature (°C), pulse, and blood pressure (systolic and diastolic, measured after the patient has been seated at rest for 10 minutes).Height (baseline only) and weight should be measured and recorded in the eCRF.

The first two infusions (Day 1 of Cycle 1 and Day 1 of Cycle 2) should be completed within 60 minutes. If well tolerated, subsequent infusions can be completed within 30 minutes. Monitoring must be conducted 1 hour before, during, and 1 hour after the first two infusions of Tremelimumab. Starting from Cycle 3, vital signs will be collected within 60 minutes before infusion, and if clinically indicated, during and 30 minutes after infusion.

Patients will be informed about the possibility of delayed symptoms after infusion and instructed to contact their study physician if they experience such symptoms.

##### 7.3.2 Physical Examination

During the screening visit, a physical examination will be conducted, which includes assessments of: 1) head, eyes, ears, nose, and throat; 2) cardiovascular system; 3) skin; 4) musculoskeletal system; 5) respiratory system; 6) gastrointestinal system; and 7) nervous system. Any abnormalities identified during the screening process will be graded according to the NCI-CTCAE version 5.0 and recorded in the eCRF using appropriate medical terminology.

In subsequent visits (and as clinically indicated), a limited, symptom-targeted physical examination will be performed. Changes from baseline will be recorded. New or worsening clinically significant abnormalities should be documented as AEs in the eCRF.

##### 7.3.3 Eastern Cooperative Oncology Group (ECOG) Performance Status

The ECOG Performance Status will be assessed during the study period (Appendix 1).

##### 7.3.4 Laboratory Safety Assessments

Assessments of serum biochemistry, hematology, coagulation, urinalysis, stool analysis, infectious disease testing, pregnancy testing, thyroid function, and blood type will be performed at a local laboratory.

If laboratory tests required for screening have not been conducted within 7 days prior to enrollment, these tests should be repeated before the administration of the study drug, and the results should be reviewed. Hematology and serum biochemistry (including liver, kidney, and cardiac function tests) should be conducted at the start of subsequent cycles and as clinically indicated, as specified in Appendix 1. The investigator may use results from the local laboratory for eligibility assessment, safety monitoring, and dosing decisions.

##### 7.3.5 Electrocardiogram

For the purpose of safety monitoring, the investigator must review all 12-lead electrocardiogram (ECG) recordings, sign, and date them. Both paper and electronic versions of the ECG will be maintained as a permanent part of the patient's study records at the study center. The 12-lead ECGs will be performed by qualified personnel at the specified time points outlined in the 'Trial Flowchart.'

All ECG assessments must be conducted after the subject has rested quietly for at least 10 minutes. The content of the ECG should include at least: heart rate, QT interval, QT corrected for heart rate (QTcF), and P-R interval. Three ECG checks are required during the screening period, with a minimum interval of 5 minutes between each. The QTcF can be calculated using the formula QTcF = QT / (heart rate)^0.33. If the subject experiences symptoms such as chest pain or palpitations, the investigator may consider additional ECGs and/or cardiac enzyme testing.

##### 7.3.6 Cardiac Ultrasound

The echocardiogram during the screening period will be conducted by a qualified physician, who will measure the Left Ventricular Ejection Fraction (LVEF).

##### 7.3.7 Pulmonary Function

The pulmonary function tests during the screening period will be conducted by a qualified physician, who will evaluate the tolerance for curative surgery.

##### 7.3.8 Adverse Events

Throughout the study, adverse events (AEs) related to the drug and curative surgery will be graded and recorded according to the NCI-CTCAE version 5.0 and the Clavien-Dindo classification system. Characterization of toxicity will include severity, duration, and time to onset.

#### 7.4 Biomarkers

The transportation, storage, and processing of blood samples for biomarker assessment, archived tumor samples, fresh tumor samples, and residual tumor tissue samples will be managed uniformly by the central laboratory. For returns, please contact the coordinating institution.

#### 7.5 Visit Window

All visits during the neoadjuvant treatment period must be conducted within a window of -3 to +7 days of the scheduled date, while postoperative visits have a window of ±14 days, unless otherwise specified. All assessments will occur on the designated visit day unless an acceptable time window is specified. Evaluations scheduled for the day of study treatment administration (Day 1) should be completed prior to the infusion/administration of the study treatment, unless otherwise stated. Laboratory and imaging results should be reviewed prior to administration.

If the scheduled study visit time points conflict with holidays, weekends, or other events/legal regulations, the visit may be rescheduled to the nearest feasible date.

#### 7.6 Unscheduled visit

Unscheduled visits may occur at any time based on the request of the patient or investigator. These visits may include: vital signs; targeted physical examinations; ECOG performance status; AE review; review of concomitant medications and procedures; imaging assessments; evaluation of disease-related systemic symptoms; and hematological and biochemical laboratory assessments. The date and reason for unscheduled visits must be recorded in the source documents.

If an unscheduled visit is necessary to assess toxicity or suspected progressive disease (PD), diagnostic tests may be conducted as deemed appropriate by the investigator, and the results will be recorded in the eCRF under unscheduled visits.

### 8 Quality Management

#### 8.1 Ethical Standards

This study will be conducted by the principal investigator and research center in full compliance with ICH E6 guidelines on Good Clinical Practice and the principles of the Declaration of Helsinki, or the legal regulations of the country where the research is conducted, prioritizing the protection of participants. The study will also adhere to the requirements outlined in ICH E2A guidelines (Clinical Safety Data Management: Definitions and Standards for Expedited Reporting).

#### 8.2 Institutional Review Board / Independent Ethics Committee

The principal investigator must submit the study protocol, informed consent form (ICF), any information provided to patients, and relevant supporting documents to the IRB/IEC for review and approval prior to the initiation of the study. Additionally, any patient recruitment materials must be approved by the IRB/IEC.

The principal investigator is responsible for providing an annual written summary of the study status to the IRB/IEC in accordance with their requirements, policies, and procedures. The investigator must also promptly notify the IRB/IEC of any protocol amendments. Serious Adverse Events (SAEs) must be reported to local health authorities and the IRB/IEC as required.

Any protocol amendments will be drafted by the investigator. All modifications must be submitted to regulatory authorities according to local requirements, along with the revised ICF template for IRB/IEC review. The investigator must obtain written documentation and approvals from the appropriate authorities (as per local regulations) and the IRB/IEC before implementing any changes, except for those necessary to eliminate direct harm to participants or those that involve only logistical or administrative changes (e.g., changes to the medical monitor or contact information).

Participants who are actively involved in the study must be informed of any changes in risks and/or scope, and they must read, understand, and sign each revised ICF to confirm their willingness to remain in the study.

#### 8.3 Informed Consent

The ICF must be signed and dated by the participant or the participant's legally authorized representative before the participant enters the study. The informed consent process for each participant should be documented in their medical records or clinical notes, and written informed consent must be obtained prior to participation in the study.

During the study, participants must re-sign the most current version of the ICF (or a supplementary document for significant new information/findings as required by applicable laws and IRB/IEC policies). For any updated or revised ICF, the informed consent process must be documented in each participant's medical records or clinical notes, and written informed consent must be obtained using the updated/revised ICF before continuing participation in the study.

A copy of each signed ICF must be provided to the participant or their legally authorized representative. All signed and dated ICFs must be retained in the study documents or site files for each participant and must be readily available for verification by study monitors.

#### 8.4 Considerations for the Informed Consent Process

• Ensure that each patient is informed in detail, both orally and in writing, about the nature, purpose, potential risks, and benefits of the research.

• Ensure that each patient understands their right to withdraw from the study at any time.

• Ensure that each patient has the right to ask questions and has time to consider the information provided.

• Ensure that each patient signs and dates the informed consent form prior to participating in the study.

• Ensure that the original signed ICF is stored in the investigator's file.

• Ensure that each patient has a signed copy of the ICF.

• Ensure that the benefits obtained from the patients entering the study and the preventive measures for any harm resulting from the trial are described in the ICF approved by the ethics committee.

### 9 Risk Control

#### 9.1 Overall Plan for Managing Safety Issues

##### 9.1.1 Inclusion/Exclusion Criteria

In this study, qualified inclusion and exclusion criteria were established to protect patient safety. Considering the non-clinical and clinical data for Tremelimumab and other PD-L1/PD-1 inhibitors, patients with a history of acute autoimmune diseases or those at risk of relapsing autoimmune conditions, patients who have undergone allogeneic stem cell or organ transplantation, and patients who received live virus vaccines within 28 days prior to randomization will be excluded. Additionally, patients with contraindications to chemotherapy will also be excluded from the study.

##### 9.1.2 Safety Monitoring Plan

All adverse events (AEs) will be defined and graded according to the NCI-CTCAE v5.0 criteria. These AEs will be monitored to assess safety in this study. Clinical laboratory test results must be reviewed prior to the start of each cycle.

In this study, all enrolled patients will undergo regular clinical assessments and standard laboratory tests before and during participation. Safety evaluations will include medical interviews, recording of AEs, physical examinations, laboratory indicators (hematology, biochemistry, etc.), and other assessments. Additionally, patients will be closely monitored for any signs or symptoms of autoimmune diseases and infections.

Tremelimumab administration will occur in settings equipped with emergency medical equipment and staffed by personnel trained to handle emergencies. All AEs must be recorded during the study (AEs from the first dose and SAEs from the signing of informed consent) until 90 days after the last dose or until another anticancer treatment is initiated, whichever occurs first.

At the end of treatment, AEs related to the study treatment that have not resolved will require ongoing follow-up until the event resolves to baseline levels or $\leqslant$ grade 1, unless the investigator determines the event is stable, the patient is lost to follow-up, withdraws consent, or it is determined that the study treatment or participation is not the cause of the AE.

Investigators must record all drug-related SAEs occurring after treatment cessation until patient death, withdrawal of consent, or loss to follow-up, whichever occurs first.

Investigators are required to report all adverse events, including pregnancy-related AEs. Potential safety issues anticipated in this study, along with measures taken to avoid or minimize such toxicities, are outlined in the following sections.

#### 9.2 Adverse Events

##### 9.2.1 Definition and Report

Adverse events (AEs) are defined as any undesirable and unexpected signs (including abnormal laboratory results), symptoms, or diseases (new or worsening) associated with the use of a study drug, regardless of their relation to the drug. Examples of AEs include:

• Worsening of chronic or intermittent pre-existing conditions, including increases in severity, frequency, or duration, and/or significant outcomes.

• New conditions discovered or diagnosed after the administration of the study drug, even if they may have existed before the study began.

• Signs, symptoms, or clinical sequelae that could interact with each other.

• Suspected signs, symptoms, or clinical sequelae related to overdose of the study drug or concomitant medications (overdose itself should not be reported as an AE or SAE).

If an AE or serious adverse event (SAE) occurs, the investigator is responsible for reviewing all relevant documentation (e.g., hospital progress notes, lab results, and diagnostic reports). The investigator must then record detailed information about the AEs in the case report form (CRF), including the nature, severity, timing, management, and outcome. SAEs must be reported per SFDA regulations, with specific requirements to report them to the hospital GCP center and ethics committee within 24 hours.

##### 9.2.2 Assessment of Severity

The investigator will assess the severity of each adverse event (AE) and serious adverse event (SAE) reported during the study. AEs and SAEs should be evaluated and graded according to the NCI-CTCAE Version 5.0 criteria. This standardized system provides a consistent framework for assessing the severity of AEs, facilitating communication and reporting throughout the study.

##### 9.2.3 Causality Assessment

Investigators are obligated to use their best clinical judgment to assess the relationship between the study drug and each adverse event (AE) or serious adverse event (SAE). They should consider and investigate other causes, such as the natural history of underlying diseases, concomitant treatments, other risk factors, and the temporal relationship between the AE or SAE and the study drug.

In the event of an SAE, each SAE should be assessed for causality, and the investigator may adjust their assessment based on follow-up information. Investigators are responsible for evaluating the causality of each AE and classifying it as "related" or "unrelated." An AE is considered related if there is a "reasonable possibility" that it may have been caused by the study drug (i.e., there are facts, evidence, or arguments suggesting a potential causal relationship).

When conducting this assessment, several factors should be considered, including:

- The temporal relationship between the AE and the administration of the study treatment/procedure.
- Whether an alternative causative agent can be identified.
- The mechanism of action of the study drug.
- Biological plausibility.

An AE should be deemed "related" to the study drug if it meets any of the following criteria; otherwise, the event should be assessed as unrelated:

- Clear evidence indicating a causal relationship, excluding other possible contributing factors.
- Evidence suggesting a causal relationship with minimal influence from other factors.

• Evidence supporting a causal relationship (e.g., the AE occurs within a reasonable time frame after administration of the study drug). However, the influence of other factors (e.g., the patient's clinical condition or other concomitant AEs) may also contribute to the occurrence of the AE.

##### 9.2.4 Follow-Up for Adverse Events

After submitting an initial report of an adverse event (AE) or serious adverse event (SAE), investigators need to actively follow up with each patient. During the last visit/contact, they should document and highlight all ongoing AEs and SAEs, which will continue to be reviewed in subsequent visits/contacts.

Follow-up is required for all AEs and SAEs until they are resolved, the patient's condition stabilizes, the event is deemed chronic, an alternative explanation is identified, the patient is lost to follow-up, or the patient withdraws consent. Investigators should ensure that follow-up includes various supplementary evaluations that can explain the nature and/or causality of the AE or SAE. These may include additional laboratory tests, diagnostic investigations, histopathological examinations, imaging studies, or consultations with other healthcare professionals.

##### 9.2.5 Laboratory Abnormalities

Abnormal laboratory results judged by the investigator to be clinically significant (such as clinical biochemistry, complete blood count, coagulation, or urinalysis) or other abnormal assessments (such as ECG, X-rays, or vital signs) will be recorded as adverse events (AEs) or serious adverse events (SAEs). This includes clinically significant laboratory abnormalities or other abnormal assessments that existed at baseline and worsened significantly during the study. The definition of clinical significance is determined by the investigator. Generally, abnormal laboratory results or other abnormal assessments refer to those that are:

• Related to clinical symptoms or signs, or

• Require active medical intervention, or

• Result in dose interruption or discontinuation, or

• Require close monitoring, more frequent follow-up evaluations, or further diagnostic investigations.

#### 9.3 Definition of Serious Adverse Event (SAE)

SAE refers to any adverse medical event at any dose level that:

• Results in death

• Is life-threatening (Note: The term "life-threatening" refers to an event where the patient is at immediate risk of death when the AE occurs, not an event that could potentially lead to death if it were more severe.)

• Requires hospitalization or prolongation of existing hospitalization (Note: Generally, hospitalization means the patient is admitted to a hospital or emergency room for observation and/or treatment that cannot be conveniently provided in a physician's office or outpatient setting, typically for at least one night.)

• Results in disability/incapacity (Note: "Disability" refers to a significant impairment of a person's ability to conduct normal life activities. This definition does not include experiences that are relatively minor in medical significance, such as uncomplicated headaches, nausea, vomiting, diarrhea, flu, and minor injuries (e.g., sprained ankle) that may disrupt or affect daily life but do not constitute significant impairment.)

• Results in congenital anomaly/birth defect

• Based on medical judgment, the investigator considers this an AE of medical significance (e.g., it may pose a risk to the patient or may require medical/surgical intervention to prevent one of the outcomes listed above).

The following situations will not be considered SAEs:

• Hospitalization for elective treatment of a pre-existing condition that has not worsened compared to baseline

• Hospitalization for social/convenience reasons

• Hospitalization for treatment related to the target disease of the study, including for transfusion support or convenience.

#### 9.4 Suspected Unexpected Serious Adverse Reaction

A suspected unexpected serious adverse reaction (SUSAR) is a serious adverse reaction that is both unexpected (i.e., not included in the product's reference safety information) and meets the definition of a serious adverse drug reaction. Its unexpectedness or seriousness is inconsistent with the information described in the investigator's brochure.

#### 9.5 The Occurrence Time, Frequency, and Methods of Obtaining Adverse Events and Serious Adverse Events

##### 9.5.1 Adverse Event Reporting Period

After signing the informed consent form, only SAEs should be reported before the administration of the study drug begins. After the administration of the study drug starts, all AEs and SAEs will be reported, regardless of their relationship to the study drug, until 90 days after the last dose of the study drug (including chemotherapy) or the start of a new anticancer treatment, whichever occurs first.

##### 9.5.2 Serious Adverse Event Report

If the investigator determines that the AE meets the definition of an SAE in the study protocol, the event must be reported immediately (within 24 hours) to the hospital's GCP center and the hospital ethics committee.

##### 9.5.3 Proactive Collection of Adverse Events

The investigator or designated personnel will inquire about AE by asking the following standard questions:

- How do you feel?
- Have you experienced any medical issues since the last visit?
- Have you taken any new medications since the last visit?

##### 9.5.4 Disease Progression

In this study population, disease progression (including fatal disease progression) that is expected to occur and measured as an efficacy endpoint should not be reported using the term AE. However, symptoms, signs, or clinical sequelae caused by disease progression should be reported as AEs.

For example, if a patient develops a pleural effusion due to lung metastasis from disease progression, the event should be reported as "pleural effusion," rather than as disease progression. If a patient experiences fatal multi-organ failure due to disease progression, the term "multi-organ failure" should be reported as an SAE, with death as the outcome, rather than reporting it as "fatal disease progression" or "death due to disease progression."

##### 9.5.5 Death

Death is an outcome and is generally not considered an event. If the only information obtained is death and the cause of death is unknown, then death can be reported as an event, such as "death," "cause of death unknown," or "unspecified cause of death."

##### 9.5.6 Pregnancy

If a female patient or the partner of a male patient becomes pregnant during the course of the study treatment or within 120 days after the last dose of tiragolumab or within 30 days after the last dose of chemotherapy, a pregnancy report form must be completed, and the outcome should be followed up. Generally, the follow-up period should not exceed 8 weeks after the expected due date. Any cases of pregnancy termination must be reported.

Although pregnancy itself is not considered an AE, all pregnancy complications or elective terminations for medical reasons should be recorded as AEs or SAEs.

Miscarriages, whether spontaneous, therapeutic, or natural, should be reported as SAEs. Similarly, if any children born to patients exposed to the investigational drug have congenital abnormalities/birth defects, these should be documented and reported as SAEs.

##### 9.5.7 Evaluation and Recording of Immune-Related Adverse Events

Due to the potential for anti-PD-1 therapy to cause autoimmune diseases, researchers consider immune-related AEs to be classified as irAEs and identified as such on the CRF AE page until 90 days after treatment cessation.

Researchers should refer to the diagnostic assessment and management guidelines for irAEs provided in Appendix 1, as irAEs are commonly seen with immune checkpoint inhibitors.

#### 9.6 Management of Adverse Events Requiring Special Attention

As a standard precaution, patients must be monitored for at least 1 hour in an area equipped with resuscitation equipment and emergency medications following the infusion of tiragolumab on Day 1 of Cycle 1 and Cycle 2. Starting from Cycle 3, monitoring for at least 30 minutes in an area with resuscitation equipment and emergency medications is required.

The management of infusion-related reactions, severe hypersensitivity reactions, and irAEs is outlined below according to NCI-CTCAE.

##### 9.6.1 Infusion-Related Reactions

Symptoms of infusion-related reactions include fever, chills/rigor, nausea, itching, angioedema, hypotension, headache, bronchospasm, urticaria, rash, vomiting, myalgia, dizziness, or hypertension. Severe reactions may include acute respiratory distress syndrome, myocardial infarction, ventricular fibrillation, and cardiogenic shock. Patients should be closely monitored for these reactions. If an infusion-related reaction occurs, the patient must be immediately transferred to an intensive care unit or equivalent setting and receive appropriate pharmacological treatment, including epinephrine, corticosteroids, intravenous antihistamines, bronchodilators, and oxygen.

**Table 3: Infusion-Related Reactions**

| NCI-CTCAE Levels | Adjustments for Tislelizumab Treatment |
|---|---|
| **Grade 1 – Mild**<br>Mild, transient reactions; does not indicate interruption of infusion; does not require intervention. | Reduce infusion rate by 50%. Closely monitor all worsening symptoms. Medical management as needed. Subsequent infusions should be administered after premedication before chemotherapy, completing the infusion at the reduced rate. |

| NCI-CTCAE Levels | Adjustments for Tislelizumab Treatment |
|---|---|
| **Grade 2 – Moderate**<br>Indicates interruption of treatment or infusion, but responds quickly to symptomatic treatment (e.g., antihistamines, NSAIDs, analgesics, intravenous fluids); suggests preventive medication should be administered within ≤24 hours. | Stop the infusion. If infusion-related reactions have resolved or the severity has decreased to Grade 1, the infusion can be resumed at 50% of the previous rate. Closely monitor all worsening symptoms. Appropriate medical management should be planned according to the following instructions. Subsequent infusions should be administered after premedication before chemotherapy, completing the infusion at the reduced rate. |
| **Grade 3 – Severe**<br>Prolonged (e.g., does not respond quickly to symptomatic treatment and/or requires a temporary interruption of infusion); symptoms recur after initial improvement; clinical sequelae suggest hospitalization. | Immediately stop the infusion. Appropriate medical management should be planned according to the following instructions. The patient should suspend treatment with the investigational drug. |
| **Grade 4 – Life-Threatening**<br>Life-threatening consequences; indicates the need for emergency treatment. | Immediately stop the infusion. Appropriate medical management should be planned according to the following instructions. The patient should suspend treatment with the investigational drug. Hospitalization is recommended. |

If the infusion rate of Tislelizumab decreases by 50% or is paused due to infusion-related reactions, the reduced rate must be maintained for all subsequent infusions, along with premedication used prior to chemotherapy. If the patient experiences a second infusion-related reaction (≥ Grade 2) at the slower infusion rate, the infusion should be stopped, and treatment with Tislelizumab should be discontinued.

NCI-CTCAE Grade 1 or 2 infusion reactions: Appropriate medical management measures should be developed based on the type of reaction. These may include but are not limited to antihistamines (e.g., diphenhydramine or equivalent), antipyretics (e.g., acetaminophen or equivalent), and if indicated, the use of corticosteroids, epinephrine, bronchodilators, and oxygen, either orally or intravenously. In the next cycle, the patient should receive oral antihistamines (e.g., diphenhydramine or equivalent) and antipyretics (e.g., acetaminophen or equivalent) as premedication and should be closely monitored for clinical signs and symptoms of infusion reactions.

NCI-CTCAE Grade 3 or 4 infusion reactions: Appropriate medical management measures should be immediately developed based on the type and severity of the reaction. These may include but are not limited to oral or intravenous antihistamines, antipyretics, corticosteroids, epinephrine, bronchodilators, and oxygen.

##### 9.6.2 Severe Allergic Reactions and Flu-like Symptoms

If an anaphylactic reaction occurs, the patient must be treated according to the best available medical practices as outlined in the complete guidelines for emergency management of allergic reactions developed by the Resuscitation Council (UK) working group [54]. Patients should be instructed to immediately report any delayed reactions to the investigator.

If a systemic allergic/anaphylactic reaction occurs (typically manifested within minutes after drug/antigen administration and characterized by: respiratory distress; throat edema; and/or severe bronchospasm; often followed by vascular collapse or shock, with previous respiratory difficulties resolving; skin manifestations such as itching and urticaria with or without edema; and gastrointestinal symptoms like nausea, vomiting, crampy abdominal pain, and diarrhea), the infusion must be stopped immediately, and arrangements should be made for the patient to withdraw from the study.

If an allergic reaction is observed, the patient will receive an injection of epinephrine and a dose of dexamethasone, followed by close monitoring and notification of the intensive care unit, with transfer if necessary.

To prevent flu-like symptoms, 25 mg of indomethacin or an equivalent dose of a non-steroidal anti-inflammatory drug (i.e., 600 mg of ibuprofen or 500 mg of naproxen sodium) may be administered 2 hours before and 8 hours after each study drug infusion. Alternative treatment for fever (i.e., acetaminophen) may be given at the investigator's discretion.

##### 9.6.3 Immune-Related Adverse Events

Immune-related AEs are of special interest in this study. If the events listed below or similar events occur, the investigator should exclude alternative explanations (eg, combination drugs, infectious disease, metabolic, toxin, PD or other neoplastic causes) with appropriate diagnostic tests, which may include but is not limited to serologic, immunologic, and histologic (biopsy) data. If alternative causes have been ruled out; the AE required the use of systemic steroids, other immunosuppressants, or endocrine therapy; and the AE is consistent with an immune-mediated mechanism of action, the irAE indicator in the eCRF should be checked.

Recommendation for diagnostic evaluation and management of irAEs is based on European Society for Medical Oncology and American Society of Clinical Oncology guidelines (Haanen et al 2017) and common immune-related toxicities are detailed in Appendix 1. For any AEs not included in Appendix 1, please refer to the American Society of Clinical Oncology Clinical Practice Guideline for further guidance on diagnostic evaluation and management of immune-related toxicities.

**Table 4: Immune-Related Adverse Events**

| **Body Organ System Affected** | **Events** |
|---|---|
| Skin (mild-common) | pruritus or maculopapular rash; vitiligo |
| Skin (moderate) | follicular or urticarial dermatitis; erythematous/lichenoid rash; acute febrile neutrophilic dermatosis |
| Skin (severe-rare) | full-thickness necrolysis/Stevens-Johnson syndrome |
| Gastrointestinal | colitis (includes diarrhea with abdominal pain or endoscopic/radiographic evidence of inflammation); pancreatitis; hepatitis; aminotransferase (ALT/AST) elevation; intestinal perforation |
| Endocrine | thyroiditis, hypothyroidism, hyperthyroidism; hypophysitis with features of hypopituitarism (eg, fatigue, weakness, weight gain); insulin-dependent diabetes mellitus; diabetic |

| Body Organ System Affected | Events |
| --- | --- |
| | ketoacidosis; adrenal insufficiency |
| Respiratory | pneumonitis/diffuse alveolitis |
| Eye | episcleritis; conjunctivitis; iritis/uveitis |
| Neuromuscular | arthritis; arthralgia; myalgia; neuropathy; Guillain-Barre syndrome;<br>aseptic meningitis; myasthenic syndrome/myasthenia gravis, meningoencephalitis; myositis |
| Blood | anaemia; leukopenia; thrombocytopenia |
| Renal | interstitial nephritis; glomerulonephritis; acute renal failure |
| Cardiac | pericarditis; myocarditis; heart failure |

If the toxicity takes too long a time to respond that the total duration of neoadjuvant therapy exceeds 12 weeks, investigational agents should be discontinued after consultation with the sponsor.

##### 9.6.4 Assessment and Management of Immune-Related Adverse Events

The following recommendations for the diagnosis and management of various immune-related adverse events (irAEs) will serve as a guideline. This document should be used in conjunction with the clinical judgment of experts (specialists experienced in the use of immunotherapy for cancer) and the guidelines or policies of individual institutions.

Standards for diagnosing irAEs include blood tests, diagnostic imaging, histopathology, and microbiological assessments to rule out other causes such as infections, progression of disease (PD), and side effects from concomitant medications. In addition to these test results, the following factors should be considered when diagnosing irAEs:

- Is there a temporal relationship between the occurrence of telegeneic monoclonal antibodies and the adverse event (AE)?
- How does the patient respond to the cessation of telegeneic monoclonal antibodies?
- Does the event recur upon re-initiation of telegeneic monoclonal antibodies?

- Is there clinical relief after corticosteroid use?
- Is the event indicative of an autoimmune endocrine disease?
- Is PD or another differential diagnosis a more reasonable explanation?

If alternative explanations for autoimmune toxicity are ruled out, the irAE fields related to AEs in the case report form (CRF) should be examined.

**Table 5: Recommended Diagnostic Tests for the Management of Potentially Related Immune-related Adverse Events**

| Immune-related Toxicity | Diagnostic Assessment Guidelines |
|---|---|
| Thyroid Disorders | Schedule and repeat thyroid function tests (TSH and T4) |
| Pituitary Inflammation | Examine the visual fields and consider the blood distribution of the pituitary endocrine axis. For patients presenting with headaches, vision disturbances, unexplained fatigue, weakness, weight loss, or nonspecific constitutional symptoms, conduct MRI of the pituitary gland and the entire brain. If abnormalities are found, consider consulting an endocrinologist. |
| Non-infectious Pneumonia | All patients presenting with new or worsening pulmonary symptoms or signs (such as upper respiratory infection, cough, dyspnea, or hypoxia) should be evaluated with high-resolution CT. Consider pulmonary function tests, including DLCO. Imaging findings are often nonspecific. Depending on the location of the lesions, bronchoscopy, bronchoalveolar lavage, or lung biopsy may be considered. If the cause is unclear, consult a pulmonologist. |
| Neurotoxicity | Conduct a comprehensive neurological examination and brain MRI for all patients with central nervous system symptoms. Review alcohol and other drug histories. Perform diabetes screening and assess serum B12/folate levels, HIV status, and thyroid function tests (TFT). Consider autoimmune serological tests. |

| Immune-related Toxicity | Diagnostic Assessment Guidelines |
|---|---|
| | Evaluate the need for brain/spine MRI/MRA, and conduct nerve conduction studies for peripheral neuropathy. If abnormalities are found, consult a neurologist. |
| Colitis | Assess dietary intake and rule out fatty diarrhea. Consider a comprehensive evaluation including FBC, UEC, LFT, CRP, TFT, stool microscopy and culture, viral PCR, C. difficile toxin, and Cryptosporidium (resistant organisms). If abdominal discomfort is present, consider imaging studies like X-ray or CT scan. For patients with bleeding, pain, or abdominal distension, consider a colonoscopy with possible biopsy and surgical intervention as appropriate. |
| Ocular Diseases | If a patient presents with acute, new, or worsening eye inflammation, blurred vision, or other visual disturbances, promptly refer them to an ophthalmologist for evaluation and treatment. |
| Hepatitis | Monitor alanine aminotransferase (ALT), aspartate aminotransferase (AST), total bilirubin, INR, and albumin levels; the frequency of testing will depend on the severity of adverse events (e.g., daily for grade 3 or 4; every 2-3 days for grade 2 until recovery). Review medication history (e.g., statins, antibiotics) and alcohol consumption. Conduct liver screening, including serological tests for hepatitis A, B, and C, PCR for hepatitis E, and assess for autoimmune markers (anti-ANA, SMA, LKM, SLA, LP, LCI) and iron studies. Consider imaging studies, such as ultrasound, to evaluate for metastasis or thromboembolism. Consult a liver specialist and consider liver biopsy if indicated. |
| Renal Toxicity | Review the patient's hydration status and medication history. Perform urinalysis and culture. Consider renal ultrasound, protein assessment (using dipstick or 24-hour urine collection), or phase-contrast microscopy. For further management assistance, consult a nephrologist. |
| Dermatology | Conduct a physical examination to consider other |

| Immune-related Toxicity | Diagnostic Assessment Guidelines |
|---|---|
| | potential causes. If a skin biopsy is necessary, refer the patient to a dermatologist for further evaluation. |
| Joint or Muscle Inflammation | Review the musculoskeletal history and perform a comprehensive musculoskeletal examination. Consider joint X-rays and other imaging studies as needed to rule out metastatic disease. Conduct autoimmune serology tests, and for further management assistance, consult a rheumatologist. If myositis, rhabdomyolysis, or myasthenia is suspected, include tests for creatine kinase (CK), erythrocyte sedimentation rate (ESR), C-reactive protein (CRP), and troponin, and consider a muscle biopsy. |
| Myocarditis | Perform an electrocardiogram (ECG), echocardiogram, and tests for creatine kinase (CK), CK-MB, and troponin (I and/or T). Consult a cardiologist for further evaluation and management. |

Treatment of Immune-Related Adverse Events (irAEs):

• Immune-related adverse events can escalate rapidly; consider treatment interruption, close monitoring, timely diagnostic assessments, and appropriate interventions for the patient.

• Following immunosuppressive therapy, irAEs should improve promptly. If there is no improvement, reassess the diagnosis, seek further expert advice, and contact the study medical monitor.

• For patients with grade 3 toxicities who improve quickly, and if there is evidence of a clinical response to the investigational treatment, reinitiation of the study drug may be considered after consulting the study medical monitor.

• Refer to the table for dosages of oral or intravenous (methyl)prednisolone. Alternative equivalent doses of other corticosteroids may be used. For steroid-refractory irAEs, consider non-steroidal immunosuppressants (e.g., mycophenolate mofetil).

• If the patient requires long-term immunosuppressive therapy, consider prophylactic antibiotics to prevent opportunistic infections.

**Table 6: Treatment of irAEs**

| **Immune-related Toxicity** | **Grade** | **Treatment Guidelines** | **Management of Investigational Drugs** |
|---|---|---|---|
| Thyroid Disorders | 1 or 2 Asymptomatic TFT abnormalities or mild symptoms | If hypothyroidism occurs, replace thyroid hormone until TSH/T4 levels return to the normal range. If the patient has thyrotoxicosis, consult an endocrinologist. If systemic symptoms arise: suspend investigational treatment, administer a beta-blocker, and consider oral prednisone 0.5 mg/kg/day for thyroid pain. Gradually taper the corticosteroid dose over 2-4 weeks. Monitor thyroid function to determine if hormone replacement therapy is needed. | Continue investigational treatment; if systemic symptoms occur, discontinue treatment. |
| | 3 or 4 Severe symptoms requiring hospitalization | Refer the patient to an endocrinologist. If hypothyroidism is diagnosed, replace with thyroid hormone at a dose of 0.5-1.6 | Suspend investigational treatment; resume medication once symptoms |

| Immune-related Toxicity | Grade | Treatment Guidelines | Management of Investigational Drugs |
|---|---|---|---|
| | | g/kg/day (for elderly patients or those with complications, a starting dose of 0.5 g/kg/day is recommended). Increase oral prednisone to 0.5 mg/kg/day for thyroid pain. Patients with thyrotoxicosis should be treated with a beta-blocker, and may also require carbimazole until the thyroiditis resolves. | improve to grade 0 or 1. |
| Pituitary Inflammation | 1 or 2<br>Mild to moderate symptoms | If hormone replacement therapy is needed, refer the patient to an endocrinologist. For patients with pituitaryitis, increase oral prednisone to 0.5-1 mg/kg/day. Gradually reduce the corticosteroid dose over at least 1 month. If there is no improvement within 48 hours, manage as grade 3 or 4. | Continue researching treatment |
| | 3 or 4<br>Severe or life-threatening | Refer the patient to an endocrinologist for assessment and | Pause research treatment for patients with |

| Immune-related Toxicity | Grade | Treatment Guidelines | Management of Investigational Drugs |
|---|---|---|---|
| | symptoms | treatment. Administer 1 mg/kg of methylprednisolone for headaches/visual disturbances caused by pituitary inflammation. Switch to oral prednisone and gradually reduce the dose over at least 1 month. Maintain hormone replacement therapy according to the endocrinologist's recommendations. | headaches/visual disturbances due to pituitary inflammation until relief/improvement to grade 2 or lower. Generally, discontinuation of medication is not required. |
| Non-infectious pneumonia | 1<br>Simple radiological changes. | Monitor symptoms every 2-3 days.<br>If signs worsen, manage as grade 2. | Consider pausing research treatment until signs improve and the cause is determined. |
| | 2<br>Symptoms: exertional dyspnea. | If an infection is suspected, start antibiotics. If symptoms/signs persist for 48 hours or worsen, administer oral prednisone at 1 mg/kg/day. Consider prophylaxis for Pneumocystis pneumonia. Gradually reduce the corticosteroid dose | Pause research treatment. If symptoms completely resolve or prednisone is maintained at ≤10 mg/day, treatment can be resumed. If symptoms persist after corticosteroid |

| Immune-related Toxicity | Grade | Treatment Guidelines | Management of Investigational Drugs |
|---|---|---|---|
| | | over at least 6 weeks. Consider preventive measures for steroid side effects, such as blood sugar monitoring and supplementation with vitamin D/calcium. | treatment, discontinue research treatment. |
| | 3 or 4<br>Severe or life-threatening symptoms<br>Apnea at rest. | Hospitalize and initiate intravenous methylprednisolone at 2-4 mg/kg/day. If no improvement or worsening occurs after 48 hours, increase infliximab to 5 mg/kg (if liver not involved). Switch to oral prednisone and gradually reduce the dose over at least 2 months. Use empirical antibiotics and consider prophylaxis for Pneumocystis pneumonia and other steroid side effects, such as blood sugar monitoring and vitamin D/calcium supplementation. | Discontinue research treatment. |
| Neurotoxicity | 1<br>Mild symptoms | - | Continue researching treatment |
| | 2 | Oral prednisone 0.5-1 | Suspend the |

| **Immune-related Toxicity** | **Grade** | **Treatment Guidelines** | **Management of Investigational Drugs** |
|---|---|---|---|
| | Moderate symptoms | mg/kg/day for treatment. Gradually reduce the dose over at least 4 weeks. Request a neurology consult. | study treatment; resume medication after symptoms relieve/improve to level 0 or 1. |
| | 3 or 4<br>Severe or life-threatening symptoms | Based on symptoms, initiate treatment with oral prednisone or intravenous methylprednisolone at 1-2 mg/kg/day. Gradually reduce the corticosteroid dose over at least 4 weeks. If there is no improvement within 72-96 hours, consider azathioprine, MMF, or cyclosporine. | Discontinue research treatment. |
| Colitis/Diarrhea | 1<br>Mild symptoms: More than 3 liquid stools per day compared to baseline, feeling well. | Symptomatic treatment: fluids, loperamide, avoid high-fiber/lactose diet. If level 1 persists for more than 14 days, manage as level 2 events. | Continue researching treatment |
| | 2<br>Moderate symptoms: 4-6 more liquid stools per day | Oral prednisone 0.5 mg/kg/day (non-enteric coated). Start treatment immediately, without waiting for | Suspend study treatment; resume medication once |

| Immune-related Toxicity | Grade | Treatment Guidelines | Management of Investigational Drugs |
|---|---|---|---|
| | compared to baseline, or abdominal pain, or blood in stool, or nausea, or nighttime episodes. | diagnostic results. Gradually taper steroids over 2-4 weeks; if symptoms recur, consider endoscopy. | relief/improvement returns to baseline level. |
| | 3<br>Severe symptoms: More than 6 liquid stools per day compared to baseline, or the need to defecate within 1 hour after eating. | Start intravenous methylprednisolone 1-2 mg/kg/day. Switch to oral prednisone and gradually taper the dose over at least 4 weeks. Consider preventing steroid side effects, such as blood sugar monitoring and supplementing with vitamin D/calcium. | Suspend study treatment; consider resuming treatment once relief/improvement returns to baseline level. |
| | 4<br>Life-threatening symptoms. | If no improvement or symptoms worsen within 72 hours, and if there are no perforation, sepsis, tuberculosis (TB), hepatitis, or NYHA Class III/IV CHF, consider infliximab 5 mg/kg; otherwise, consider other immunosuppressants: MMF or tacrolimus. Please consult a gastroenterologist for colonoscopy/sigmoido | Discontinue research treatment. |

| Immune-related Toxicity | Grade | Treatment Guidelines | Management of Investigational Drugs |
|---|---|---|---|
| | | scopy. | |
| Skin Reaction | 1<br>Rash, with or without accompanying symptoms, affecting <10% of body surface area (BSA). | Avoid skin irritation and sun exposure; it is recommended to use topical moisturizers. | Continue researching treatment |
| | 2<br>Rash covering 10%-30% of body surface area (BSA). | Avoid skin irritation and sun exposure; it is recommended to use topical moisturizers.<br><br>Topical steroids (medium-potency cream once daily or high-potency cream twice daily) ± oral or topical antihistamines for itching. Consider short-term oral steroids. | Continue researching treatment |
| | 3<br>Rash covering >30% of body surface area (BSA) or Grade 2, accompanied by significant symptoms. | Avoid skin irritation and sun exposure; it is recommended to use topical moisturizers. Based on clinical judgment, decide whether to start using steroids:<br>Moderate symptoms: Oral prednisone 0.5-1 mg/kg/day for 3 days, then gradually taper | Pause the study treatment. After discussing with the study medical monitor, if the adverse event (AE) has resolved or improved to a mild rash |

| Immune-related Toxicity | Grade | Treatment Guidelines | Management of Investigational Drugs |
|---|---|---|---|
| | | over 2-4 weeks. Severe symptoms: Intravenous methylprednisolone 0.5-1 mg/kg/day; switch to oral prednisone and gradually reduce the dose over at least 4 weeks. | (Grade 1 or 2), treatment may be resumed. |
| | 4 Necrosis of the skin covering >30% of body surface area (BSA), accompanied by related symptoms (such as erythema, purpura, epidermal shedding). | Initiate intravenous methylprednisolone at 1-2 mg/kg/day. Transition to oral prednisone, tapering the dose over at least 4 weeks. Hospitalize the patient and urgently consult dermatology. | Discontinue research treatment. |
| Hepatitis | 1 ALT or AST > ULN to 3×UL | Recheck LFT within one week, and recheck LFT before the next dose to confirm no deterioration has occurred. If LFT worsens, recheck every 48-72 hours until improvement is observed. | If LFT shows no change or improvement, continue the study treatment. If LFT worsens, pause the study treatment until improvement is observed. |
| | 2 | For persistent | Pause the study |

| Immune-related Toxicity | Grade | Treatment Guidelines | Management of Investigational Drugs |
|---|---|---|---|
| | ALT or AST 3-5× ULN | ALT/AST elevation, consider oral prednisone at 0.5 - 1 mg/kg/day for 3 days, then gradually taper over 2-4 weeks.<br>For elevated ALT/AST, initiate oral prednisone at 1 mg/kg/day and taper over 2-4 weeks. If LFT worsens, adjust the dosage based on clinical judgment. | treatment; once there is relief/improvement to baseline levels, treatment can be resumed. Gradually reduce the prednisone dosage to ≤ 10 mg. |
| | 3<br>ALT or AST >5-20 × ULN | If ALT/AST < 400 IU/L and bilirubin/INR/albumin are normal: start oral prednisone at 1 mg/kg and gradually taper the dose over at least 4 weeks.<br>If ALT/AST > 400 IU/L or bilirubin/INR is elevated/albumin is low: start intravenous (methyl)prednisolone at 2 mg/kg/day. Once LFT improves to grade 2 or below, switch to oral prednisone and gradually taper the dose over at least 4 weeks. | Pause the study treatment until improvement to baseline level. |
| | 4<br>ALT or AST > | Start intravenous methylprednisolone at | Discontinue research |

| Immune-related Toxicity | Grade | Treatment Guidelines | Management of Investigational Drugs |
|---|---|---|---|
| | 20 × ULN | 2 mg/kg/day. Switch to oral prednisone and gradually taper the dose over at least 6 weeks. | treatment. |
| | If LFT continues to worsen despite steroid use, consider reassessing the diagnosis and exploring alternative treatments or interventions:<br>• If on oral prednisone, switch to pulse IV methylprednisolone.<br>• If on IV steroids, increase MMF to 500-1000 mg twice daily.<br>• If there is worsening after starting MMF, consider adding tacrolimus.<br>The duration and dosage of steroid therapy will depend on the severity of the events. | | |
| Nephritis | 1<br>Creatinine 1.5 times baseline or > ULN or 1.5 times ULN. | Recheck creatinine weekly. If symptoms worsen, manage according to the following criteria. | Continue researching treatment |
| | 2<br>Creatinine > 1.5 to 3 times baseline or > 1.5 to 3 times ULN. | Ensure hydration and recheck creatinine within 48-72 hours; if no improvement, consider 24-hour urine collection for creatinine clearance. Discuss with a nephrologist about the need for a kidney biopsy. If due to investigational drugs, initiate oral prednisone 0.5-1 mg/kg and taper | Pause investigational treatment. If not due to drug toxicity, restart treatment. If due to investigational drugs and symptoms have improved to baseline levels, resume the investigational |

| **Immune-related Toxicity** | **Grade** | **Treatment Guidelines** | **Management of Investigational Drugs** |
|---|---|---|---|
| | | over at least 2 weeks. Recheck creatinine/U&E every 48-72 hours. | drug while tapering prednisone to <10 mg. |
| | 3<br>Creatinine > 3 times baseline or > 3 to 6 times ULN | Admit the patient for monitoring and fluid balance restoration; recheck creatinine daily. Consult a nephrologist to discuss the necessity of a kidney biopsy. If condition worsens, initiate IV (methyl)prednisolone 1-2 mg/kg and taper corticosteroids over at least 4 weeks. | Pause investigational treatment until the cause is identified. If the investigational drug is suspected, discontinue the treatment. |
| | 4<br>Creatinine > 6 times ULN | Based on tier three criteria, the patient should be treated in a facility capable of providing renal replacement therapy. | Discontinue research treatment. |
| Diabetes/Hyperglycemia | 1<br>asting blood glucose value ULN reaches 160 mg/dL; ULN up to 8.9 mmol/L. | Closely monitor and manage according to local guidelines. It is recommended to check C-peptide, glutamic acid decarboxylase antibodies, and islet cell antibodies. | Continue researching treatment |
| | 2<br>Fasting blood glucose value | At least weekly blood glucose level monitoring is | Continue to investigate treatment |

| Immune-related Toxicity | Grade | Treatment Guidelines | Management of Investigational Drugs |
|---|---|---|---|
| | ULN reaches 160-250 mg/dL; 8.9-13.9 mmol/L. | recommended. Management should be conducted according to local guidelines | options. If hyperglycemia worsens, discontinue treatment. If blood glucose stabilizes at baseline or levels 0 or 1, resume treatment |
| | 3<br>Fasting blood glucose value ULN reaches 250-500 mg/dL; 13.9-27.8 mmol/L. | Arrange for patient admission and refer to a diabetes specialist for management of hyperglycemia. Glucocorticoids may exacerbate hyperglycemia and should be avoided | Pause investigational treatment until the patient's hyperglycemia symptoms improve and blood glucose stabilizes at baseline or levels 0 or 1. |
| | 4<br>Fasting blood glucose value ULN exceeds 500 mg/dL; greater than 27.8 mmol/L. | Arrange for patient admission and establish local diabetes emergency management protocols. Refer the patient to a diabetes specialist for insulin maintenance and monitoring | |
| Retinotoxicity | 1<br>Asymptomatic, with abnormal findings on eye examination/testing. | Consider other causes and prescribe local treatment as needed. | Continue researching treatment |

| Immune-related Toxicity | Grade | Treatment Guidelines | Management of Investigational Drugs |
|---|---|---|---|
| | 2<br>Anterior uveitis or mild symptoms | Refer the patient to an ophthalmologist for assessment and local corticosteroid treatment. Consider a short course of oral steroids. | Continue researching treatment; if symptoms worsen or visual disturbances occur, discontinue treatment. |
| | 3<br>Posterior uveitis or panuveitis with significant symptoms. | Urgently refer the patient to an ophthalmologist. Start oral prednisolone at 1-2 mg/kg and taper over at least 4 weeks. | Pause treatment research until improvement reaches grade 0 or 1. |
| | 4<br>Affected eye blindness (at least 20/200) | Initiate intravenous (methyl)prednisolone at 2 mg/kg/day. Transition to oral prednisolone, tapering over at least 4 weeks. | Discontinue research treatment. |
| Pancreatitis | 1<br>Asymptomatic, abnormal blood test results. | Monitor pancreatic enzymes | Continue researching treatment |
| | 2<br>Abdominal pain, nausea, and vomiting | Arrange for hospitalization for emergency treatment. Start intravenous administration of (methyl)prednisolone at 1-2 mg/kg/day. If amylase/lipase improves to grade 2, | Discontinue research treatment. |

| Immune-related Toxicity | Grade | Treatment Guidelines | Management of Investigational Drugs |
|---|---|---|---|
| | | switch to oral prednisolone and gradually taper over at least 4 weeks. | |
| | 3<br>Acute abdominal pain, emergency surgery | Arrange for hospitalization for emergency treatment and appropriate referral. | Discontinue research treatment. |
| Arthritis | 1<br>Mild pain accompanied by inflammation and swelling. | Manage according to local guidelines. | Continue researching treatment |
| | 2<br>Moderate pain accompanied by inflammation and swelling, with limited fine motor activities due to instrument use. | Manage according to local guidelines. Consider referring the patient to a rheumatologist. If symptoms worsen during treatment, handle as a level 3 event. | Continue treatment; if symptoms persist in worsening, pause research treatment until symptoms improve to baseline or level 0 or 1. |
| | 3<br>Severe pain accompanied by inflammation or permanent joint damage, with limitations in daily activities. | Refer the patient to a rheumatologist for evaluation and treatment. Initiate oral prednisone at 0.5-1 mg/kg, tapering the dose gradually over at least 4 weeks. | Pause research treatment until symptoms improve to level 0 or 1. |
| Mucositis/Oral mucositis | 1<br>Mild symptoms | Consider local treatment or analgesics | Continue researching |

| Immune-related Toxicity | Grade | Treatment Guidelines | Management of Investigational Drugs |
|---|---|---|---|
| | or isolated test results. | based on local guidelines. | treatment |
| | 2<br>Moderate pain, reduced oral intake, and limited use of instruments. | Based on local guidelines, administer analgesics, local treatments, and oral hygiene care. Ensure adequate hydration. If symptoms worsen or if signs of sepsis or bleeding occur, manage as a grade 3 event. | Continue researching treatment |
| | 3<br>Severe pain, limited intake of food and fluids, and restricted daily activities. | Arrange for hospitalization to receive appropriate treatment. Initiate intravenous (methyl)prednisolone at 1-2 mg/kg/day. If symptoms improve to grade 2, switch to oral prednisolone and taper off gradually over at least 4 weeks. | Suspend study treatment until improvement to grade 0 or 1. |
| | 4<br>Life-threatening complications or dehydration. | Arrange for hospitalization for emergency care. If there are no contraindications to infection, consider intravenous corticosteroids. | Discontinue research treatment. |
| Myositis/Rhabdomyolysis. | 1<br>Mild symptoms | Consider local treatment or analgesics | Continue researching |

| **Immune-related Toxicity** | **Grade** | **Treatment Guidelines** | **Management of Investigational Drugs** |
|---|---|---|---|
| | or results from simple tests. | based on local guidelines. | treatment |
| | 2<br>Moderate weakness with or without pain. | If CK reaches 3×ULN or worse, start oral prednisone at 0.5-1 mg/kg, gradually tapering over at least 4 weeks. | Pause the study treatment until improvement to grade 0 or 1. |
| | 3 or 4<br>Severe weakness, self-limiting. | Admit the patient and start oral prednisone at 1 mg/kg. Consider IV (methyl)prednisolone at 1-2 mg/kg/day for severe activity limitation or swallowing difficulties as maintenance therapy. If symptoms do not improve, escalate immunosuppressive treatment and gradually taper oral steroids over at least 4 weeks. | Pause the study treatment until improvement to grade 0 or 1. If there is any evidence of cardiac involvement, discontinue treatment. |
| Myocarditis | <2<br>Asymptomatic but with significantly elevated CK-MB or troponin, or clinically significant ventricular conduction | Initiate cardiac evaluation with repeat serum tests under close monitoring; consider referral to a cardiologist. If myocarditis is confirmed, classify as grade 2. | Pause the study treatment. If myocarditis is confirmed, permanently discontinue treatment for patients with moderate to severe |

| Immune-related Toxicity | Grade | Treatment Guidelines | Management of Investigational Drugs |
|---|---|---|---|
| | delay. | | symptoms. For asymptomatic or mildly symptomatic patients, do not resume treatment with relatlimab if cardiac parameters have not returned to baseline. |
| | 2<br>Symptoms after light to moderate physical activity. | Admit the patient and initiate oral prednisone or IV (methyl)prednisolone at a dose of 1-2 mg/kg/day. Consult a cardiologist and manage heart failure symptoms according to local guidelines. If there is no immediate relief, switch to pulse dosing of (methyl)prednisolone at 1 g/day and consider increasing MMF, infliximab, or antithymocyte globulin. | |
| | 3<br>Severe symptoms after light physical activity. | | |
| | 4<br>Life-threatening. | | |

### APPENDIX 1: ECOG PERFORMANCE STATUS

| Grade | Description |
|---|---|
| 0 | Fully active, able to carry on all pre-diseases performance without restriction. |
| 1 | Restricted in physically strenuous activity but ambulatory and able to carry out work of a light or sedentary nature, e.g., light housework, office work. |
| 2 | Ambulatory and capable of all self-care but unable to carry out any work activities; up and about more than 50% of waking hours |
| 3 | Capable of only limited self-care; confined to bed or chair more than 50% of waking hours |
| 4 | Completely disabled; cannot carry on any self-care; totally confined to bed or chair |
| 5 | Deaths |

Source: Oken MM, Creech RH, Tormey DC, et al. Toxicity and Response Criteria of the Eastern Cooperative Oncology Group. Am J Clin Oncol. 1982;5(6):649-55

### APPENDIX 2: CONTRACEPTIVE GUIDELINES AND DEFINITIONS OF "WOMEN OF CHILDBEARING POTENTIAL" AND "WOMEN NOT OF CHILDBEARING POTENTIAL."

#### Contraceptive Guidelines

The Clinical Trial Facilitation Group's recommendations on contraception and pregnancy testing in clinical research include the use of highly effective contraceptive methods. These methods include:

- Combined hormonal contraception (with estrogen and progestin) to suppress ovulation (oral, vaginal, or transdermal)
- Progestin-only hormonal contraception to suppress ovulation (oral, injectable, or implantable)
- Intrauterine device (IUD)
- Intrauterine hormone-releasing system
- Bilateral tubal ligation
- Male partners with vasectomies, provided the partner is the only potential participant and has undergone medical evaluation confirming the success of the surgery.
- Abstinence (defined as avoiding heterosexual intercourse during the entire exposure related to the study treatment) is also mentioned.

It is important to note that barrier methods (including male and female condoms, with or without spermicides) are not considered highly effective contraceptive measures and, if used, must be combined with one of the highly effective methods listed above.

#### Definitions of "Women of Childbearing Potential" and "Women not

**of Childbearing Potential."**

As defined in this protocol, "women of childbearing potential" refers to female patients who are physiologically capable of becoming pregnant. Conversely, "women not of childbearing potential" are those who meet any of the following criteria:

- Surgical sterilization (i.e., via bilateral tubal ligation, bilateral oophorectomy, or hysterectomy)
- Postmenopausal status, defined as:

  Age ≥ 55 years, with natural menopause having occurred for ≥ 12 months, or

  Age < 55 years, with no menstrual periods for ≥ 12 consecutive months, FSH levels > 30 IU/mL, and exclusion of other medical reasons for amenorrhea for ≥ 12 months, such as polycystic ovary syndrome or hyperprolactinemia.

If FSH testing is needed to confirm postmenopausal status, individuals using hormonal contraception or hormone replacement therapy should be excluded.

Source: Clinical Trial Facilitation Group (CTFG). Recommendations on Contraception and Pregnancy Testing in Clinical Trials. September 15, 2014.

### APPENDIX 3: THE RESPONSE EVALUATION CRITERIA IN SOLID TUMORS (RECIST) GUIDELINES, VERSION 1.1

The text below is obtained from the following reference:

Eisenhauer EA, Therasse P, Bogaerts J, et al.New response evaluation criteria in solid tumours: revised RECIST guideline (version 1.1).Eur J Cancer.2009;45(2):228-47.

**Definition**

Response and progression will be evaluated in this study using the international criteria proposed by the RECIST Committee (Version 1.1). Changes in only the largest diameter (uni-dimensional measurement) of the tumor lesions are used in the RECIST.

Note: Lesions are either measurable or non-measurable using the criteria provided below. The term “evaluable” in reference to measurability will not be used because it does not provide additional meaning or accuracy.

Measurable Disease

Tumor lesions: At least one dimension not less than the lower limit (of the instrument measurement) (the longest diameter on the measuring instrument will be recorded) must be accurately measured:

- 10 mm by CT scan (irrespective of scanner type) and MRI (no less than double the slice thickness and a minimum of 10 mm)
- 10 mm caliper measurement by clinical examination (when superficial)
- 20 mm by chest X-ray (if clearly defined and surrounded by aerated lung)

Malignant lymph nodes: To be considered pathologically enlarged and measurable, a lymph node must be ≥ 15 mm in short axis when assessed by CT scan (CT scan slice thickness recommended to be no greater than 5 mm). At baseline and in follow-up, only the short axis will be measured and followed.

Non-measurable Disease

All other lesions (or sites of disease), including small lesions (longest diameter ≥ 10 to < 15 mm with conventional techniques or < 10 mm using CT scan), are considered non-measurable disease. Leptomeningeal disease, ascites, pleural effusion or pericardial effusion shown by spiral CT, inflammatory breast disease, lymphangitic involvement of skin or lung, abdominal masses/abdominal organomegaly identified by physical examination that is not measurable by reproducible imaging techniques are all non-measurable.

Bone lesions:

- Bone scan, PET scan, or plain films are not considered adequate imaging techniques to measure bone lesions. However, these techniques can be used to confirm the presence or disappearance of bone lesions.
- Lytic bone lesions or mixed lytic-blastic lesions, with identifiable soft tissue components, that can be evaluated by cross sectional imaging techniques such as CT or MRI can be considered measurable lesions if the soft tissue component meets the definition of measurability described above.
- Acute bone lesions are non-measurable

Cystic lesions:

- Lesions that meet the criteria for simple cysts defined by X-ray should not be considered as malignant lesions (neither measurable nor non-measurable) since they are, by definition, simple cysts.
- Cystic lesions thought to represent cystic metastases can be considered measurable if they meet the definition of measurability

described above. However, if non-cystic lesions are present in the same patient, these are preferred for selection as target lesions. -

Lesions with prior local treatment:

Tumor lesions situated in a previously irradiated area, or in an area pertaining to other loco-regional therapy, are usually not considered measurable disease unless they are proven to still be present. The protocol should detail the conditions under which such lesions would be considered measurable disease.

Target Lesions

All measurable lesions up to a maximum of 2 lesions per organ and 5 lesions in total, should be identified as target lesions and recorded and measured at baseline. Target lesions should be selected based on their size (lesions with the longest diameter), be representative of all involved organs, but in addition should be those that lend themselves to reproducible repeated measurements.

Lymph nodes merit special mention since they are normal anatomical structures that may be visible by imaging even if not involved by tumor. Pathological nodes that are defined as measurable and may be identified as target lesions must meet the criterion of a short axis of ≥ 15 mm by CT scan. Only the short axis of these nodes will contribute to the baseline sum. The short axis of the node is the diameter normally used by radiologists to judge if a node is involved by solid tumor. Nodal size is normally reported as 2 dimensions in the plane in which the image is obtained (for CT scan, this is almost always the axial plane; for MRI the plane of acquisition may be transverse, sagittal, or coronal). The smaller of these measures is the short axis. For example, an abdominal node which is reported as being 20 mm × 30 mm has a short axis of 20 mm and qualifies as a malignant, measurable node. In this example, 20 mm should be recorded as the node measurement. All other pathological nodes (those with short axis ≥ 10 mm but < 15 mm) should be considered non-target lesions. Nodes that have a short axis < 10 mm are considered nonpathological and should not be recorded or followed.

A sum of the diameters (longest for non-nodal lesions, short axis for nodal lesions) for all target lesions will be calculated and reported as the baseline sum diameters. If lymph nodes are to be included in the sum, then as noted above, only the short axis is added into the sum. The baseline sum diameters will be used as reference to further characterize any objective tumor regression in the measurable dimension of the disease.

Non-Target Lesions

All other lesions (or sites of disease) including pathological lymph nodes should be identified as non-target lesions and should be recorded at baseline. Measurements are not required, and these lesions should be followed as “present”, “absent”, or in rare cases “unequivocal progression” (more details to follow). In addition, it is possible to record multiple non-target lesions involving the same organ on the case record form (e.g., “multiple enlarged pelvic lymph nodes” or “multiple liver metastases”).

**Guidelines for Evaluation of Measurable Disease**

All measurements should be recorded in metric notation, using calipers if clinically assessed. All baseline evaluations should be performed as close as possible to the treatment start and never more than 4 weeks before the beginning of the treatment.

The same method of assessment and the same technique should be used to characterize each identified and reported lesion at baseline and during follow-up. All the lesions must be evaluated by imaging rather than only through clinical examination, unless the lesions are assessable only by clinical examination and cannot be imaged.

Clinical lesions:

Clinical lesions that are superficial and below P 10 mm in diameter (e.g., skin nodules) are considered to be measured using calipers. For the case of skin lesions, documentation by color photography including a scale to measure the size of the lesion is suggested. As noted above, when lesions can be evaluated by both clinical examination and imaging, imaging evaluation should be undertaken since it is more objective and may also be used for post-treatment review of study endpoints.

- Chest X-ray: chest CT is preferred for measuring lesions on chest X-ray and chest CT, particularly in the event of progression of an important treatment endpoint because CT is more sensitive than X-ray, particularly in identifying new lesions. Of course, lesions on chest X-ray can also be measurable if they are clearly defined and surrounded by aerated lung. -

- CT, MRI: CT is the best currently available and reproducible method to measure lesions selected for response assessment. This guideline has defined measurability of lesions on CT scan based on the assumption that CT slice thickness is 5 mm or less. When CT scans have a slice thickness greater than 5 mm, the minimum size for a measurable lesion should be twice the slice thickness. MRI is also acceptable in certain situations (e.g., for body scans).

- Ultrasound: Ultrasound is not useful in assessment of lesion size and should not be used as a method of measurement. Ultrasound examinations cannot be reproduced in their entirety for the review at a later date, and because the results are operator dependent, it cannot be guaranteed that the same technique and measurements will be taken from one assessment to the next. If new lesions are identified by ultrasound in the course of the study, confirmation by CT or MRI is advised. If there is concern about radiation exposure at CT, MRI may be used instead of CT in certain cases.

- Endoscopy, laparoscopy: The utilization of these techniques for solid tumor evaluation is not advised. However, they can be useful to confirm complete pathological response when biopsies are obtained or to determine relapse after complete response (CR) or surgical resection.

- Tumor markers: Tumor markers alone cannot be used for response evaluation in solid tumors. If markers are initially above the upper normal limit, they must normalize for a patient to be considered in CR. Because tumor markers are disease specific, instructions for their measurement techniques should be incorporated into protocols on a disease specific basis.

Specific guidelines for changes in CA-125 (in recurrent ovarian cancer) and prostate-specific antigen (in recurrent prostate cancer) have been published. In addition, the Gynecologic Cancer Intergroup has developed CA-125 progression criteria which are to be integrated with objective tumor assessment for use in firstline trials in ovarian cancer.

- Cytology, histology: These techniques can be used to differentiate between PR and CR in rare cases if required by protocol (for example, residual lesions in tumors like germ cell tumors, where known residual benign tumors can remain). When effusions are known to be a potential side effect of treatment (e.g, with certain taxane compounds or angiogenesis inhibitors), the cytological confirmation of the neoplastic origin of any effusion that appears or worsens during treatment can be considered if the measurable tumor has met criteria for response or stable disease to differentiate between response (or stable disease) and PD.

**Response Evaluation Criteria**

**<u>Evaluation of Target Lesions</u>**

- Complete Response (CR): Disappearance of all target lesions. All pathological lymph nodes (whether target or not) must have reduction in short axis to < 10mm. -

- Partial response (PR): At least a 30% decrease in the sum of diameters of target lesions, taking as reference the baseline sum diameters

- Progressive disease (PD): At least a 20% increase in the sum of diameters of target lesions, taking as reference the smallest sum on study (this includes the baseline sum if that is the smallest on study). In addition to the relative increase of 20%, the sum must also demonstrate an absolute increase of at least 5 mm. (Note: The appearance of 1 or more new lesion(s) is also considered progression).

- Stable disease (SD): Neither sufficient shrinkage to qualify for PR nor sufficient increase to qualify for PD, taking as reference the smallest sum diameters while on study

- Lymph nodes: Lymph nodes identified as target lesions should always have the actual short axis measurement recorded (measured in the same anatomical plane as the baseline examination), even if the nodes regress to below 10mm on study. This means that when lymph nodes are included as target lesions, the “sum” of lesions may not be zero even if CR criteria are met, since a normal lymph node is defined as having a short axis of < 10 mm. Case report recorded in a separate section where, to qualify for CR, each node must achieve a short axis < 10 mm. For PR, SD and PD, the actual short axis measurement of the nodes is to be included in the sum of target lesions.

Target lesions that become “too small to measure”: While on study, all lesions (nodal and non-nodal) recorded at baseline should have their actual measurements recorded at each subsequent evaluation, even when very small (e.g., 2 mm). However, sometimes lesions or lymph nodes which are recorded as target lesions at baseline become so faint on CT scan that the radiologist may not feel comfortable assigning an exact measurement and may report them as being “too small to measure”.

When this occurs, it is important that a measurement be recorded on the eCRF. If it is the opinion of the radiologist that the lesion has likely disappeared, the measurement should be recorded as 0 mm. If the lesion is believed to be present and is faintly seen but too small to measure, a default value of 5 mm should be assigned (Note: It is less likely that this rule will be used for lymph nodes since they usually have a definable size when normal and are frequently surrounded by fat, such as in the retroperitoneum; however, if a lymph node is believed to be present and is faintly seen but too small to measure, a default value of 5 mm should be assigned in this circumstance as well). This default value is derived from the 5 mm CT slice thickness (but should not be changed with varying CT slice thickness). The measurement of these (“too small to measure”) lesions is potentially nonreproducible; therefore, providing this default value will prevent false responses or progressions based upon measurement error. To reiterate, if the radiologist is able to provide an actual measurement, that measurement should be recorded, even if it is below 5 mm.

- Lesions that split or coalesce on treatment: When non-nodal lesions “fragment,” the longest diameters of the fragmented portions should be added together to calculate the sum (of diameters) of target lesions. Similarly, as lesions coalesce, a plane between them may be maintained before that which would aid in obtaining maximal diameters of each lesion. If the lesions have truly coalesced completely and are no longer separable, the vector of the longest diameter in this instance should be the longest diameter for the "coalesced lesion"

Evaluation of Non-Target Lesions

While some non-target lesions may be measurable, they may not be measured and instead be assessed only qualitatively at the timepoints specified in the protocol.

- CR: Regression of all non-target lesions and normalization of tumor marker level. All lymph nodes must meet non-pathological criteria in size (< 10 mm short axis).

- PD: Unequivocal progression (as detailed below) of existing non-target lesions.(Note: The appearance of 1 or more new lesion(s) is also considered progression.)

- Non-CR/Non-PD: Persistence of 1 or more non-target lesion(s) and/or maintenance of tumor marker level above the normal limits

- When the patient also has measurable disease: In this setting, to achieve “unequivocal progression” on the basis of the non-target disease, there must be an overall level of substantial worsening in non-target disease such that, even in presence of SD or PR in target disease, the overall tumor burden has increased sufficiently to merit discontinuation of therapy. The size of 1 or more non-target lesion(s) modestly“increase”, but is usually not sufficient to qualify for unequivocal progression status. The designation of overall progression solely on the basis of change in non-target disease in the face of SD or PR of target disease will therefore be extremely rare.

- When the patient has only non-measurable disease: This circumstance arises in some Phase 3 trials when it is not a criterion of enrollment to have measurable disease. The same general concept applies here as noted above; however, in this instance there is no measurable disease assessment to factor into the interpretation of an increase in non-measurable disease burden. Because worsening in non-target disease cannot be easily quantified (by definition: if all lesions are truly non-measurable), a useful test that can be applied when assessing patients for unequivocal progression is to consider if the increase in overall disease burden based on the change in non-measurable disease is comparable in magnitude to the increase that would be required to declare PD for measurable disease: i.e., an increase in tumor burden representing an additional 73% increase in “volume” (which is equivalent to a 20% increase in diameter in a measurable lesion). Such examples include an increase in a pleural effusion from “trace” to “large”, an increase in lymphangiopathy from localized to widespread, or may be described in protocols as “sufficient to require a change in therapy”. If “unequivocal progression” is observed, the patient should be considered to have had overall PD at that point. While it would be ideal to have objective criteria to apply to non-measurable disease, the very nature of that disease makes it impossible to do so; therefore, the increase must be substantial.

**New lesions**

The appearance of new malignant lesions denotes PD; therefore, some comments on detection of new lesions are important. There are no specific criteria for the identification of new lesions on X-ray; however, the finding of a new lesion should be unequivocal: i.e., not attributable to differences in scanning technique, change in imaging modality or findings thought to represent something other than tumor (for example, some “new” bone lesions may be simply healing or flare of preexisting lesions). This is particularly important when the patient’s baseline lesions show PR or CR. For example, necrosis of a liver lesion may be reported on a CT scan report as a “new” cystic lesion, which it is not.

A lesion identified on a follow-up study in an anatomical location that was not scanned at baseline is considered a new lesion and will indicate PD. An example of this is the patient who has visceral disease at baseline and while at the time of study entry has a CT or MRI brain scan ordered that reveals metastases. The patient's brain metastases can be considered evidence of PD even if he or she did not have brain imaging at baseline.

If a new lesion is equivocal, for example because of its small size, continued therapy and follow-up evaluation will clarify if it represents a truly new disease. If repeat scans confirm that there is definitely a new lesion, then progression should be declared using the date of the initial scan.

Evaluation of Best Overall Response

The best overall response is the best response recorded from the start of the study treatment until the end of treatment considering various factors for confirmation. On occasion, a response may not be documented until after the end of treatment, so protocols should be clear if post-treatment assessments are to be considered in determination of best overall response. Protocols must specify how various new therapies introduced before progression will affect best response designation. The patient's best overall response assignment is related to the findings of both target and non-target lesions and will also take into consideration the appearance of new lesions. Furthermore, due to the nature of the study and the protocol requirements, it may also require confirmatory measurement. Specifically, in nonrandomized study where response is the primary endpoint, confirmation of PR or CR is needed to deem either one the "best overall response".

The best overall response is determined once all the data for the patient are known. Best response determination in studies where confirmation of CR or PR is not required: Best response in these studies is defined as the best response across all timepoints (for example, a patient who has SD at first assessment, PR at second assessment, and PD on last assessment has a best overall response of PR). When stable disease is believed to be the best response, it must also meet the protocol-specified minimum time from baseline. If the protocol-specified minimum time is not met when SD is otherwise the best timepoint response, the patient's best response depends on the subsequent assessments. For example, a patient who has SD at first assessment, PD at second and does not meet minimum duration for SD, will have a best response of PD. The same patient lost to follow-up after the first SD assessment would be considered not evaluable.

| Target Lesions | Non-Target Lesions | New lesions | Overall Response |
|---|---|---|---|
| CR | CR | No | CR |
| CR | Non-CR/non-PD | No | PR |
| CR | Not evaluated | No | PR |
| PR | Non-PD or not all evaluated | No | PR |
| SD | Non-PD or not all evaluated | No | SD |
| Not all evaluated | Non-PD | No | NE |
| PD | Any | Yes or No | PD |
| Any | PD | Yes or No | PD |
| Any | Any | Yes | PD |

Abbreviations: CR, complete response; NE, not evaluable; PD, progressive disease; PR, partial response; SD, stable disease.

When nodal disease is included in the sum of target lesions and the nodes decrease to "normal" size (< 10 mm), they may still have a measurement reported on scans. This measurement should be recorded even though the nodes have resolved in order to not to overstate progression should it be based on increase in the size of the nodes. As noted earlier, this means that patients with CR may not have a total sum of "zero".

In studies where confirmation of response is required, repeated "not evaluable (NE)" timepoint assessments may complicate best response determination. The analysis plan for the study must address how missing data/assessments will be addressed in determination of the response and progression. For example, in most studies it is reasonable to consider a patient with timepoint responses of PR-NE-PR as a confirmed response.

Patients with a global deterioration of health status requiring discontinuation of treatment without objective evidence of PD at that time should be reported as "symptomatic deterioration". Every effort should be made to confirm objective progression even after discontinuation of treatment. Symptomatic deterioration is not a descriptor of an objective response: it is a reason for stopping study therapy.

Conditions that define “early progression”, “early death”, and “un-evaluability” are study specific and should be clearly described in each protocol (depending on treatment duration, treatment cycle setting).

In some circumstances it may be difficult to distinguish residual disease from normal tissue. When the evaluation of CR depends upon the results, it is recommended that the residual lesion be investigated (fine-needle aspiration/biopsy) before assigning a status of CR. For equivocal findings of progression (e.g., very small and uncertain new lesions; cystic changes and necrosis in existing lesions, etc), treatment may continue until the next scheduled assessment timepoint. If progression is confirmed at the next scheduled assessment, the date of progression should be the earlier date when progression was suspected.

**Confirmatory Measurement/Duration of Response**

<u>Confirmation</u>

In nonrandomized studies where response is the primary endpoint, confirmation of PR and CR is required to identify whether responses are the result of measurement error or not. This will also permit appropriate interpretation of results in the context of historical data where response has traditionally required confirmation in such studies. However, in all other circumstances, i.e., in randomized studies (Phase 2 or 3) or studies where SD or progression are the primary endpoints, confirmation of response is not required since it will not add value to the interpretation of study results. However, elimination of the requirement for response confirmation may increase the importance of central review to protect against bias, in particular in studies that are not blinded.

In the case of SD, measurements must have met the SD criteria at least once after study entry at a minimum interval (in general not less than 6 weeks).

<u>Duration of Overall Response</u>

The DOR is measured from the time measurement criteria are first met for CR/PR (whichever is first recorded) until the first date that recurrent or progressive disease is objectively documented (taking as reference for PD the smallest measurements recorded on study).

The duration of overall CR is measured from the time measurement criteria are first met for CR until the first date that recurrent disease is objectively documented.

Duration of Stable Disease

Stable disease is measured from the start of the treatment (from date of enrollment) until the criteria for progression are met, taking as reference the smallest sum on study (if the baseline sum is the smallest, this is the reference for calculation of PD).

The clinical relevance of the duration of stable disease varies in different studies and diseases. If the proportion of patients achieving stable disease for a minimum period is an endpoint of importance in a particular study, the protocol should specify the minimal time interval required between 2 measurements for determination of stable disease.

Note: The DOR and duration of stable disease as well as the progression-free survival are influenced by the frequency of follow-up after baseline evaluation. It is not in the scope of this guideline to define a standard follow-up frequency. The frequency should take into account many parameters including disease types and stages, treatment cycles, and standard practice. However, these limitations of the precision of the measured endpoint should be considered if comparisons between studies are to be mad.
